# Cost-effectiveness of end-game strategies against sleeping sickness across the Democratic Republic of Congo

**DOI:** 10.1101/2024.03.29.24305066

**Authors:** Marina Antillon, Ching-I Huang, Samuel A. Sutherland, Ronald E. Crump, Paul E. Brown, Paul R. Bessell, Emily H. Crowley, Rian Snijders, Andrew Hope, Iñaki Tirados, Sophie Dunkley, Paul Verlé, Junior Lebuki, Chansy Shampa, Erick Mwamba Miaka, Fabrizio Tediosi, Kat S. Rock

**Affiliations:** Department of Epidemiology and Public Health, Swiss Tropical and Public Health Institute, Allschwil, Basel-Land, Switzerland; University of Basel, Basel, Switzerland; Zeeman Institute, University of Warwick, Coventry, UK; Mathematics Institute, University of Warwick, Coventry, UK; Warwick Medical School, University of Warwick, Coventry, UK; Independent consultant, Edinburgh, UK; Institute of Tropical Medicine, Antwerp, Belgium; Liverpool School of Tropical Medicine, Liverpool, UK; Programme National de Lutte contre la Trypanosomiase Humaine Africaine, Kinshasa, Democratic Republic of Congo

**Author notes:** These authors contributed equally to this work (Ching-I Huang, Samuel A. Sutherland, Ronald E. Crump).

**Keywords:** sleeping sickness, elimination of transmission, economic evaluation, modelling, Democratic Republic of Congo

## Abstract

**Background:** *Gambiense* human African trypanosomiasis (gHAT) is marked for elimination of transmission (EoT) by 2030. We examined the cost-effectiveness (CE) of EoT in the Democratic Republic of Congo, which has the highest global gHAT burden.

**Methods:** In 165 health zones (HZs), we modelled the transmission dynamics, health outcomes, and economic costs of six strategies during 2026–40, including the cessation of activities after cases reported reach zero. Uncertainty in CE was assessed within the net monetary framework, which presents the optimal strategies at a range of willingness-to-pay (WTP) values, denominated in costs per disability-adjusted life-year averted. We assessed the optimal strategy for CE and EoT in each health zone separately, but we present results by health zone as well as aggregated by coordination and for the whole country.

**Results:** Status quo strategies, CE strategies (WTP=$500), and strategies with a high probability of EoT by 2030 are predicted to yield EoT by 2030 in 118 HZs, 123 HZs, and 134 HZs respectively, at a cost by 2040 of $67.8M [95% PI: $36.7M–113M], $93.3M [95% PI: $51.6M–153M], $185M [95% PI: $111M–309M]. A more lenient timeline of EoT by 2040 could lead to EoT in 152 HZs at a cost of $158M [95% PI: $91.6-265M], leaving 13 HZs shy of the goal. Costs would have to be front-loaded; in 2026, while status quo strategies would cost $8.75M [95% PI: $7.01M–11.2M], elimination strategies would cost $27.0 [95% PI: $21.0M–35.2M]. Investing in EoT by 2030 is predicted to reduce 68% of gHAT deaths from 7979 [95% PI: 770–27,868] with status quo strategies to 2576 [95% PI: 255–9133].

**Conclusions:** The current arsenal of tools could make considerable progress to maximise the probability of EoT by 2030, but select health zones are facing a low probability of EoT even with more ambitious strategies. Investments need to be front-loaded, but we would witness considerable returns on investment by 2040.

## Background

*Gambiense* human African trypanosomiasis (gHAT), commonly known as sleeping sickness, is an enduring infectious disease that has afflicted certain West and Central African populations for centuries. Despite notable advancements in diagnosis, treatment modalities, and interventions, the disease continues to pose a significant threat to infected individuals lacking timely access to curative medications.

Throughout the 20th century, gHAT witnessed three epidemics, with the most recent surge occurring during the 1990s, during which a staggering 37,385 cases were documented across Africa in 1998, 26,318 of which were in the Democratic Republic of Congo (DRC). Encouragingly, the period between 2008 and 2020 marked a transformative phase in gHAT control strategies, leading to a substantial reduction in reported cases. In 2024, only 546 cases were recorded globally, 330 of which originated in the DRC [19, 58].

The progressive decline in both the overall burden of gHAT and the share borne by the DRC offers optimism to the global health community, suggesting that the national control programme in the DRC has made significant strides towards achieving elimination [19, 58]. Aligned with the London Declaration on Neglected Tropical Diseases, a meeting targeting the elimination of various neglected tropical diseases, gHAT was identified as a key priority for achieving elimination of transmission (EoT) by 2030 [54]. More recently, this target has been reiterated in the World Health Organization’s (WHO’s) 2030 roadmap for NTDs [57]. Previous studies have demonstrated that the observed reductions in reported cases are indicative of a decline in the underlying, albeit unobservable, transmission of gHAT, even in regions where screening coverage has diminished [14]. The currently available arsenal of tools exhibits promising potential in combating transmission throughout the DRC [23].

This analysis aims to evaluate the feasibility and cost-effectiveness of achieving the elimination of gHAT in the DRC. The study focuses on assessing six plausible strategies for gHAT control and elimination at the health-zone level. The strategies comprise different combinations of passive screening (PS) in fixed health facilities, active screening (AS) by mobile teams, treatment, and vector control (VC) using Tiny Targets, which attract and kill tsetse that transmit the parasite via insecticide-impregnated fabric [31, 48]. Building upon prior health economic analyses, which concentrated on five health zones with different risk levels, this study greatly expands the analyses to encompass 165 health zones across the DRC which have had reported cases [3]. The present study therefore provides a full DRC-wide picture of the costs, cost-effectiveness and probability of elimination.

To enhance the accuracy and reliability of the findings, the model fitting and projections are refined, using an additional four years (2017-2020) of gHAT case and screening data [14, 15]. Furthermore, the model accounts for epidemiological uncertainties related to potential non-human animal transmission by utilising an “ensemble model” approach. Stochastic projections are employed to better estimate variations in the timing of achieving elimination. In short, by using a comprehensive modelling framework, this study examines the intricate interplay among epidemiological dynamics, economic considerations, and temporal factors to inform decision-making regarding gHAT strategies to achieve EoT.

## Methods

The analysis process for each health zone is shown in Figure 1, consistent with our previous analyses [3, 14, 23]. In Step 1, the ensemble transmission model was re-fitted to 165 health zones; updates from the previously published analysis are summarised in Supplementary Materials, Section A.1. In Step 2, we simulated the current strategies and alternatives with the stochastic model as shown in Figure 1B and Figure 1C. In Step 3, we apply the simulated infections, cases reported, and undetected cases, to a treatment outcome model. A glossary of epidemiology and health economic terms is found in the appendix, Supplementary Note 1.

**Fig. 1.**
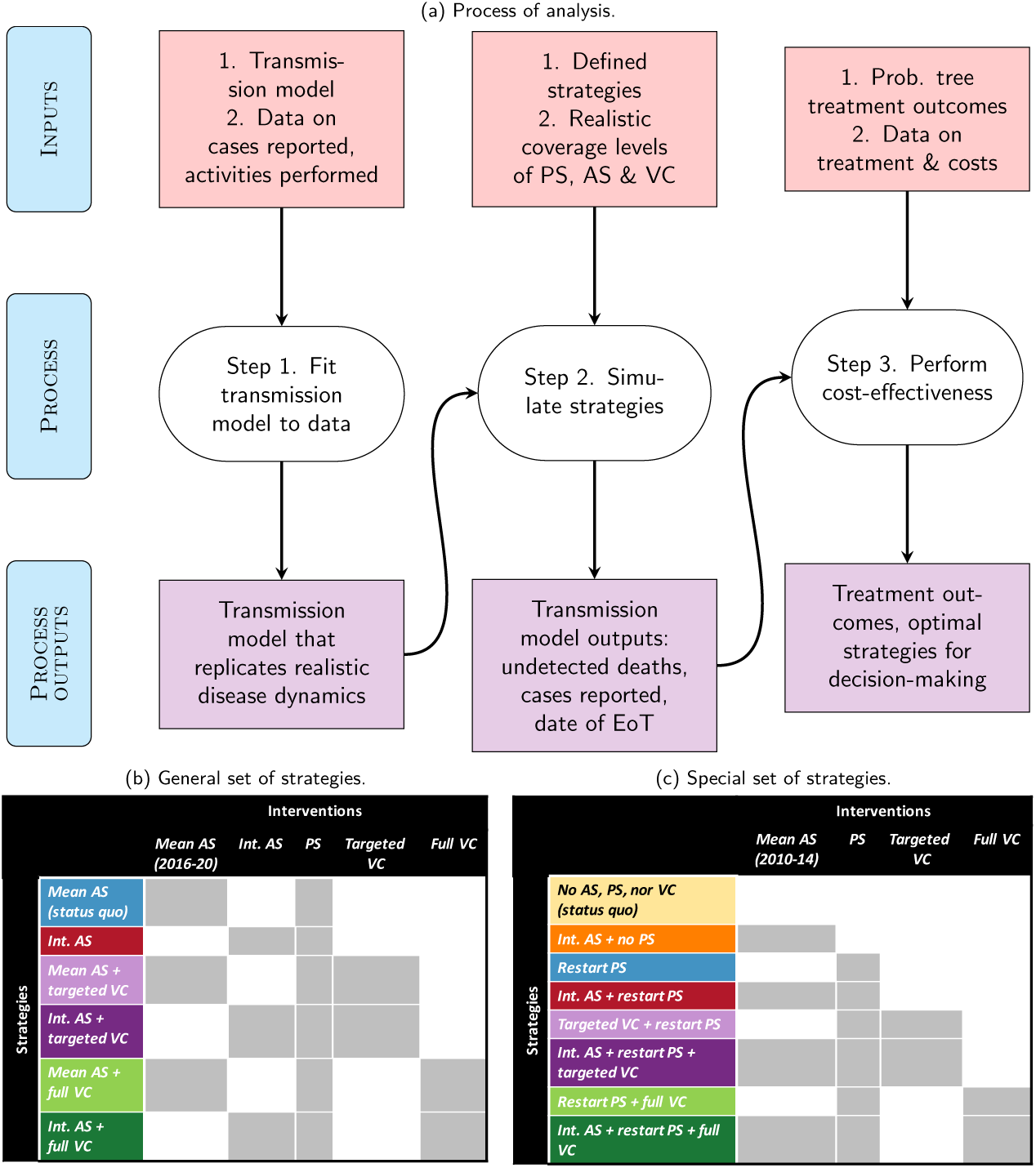
a) Process of analysis. b) Future strategies simulated in most health zones. *Mean AS* is equal to the mean coverage of AS between 2016–2020, *Int. AS* is 30% or the historical maximum coverage between 2000–2020, whichever is higher. c) Future strategies simulated in Ango, and Doruma health zones of the Bas Uélé region. We do not mean AS from 2016–2020 in these health zones as there was no AS during this period, hence *Restart PS* is effectively equivalent to this strategy. In these health zones, we take *Int. AS* to be the mean of 2010–2014 when MSF was operating in the region. In all health zones, the *Targeted VC* strategy only simulates VC along rivers in regions with high case density, and because the cases may be diffuse in some health zones, this strategy is not present in some health zones. Whereas the *Full VC* strategy involves the deployment of Tiny Targets throughout all large rivers in a health zone, regardless of the density of the cases. *Intervention cessation:* All strategies assume that AS will cease after 3 years of AS with zero cases in either AS or PS, followed by another AS in year 5 with no cases. RS is triggered if a case is found in PS and stops using the same 3+1 algorithm. VC stops after 3 years of no cases. PS is stopped 5 years after AS and RS have ceased. See the glossary and Table 8 for details of abbreviations. AS: active screening, PS: passive screening, VC: vector control, EoT: elimination of transmission.

### Step 1: Fitting the transmission model to health zones

#### Data

Historical case data by year and location for 2000–2020 was acquired from the WHO HAT Atlas. HAT Atlas data included records of the populations seen in mobile AS activities and PS activities in clinics in fixed locations [58]. We mapped location names (province, health zone, and health area) and geolocations onto two shapefiles provided by the American Red Cross [2] to aggregate records including AS numbers and case detections from both AS and PS by year in 519 health zones (see Supplementary Table 1). Health zones are delineated by the Ministry of Health to provide a unit of public health administration and to have a referral hospital for the treatment of all illnesses that cannot be handled in a health post, usually the first point of contact between the population and the healthcare system. Health areas are sub-units within health zones with at least one health post each.

For the data collected between 2015–2019, the HAT Atlas also includes staging information for the majority of cases. Stage-dependent data was missing in other years because a) before 2015 staging information was not routinely digitised even though staging would have been performed, as treatment was stage-dependent, and b) since 2020 many cases are eligible for fexinidazole, which no longer requires staging for treatment, and therefore this information is not collected any more.

For more details on the data and data extraction, see Supplementary Methods, Section A.2.

#### Settings

Out of 519 health zones in the DRC, 281 health zones reported either a) active screening, and/or b) one or more cases of sleeping sickness between 2000–2020 according to the HAT Atlas. Of those, 190 health zones had 10 or more data points consisting of a) years of AS, yielding any number of cases, including zero, plus b) years with one or more cases of gHAT reported from PS in fixed health facilities, or clinics, that have the capacity for parasitological (microscopy) confirmation. Following recommendations from the national gHAT programme (PNLTHA-DRC), we excluded several health zones in urban areas, primarily around Kinshasa and Mbuyi-Mbuyi, the capital of Kasaï Oriental Province. The programme believes that case reports in those health zones were likely due to importations from rural areas and the presence of a diagnostic hospital in those regions (see Supplementary Table 2). After these exemptions, 165 health zones remained, encompassing 131,941 cases, or 94% of all cases reported in 2000–2020.

The 165 health zones were distributed across 11 “coordinations”, which are subunits used by PNLTHA-DRC to coordinate intervention activities across the country. Coordinations were delineated in the early 2000s and are contiguous with, or parts of, the provinces before 2015, dividing these former provinces into approximately equal areas (see Supplementary Figure 1 in Supplementary Methods, Section A.3). Within coordinations, health zones are an administrative subunit. Although initially serving approximately 100,000 people, health zones in our analysis have a median of 210 thousand people in 2022 due to population growth, for a total of 37.6 million people (see Supplementary Table 3).

Two health zones are handled separately in this analysis: Ango in the Bas Uélé province of Isangi coordination, and Doruma in Haut Uélé, which we will consider part of Isangi coordination for the purpose of this analysis, although Doruma officially does not belong to any coordination. Together, we term this region “the Bas Uélé region” for this analysis. Historically, activities in these health zones were mostly handled by MSF instead of PNLTHA and screening and treatment activities have been absent between 2015 and 2022, when some exploratory research activities were initiated. No VC has ever been done in this region. For these health zones, special considerations were taken when re-fitting the data and an alternative set of strategies was simulated that take into account the distinct history of activities in these health zones.

Although the basic unit of analysis is the health zone, results are also aggregated by coordinations, provinces, former provinces, and for the whole country.

#### Transmission model

For this study, we used two variants of the previously published Warwick gHAT model [14, 15] consisting of a mechanistic, deterministic modelling framework to explicitly simulate transmission between humans and possibly animals via tsetse vectors (see Supplementary Figure 2 and the Supplementary Methods, Section A.4). Model parameterisation was performed individually for each model variant (models with and without possible animal transmission) and was updated compared to previous publications by fitting to WHO HAT Atlas data from 2000–2020 for each health zone in the DRC that had sufficient data—at least 10 or 13 data points for the models without and with animal transmission, respectively (where any year with AS and any year with non-zero passive case detection count as individual data points). This gave 165 health zones with a fit using the model without animal transmission and 156 that were fitted to both model variants. More details of the statistical fitting procedure are provided in the Supplementary Methods, Section A.5. The “ensemble” model consists of posteriors from both models. The proportion of samples from both models was determined by a statistical method (Bayes factors) that measures the relative goodness of fit of each model to the data.

### Step 2: Epidemiological projections

Stochastic simulations, which capture chance events at very low prevalence, were performed using the fitted model from Step 1. Technical details on the projections are included in Supplementary Methods, Section A.6. The list of strategies is refined compared to those presented in previous work, through conversations with PNLTHA-DRC, NGO, and academic partners [23].

Projections of up to eight plausible gHAT intervention strategies per health zone were simulated from 2026–2055 and are listed in Figure 1B; furthermore, each activity is further explained in the Supplementary Methods, Section A.7, and in particular in Supplementary Table 8. Each of our strategies represents a specific combination of three available interventions at different levels: 1) AS with mobile units moving from village to village, 2) PS in fixed health facilities with serological (RDT or CATT) tests and parasitology, 3) VC via Tiny Targets. Treatment of detected cases is included in all strategies.

AS is simulated at either a mean (*Mean AS*) coverage, targeting villages with case reporting in the last 5 years, or an intensified level of AS (*Intensified AS*) which we assume is achieved by increasing turnout, within target villages, additionally screening neighbouring villages and/or screening in locations which historically had case reporting. The *Mean AS* coverage was set equal to the mean coverage of the last five years of data (2016–2020), and the *Intensified AS* coverage was set equal to the highest coverage within a health zone in the period of 2000–2020 or 30%, whichever is highest. Four out of the six strategies also include VC, combining *Mean AS* or *Intensified AS* with *Targeted VC* or *Full VC*. *Targeted VC* is the deployment of VC along large rivers (defined here as an average long-term discharge estimate for a river reach of *>* 20 m^3^/s [30]) with a case density of at least 1 case per 10 km of the treated riverbank. *Full VC* is the deployment of VC along all large rivers in the health zone. The *Full VC* strategy was considered in 155 of the 165 health zones we analysed, and the targeted strategy was considered in the 45 health zones where the amount of river meeting the above criterion was both non-zero and less than the extent considered as *Full VC*. For more details, see Supplementary Methods, Section A.7.1. Besides AS and VC, PS is simulated to continue at estimated 2020 levels into the future based on the 2019 WHO survey of clinics; this level reflects some improvements of PS over the period of 2000-2020, estimated by model fitting. The quality of PS after 2020 is projected to remain the same.

Lastly, cessation of AS and VC strategies are enacted after three years of no cases. For AS, one more screening is deployed on the fifth year of no cases, per WHO recommendations for cessation [54]. PS is then ceased in all but one clinic in each health zone after eight years of zero case reports within the health zone.

The status quo (comparator) strategy differs in some locations, as some locations have already conducted VC in addition to medical interventions. Furthermore, ten health zones have no large rivers and therefore are not deemed suitable for deployment of VC. For the two health zones in the Bas Uélé region of Isangi coordination, we have simulated an alternative set of strategies, as listed in Figure 1C, because the status quo is “No AS, PS, nor VC”, and we had to consider the potential benefits of adding PS, which was present in all strategies of all other health zones. The details of the specific health zones with different strategies can be found in Supplementary Methods, Section A.7.

For an overview of the operational inputs in each health zone, see Supplementary Table 9, and for the specifics in each health zone, see Supplementary Note 2 and the graphical user interface that supplements this analysis (https://hatmepp.warwick.ac.uk/DRCCEA/v8/).

### Step 3: Cost-effectiveness analysis

The analysis for each health zone is governed by a decision tree as shown in the Supplementary Methods, Section A.8, Supplementary Figures 4-5. The decision tree invokes the transmission model detailed above as well as a treatment model depicted in Supplementary Figure 6.

#### Health outcomes

Health burden is denominated in disability-adjusted life years (DALYs), but we also report cases and deaths as intermediate outcomes (see Supplementary Methods, Section A.9). To calculate the probability of each treatment outcome, we used the outputs of the transmission model as inputs in a probability tree model of disease outcomes (see Supplementary Methods, Section A.10 and Supplementary Figure 6) and as done in previous work [3]. We simulated the disease process separately for stage 1 and stage 2 of the disease, including steps to sort patients into the appropriate treatment [53], treatment success or failure, diagnosis in the event of treatment failure, and progression to rescue treatment. All parameters are listed in Table 1, and more details are provided in Supplementary Note 2 for health-zone-specific parameters and Supplementary Note 3, the parameter glossary.

**Table 1:**
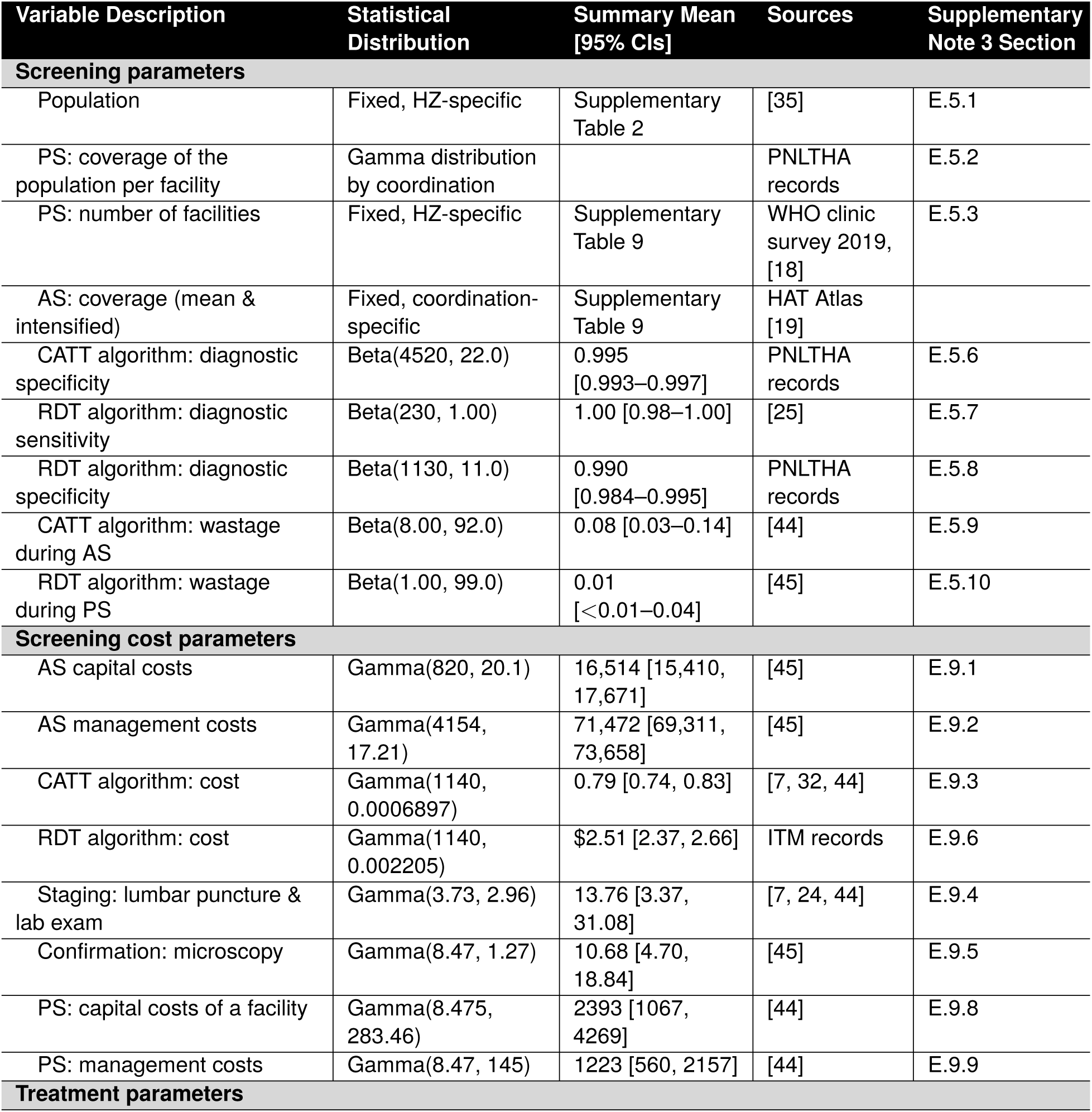
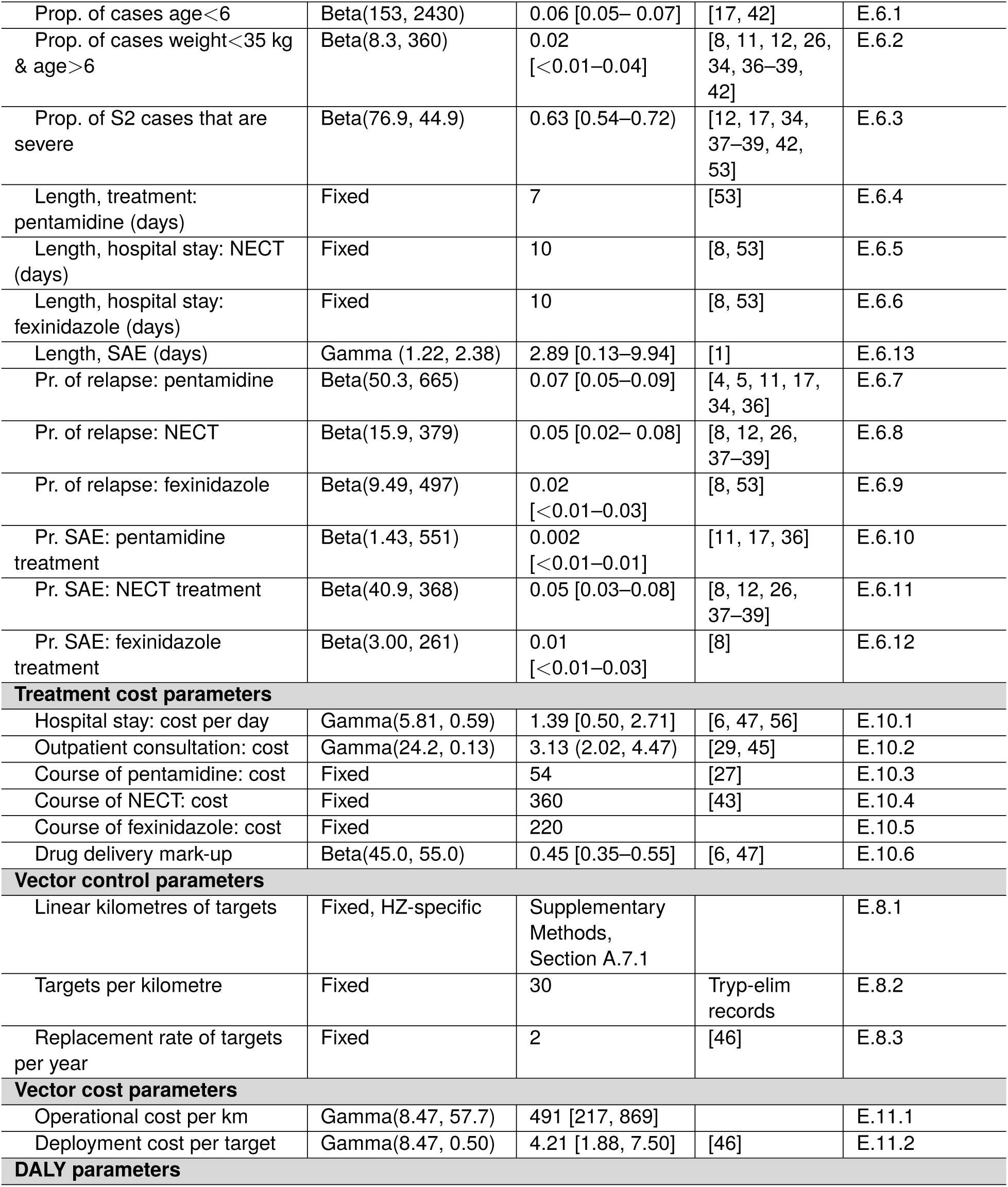
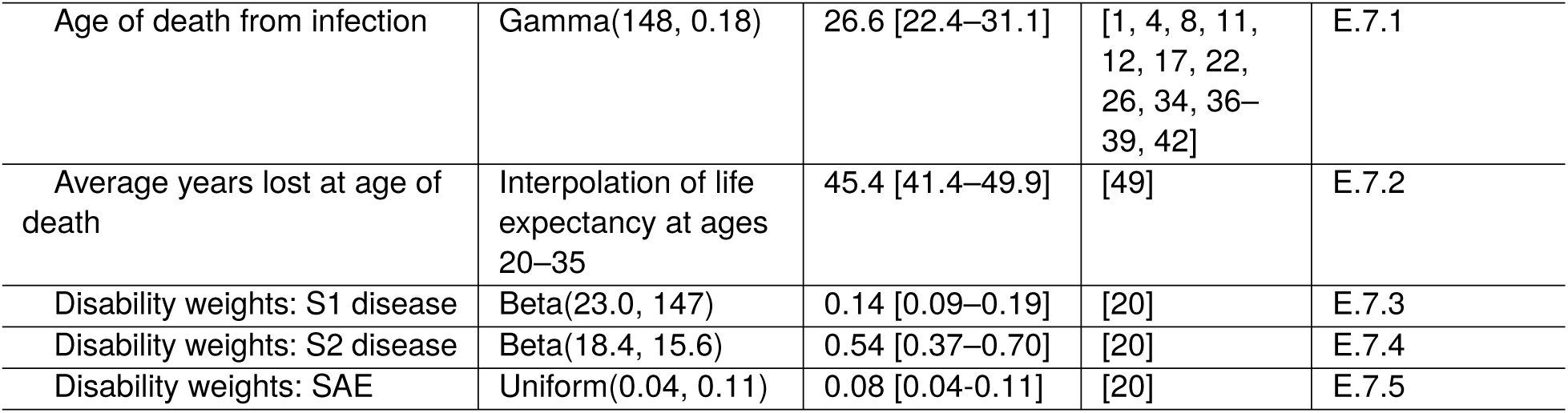
Model Parameters. For further details and sources, see Supplementary Note 3. CIs intervals. All values given to 3 significant figures. AS & PS: active and passive screening, respective control, PNLTHA: Programme de Lutte contre la Trypanosomie Humaine Africaine, NECT: nifurtimo combination therapy, CATT: card agglutination test for trypanosomiasis, S1 & S2: stage 1 & 2 dis disability-adjusted life-years, SAE: severe adverse events, RDT: Rapid Diagnostic Test

#### Economic Costs

We developed a cost function that incorporates the costs presented at each step of the transmission and treatment models, including fixed and variable costs of operating preventive and therapeutic activities (see Supplementary Methods, Section A.10). Cost data for AS, PS, VC, and treatment were acquired from the literature and conversations with programme staff ([44, 45], among others) and the unit costs and cost functions are detailed in the Supplementary Methods. For individuals screened during PS specifically, costs were deduced from two data sources: the historical records of clinics capable of screening with serological tests (RDT or CATT) [13], and records acquired from the PNLTHA for the years 2019–2020. Disease costs include diagnosis, confirmation, and staging (lumbar puncture) if required, as well as the cost of the drug itself and the administration by health care personnel (nurse or community health care worker).

We performed our analysis from the perspective of health- or intervention-delivery payers collectively (the government, the WHO, donors, etc). Although some activities have primarily been performed by foreign research partners in the past, we excluded any research personnel costs and calculated the value of their labour in local costs. Costs were inflated and converted to 2024 US dollars as detailed in Supplementary Note 3, Section E.1.

#### Cost-effectiveness estimates

For each health zone, we have run the transmission model 20,000 times, constituting 10 stochastic iterations per draw for 2,000 samples from a joint parameter distribution. Analogously, we sampled from the treatment costs and outcome parameters 20,000 times to propagate treatment- and cost-related uncertainty and calculate the costs of AS, PS, and VC (when applicable), which are detailed in Supplementary Methods, Section A.10. Lastly, we use the net monetary benefits (NMB) framework, which monetises DALYs averted (health benefits) according to a pre-defined willingness-to-pay (WTP) per DALY averted and subtracts the additional cost to deploy that strategy [9]. In this study, we have determined the optimal strategies at investment levels, which encompasses the usual range of values considered to be “cost-effective” for a country with a GDP per capita of $577 in 2021 [33, 55, 56]. Unlike computing single incremental cost-effectiveness ratios (ICERs), this takes into account all uncertainty and refrains the analysis from invoking any single WTP threshold as the cost-effective threshold. To determine the optimal strategy for a health zone across all iterations, one selects the strategy with the maximum NMB, averaged over all iterations.

We discount both costs and health outcomes at a rate of 3% annually. We adopt a time horizon that is relatively long (2026–2040), as opposed to the conventional 10 years, in order to assess the returns on investments in both augmented disease control and elimination after the 2030 goal has been reached.

Once we have the cost-effectiveness results for each health zone, we aggregate health outcomes such as cases, DALYs, and deaths, and cost outcomes in dollars, by coordination, province, and for the whole country. However, we did not aggregate the health and cost outcomes by strategy, as the costs and benefits of implementing a single strategy everywhere would not reflect any decision-making that would be made. Rather, we aggregated the costs and benefits by willingness-to-pay or EoT outcomes, which we called “objectives”. Since what is likely to be decided is that the country will implement strategies that align with a specific WTP or EoT objective that is consistent everywhere, we conditioned the total outcomes and total costs by the objectives. In other words, for the costs and health benefits at the coordination, province, and national level, we take the strategy in each health zone that would be optimal either at each WTP ($0–1500) or for the EoT objective we take the cheapest strategy that leads to EoT with >90% probability.

With the designated set of strategies, we then add the health outcomes and the costs of all the health zones included in each geographic designation. In health zones where strategies yield *<*90% probability of EoT by 2030, the strategy with the maximum probability of EoT by 2030 is selected. The aggregations are performed assuming full correlation (rank=1 covariance structure) between the health zones to compute the 95% predictive intervals and expected values. When we did this, the sum of the cases reported more adequately fit the sum of the cases reported at the coordination, provincial, and national levels compared to assuming total independence between health zones.

## Results

### Step 1: Fitting the model

The fitted model predictions for a sample health zone of Kikongo are available in Supplementary Figure 7 in the Supplementary Results, Section B.1. Modelling shows that in addition to decreasing numbers of PS- and AS-detected cases, the new infections are also decreasing, and since Kikongo only started VC in mid-2019, and our data only runs until 2020, this is pre-dominantly due to screening, either in mobile teams or in fixed health centres. Similar results are available for all health zones in the GUI https://hatmepp.warwick.ac.uk/DRCCEA/v8/. Model selection results are found in the Supplementary Results, Section B.2. Broadly speaking, for the 156/165 health zones that had enough data to fit the animal model, only one health zone (Bokoro) showed strong evidence of a contribution of animals to transmission, and 8 other health zones showed weak evidence of animal dynamics in transmission, as shown in Supplementary Figure 8. This contrasts with our previous publication, where 24 health zones showed more than weak support for animal dynamics in transmission [15]. In 29 health zones, although there is no strong evidence of animal dynamics, there is weak evidence of an absence of animal dynamics, yielding considerable uncertainty on this question. In Bokoro, there is both strong evidence for the role of animals, and more than a 10 percentage point reduction in the probability of EoT by 2030 if animals play a role. In 31 health zones, there is considerable uncertainty on animal dynamics (weak evidence for either model) and also a large reduction (of 10 percentage points or more) of the probability of EoT by 2030 in the case that animals do play a role in transmission. The health zones with both uncertainty about the role of animals in transmission and with non-negligible differences in EoT predictions are found across the country, with no discernible geographic pattern.

The detected cases, the detected deaths, and the undetected deaths according to the model are shown in Supplementary Figures 10.

### Step 2 and 3. Epidemiological projections and cost-effectiveness predictions for example health zone: Kikongo

The projections of detected cases and new infections are shown for an example health zone of Kikongo in Supplementary Figure 7, as mentioned above, and full intermediate and final health outcomes are shown in Supplementary Table 25. In Kikongo, the status quo strategy was considered to be *Mean AS*, as the VC from 2019 was scaled back to cover less than 1% of past cases in 2022. *Mean AS* results in 47 [95% PI: 0–197] total cases and 1129 [95% PI: 0–5086] DALYs at a cost of $1.68M [95% PI: 728K–2.76M] (not discounted), which makes it the minimum cost strategy. *Mean AS + Targeted VC* is the only cost-effective strategy, resulting in 29 [95% PI: 0–106] total cases and 696 [95% PI: 0–2669] DALYs, while costing $1.65M [95% PI: 756K–2.73M] (not discounted); therefore, *Mean AS + Targeted VC* dominates the *Int. AS* strategy because it costs less and averts more DALYs. *Mean AS + Targeted VC* therefore results in an ICER of $44/DALYs averted. *Mean AS + Full VC* and *Int. AS + Full VC* have ICERs of $13,839 and $22,072 per DALY averted after discounting costs and DALYs, indicating that these strategies are not cost-effective at WTP thresholds equivalent to values under three times the GDP per capita of DRC. *Int. AS + Targeted VC* is weakly dominated by *Int. AS + Full VC*, because although it averts DALYs at a lower overall cost than *Int. AS + Full VC*, *Int. AS + Targeted VC* averts DALYs at a higher ICER than *Int. AS + Full VC*. A step-by-step explanation of weak dominance can be found in [3] in the Supplementary Methods, page 46.

Accounting for uncertainty in the choice of strategy, *Mean AS + Targeted VC* is optimal in 55% of the iterations at a cost-minimizing WTP (WTP=$0). However, at a WTP=$250 and above, the mean net monetary benefit is highest for *Mean AS + Full VC*, and its probability of being optimal is 45% and higher.

In terms of EoT by 2030, the model predicts that Kikongo has a 37% probability of reaching EoT by 2030 with the status quo strategy of *Mean AS* and with an expected year of elimination of 2035 [95% PI: 2023–After 2057]. The lower expected year of elimination indicates that there is a small chance that EoT has already been reached. With the cost-effective strategy (*Mean AS + Targeted VC*) there is a 53% probability of reaching EoT by 2030, with an expected year of elimination of 2032 [95% PI: 2023–2053]. Implementing the more expensive strategies would not be cost-effective, but it would increase the probability of EoT by 8–19 percentage points over *Mean AS + Targeted VC*, yet no strategy considered in the analysis is predicted to yield a probability of EoT by 2030 of 90%.

Another example is the health zone of Kwamouth is shown in Supplementary Table 26 in Supplementary Section B.4, as this health zone was featured in a previous CEA [3]. Results discussing the special health zones of Ango and Doruma individually can be found in Supplementary Section B.5. Individual results by health zone are displayed in our GUI https://hatmepp.warwick.ac.uk/DRCCEA/v8/.

### Step 3: Optimal strategies for all health zones

The optimal strategy for each health zone is shown in Figure 2 for the status quo, the minimum-cost strategy (WTP=$0), and the strategy that would be optimal at WTP=$250 and WTP=$500, as well as the strategies that maximise EoT by 2030 and 2040. Individual results by health zone are displayed in the “Cost-effectiveness” tab of our GUI https://hatmepp.warwick.ac.uk/DRCCEA/v8/, in addition to results for additional WTP values.

**Fig. 2.**
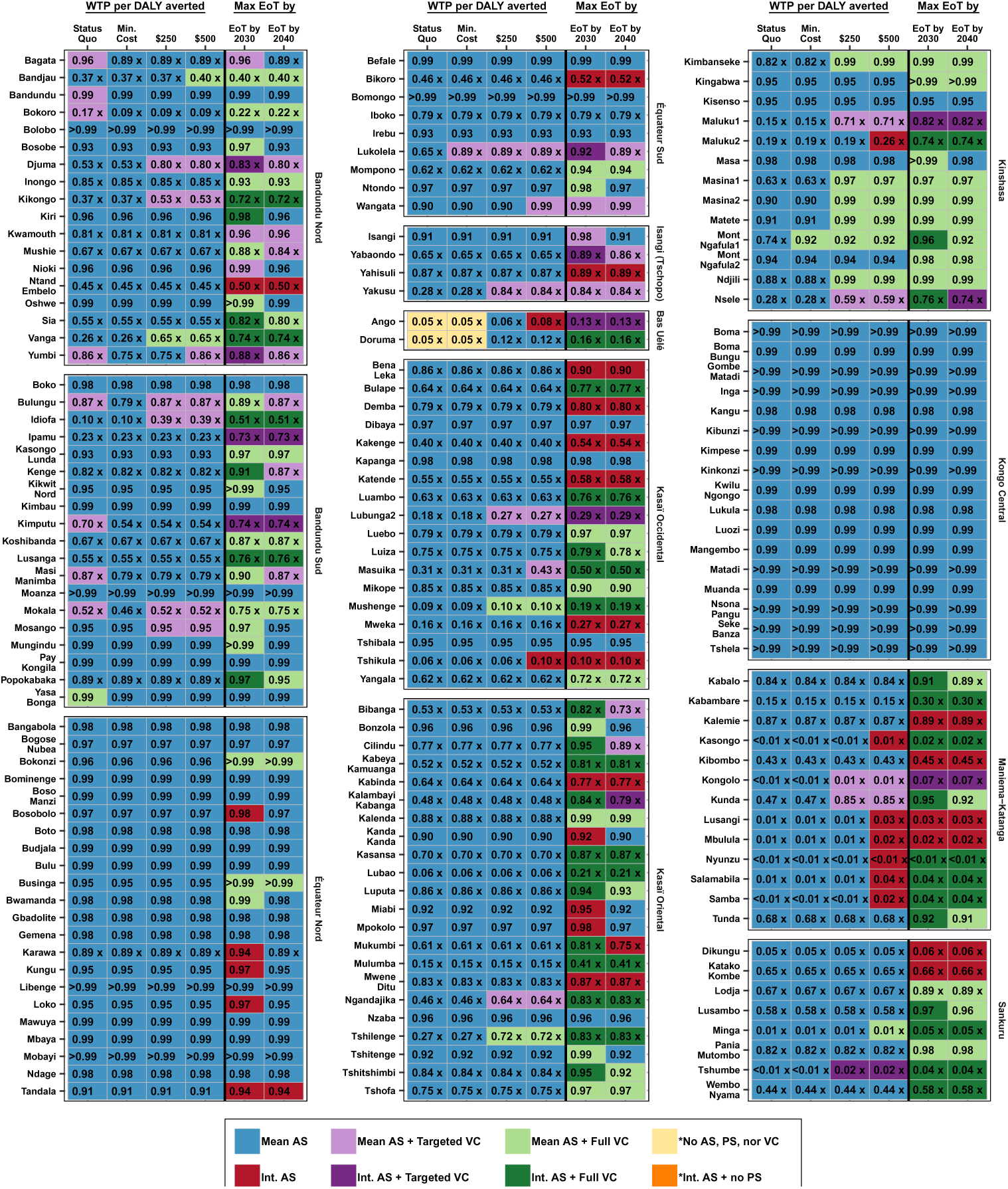
Optimal strategies in each health zone of DRC compared to the status quo strategy (first column) depending on the level of investment (denominated by USD per DALYs averted), or by different target dates to maximize the probability of EoT. Colours represent the optimal strategy, and numbers represent the probability of achieving EoT with that colour strategy by 2030. Strategies that do not reach a 90% probability of achieving EoT by 2030 are marked with an ‘x’. AS: active screening, PS: passive screening, VC: vector control, DALY: disability-adjusted life-year, WTP: willingness-to-pay (denominated in USD per DALY averted), EoT: elimination of transmission.

#### Cost-effective strategies across willingness-to-pay levels

To minimise costs, the analysis indicates that VC should be deployed in two health zones Lukolela in Équateur Sud (with *Mean AS + Targeted VC* and Mont Ngufula 1 in Kinshasa (with *Mean AS + Full VC*) but it does not recommend additional AS anywhere. On the other hand, in Yasa Bonga, where VC has been in place since mid-2015, the analysis suggests that VC should cease to minimise costs with no predicted sacrifice in the probability of elimination by 2030. In Bandundu and Yasa Bonga health zones, where VC has been in place since mid-2019, the analysis suggests that VC should cease in order to minimise costs with no decrease in the probability of achieving EoT by 2030. In Bagata, Bokoro, Yumbi, Bulungu, Kimputu, Masi Manimba, and Mokala, the algorithm would suggest removing VC to save money, but this would decrease the probability of EoT by 2030.

Compared to the minimum cost strategies, the analysis recommends more ambitious strategies in 24 and 37 health zones at WTP=$250 and WTP=$500, respectively. At WTP=$250 the model would recommend the addition of VC in 20 health zones (Djuma, Kikongo, and Vanga in Bandundu Nord; Idiofa and Mosango in Bandundu Sud; Yakusu in the Tshopo region of Isangi; Lubunga 2 and Mushenge in Kasaï Occidental; Ngandajika and Tschilenge in Kasaï Oriental; Kimbanseke, Maluku 1, Masina 1, Masina 2, Matete, Ndjili, and Nsele in Kinshasa; Kolongo and Kunda in Maniema-Katanga; and Tschumbe in Sankuru. At the level of investment equivalent with WTP=$250, the analysis would only recommend the addition of AS in Ango and Doruma (in the Bas Uélé region). In two health zones, Bulungu and Mokala in Bandundu Sud, the status quo is CE at $250 although it is not the minimum cost strategy.

If the strategies optimal at WTP=$500 would be adopted that would constitute making the changes to Lukolela and Mont Ngafula 1 that would minimize costs by 2040; leaving the *Status Quo* in Bulungu and Mokala which are CE at the WTP=$250 and leaving the *Status Quo* strategy in Yumbi (which is neither cost-minimizing nor cost-effective at a lower WTP); as well as adopting the changes recommended at the WTP=$250 level except for two health zones. In Ango in the Bas Uéle region of Isangi, *Int. AS* would be adopted rather than the *Mean AS* that would be adopted at a lower WTP of $250. In 4 further health zones, VC must be added (Bandjau in Bandundu Nord, Wangata in Equateur Sud, Masuika in Kasaï Occidental, and Minga in Sankuru). Furthermore, in 8 health zones, the analysis would recommend intensifying AS (Tshikula in Kasaï Occidental, Maluku 2 in Kinshasha; and Kasongo, Lusangi, Mbulula, Nyunzu, Samabila, and Samba in Maniema-Katanga).

#### Strategies to maximise the probability of EoT

In order to maximize the probability of meeting the EoT goal by 2030, 119 health zones need a change compared to *Status Quo*. None of these changes will be cost-saving, 7 of those changes be CE at WTP=$250 – all but one located in Kinshasa – and an additional 5 changes will be CE at a WTP=$500. The remaining 107 changes will be neither cost-saving nor cost-effective; in other words, the extra cost cannot be justified solely by the additional disease burden averted. Overall, the highest number of health zones where EoT-maximising strategies are not CE are in Kasaï Oriental (21), Kasaï Occidental (14), Bandundu Nord (14), Bandundu Sud (14), and Maniema Katanga (10) coordinations.

Notably, in 66 health zones, the strategy that maximises EoT by 2030 has a *<*90% estimated probability of reaching EoT by 2030. The largest number of those health zones are in Kasaï Occidental (13), Kasaï Oriental (11), and Maniema-Katanga (10), while the rest are in Bandundu Nord (9), Bandundu Sud (7), Bas Uélé region (both health zones), Kinshasa (3), and Sankuru (6). All the health zones in Kongo Central have more than a 90% probability of reaching EoT by 2030 with *Status Quo* strategies, and the same is true for all but the Karawa health zone in Équateur Nord.

The difference between striving for EoT by 2030 versus a more lenient timeline by 2040 is that 20 health zones need less ambitious strategies, and 24 health zones will not need any change at all. Overall, in order to reach EoT by 2040, 94 health zones need a change compared to status quo strategies and 92 health zones would need more ambitious strategies to reach EoT by 2040 compared to the minimum cost strategies. 11 of those changes will not be cost-saving but will be CE at WTP=$250 or WTP=$500; the remaining 81 changes will not be CE at WTP<$500.

#### Geographic distribution of strategies and expected number of health zones reaching EoT

Figure 3 displays the geographic distribution of optimal strategies at different investment levels and summarises the optimal strategy at WTP values of $0, $250, and $500, in addition to the 2030 and 2040 EoT targets. The health zones coloured with dashed lines are those that have *<*90% probability of reaching the 2030 or 2040 goal. We expect that 118, 117, 122, and 123 health zones will reach EoT by 2030 with the *Status Quo*, the minimum cost, and the CE strategies at WTP of $250 or $500 (Figure 3). With strategies meant to maximise the probability of EoT, up to 134 health zones could reach EoT by 2030. However, our analysis suggests that, although 118 health zones would need to switch strategies to maximise the probability of EoT by 2030, because of the probabilistic nature of elimination, the change in strategies would only increase the number of health zones that would reach EoT by 2030 by 16 for a final estimate of 134 health zones (Figure 3).

**Fig. 3.**
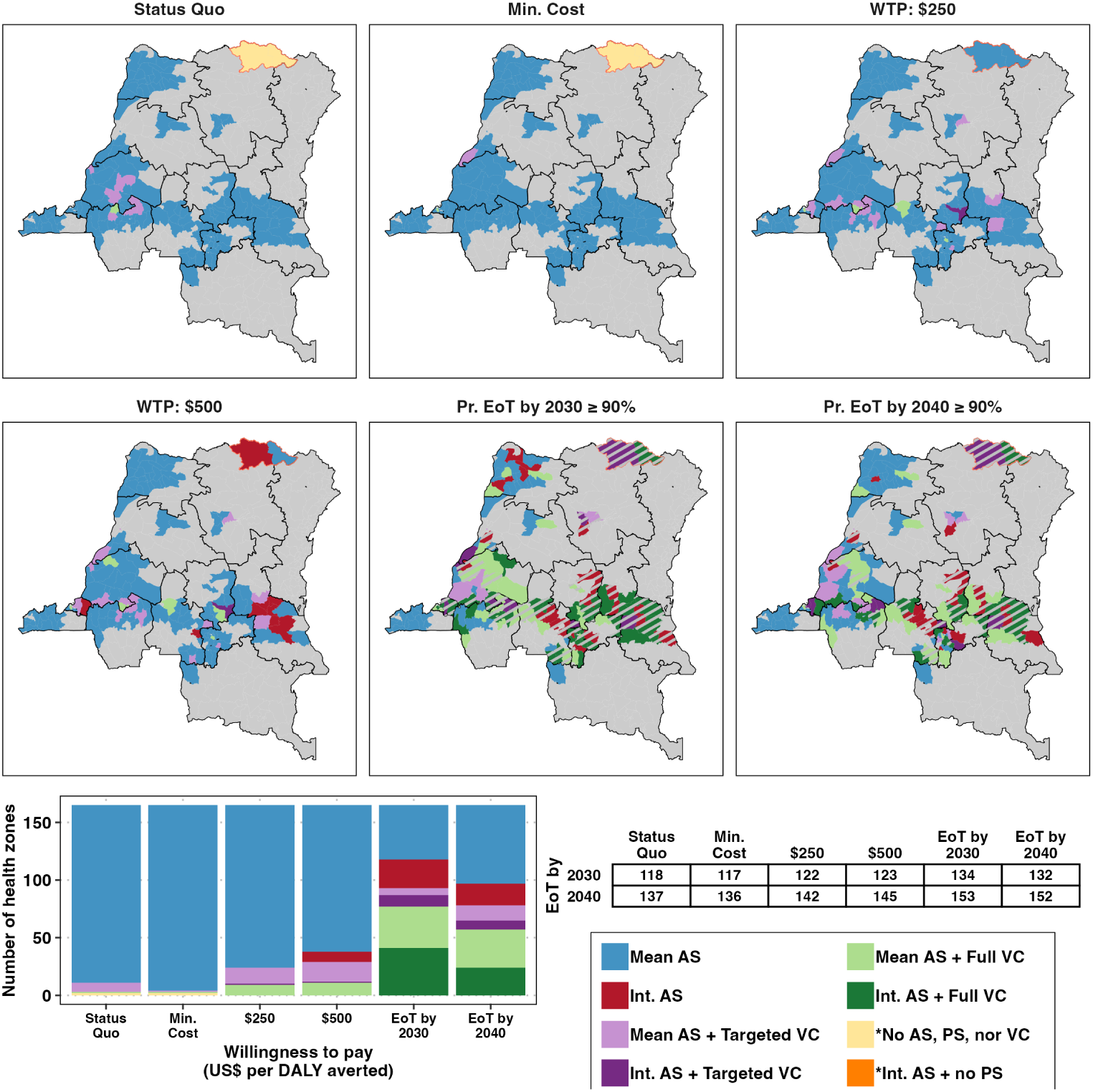
Maps and bar charts of optimal strategies according to economic or elimination goals for the whole of the DRC (time horizon 2026–2040 and 3% discounting, based on models estimated to 2000–2020 data). Maps show the optimal strategies depending on the status quo, minimum cost, and a WTP of $250 and $500, respectively. The final map shows the most efficient strategy with the maximum probability of EoT by 2030 and 2040 in each health zone of the DRC. Striped health zones are those where the probability of EoT by 2030 is less than 90%. Shapefiles used to produce this map were provided by Nicole Hoff and Cyrus Sinai under a CC-BY licence (current versions can be found at https://data.humdata.org/dataset/drc-health-data). The table denotes the expected number of health zones that will meet the EoT goal by either 2030 or 2040 under the different objectives. An interactive map can be found at https://hatmepp.warwick.ac.uk/DRCCEA/v8/. AS: active screening, PS: passive screening, VC: vector control, DALY: disability-adjusted life-year, WTP: willingness-to-pay (denominated in USD per DALY averted), EoT: elimination of transmission.

If EoT were deferred until 2040, 137, 136, and 145 health zones would reach the goal with status quo, minimum-cost, and cost-effective strategies at WTP=$500. With EoT-maximising strategies, 152 health zones are expected to reach EoT by 2040 with *≥* 90% probability. Overall, 31 health zones will not reach EoT by 2030 and 13 health zones are not predicted to reach by 2040 under any approach, and these health zones are primarily concentrated in the coordinations of the east of the DRC (Bas Uélé region, Kasaï Oriental, Maniema-Katanga, and Sankuru).

### Health outcomes and costs at different levels of investment

Figure 4 shows the results by year and aggregated for the period of 2026-2040. Our GUI https://hatmepp.warwick.ac.uk/DRCCEA/v8/ under different assumptions and additional WTP values.

**Fig. 4.**
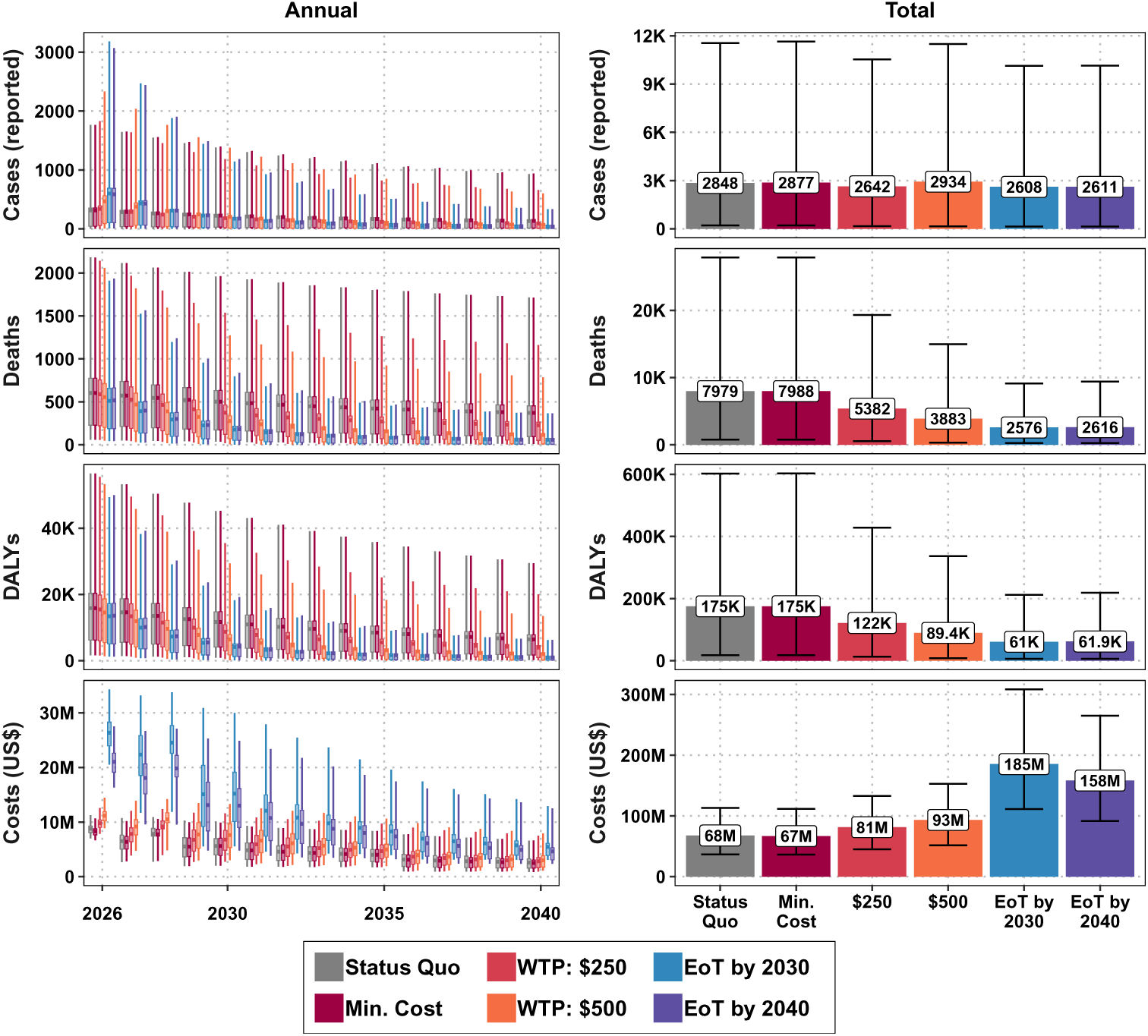
Projected total cases reported, deaths, DALYs and economic costs in all the DRC with optimal strategies at different investment levels over time and for the period of 2026-2040. Values here show numbers of projected cases, deaths, DALYs and costs (undiscounted) over the time horizon 2026–2040 based on fits to 2000–2020 data. DALY: disability-adjusted life-year, WTP: willingness-to-pay (denominated in USD per DALY averted), EoT: elimination of transmission.

#### Status quo

We expect 313 [95% PI: 1–1765] reported cases in 2026 with status quo strategies, in addition to 661 [95% PI: 59–2349] deaths, most of which would be undetected. During 2026-2040 under status quo strategies, we expect 2848 [95% PI: 210–11,547] reported cases, 7979 [95% PI: 770–27,868] deaths or 175K [95% PI: 17.9K–602K] discounted DALYs. We predict 1405 [95% PI: 2–4474] deaths occur in Bas Uélé alone; in other words, 17.6% of all deaths are attributable to a region constituted of two health zones, under-scoring the disproportionate burden in the region (Supplementary Figure 14).

Costs under the status quo strategies will be $8.75M [95% PI: $7.00M-11.3M] in 2026. After accounting for the fact that AS and VC operations will cease in health zones when no cases are reported, and PS clinics will be reduced to 1 in those same health zones 5 years after AS ceases, costs are predicted to amount to $67.8M [95% PI: $36.7M-113M] by 2040 (undiscounted).

#### Optimising for cost-effectiveness

Under minimum-cost strategies, reported cases are expected to decrease, amounting to 2877 [95% PI: 210–11,643 in 2026–2040. Under strategies consistent with WTP=$250 and WTP=$500, we estimate that by 2040 there will be 2579 [95% PI: 152–10385] and 2889 [95% PI: 159–11,274] cases reported, respectively.

Strategies consistent with higher WTP would lead to lower numbers of deaths and DALYs (Figures 4). Minimum-cost and CE strategies at WTP=$250 and WTP=$500 will yield deaths totalling 7988 [95% PI: 770–27,879], 5504 [95% PI: 583–19,571], 4076 [95% PI: 3322–15,547] by 2040, respectively, and will yield discounted DALYs totalling 175K [95% PI: 17.9K–603K], 124K [95% PI: 13.9K–434K], 93K [95% PI: 8.23K–348K] by 2040, respectively.

In 2026, minimum-cost strategies will cost $8.10M [95% PI: $6.52M–10.4M] compared to *Status Quo* strategies that will cost $8.56M [95% PI: $6.86M–11.0M], and by 2040, minimum-cost strategies will cost $64.5M [95% PI: $34.7M–108M], or $1M less than the *Status Quo*. Under strategies consistent with WTP=$250 and WTP=$500, the total cost will be $81.4M [95% PI: $45.0M–133M] and $93.3M [95% PI: $51.6M–153M], respectively.

#### Optimising for probability of EoT by 2030 and by 2040

Strategies chosen to maximise the probability of EoT by 2030 would bring cases reported to 2608 [95% PI: 146–10,136] and will bring deaths down to 2576 [95% PI: 255–9133], and burden down to 60.9K [95% PI: 6.42K–212K] DALYs, at a cost of $196M [95% PI: $118M– 326M]. Compared to the status quo, EoT by 2030 would yield 241 fewer reported cases while averting 5403 deaths and 114K DALYs for an additional cost of $118M, equalling a cost of $185M [95% PI: $111M–$309] by 2040.

Notably, however, the additional costs will have to be front-loaded; in the year 2026, the status quo will cost $8.75M [95% PI: $7.01M–11.2M] while EoT-maximising strategies would cost $27.0M [95% PI: $21.0M–35.2M] and 21.7M [95% PI: 16.8M–28.4M], depending on whether the goal is to reach EoT by 2030 or 2040, respectively.

Deferring EoT until 2040 saves only $27.1M and will accrue 2611 [95% PI: 146–10,141] reported cases while incurring 2616 [95% PI: 255–9412] deaths and 61.9K [95% PI: 6.42K– 218K] DALYs at a cost of $158M [95% PI: $91.6M–265M] by 2040. Compared to EoT by 2030, EoT by 2040 strategies would incur 3 more cases reported, 39 more deaths, and 913 more DALYs.

#### Health outcomes and costs in different coordinations

All results are available stratified by coordination in Supplementary Figures 12-19. Our GUI https://hatmepp.warwick.ac.uk/DRCCEA/v8/ shows results for health zones individually, as well as aggregated by coordinations and provinces, under different assumptions and additional WTP values. Expected case trends throughout the country show a decline even with status quo or cost-minimizing strategies, with the notable exception of the Bas Uélé region, which will reap the largest health benefits if more ambitious strategies are implemented.

### Breakdown of costs and resources needed

#### By coordination

Most coordinations are expected to soon need an infusion of resources to meet the EoT goal by 2030 or 2040, with the notable exception of Kongo Central (Supplementary Figures 19). The largest portion of the costs at any level of investment will go to AS followed by VC activities if EoT is to be achieved (see Supplementary Figures 20). For minimum-cost or cost-effective strategies, PS costs almost equal AS costs, and VC costs are small. The distribution differs substantially by coordination (see Supplementary Figures 21). An effort to maximise EoT by 2030 would mean costs will be incurred primarily in Kasaï Oriental, Maniema-Katanga, and Bandundu Nord.

#### Resources needs

We have estimated the number of drugs and screening tests that will be required (Supplementary Figures 22–23). NECT and fexinidazole will continue to figure prominently in treatment until a new drug, acoziborole, is approved, needing an expected 144 doses of NECT, 172 doses of fexinidazole, and 6 doses of pentamidine (for those too young for fexinidazole) in 2026 under status quo strategies. Also in status quo, 2.34M tests will be needed for AS and 235K tests will be needed for PS.

## Discussion

This is the first analysis considering the cost-effectiveness of country-wide elimination of gHAT using location-specific health zone data in the DRC, the country with the highest burden in the world. We find that current strategies are on track to eliminate the disease by 2030 in 118 health zones, and that up to 134 health zones could reach elimination of transmission by 2030 with the right strategies. Reaching elimination of transmission by 2030 would reduce deaths expected in DRC by 68%, from 7979 [95% PI: 770–27,868] with status quo strategies to 2576 [95% PI: 255–9133]. Our analysis also outlines the costs and consequences of delaying EoT until 2040.

The findings suggest that strategies likely to achieve EoT by 2030 will require an appreciable increase in economic resources. While status quo strategies have an expected mean cost of $8.75M in 2026, strategies to reach EoT have an expected mean cost of $27.0M in 2026, more than tripling the costs. These expanded strategies are predicted to have the greatest impact in the eastern coordinations, where limited activities have occurred in recent years and uncertainty in current transmission levels favours the adoption of more intense strategies. While delaying the EoT target until 2040 could save an estimated $27.1M compared to implementing activities aimed at achieving elimination by 2030, it is crucial to consider potential unforeseen events in the extra decade that may undermine the political will or operational feasibility to sustain these activities.

Compared to previous modelling studies [14, 23], this analysis demonstrates a more pessimistic outlook regarding EoT. Despite now having more data on cases from recent AS and PS that display an optimistic downward trend, our new reservations on the probability of EoT by 2030 arise from several factors related to modelling and analysis changes which we believe better capture the uncertainty in the predictions: a) previous analyses were performed with a deterministic model, while this analysis is performed with a stochastic model, which makes us less optimistic when aiming for a high probability of EoT, b) our more refined estimates of the potential impact of VC based on case and river proximity, and c) 2024–2030 is a shorter time horizon than 2020–2030 – the previous time horizon in other analyses. Especially concerning the importance of timing, our present analysis shows how the proximity to 2030 is becoming a hindrance – with strategies aimed at maximising the probability of EoT by 2030, 134 health zones are expected to reach EoT by 2030, and an additional 19 health zones are forecast to reach EoT by 2040.

Operational feasibility emerges as a major challenge. Although effective tools exist and there is potential for increased donor funding to support expanded activities, the convergence of regions requiring new strategies with issues of inaccessibility and/or civil unrest presents a notable hurdle. Rapidly scaling up trained personnel to carry out these activities remains a crucial operational challenge that must be addressed. Whilst the treatment costs are limited, timely access to treatment is pivotal for strategies to remain cost-effective, as treatment is the mechanism through which deaths and DALYs are averted and through which further cases are prevented in the health zones where no VC is implemented.

Additionally, this analysis confirms previous results, showing that VC could play a pivotal role in achieving elimination, as it is the key determinant in reducing the tsetse population. To alleviate the burden of disease, strengthening AS efforts is beneficial in the short term by identifying and treating individuals currently infected and ill, even if they are unlikely to transmit the infection whilst the tsetse population is suppressed via VC.

### Limitations

This study has limitations inherent to its scope and methodology, categorised into three main areas:

#### 1. Past data

Whilst we account for the underreporting of cases, the study acknowledges that the mechanistic models cannot extrapolate to regions with no or minimal historical case data due to insufficient surveillance. Alternative approaches (e.g. geostatistical modelling using satellite imagery) may offer better quantitative support for the absence of suitable tsetse habitat and gHAT transmission. In the Bas Uélé region, there are challenges in data interpretation as alternative screening algorithms were performed by MSF during 2007–2014 followed by no subsequent surveillance, leading to large uncertainty in modelled results. Exploratory screening in regions with little or no recent screening data could help refine model predictions, however, the strength of data to reduce uncertainty will be linked to the number of people screened.

#### 2. Future strategies

The study assumes the perfect application of tools and robust quality assurance in the future. In addition to video confirmation of case diagnosis, which has been gradually rolled out in the DRC since 2015, further maintenance and development of the quality assurance system will be needed. For treatment, all diagnosed cases were simulated to receive medication promptly, requiring sufficient stocks near where cases are diagnosed; we did not simulate treatment with acoziborole which we hope will be available in the near future. If the same algorithm is used for diagnosis, we would expect projected total treatments and transmission dynamics to remain the same if acoziborole becomes available. Operational feasibility is identified as a major hurdle, with coordinations such as Sankuru, Maniema-Katanga, and the Bas Uélé region posing significant challenges in terms of accessibility and security. These factors can impact the implementation of interventions and may result in overambitious projections for these regions. Throughout the country emphasis should be placed on the importance of population turnout in active screening to improve the efficiency of this intervention [41], particularly as community knowledge wanes with the increasing rarity of the disease; the model currently accounts for continued AS turn out based on past data. Other interventions that are under consideration for the future but do not feature in the present analysis are the community-based deployment of VC targets [50–52], a “screen-and-treat” intervention that would circumvent confirmation and staging if the new acoziborole treatment is approved and deemed suitably safe [28], and new laboratory-based diagnostics [21, 40]. These future strategies should be considered in future simulation studies, particularly when information on implementation algorithms for screen-and-treat and roll-out dates for acoziborole can be better defined.

#### 3. Model formulation and use

The analysis does not consider the cost of end-game diagnostics with higher specificity, which could improve the positive predictive and negative predictive value at low prevalence. As this diagnostic would be present in all possible strategies to the same degree, it is not expected to influence the selection of optimal strategies but would slightly affect the economic cost estimation. Likewise, post-elimination surveillance is not explicitly modelled, although we assume that PS will continue at reduced levels. One potential issue is treatment completion, as there have been instances where treatment access was *<*100% due to logistical issues or delays, which can impact the model fitting process and potentially contribute to ongoing transmission and disease burden. Resolving delayed or under-treatment in the future, however, is a matter of organisational capacity between pharmaceutical companies, the WHO, and the country programme. Ensuring sufficient spare stock at the right points in the supply chain is not considered in this analysis, however, this modelling work could be adapted to support stock planning.

Future work will also encompass modelling at smaller spatial scales as, although analysis at the health-zone level aligns with the scale at which elimination as a public health problem is assessed for a country, health areas align more closely with operational decisions made for interventions. Health-area modelling requires fitting to around 1,200 regions rather than the 165 regions for a health-zone analysis. Whilst this is the goal of future modelling work and the proof-of-principle for the modelling methodology has been demonstrated [16], it is beyond the scope of the present analysis to expand to this granularity. Overall, we believe our health zone-modelled trends provide very similar outputs to modelling at a smaller scale and aggregating [16]. In this regard, our predictions are potentially conservative, as the model assumes larger operations will remain in place longer than would probably be the case in reality. A cost-effectiveness analysis at the health-area level might, therefore, be more optimistic as additional interventions could be targeted at a smaller scale.

## Conclusion

This manuscript presents a comprehensive analysis evaluating the feasibility and cost-effectiveness of achieving the elimination of gHAT in the DRC. Despite operational challenges, this study presents a compelling case for the sensible utilisation of resources in support of gHAT elimination of transmission by 2030. This analysis reveals that, overall, the EoT of gHAT by 2030 appears to be epidemiologically feasible with the existing toolkit in a majority of health zones of DRC. Overall, 119 health zones require strategy modifications to maximise the probability of EoT by 2030 (Figure 2), in 12 health zones, those changes will be cost-effective. To reach EoT by 2040, 94 health zones require strategy modifications, where 13 health zones would make changes that are cost-saving or cost-effective, and the savings compared to aiming for EoT by 2030 would amount to approximately 15% of the costs. We show that substantial investments aimed at achieving EoT do not need to be sustained for more than a decade. By targeting high-incidence areas and implementing appropriate interventions, such as VC and intensified AS strategies, major progress can be made towards eliminating gHAT transmission, ultimately leading to substantial improvements in public health.

## Declarations

### Ethics approval and consent to participate

Ethics approval was granted by the University of Warwick Biomedical and Scientific Research Ethics Committee (application number BSREC 80/21-22) to use the previously collected DRC country HAT data, provided through the framework of the WHO HAT Atlas [19], in this secondary modelling analysis. No new data collection took place within the scope of this modelling study.

### Clinical Trial

Not applicable.

### Consent for Publication

Not applicable.

### Availability of data and material

Information about the WHO HAT Atlas data used for fitting is described in the Supplementary Information. Data cannot be shared publicly because they were aggregated from the World Health Organisation’s HAT Atlas which is under the stewardship of the WHO; our data-sharing agreement does not allow us to share that data. WHO HAT Atlas data include identifiable data. Data are available from the WHO (contact or visit https://www.who.int/health-topics/human-african-trypanosomiasis/) for researchers who meet the criteria for access to confidential data, including secure computational facilities and an existing relationship to the national sleeping control program of the DRC. The time frame for response would depend on the WHO’s timelines and workloads. Clinical outcomes and costs (listed in Table 1) were simulated using estimates from the literature and are described in Supplementary Note 3: Parameter Glossary. Assumptions and estimates were parameterised according to conventions in the economic evaluation literature [10]. All modelled outputs and code are available via Open Science Framework: https://osf.io/ezjxb/.

## Author’s contributions

MA, SAS, CH, REC and KSR developed the software and performed the analyses. MA and PEB visualised the results. EMM, JL, CS, SAS, PRB and RS curated the data. REC, SAS, CH and MA analysed the data. MA, RS, SAS, PRB, REC, and KSR developed the methods. MA, KSR, EHC and FT wrote the original draft. MA, KSR and FT conceptualised the study. AH, IT, SD, PV read, reviewed and gave substantial comments on the draft. All authors read, reviewed, and approved the final version for publication.

## Acknowledgements

The authors thank PNLTHA for original data collection, WHO for data access (in the framework of the WHO HAT Atlas [19]), Cyrus Sinai and Nicole Hoff from UCLA Fielding School of Public Health for providing health zone level shape files (current versions can be found at https://data.humdata.org/dataset/drc-health-data). Calculations were performed using the sci-CORE (http://scicore.unibas.ch/) Scientific Computing Center at the University of Basel. The authors also thank Steve Torr and Alex Shaw for their valuable insights on specific topics in this article. For the purpose of open access, the authors have applied a Creative Commons Attribution (CC-BY 4.0) licence to any Author Accepted Manuscript version arising from this submission.

