## Supplement for "Cost-effectiveness of end-game strategies against sleeping sickness across the Democratic Republic of Congo"

**C Supplementary Note 1: Glossary of Technical Terms** **74**

**D Supplementary Note 2: Health Zone-Specific Parameters** **75**

**E Supplementary Note 3: Parameter Glossary** **79**

**F Supplementary Note 4: NTD PRIME Criteria** **104**

**G Supplementary Note 5: CHEERS Checklist** **105**

**Supplementary References** **110**

### A Supplementary Methods

#### A.1 Key updates to the modelling in this study

↩ Return to the Supplement Table of Contents

##### Data extraction

We now have four more years of data (covering 2000–2020) compared to previous modelling studies for the DRC (2000–2016) [1, 2]. In the current data extraction, we have more places which have "unknown" active screening (AS) numbers. This occurs where the apparent prevalence in AS was >10% or the recorded number screened was <20. In previous studies [1, 2], we assumed that these new cases arose from people being recorded in their home health zone, although they were screened in a neighbouring health zone. We still believe this assumption to be appropriate, but we set the number of people screened in those "home health zones" to be "missing", which allowed us to impute the number of people that would have to be tested in that health zone, given the current prevalence, to generate the number of new active cases that was recorded. Data extraction is further described in Section A.2 of these Supplementary Methods.

The Bas-Uélé region of Isangi coordination comprises the health zones of Ango, Bili, Doruma, Ganga, Poko and Titule. Of these, only Ango and Doruma had sufficient data points in the final data set to be analysed. Médecins Sans Frontières (MSF) were active in the Bas-Uélé region from 2008–2014. Some modifications were made to the aggregated data in this region:

1. Based on information from an MSF report [3], data from Ango and Ganga in 2014 were modified to include only confirmed cases, with an assumed specificity of 100%.
2. Implausibly high numbers of cases were detected from passive screening in Ango (in 2008) and Ganga (in 2010 and 2011), we assumed that these arose from widespread screening by MSF within the health centres (effectively active screening) and consequently transferred them to the active screening data. This also required the number of people screened in those HZ–year combinations to be estimated within the model fitting process.
3. In early 2024, exploratory screening was performed in the Ango health zone. Information received from this screening indicated that no staging had taken place as part of MSF's activities. In light of this, staging information was removed from the data in the Bas-Uélé region.

##### Fitting assumptions and process

4. **Endemic Equilibrium.** We now assume no activity (and in particular no PS improvement) before the first detection year in all health zones where the first detection is after 2000. In all other health zones with detections in 2000, we assume some step improvement to PS in 1998 in line with our previous analyses [1, 2].
    4.1. In Kasaï Occidental and Kasaï Oriental coordination, the model assumed transmission remained at endemic equilibrium until the year when cases were first reported. This was because of a lack of accessibility preventing interventions from taking place in the early 2000s.
    4.2. In the Bas-Uélé region of Isangi coordination, passive screening (PS) began in 2004. As this region is difficult to access and hence no interventions were taking place in the early 2000s, we assumed that transmission was in equilibrium until 2004 instead of 1998.
5. **Specificity in AS.** Starting from the second published "Warwick gHAT model" [4] it was assumed that AS diagnostic specificity would increase to 100% due to additional cross-checking of confirmed cases at very low prevalence. A somewhat arbitrary threshold "fewer than 2 reported cases per 10,000 above the expected incidence of false positives based on the level of screening" was used in previous analyses if it was not clear when this might happen. More recently, the rollout of video confirmation, starting in Bandundu Nord and Sud coordinations, has provided a mechanism for this to happen. In this present study, we therefore assume fixed years when specificity would increase to 100% rather than using the previous threshold; this is 2015 in Yasa Bonga and Mosango health zones, 2018 for the remainder of Bandundu Nord and Sud coordinations, 2018 for Kongo Central and Kinshasa coordinations, and 2024 elsewhere.
    5.1. In previous modelling studies in the DRC [1, 2], we assumed that MSF were active in the health zones in the Bas-Uélé region (including in Ango and Doruma) of Isangi coordination from 2008 until 2012, and the PNLTHA was active for two years in 2013 and 2014. As a result of updated information, in this study MSF was assumed to be active from 2008 until 2014, with no AS by PNLTHA in this region. Two AS diagnostic algorithm

specificity values were fitted: for MSF and PNLTHA activities, with the MSF specificity being lower. In this study, the specificity of MSF and PNLTHA activity are included in the fitting, however values for the PNLTHA specificity – higher than that of the MSF specificity – result from the prior distribution (and were included solely to provide variable estimates for use in future projections). In 2014, MSF changed their diagnostic algorithm in Ango and Ganga and although some unconfirmed cases were reported (removed from the data used here), the specificity for the analysed (confirmed) cases in these HZs was taken to be 100%.

6. **Passive screening**

6.1. We now include gradual improvements in passive detection (from the start of activities to 2020) in more coordinations, expanding from Bandundu Nord, Bandundu Sud and Kongo Central coordinations in previous studies [1, 2], and now adding in Équateur Nord, Équateur Sud, Kinshasa, Kasaï Oriental.

6.2. Since 2014, there has been no AS and no PS (no tests available) in the Bas Uélé region and PS has been simulated as absent in the model to reflect this.

6.3. We are now using a smaller overdispersion value for passive case observations, which results in less predicted variation in case reporting. This applies to all the coordinations except for the Bas Uélé region, where we keep the previous overdispersion value (and have more uncertainty).

#### Projections

7. This study includes more granular details about vector control (VC), particularly for the simulation of partial coverage of health zones with Tiny Target deployments (both past and future).
8. We now use a stochastic model for doing projections which tracks integer numbers of infections and is better at capturing the distribution in elimination times as it has a clearly defined time when zero is reached.
9. In our GUI (https://hatmepp.warwick.ac.uk/DRCCEA/v8/ we now show aggregations for coordinations or the whole country as well as individual health zones. Aggregations are computed by summing up outcomes across different health zones based on the optimal strategy in each health zone to achieve the specific objective – e.g. not all health zones would have the same optimal strategy to achieve the EoT by 2030 objective.
10. In previous studies for the DRC we have often used only one model [1, 5, 6]. Here our default outputs are from an “ensemble” model which includes outputs from the models with and without transmission from animals based on the weighted model evidence for each health zone, as done in Crump et al. [2].

#### A.2 New data extraction

↩ Return to the Supplement Table of Contents

The methods presented in this section are adapted from the previous DRC fitting study by Crump et al. [1] which used data from 2000–2016. Here we extend the same method, producing both a health-zone level and health-area level extraction to allow more flexibility for future work, and also to include data from 2000–2020.

##### A.2.1 Data

**HAT Atlas data** The DRC WHO HAT Atlas data [7] were provided in a spreadsheet with rows representing case reports or screening events. Each record contains columns for the number of people screened, the number of cases reported at each stage, the type of screening method (active or passive), and the year. Each record also contains multiple fields describing the location: Province, Health Zone, Health Area, Location name, Neighbourhood, Quarter, Sector, Group, District, Territory, and a geolocation (latitude and longitude). In total, there were 153,876 records; of which 146,256 (95.0%) had their geolocation field populated. Note that health areas should be contained within exactly one health zone which is also contained in exactly one province and one coordination. Coordinations are not necessarily contained within one province, and vice versa.

Entries for PS with no cases detected, and entries for AS where both the number of people screened and the number of cases detected are either zero or missing, were dropped from the dataset. After removing these records, we are left with 125,045 case and screening records (119,669 of which had geolocation data, 95.7%). Among these entries, there were 24,568 unique combinations of values in the geolocation, province, health zone, health area, location and territory fields, of which 21,575 had geolocation information and 2,993 did not.

| Recorded region identifiers: | | | Number |
|---|---|---|---|
| Province | Health zone | Health area | n |
| ✓ | ✓ | ✓ | 120597 |
| ✓ | ✓ | | 3213 |
| ✓ | | ✓ | 14 |
| ✓ | | | 1221 |

**Supplementary Table 1**: Number of WHO HAT Atlas records with different combinations of the province, health zone and health area recorded.

**Shapefiles** To match geolocations to health zones (HZs) and health areas (HAs), we used two shapefiles provided by the American Red Cross (ARC) [8]. The first of these is a map of HZ boundaries, with 519 records covering the whole DRC. The second is a map of HAs covering most of the country (all the DRC except Lualaba and Kasaï-Central provinces, most of Sankuru and Nord-Kivu provinces, portions of the north of former Équateur province, and Poko and Monkoto HZs). Both shapefiles provided fields stating the name of the province and HZ, and, in the case of the area map, also the HA. The province and HZ information in both shape files were used to augment the data with the coordination. The ARC HA file has more granular boundaries (HAs are smaller than HZs), so wherever possible we used the HZ and HA data from this file, however, it does not cover the whole of the DRC. The ARC HZ file, on the other hand, has slightly less granular boundaries and some inconsistencies with the HA shapefile in places; by comparing both shapefiles to maps with rivers (which are often used as boundary definitions) it is assumed that the HA shapefile is also more accurate. Nevertheless, the HZ shapefile is complete and covers the whole country. We also used the combined information from these two shapefiles to identify smaller gaps in HZs which are partially mapped on the HA shapefile. In particular, any contiguous gaps of at least 50km$^2$ in the HA shapefile which fell into the same zone in the HZ file were combined into pseudo-HAs and numbered to catch the cases that fell into unmapped HAs.

**Additional geographic information** The following geographical information sourced from the Humanitarian Data Exchange was also used [8]:

- a HZ shapefile from the United Nations Office for the Coordination of Humanitarian Affairs (OCHA);

- an OCHA file of geolocations of localities; and
- a file of geolocations of health facilities from the Global Healthsite Mapping Project.

These data were used to assist in matching and locating the gHAT data, by providing alternative spellings of names and potential geolocations for non-geolocated gHAT locations. The locality and health facility lists were concatenated, and this enlarged locality set and the OCHA HZ map were assigned geographical identifiers as per our shapefile of choice.

**Matching HAT Atlas records to DRC shapefile** Similarly to [1], the names of all locations in both the data and the shapefiles were normalised to facilitate matching between different data sources. The normalisation process consisted of removing any diacritics, converting all names to lowercase, trimming superfluous whitespace, replacing Roman numerals with Arabic numerals, and normalising some spellings by removing leading ‘m’, ‘n’, or ‘g’ when followed by a consonant, and replacing leading ‘ts’ with ‘s’. Additionally, some alternative names and spellings were manually added as they were noticed.

Location records were then placed on the two shapefiles provided by the ARC, and the names of the matching HAs were stored. Locations were then matched in a series of stages, starting with the most precise matches and slowly becoming more permissive until the majority of data points had been matched.

1. Locations with known HZ and HA, which match a HA on the ARC HA shapefile, and where the recorded HZ and HA match the geolocated HZ and HA on the ARC HA shapefile. These locations were matched to the recorded HZ and HA. This matched 4,441 (34.5% of records with geolocation) locations.
2. Locations with known HZ and HA, which match a HA on the ARC HA shapefile, where the recorded HZ matches the geolocated HZ, and where the geolocation is within 5km of the recorded HA on the ARC HA shapefile. These locations were matched to the recorded HZ and HA. This matched a further 2,117 (9.8% of records with geolocation) locations. [Cumulatively: 9,558 or 44.3%]
3. Next, we used the location information. The long list of locations provided by the additional data sources was placed onto the same ARC shapefiles. Data records were then matched to any locations on this list with matching location and territory records (after normalisation), and when the record geolocation was within 10km of the geolocation associated with the location. Again, we only consider records where the recorded HZ matched the geolocated HZ on the ARC HA shapefile. This then gives three possible candidate HAs: the one stated in the record, the one matched on the HA shapefile, and the one matched by location. If two of these agreed we used this value, however, if all three still disagreed then we chose to trust the HA matched with by geolocation. These records were matched to the recorded HZ and the HA as described above. This matched a further 662 (3.1% of records with geolocation) locations. [Cumulatively: 10,220 or 47.4%]
4. The remaining records which match a HA on the ARC HA shapefile, and where the recorded HZ and geolocated HZ on the HA map agree, were matched according to their geolocation. The records were matched to the recorded HZ and the geolocated HA according to the ARC HA shapefile. This matched a further 5462 (25.3% of records with geolocation) locations. [Cumulatively: 15,682 or 72.7%]
5. Now we consider records where the recorded HZ does not match the geolocated HZ but is nearby. Similarly to step 2, we consider records where the geolocation falls within 5km of the recorded HZ according to the ARC HA shapefile. We then perform three matching steps analogous to steps 1–3 for these close HZ matches.
    - We then consider records where the recorded HA matches the geolocated HA. These records are matched to the recorded HZ and the recorded HA. This matched a further 74 (0.3% of records with geolocation) locations. [Cumulatively: 15,756 or 73.0%]
    - We then consider records where the geolocation is within 5km of the recorded HA according to the ARC HA shapefile. These records are matched to the recorded HZ and the recorded HA. This matched a further 446 (2.1% of records with geolocation) locations. [Cumulatively: 16,202 or 75.1%]
    - We then consider records where the location matches a nearby location as in step 3 above. To match the HZ for these records, we apply the following steps. If the record has entries for both HZ and HA, we match to the recorded HZ. If not, we check whether the record’s geolocation matched an area present on

the ARC HA shapefile. If so, we match to the geolocated HZ using the ARC HA shapefile, if not we use the ARC HZ shapefile. To match the HA, then as above, if the recorded HA and the HA from the locality agree then we use this, otherwise we fall back to the geolocated HA according to the ARC HA shapefile. This matched a further 54 (0.3% of records with geolocation) locations. [Cumulatively: 16,256 or 75.3%]

6. After mapping all the records where the recorded health zone matched the ARC HA shapefile, we were left with the records which either fell in the gaps between health zone polygons or have inconsistent data.
   - First we considered locations where the recorded health zone matched the ARC HZ shapefile, and where they fall into an area which is not mapped on the ARC HA file. These records are matched to the recorded HZ and to the appropriate pseudo-HA based on their geolocation. This matched a further 2635 (12.2% of records with geolocation) locations. [Cumulatively: 18,891 or 87.6%]
   - Now we consider locations where the geolocation is within 5km of the recorded health zone according to the ARC HZ shapefile, and where the location does not map onto the ARC HA shapefile. These records are also matched to the recorded health zone. To assign a health area, we find the nearest health area (or pseudo-HA) which falls in the assigned health zone (according to its boundary in the ARC HZ shapefile), and use this. This matched a further 292 (1.4% of records with geolocation) locations. [Cumulatively: 19,183 or 88.9%]
7. For locations with inconsistent data:
   - Firstly, locations where the recorded health area matches the geolocated health area according to the ARC HA shapefile. For these, we used the recorded health area and used the geolocated health zone according to the ARC HA shapefile (to match the health area). This matched a further 192 (0.9% of records with geolocation) locations. [Cumulatively: 19,375 or 89.8%]
   - Next, we match to the locality list and find any locations where the locality and territory match and the geolocations are within 5km of each other. If the health area of the locality according to the ARC HA shapefile matches either the recorded HA or the geolocation HA then we select that as the health area. Note that this includes the possibility of the geolocation and locality both matching the same pseudo-HA. To match the health zone, we use the health zone of the health area (or pseudo-HA) we have just assigned it to, either from the ARC HA map if we matched a real health area, or from the ARC HZ map if the location is not on the ARC HA shapefile. This matched a further 196 (0.5% of records with geolocation) locations. [Cumulatively: 19,471 or 90.2%]
8. Finally, if we cannot reconcile the recorded data with the geolocations, on the advice of the WHO we accept the geolocations as the more authoritative source, matching the health area (or pseudo-HA) according to the ARC HA shapefile, and matching the health zone according to the ARC HA shapefile if possible, or the ARC HZ shapefile if not. This matched the final 2,104 locations with geolocation data present, giving a total of 21,575.
9. For records with no geolocation data available:
   - First, we try to match the locality data as above, matching the locality and territory columns. For any results with exactly one match, we use the geolocation from this locality to assign a health area or pseudo-HA from the ARC HA shapefile, and health zone from the same shapefile if possible or from the ARC HZ shapefile if not. This matched 49 (1.6% of records without geolocation) locations.
   - Now, we instead match by location (but not territory) and health zone, again keeping any unique matches and using the geolocation fromt he territory. This matched a further 11 (0.4% of records without geolocation) locations. [Cumulatively: 60 or 2.0%]
   - Now, we match by location and province. We match any locations from the location list with the same (normalised) name, and where the recorded province in the data record matches any of the former province name, province name, or co-ordination name of the location according to either of the ARC shapefiles. Again, we filter for unique matches and match according to the geolocation of the matched location. This matched a further 19 (0.6% of records without geolocation) locations. [Cumulatively: 79 or 2.6%]

10. Finally, we matched the remaining records to the health zone and health area polygon according to the health zone or health area stated in WHO HAT Atlas. This allowed us to match 2,710 records (90.5% of records without a specific geolocation), giving a final cumulative total of 2,789 matched ungeolocated records, making 93.2% of non-geolocated records or 99.2% of all records. The remaining records (totalling 316 cases and 193,035 people screened) were missing the majority of location fields and were deemed unable to be matched, so were not included in our data.

**Missing screening data** In some HZs there are instances of years of ASs with either very high (or impossible) prevalences (e.g. >10% or with active case numbers exceeding the number of people recorded as having been screened) or where the total number of people actively screened is extremely low (i.e. <20 people). This occurs in 56 HZs for 1 or more years. For these screenings we denote the number of people screened as "unknown" and this is later inferred during model fitting (see A.5) as has been done in previous studies.

##### A.2.2 Post-processing of data in Bas-Uélé region health zones.

Médecins Sans Frontières (MSF) were active in the Bas-Uélé region from 2007–2014, inclusive. This region comprises the Ango, Bili, Doruma, Ganga, Poko and Titule health zones. Based on information received from PNLTHA, patterns in the data, and a report by MSF [3] we modified the extracted and aggregated data for these health zones as follows:

1. All cases detected by passive screening in Ango in 2008 and Ganga in 2010 and 2011 were transferred to the active screening results. Information received indicated that active screening was effectively carried out in the medical facilities in these health zones, and disproportionately high numbers of PS cases were recorded in these years.

2. Information was received from PNLTHA that no staging was carried out in the Bas Uélé region. Therefore, all historical case records were treated as having an unknown stage regardless of whether they were recorded in active or passive screening.

3. In 2014, MSF active screenings in Ango and Ganga were more rigorous, with parasitological confirmation of some cases. However, the serological positives were still recorded as cases in data sent to WHO; the HAT Atlas does not have the split of parasitological or serological cases available. In our analysis, the 2014 active screening data in Ango and Ganga was modified to include only parasitologically confirmed cases as detailed in a report by MSF[3], still without staging information:

| Health zone | Ango | Ganga |
|---|---|---|
| Number of AS parasitological cases in 2014 | 25 | 36 |
| Number of people actively screened in 2014 | 6455 | 13672 |

As outlined in section A.1, this differs from earlier studies, which used the unedited WHO HAT Atlas data for the Bas-Uélé region health zones and assumed that MSF was only present/active here until 2012. There has been no active screening or passive screening in these health zones since the end of MSF activities in 2014.

#### A.3 Locations

↩ Return to the Supplement Table of Contents

**Health zones included.** We have included 165 health zones in which 37.5M people live, and which have records of 131,941 cases in 2000–20. Our population data comes from the United Nations Office for the Coordination of Humanitarian Affairs censuses for national vaccine days [9].

**Health zones excluded.** We have omitted any health zones with less than 10 years of either AS or PS detection records. Therefore, our analysis omits 330 health zones, 69.9M people, and 3,789 cases – approximately 3% of all cases reported in 2000–20 (see Supplementary Table 2).

| | | Included | | | Excluded - insufficient data | | | Excluded - urban locale | | |
|---|---|---|---|---|---|---|---|---|---|---|
| **Coordination** | **Total No. HZ** | **No. HZ** | **Cases 2000-2020** | **Populati (mil-lions)** | **No. HZ** | **Cases 2000-2020** | **Populati (mil-lions)** | **No. HZ** | **Cases 2000-2020** | **Populati (mil-lions)** |
| Bandundu Nord | 20 | 18 | 36369 | 3.6 | 2 | 6 | 0.2 | 0 | 0 | 0.0 |
| Bandundu Sud | 32 | 19 | 30456 | 4.8 | 12 | 27 | 2.4 | 1 | 64 | 0.3 |
| Equateur Nord | 39 | 22 | 19340 | 4.7 | 17 | 21 | 3.6 | 0 | 0 | 0.0 |
| Equateur Sud | 30 | 9 | 1622 | 1.3 | 21 | 55 | 4.0 | 0 | 0 | 0.0 |
| Isangi - Bas-Uélé | 6 | 2 | 3297 | 0.2 | 4 | 3272 | 0.5 | 0 | 0 | 0.0 |
| Isangi - Tschopo | 29 | 4 | 2792 | 0.7 | 25 | 55 | 3.9 | 0 | 0 | 0.0 |
| Kasai Occidental | 45 | 18 | 6314 | 4.4 | 26 | 66 | 6.5 | 1 | 172 | 0.3 |
| Kasai Oriental | 35 | 22 | 17415 | 6.4 | 5 | 31 | 1.0 | 8 | 2874 | 3.1 |
| Kinshasa | 36 | 13 | 3027 | 4.2 | 9 | 108 | 1.8 | 14 | 1107 | 5.0 |
| Kongo Central | 30 | 17 | 4673 | 2.6 | 13 | 40 | 1.9 | 0 | 0 | 0.0 |
| Maniema Katanga | 29 | 13 | 4583 | 3.3 | 16 | 30 | 3.3 | 0 | 0 | 0.0 |
| Sankuru | 16 | 8 | 2053 | 1.3 | 8 | 17 | 1.1 | 0 | 0 | 0.0 |
| No Coordination | 172 | NA | 0 | 0.0 | 172 | 58 | 39.7 | 0 | 0 | 0.0 |
| **Total** | **519** | **165** | **131941** | **37.5** | **330** | **3786** | **69.9** | **24** | **4217** | **8.7** |

**Supplementary Table 2**: Summary of demographics characteristics and HAT case burden of health zones that were included compared to those excluded from the analysis. The health zones were omitted if there were fewer than 10 data points: in other words the years of AS reports available plus the years of PS reports available equalled less than 10, or if we did not believe that transmission could take place in the health zone because it was urban. Fewer than 3% of cases occurred in health zones with insufficient data and another 3% occurred in health zones that we have excluded because of their urban locale, where we believe there is no transmission. Abbreviations: HZ: health zones (zone de santé in the original French). AS: active screening, PS: passive screening.

For the model with animal transmission, we needed at least 13 years of either AS or PS detection records. Therefore, for 10 health zones, that have been included in the overall analysis, simulations come from only the model without animal transmission.

Moreover, health zones with no major rivers or which are too urban to sustain tsetse populations are not simulated. These are primarily in urban Kinshasa, in Kasaï Orientale coordination around the area of Mbuji-Mayi, a mining town without any rivers, and Kikwit Sud in Bandundu Sud. The cases reported there are approximately 4217, approximately 3% of all cases reported in 2000–20 (see Supplementary Table 2). We believe that these cases were reported due to the presence of large hospitals capable of diagnosing and treating people, but that these individuals were infected with the disease in other regions that could sustain transmission.

Therefore, we have modelled transmission in the areas of DRC that constitute 94% of all the HAT Atlas cases in 2000-2020.

**Population.** It must be noted that estimating the population of the DRC is difficult, as the country has not held a census since 1984, and many other forms of population surveillance are inconsistent throughout the country. Our data on the population for each health zone comes from the Office for the Coordination of Humanitarian Affairs [9], and a summary of the population per health zone is shown in Supplementary Table 2. We have assumed that the population has a 3% annual growth rate, compounded annually. Our total population comes out to 117.6 million, which roughly aligns with the population estimate from the CIA World Factbook estimate for 2023 [10] although it is higher than the UN World Population Prospects [11].

**Coordinations.** For the purpose of organising gHAT control and elimination efforts, the country is partitioned into eleven regions known as coordinations. See Supplementary Figure 1.

| Coordination | No. HZ | Pop. per HZ (thousands) | Pop. subtotal (millions) | 2000-2020 Cases per HZ[a] | 2000-2020 Sum cases | 2016-2020 Cases per HZ[a] | 2016-2020 Sum cases |
|---|---|---|---|---|---|---|---|
| Bandundu Nord | 18 | 180 [110–367] | 3.60 | 1748 [37–7186] | 36369 | 66 [3–210] | 1278 |
| Bandundu Sud | 19 | 249 [154–358] | 4.80 | 592 [10–6827] | 30456 | 36 [1–183] | 1194 |
| Equateur Nord | 22 | 188 [84–427] | 4.70 | 474 [25–4298] | 19340 | 2 [0–29] | 113 |
| Equateur Sud | 9 | 171 [45–212] | 1.30 | 92 [19–550] | 1622 | 2 [0–15] | 34 |
| Isangi - Bas-Uélé[b] | 2 | 110 [88–133] | 0.20 | 1648 [1390–1907] | 3297 | 0 [0–0] | 0 |
| Isangi - Tschopo[b] | 4 | 198 [106–208] | 0.70 | 579 [41–1593] | 2792 | 23 [0–55] | 101 |
| Kasai Occidental | 18 | 230 [117–420] | 4.40 | 180 [9–1982] | 6314 | 12 [0–85] | 446 |
| Kasai Oriental | 22 | 265 [153–543] | 6.40 | 472 [56–4567] | 17415 | 8 [1–58] | 307 |
| Kinshasa | 13 | 338 [90–594] | 4.20 | 98 [35–858] | 3027 | 5 [0–43] | 143 |
| Kongo Central | 17 | 133 [83–251] | 2.60 | 249 [15–912] | 4673 | 4 [0–51] | 169 |
| Maniema Katanga | 13 | 237 [107–417] | 3.30 | 391 [31–691] | 4583 | 14 [0–74] | 309 |
| Sankuru | 8 | 150 [95–239] | 1.30 | 218 [13–773] | 2053 | 20 [1–104] | 331 |
| **Total** | **165** | **210 [45-594]** | **37.58** | **342 [9-7186]** | **131941** | **9 [0-210]** | **4425** |

[a] Cases are shown per health zone: median [minimum-maximum].

[b] Isangi coordination has been separated into two subregions in this analysis. Bas-Uélé is constituted of Ango health zone in Bas-Uélé Province and Doruma health zone in Haut-Uélé Province. Tshopo is constituted of Isangi, Yabaondo, Yahisuli, and Yakusu health zones in Tschopo province.

**Supplementary Table 3**: Summary of the demographic characteristics and recent vs complete case burden in health zones in the analysis, stratified by the coordinations delineated the programme national de lutte contre la Trypanosomiase humaine africaine (PNLTHA-RDC). Abbreviation: HZ: health zone (zone de santé in the original French).

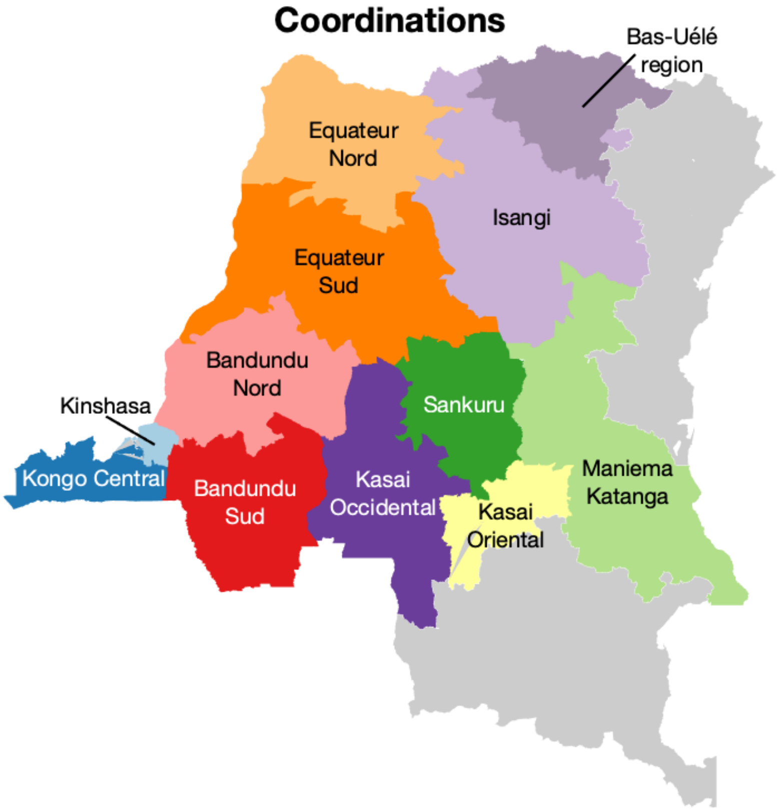

**Supplementary Figure 1**: Map of gHAT coordinations in the DRC. N.B. Isangi is a single coordination, however, we denote the region within Isangi we refer to as the Bas-Uélé region for context. Shapefiles used to produce this map were provided by Nicole Hoff and Cyrus Sinai under a CC-BY licence (current versions can be found at https://data.humdata.org/dataset/drc-health-data)

#### A.4 Model formulation

↩ Return to the Supplement Table of Contents

In this study, we use two mechanistic transmission model variants, both of which have a group of people at high risk of gHAT infection who do not participate in AS and a low-risk group of people who participate at random. The model variants differ concerning the presence or absence of non-human animal transmission cycles [2]. Below, we show the diagram and model equations, both reproduced from [2].

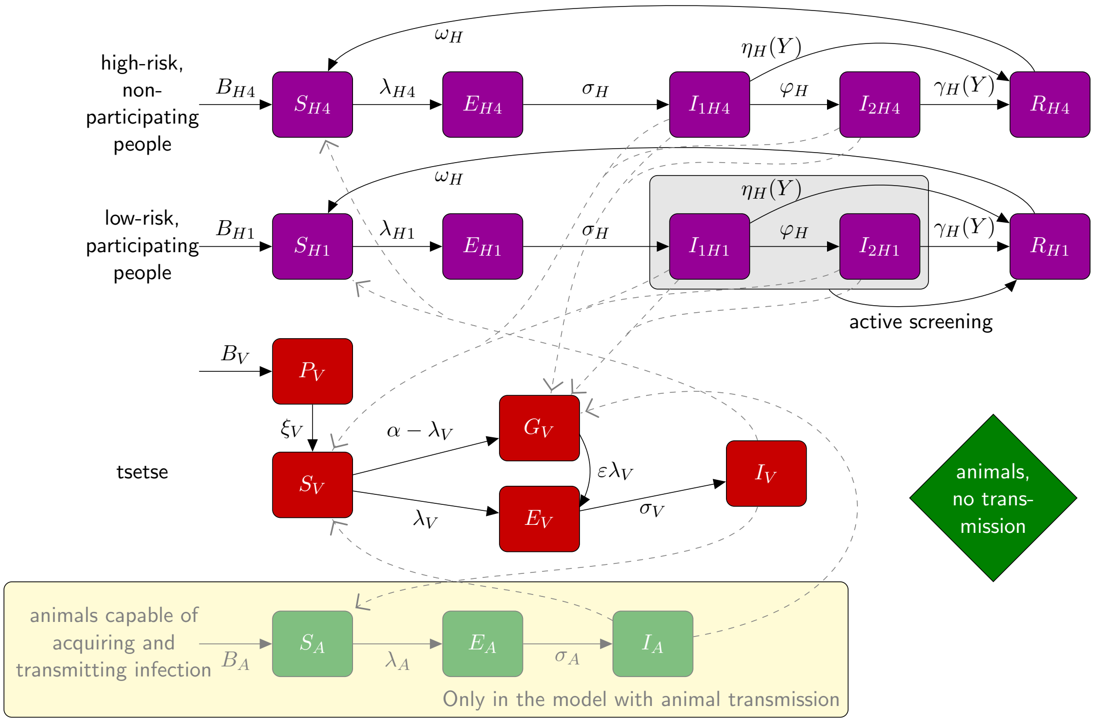

**Supplementary Figure 2**: Warwick gHAT intervention model compartmental diagram. Purple boxes denote human infection/risk compartments, red boxes denote tsetse infection compartments, and green boxes denote non-human animal infection compartments (only in the model variant with possible animal transmission). Solid lines represent the transition between infection states, and dashed lines are transmission pathways. Abbreviations and parameters: see text. Reproduced from [2] under a CC-BY licence.

For this set of model compartments, we can describe both deterministic and stochastic variations of the model. Below are the ODEs describing the deterministic dynamics. It was this version of the model which was used for fitting, whereas the analogous stochastic model, simulated using tau-leaping with a one-day time step, was used for sampling and projections into the future. More detail on the advantages and disadvantages of the deterministic and stochastic models is presented in Davis et al. [12], however, this previous work has demonstrated that using the deterministic model for fitting followed by the stochastic model for projections provides the speed of the deterministic fitting process but the integer outputs and stochastic variation generated by the stochastic model.

$$
\text{Humans}\quad\left\{
\begin{array}{lcl}
\dfrac{\mathrm{d}S_{Hi}}{\mathrm{d}t} & = & \mu_H N_{Hi} + \omega_H R_{Hi} - \alpha m_{\mathrm{eff}} f_i \dfrac{S_{Hi}}{N_{Hi}} I_V - \mu_H S_{Hi} \\
\dfrac{\mathrm{d}E_{Hi}}{\mathrm{d}t} & = & \alpha m_{\mathrm{eff}} f_i \dfrac{S_{Hi}}{N_{Hi}} I_V - (\sigma_H + \mu_H) E_{Hi} \\
\dfrac{\mathrm{d}I_{1Hi}}{\mathrm{d}t} & = & \sigma_H E_{Hi} - (\varphi_H + \eta_H(Y) + \mu_H) I_{1Hi} \\
\dfrac{\mathrm{d}I_{2Hi}}{\mathrm{d}t} & = & \varphi_H I_{1Hi} - (\gamma_H(Y) + \mu_H) I_{2Hi} \\
\dfrac{\mathrm{d}R_{Hi}}{\mathrm{d}t} & = & \eta_H(Y) I_{1Hi} + \gamma_H(Y) I_{2Hi} - (\omega_H + \mu_H) R_{Hi}
\end{array}
\right.
$$

$$
\text{Animals}\quad\left\{
\begin{array}{lcl}
\dfrac{\mathrm{d}S_A}{\mathrm{d}t} & = & \mu_A N_A - \alpha m_{\mathrm{eff}} f_A \dfrac{S_A}{N_A} I_V - \mu_A S_A \\
\dfrac{\mathrm{d}E_A}{\mathrm{d}t} & = & \alpha m_{\mathrm{eff}} f_A \dfrac{S_A}{N_A} I_V - (\sigma_A + \mu_A) E_A \\
\dfrac{\mathrm{d}I_A}{\mathrm{d}t} & = & \sigma_A E_A - \mu_A I_A
\end{array}
\right.
$$

$$
\text{Tsetse}\quad\left\{
\begin{array}{lcl}
\dfrac{\mathrm{d}P_V}{\mathrm{d}t} & = & B_V N_H - (\xi_V + \frac{P_V}{K}) P_V \\
\dfrac{\mathrm{d}S_V}{\mathrm{d}t} & = & \xi_V \mathbb{P}(\text{survive pupal stage}) P_V - \alpha S_V - \mu_V S_V \\
\dfrac{\mathrm{d}E_{1V}}{\mathrm{d}t} & = & \alpha\big(1 - f_T(t)\big) p_V \big(\sum_i f_i \dfrac{(I_{1Hi} + I_{2Hi})}{N_{Hi}} + f_A \dfrac{I_A}{N_A}\big)(S_V + \varepsilon G_V) \\
 & & \quad -(3\sigma_V + \mu_V + \alpha f_T(t)) E_{1V} \\
\dfrac{\mathrm{d}E_{2V}}{\mathrm{d}t} & = & 3\sigma_V E_{1V} - (3\sigma_V + \mu_V + \alpha f_T(t)) E_{2V} \\
\dfrac{\mathrm{d}E_{3V}}{\mathrm{d}t} & = & 3\sigma_V E_{2V} - (3\sigma_V + \mu_V + \alpha f_T(t)) E_{3V} \\
\dfrac{\mathrm{d}I_V}{\mathrm{d}t} & = & 3\sigma_V E_{3V} - (\mu_V + \alpha f_T(t)) I_V \\
\dfrac{\mathrm{d}G_V}{\mathrm{d}t} & = & \alpha(1 - f_T(t))\Big(1 - p_V\big(\sum_i f_i \dfrac{(I_{1Hi} + I_{2Hi})}{N_{Hi}} + f_A \dfrac{I_A}{N_A}\big)\Big) S_V \\
 & & \quad -\alpha\Big(f_T(t) + \big(1 - f_T(t)\big) p_V \varepsilon \big(\sum_i f_i \dfrac{(I_{1Hi} + I_{2Hi})}{N_{Hi}} + f_A \dfrac{I_A}{N_A}\big)\Big) G_V \\
 & & \quad -\mu_V G_V
\end{array}
\right. \tag{1}
$$

The function which describes the probability of a host-seeking tsetse both hitting a Tiny Target and dying as a result, $f_T$, is time-dependent ($t$, in days) from when the targets were first deployed:

$$
f_T(t) = f\text{max}\left(1 - \frac{1}{1 + \exp(-0.068(\text{mod}(t, 182.5) - 127.75))}\right) \tag{2}
$$

and $f\text{max}$ is the maximum daily probability of contacting a Tiny Target and dying as a result. $f_T$ modifies all the bite rates $\alpha$ in our tsetse equations to produce an additional Tiny-Target-induced mortality for tsetse. The choice of parameterisation of this function in different locations is described in the next section.

##### A.4.1 Assumptions about past interventions

###### Regions with no transmission

In the previous model fitting studies [1, 2], it was assumed that no transmission took place in the health zones that constituted urban Kinshasa. In this study we have added a few health zones into the analysis where we believe

| Notation | Description | Value | |
|---|---|---|---|
| $N_H$* | Total human population size in 2015 | Fixed for each health zone | [9] |
| $\mu_H$ | Natural human mortality rate | $5.4795 \times 10^{-5}$ days$^{-1}$ | [13] |
| $B_H$ | Total human birth rate | $= \mu_H N_H$ | |
| $\sigma_H$ | Human incubation rate | 0.0833 days$^{-1}$ | [14] |
| $\varphi_H$ | Stage 1 to 2 progression rate | 0.0019 days$^{-1}$ | [15, 16] |
| $\omega_H$ | Recovery rate or waning-immunity rate | 0.006 days$^{-1}$ | [17] |
| Sens | Active screening diagnostic sensitivity | 0.91 | [18] |
| $B_V$† | Tsetse birth rate (per capita rate of depositing new pupae) | 0.0505 days$^{-1}$ | [4] |
| $\xi_V$ | Rate of pupal development to adult flies | 0.037 days $^{-1}$ | [4] |
| $K$‡ | Pupal carrying capacity | $= 111.09 N_H$ | [4] |
| $\mathbb{P}$(pupating) | Probability of a pupa surviving to emerge as an adult fly | 0.75 | [4] |
| $\mu_V$ | Tsetse mortality rate | 0.03 days$^{-1}$ | [14] |
| $\sigma_V$ | Tsetse incubation rate | 0.034 days$^{-1}$ | [19, 20] |
| $\alpha$ | Tsetse bite rate | 0.333 days$^{-1}$ | [21] |
| $p_V$ | Probability of tsetse infection per single infective bite | 0.065 | [14] |
| $\varepsilon$ | Reduced susceptibility factor for non-teneral (previously fed) flies | 0.05 | [22] |
| $f_H$ | Proportion of blood-meals on humans | 0.09 | [23] |
| $\eta_H^{\text{pre}}$ | Treatment rate from stage 1, pre-1998 | 0 | Assumed |
| $\text{disp}_{\text{act}}$§ | Overdispersion parameter for active detection | $4 \times 10^{-4}$ | [1] |
| $\text{disp}_{\text{pass}}$§ | Overdispersion parameter for passive detection | $1.68 \times 10^{-5}$¶ | [1] |
| **Parameters specific to the model with animal transmission...** | | | |
| $\mu_A$ | Natural animal mortality rate | 0.0014 days$^{-1}$ | Assumed |
| $\sigma_A$ | Animal incubation rate | 0.0833 days$^{-1}$ | [14] |

*The model is internally scaled such that the population size in all years corresponds to the population in 2015 (outputs are back-transformed to reflect an assumed annual population growth rate of 3% across the DRC).
†The value of $B_V$ was chosen to maintain constant population size in the absence of vector control interventions.
‡The value of $K$ was chosen to reflect the observed bounce back rate.
§Over-dispersion values were originally chosen based on a comparison of the median of the distributions of log posterior probability from MCMC runs with $\rho$ fixed at a range of values for two example health zones under the model without animal transmission [1].
¶The over-dispersion value for passive screening in the Bas-Uele region is $2.8 \times 10^{-5}$.

**Supplementary Table 4**: Model parameterisation (fixed parameters). Notation, a brief description, and the values used for fixed parameters. This table is updated from [1, 2].

| Notation | Description | Prior distribution* | Percentiles of prior distribution [2.5, 50 & 97.5%] | Unit |
|---|---|---|---|---|
| $R_0$ | Basic reproduction number (NGM approach) | $1 + \text{Exp}(10)$ | [1.003, 1.069, 1.369] | - |
| $r$ | Relative bites taken on high-risk humans | $1 + \Gamma(3.68, 1.09)$ | [2.015, 4.654, 10.028] | - |
| $k_1$ | Proportion of low-risk people | $\text{B}(16.97, 3.23)$ | [0.6564, 0.8514, 0.9609] | - |
| $\eta_H^{\text{post}}$† | Treatment rate from stage 1, post endemic equilibrium | $\Gamma(3.54, 5.32 \times 10^{-5})$ | [4.59, 17.1, 42.9]$\times 10^{-5}$ | days$^{-1}$ |
| $\gamma_H^{\text{post}}$ | Combined treatment and disease-induced death rate from stage 2, 1998 onwards | $\Gamma(6.2082, 0.001)$ | [2.33, 5.88, 12.0]$\times 10^{-3}$ | days$^{-1}$ |
| $\gamma_H^{\text{pre}}$ | Combined treatment and disease-induced death rate from stage 2, pre-1998 | $\Gamma(6.2082, 0.001)$ | [2.33, 5.88, 12.0]$\times 10^{-3}$ | days$^{-1}$ |
| Spec | Active screening diagnostic specificity | $0.998 + (1 - 0.998)\,\text{B}(7.23, 2.41)$ | [0.9989, 0.9995, 0.9999] | - |
| $u$ | Proportion of stage 2 cases reported from passive screening | $\text{B}(20, 40)$ | [0.2208, 0.3315, 0.4564] | - |
| $d_{\text{change}}$† | Midpoint year for passive improvement | $2000 + (2017 - 2000)\,\text{B}(5, 6)$ | [2003.2, 2007.7, 2012.5] | year |
| $\eta_{H_{\text{amp}}}$† | Relative improvement in passive screening stage 1 detection rate | $\Gamma(2.5133, 1.3216)$ | [0.556, 2.893, 8.509] | - |
| $\gamma_{H_{\text{amp}}}$† | Relative improvement in passive screening stage 2 detection rate | $\Gamma(2.3095, 0.5727)$ | [0.198, 1.137, 3.493] | - |
| $d_{\text{steep}}$† | Speed of improvement in passive screening detection rate | $\Gamma(39.57, 0.0270)$ | [0.761, 1.058, 1.424] | years$^{-1}$ |
| **Parameters specific to the model with animal transmission...** | | | | |
| $f_A$ | Proportion of blood meals on reservoir animals | $\text{B}(1.3, 1.3)$ | $[0.046, 0.5, 0.954]$ | - |
| $k_A$ | Relative size of animal reservoir population | $\Gamma(1.26, 19.3)$ | $[1.18, 18.3, 81.4]$ | - |

*Where Exp(.), $\Gamma(.)$ and B(.) are the exponential, gamma (parameterised with shape and scale) and beta distributions, respectively.
†Former province-specific priors were originally used for $\eta_H^{\text{post}}$, $d_{\text{change}}$, $\eta_{H_{\text{amp}}}$, $\gamma_{H_{\text{amp}}}$ and $d_{\text{steep}}$; prior distributions and percentiles for the former province of Bandundu presented (i.e., Bandundu Nord and Bandundu Sud coordinations), see Table 6 for other coordinations.

**Supplementary Table 5**: Model parameterisation (fitted parameters). Notation, brief description, and information on the prior distributions for fitted parameters. This table is updated from Crump et al. [2].

| **Parameter** | | |
|---|---|---|
| **Coordination(s)** health zone(s) | **Prior distribution*** | **Percentiles of prior distribution** |
| $\eta_H^{\text{post}}$ – Treatment rate from stage 1, post endemic equilibrium | | |
| **Bandundu Nord & B. Sud** | $\Gamma(3.54, 5.32\times10^{-5})$ | $[4.59, 17.1, 42.9]\times10^{-5}$ |
| **Équateur Nord & É. Sud** | $\Gamma(4.92, 4.51\times10^{-5})$ | $[7.12, 20.7, 45.7]\times10^{-5}$ |
| **Isangi** | $\Gamma(1.16, 9.27\times10^{-5})$ | $[4.24, 79.0, 373]\times10^{-6}$ |
| **Kasaï Occidental** | $\Gamma(10.9, 3.03\times10^{-5})$ | $[1.64, 3.20, 5.53]\times10^{-4}$ |
| **Kasaï Oriental** | $\Gamma(2.90, 5.87\times10^{-5})$ | $[3.38, 15.1, 41.5]\times10^{-5}$ |
| **Kinshasa** | $\Gamma(1.26, 8.91\times10^{-5})$ | $[5.44, 84.4, 376]\times10^{-6}$ |
| **Kongo Central** | $\Gamma(12.0, 2.89\times10^{-5})$ | $[1.78, 3.36, 5.68]\times10^{-4}$ |
| **Maniema-Katanga** | $\Gamma(4.25, 4.85\times10^{-5})$ | $[5.90, 19.0, 44.3]\times10^{-5}$ |
| Health zones in Katanga† | $\Gamma(1.29, 8.79\times10^{-5})$ | $[5.88, 86.2, 376]\times10^{-6}$ |
| **Sankuru** | $\Gamma(2.90, 5.87\times10^{-5})$ | $[3.38, 15.1, 41.5]\times10^{-5}$ |
| $d_{\text{change}}$ – Midpoint year for improvement in passive detection rate | | |
| **Bandundu Nord & B. Sud** | $2000 + (2017 - 2000)\,\text{B}(5, 6)$ | $[2003.2, 2007.7, 2012.5]$ |
| **Équateur Nord & É. Sud** | $2000+(2020 - 2000)\,\text{B}(2.79, 23.1)$ | $[2000.4, 2002, 2005]$ |
| **Kasaï Oriental** | $2000+(2020 - 2000)\,\text{B}(2.79, 23.1)$ | $[2000.4, 2002, 2005]$ |
| **Kinshasa** | $2000+(2020 - 2000)\,\text{B}(2.79, 23.1)$ | $[2000.4, 2002, 2005]$ |
| Masa health zone‡ | Fixed parameter value, $d_{\text{change}} = 2015.5$ | |
| **Kongo Central** | Fixed parameter value, $d_{\text{change}} = 2015.5$ | |
| $\eta_{H_{\text{amp}}}$ – Relative improvement in passive stage 1 detection rate | | |
| **Bandundu Nord & B. Sud** | $\Gamma(2.51, 1.32)$ | $[0.556, 2.89, 8.51]$ |
| **Équateur Nord & É. Sud** | $\Gamma(1, 2.17)$ | $[0.055, 1.510, 8.010]$ |
| **Kasaï Oriental** | $\Gamma(1, 2.17)$ | $[0.055, 1.510, 8.010]$ |
| **Kinshasa** | $\Gamma(1, 2.17)$ | $[0.055, 1.510, 8.010$ |
| Masa health zone | $\Gamma(2.51, 1.32)$ | $[0.556, 2.89, 8.51]$ |
| **Kongo Central** | $\Gamma(2.51, 1.32)$ | $[0.556, 2.89, 8.51]$ |
| $\gamma_{H_{\text{amp}}}$ – Relative improvement in passive stage 2 detection rate | | |
| **Bandundu Nord & B. Sud** | $\Gamma(2.31, 0.57)$ | $[0.198, 1.14, 3.49]$ |
| **Équateur Nord & É. Sud** | $\Gamma(1, 1.0014)$ | $[0.0254, 0.6943.69]$ |
| **Kasaï Oriental** | $\Gamma(1, 1.0014)$ | $[0.0254, 0.6943.69]$ |
| **Kinshasa** | $\Gamma(1, 1.0014)$ | $[0.0254, 0.6943.69]$ |
| Masa health zone | $\Gamma(2.31, 0.57)$ | $[0.198, 1.14, 3.49]$ |
| **Kongo Central** | $\Gamma(2.31, 0.57)$ | $[0.198, 1.14, 3.49]$ |
| $d_{\text{steep}}$ – Speed of improvement in passive detection rate | | |
| **Bandundu Nord & B. Sud** | $\Gamma(39.6, 2.70\times10^{-2})$ | $[0.761, 1.06, 1.42]$ |
| **Équateur Nord & É. Sud** | $\Gamma(15.7, 0.51)$ | $[4.55, 7.84, 12.4]$ |
| **Kasaï Oriental** | $\Gamma(15.7, 0.51)$ | $[4.55, 7.84, 12.4]$ |
| **Kinshasa** | $\Gamma(15.7, 0.51)$ | $[4.55, 7.84, 12.4]$ |
| **Kongo Central** | $\Gamma(15.7, 0.51)$ | $[4.55, 7.84, 12.4]$ |
| $\text{Spec}_{\text{MSF}}$ – Specificity of MSF active screening algorithm | | |
| **Isangi** – Bas-Uele region§ | $\text{B}(299, 2.87)$ | $[0.977, 0.992, 0.998]$ |

*Where $\Gamma(.)$ and $\text{B}(.)$ are the gamma (parameterised with shape and scale) and beta distributions, respectively.
†Kabalo, Kongolo, Mbulula and Nyunzu health zones.
‡Masa health zone is in Kinshasa coordination but Kongo Central province. It was assumed to be subject to passive screening improvement in the same period as Kongo Central.
§Ango, Ganga and Doruma health zones, where intervention activities were carried out by MSF.

**Supplementary Table 6**: Model parameterisation (fitted parameters): coordination or health zone specific priors. Adapted from Crump et al. [2].

local transmission is possible due to tsetse presence: Nsele, Masa, Mont Ngafula 1 & 2, and Maluku 1 & 2. These are denoted as “no transmission” health zones in our graphical user interface maps https://hatmepp.warwick.ac.uk/DRCCEA/v8/.

Health zones with no data or insufficient data for fitting the model are excluded, as described in Section A.2. There may be some infection in these locations, although we believe that most of the “no data” health zones are unlikely to have transmission. The “no data” and “<10 data point” health zones can be viewed in our graphical user interface maps https://hatmepp.warwick.ac.uk/DRCCEA/v8/.

##### Passive detection

The integration of gHAT case detection through peripheral health centres using RDTs is an area with limited documented evidence in the DRC [24, 25]. While positive effects have been observed in other countries, such as Chad [26, 27], the follow-up for infection confirmation among RDT-positive cases in the DRC is reported to have a high level of attrition if the health centre where someone was screened does not have immediate confirmation available [28]. Despite this, there is evidence, particularly from the passive case staging data over time, that time to detection through PS has decreased over time [1, 29].

In the same manner as our previous study [2], for improvements between the start of activities and 2020 we use the same following equations to describe transmission rates from infected classes:

$$\eta_H(Y) = \eta_H^{\text{post}} \left[ 1 + \frac{\eta_{H_{\text{amp}}}}{1 + \exp\left(-d_{\text{steep}}(Y - d_{\text{change}})\right)} \right] \tag{3}$$

$$\gamma_H(Y) = \gamma_H^{\text{post}} \left[ 1 + \frac{\gamma_{H_{\text{amp}}}}{1 + \exp\left(-d_{\text{steep}}(Y - d_{\text{change}})\right)} \right] \tag{4}$$

We assume that all stage 1 infections are either reported as cases or progress to stage two, but that some of the exits from stage 2 are due to death from gHAT disease. In 1998 the reporting probability for an exit from stage 2 is given by $u$, however as the exit rate from stage 2 increases this reporting probability does not stay constant, but increases (proportionally more people would be detected and treated with higher exit rates). When we compute reporting rates from stage 2 we therefore use the following:

$$\text{Death rate} = (1-u)\gamma_H^{\text{post}} \tag{5}$$

$$\text{Stage 2 reporting incidence} = (\gamma_H(Y) - \text{Death rate})(I_{2H1} + I_{2H4}) \tag{6}$$

##### The Bas Uélé region

Previously, it was assumed that screening activities in the former province of Orientale (current-day Bas Uélé) had been carried out by Médecins Sans Frontières (MSF) until 2012, allowing us to differentiate between MSF-specific specificity and sensitivity parameters and those associated with PNLTHA activities in the subsequent years. The specificity in these two periods was fitted with the MSF-specific specificity constrained to be lower than the PNLTHA specificity, while the sensitivity values were assumed to have constant values (0.91 in all years). In these fits we have refined this assumption based on patterns in the data and information received from PNLTHA and from an MSF report [3], with MSF-lead activity being limited to some health zones in the Bas-Uélé region: Ango, Bili, Doruma, Ganga, Poko and Titule. MSF were active in these health zones between 2007–2014, and there have been no gHAT control activities, neither active nor PS, in this region after this. As a result, there is no information on the specificity of PNLTHA screening, and so this parameter reflects the prior distribution (our prior belief).

##### Historical VC

$f_{\text{max}}$ in Equation 2 is chosen such that the tsetse population after one year is 80% multiplied by the proportion of recent gHAT cases coverage by the intervention, except for in Yasa Bonga in Bandundu Sud coordination which already has a measured 90% reduction in the intervention area [30].

To calculate an estimate of the case coverage of the intervention areas, we applied the following algorithm: First, we selected the river segments in the DRC where VC had previously been applied from the HydroRivers dataset

| Health zone | Year started | Tsetse reduction after 1 year | Estimated coverage (based on 2016–2020 cases) |
|---|---|---|---|
| Mosango | 2015.5 | assumed 80% | 11% |
| Yasa Bonga | 2016.5 | reported 90% | 73% |
| Kenge | 2017.5 | assumed 80% | 28% |
| Masi Manimba | 2018 | assumed 80% | 53% |
| Bandundu | 2019.5 | assumed 80% | 66% |
| Bolobo | 2019.5 | assumed 80% | 14% |
| Kikongo | 2019.5 | assumed 80% | 18% |
| Kwamouth | 2019.5 | assumed 80% | 46% |
| Bulungu | 2021 | assumed 80% | 28% |
| Kimputu | 2021 | assumed 80% | 35% |
| Mokala | 2021 | assumed 80% | 15% |
| Bagata | 2021.5 | assumed 80% | 47% |
| Bokoro | 2021.5 | assumed 80% | 15% |
| Ipamu | 2021.5 | assumed 80% | 10% |

**Supplementary Table 7**: List of health zones with previous vector control (VC) deployments. This table includes health zones which have had “inadvertent VC” as boundary rivers had VC due to official deployments in neighbouring health zones. The year started is our modelled start time to the nearest half year, the tsetse reduction after 1 year is assumed to be 80% in all health zones apart from Yasa Bonga where a 90% reduction was reported. Case coverage is estimated using geolocated case data from 2016–2020 as described above.

and then created a 5km buffer zone around these areas. For each health zone, we then intersected these buffer zones with the health zone and counted the number of cases that fell inside this buffer between 2016 and 2020. Any cases that fell into multiple buffers (either for the same or multiple health zones) were split evenly between these buffers and counted fractionally to the relevant health zones. The total number of cases inside the buffers was then compared to the total number of cases in the health zone for the same period to estimate a fractional case coverage. The 80% (90% in Yasa Bonga) reduction was then scaled by this coverage to produce a new estimate for $f_{\text{max}}$.

#### A.5 Model fitting

↩ Return to the Supplement Table of Contents

An adaptive Metropolis-Hastings Markov chain Monte Carlo (MCMC) algorithm was used to fit two deterministic transmission model variants to epidemiological data as in previous modelling studies in the DRC [1, 2]. The model variants differ with regard to the presence or absence of animals contributing to gHAT transmission. Since these earlier studies, the data has been updated to span the years 2000 to 2020 (rather than 2000 to 2016) before aggregation at the health zone level within a year. As before, model fitting was carried out independently within each health zone.

Our ODE models were run from their endemic equilibrium. Two chains were run in the MCMC, and they were initialised using the fixed parameters and by random perturbations around supplied, individually valid, initial values of each parameter being fitted, rejecting those parameter sets that do not produce a valid posterior probability.

##### Likelihood

As described in Crump et al. [2], eight parameters; $R_0, r, \eta_H, \gamma_H, b_{\gamma_{H0}}, k_1, u$, and Spec were fitted in all health zones for both models. A further two parameters; $k_A$ and $f_A$, were fitted in all health zones for the model with animal transmission. Additional parameters were included as required (combinations of $d_{\text{change}}$, $\eta_{H_{\text{amp}}}$, $\gamma_{H_{\text{amp}}}$, $d_{\text{steep}}$ and $\text{Spec}_{\text{MSF}}$ as appropriate, see above).

For fitting the model to case data, we transform model ODE solutions (for S1.2.1) into annual case reporting denoted $A_{M1}$, $A_{M2}$, for active stage 1 and stage 2 and $P_{M1}$, $P_{M2}$, for passive stage 1 and 2. Since we always know the stage (1 or 2) in the model simulations, there is no requirement for a “U” (unknown stage) category for the model. These are computed using solutions to the ODEs for the given set of parameters aggregated across a year.

Detections relate to the transfer from infectious categories to the recovered category – the new annual reported case incidence. This is either by passive detection from stage 1 for year $Y$:

$$P_{M1}(Y) = \int_Y^{Y+1} \eta_H(Y)\big(I_{1H1}(t) + I_{1H4}(t)\big)\,\mathrm{d}t,$$

passive detection from stage 2

$$P_{M2}(Y) = \int_Y^{Y+1} (\gamma_H(Y) - \text{Death rate})\big((I_{2H1}(t) + I_{2H4}(t)\big)\,\mathrm{d}t,$$

or by AS from the low-risk ($H1$) group in year $Y$

$$A_{M1}(Y) = z(Y)\text{Sens}I_{1H1}(Y) + z(Y)(1 - \text{Spec})\big(k_1 N_H - I_{1H1}(Y) - I_{2H1}(Y)\big)$$

and

$$A_{M2}(Y) = z(Y) \times \text{Sens} \times I_{2H1}(Y)$$

with variable AS coverage by year, $z(Y)$ and fixed diagnostic sensitivity. $A_{M1}$ also contains any false positives that may have been incorrectly identified from non-infected people based on the high but imperfect specificity of the AS algorithm. We assume in the DRC that all false positives would be assigned to be stage 1 and treated, however, in the model false positives stay in the susceptible category, unlike true cases which move to the recovered category.

The log-likelihood function used in the adaptive Metropolis-Hastings MCMC contained two terms in each year for which reported case numbers were available for each source of reported cases (active or PS). These were:

- a beta-binomial probability that the total number of cases reported in that year for that source came from the available population (either the reported number of people actively screened for AS or the health zone population for PS) with probability calculated from solving the ODE for the current set of parameters, and
- a binomial probability that reported stage 1 cases come from the total number of reported staged cases, where the probability parameter again comes from the solution of the ODE. In many years, staging is unknown, so this part of the log-likelihood will return zero and not contribute to our calculation. In some years, we have only partially known staging information.

This formulation allowed over-dispersion in the observed cases to be included, via the beta-binomial distribution, and any proportion of cases with reported disease stage to be appropriately accounted for (assuming that the reporting of staging information is independent of the disease stage). The log-likelihood function was as follows:

$$\begin{aligned} LL(\theta|x) = &\ \log(P(x|\theta)) \\ \propto &\sum_{i=2000}^{2016}\left(\log\left[\mathrm{BetaBin}\left(A_{D1}(i)+A_{D2}(i)+A_{DU}(i); z(i), \frac{A_{M1}(i)+A_{M2}(i)}{z(i)}, \mathrm{disp}_{\mathrm{act}}\right)\right]\right. \\ &+\log\left[\mathrm{Bin}\left(A_{D1}(i); A_{D1}(i)+A_{D2}(i), \frac{A_{M1}(i)}{A_{M1}(i)+A_{M2}(i)}\right)\right] \\ &+\ \log\left[\mathrm{BetaBin}\left(P_{D1}(i)+P_{D2}(i)+P_{DU}(i); N_H, \frac{P_{M1}(i)+P_{M2}(i)}{N_H}, \mathrm{disp}_{\mathrm{pass}}\right)\right] \\ &\left.+\log\left[\mathrm{Bin}\left(P_{D1}(i); P_{D1}(i)+P_{D2}(i), \frac{P_{M1}(i)}{P_{M1}(i)+P_{M2}(i)}\right)\right]\right) \end{aligned}$$

The model takes parameterisation $\theta$, $x$ is the data, $P_{Dj}(i)$ and $A_{Dj}(i)$ are the number of cases detected by passive or AS (of stage $j$, which may be 1, 2 or unknown, $U$) in a year $i$ of the data. $P_{Mj}(i)$ and $A_{Mj}(i)$ are the number of cases detected by passive or AS (of stage $j$) in year $i$ of the model, and $z(i)$ is the number of people actively screened in year $i$. $\mathrm{BetaBin}(m; n, p, \rho)$ gives the probability of obtaining $m$ successes out of $n$ trials with probability $p$ and overdispersion parameter $\rho$. The overdispersion accounts for a larger variance than under the binomial. The probability density function of this distribution is given by:

$$\mathrm{BetaBin}(m; n, p, \rho) = \frac{\Gamma(n+1)\Gamma(m+a)\Gamma(n-m+b)\Gamma(a+b)}{\Gamma(n-m+1)\Gamma(n+a+b)\Gamma(a)\Gamma(b)}$$

where $a = p(1/\rho - 1)$ and $b = a(1-p)/p$.

##### Missing active screening numbers

There are instances in the data where the number of cases from within year $t$ ($A_D(t) = A_{D1} + A_{D2}$) is not consistent with the number of people recorded as having been screened in that year for that health zone ($z(t)$), i.e. (i) $A_D(t) > z(t)$ or (ii) $A_D(t)/z(t)$ is a much higher prevalence than is biologically realistic for gHAT (e.g. more than 10%).

In the previous DRC modelling studies [1, 2], we did not take any action concerning the very high prevalence in some years within a health zone. Instead, we considered only two scenarios:

1. If $A_D(t) < 20$ and $z(t) \leq A_D(t)$ we assumed that these people attended a screening outside their home health zone and that the record has been allocated correctly to their home health zone. in this case we set $z(t) = A_D(t)$.
2. Where $z(t) = 0$ and $A_D(t) > 0$, then we imputed the number of negative tests.

In the current study, we have chosen to impute the number of negative tests in more situations. We still believe that where the number of cases reported is low these people probably attended screening elsewhere, but imputing a missing screening value, in our independent health zone analyses, is expected to reflect the model's underlying prevalence better. We, therefore, imputed the number of negative tests for a year within a health zone where:

1. the number of cases from was more than 10% of the number screened ($A_D(t) > 0.1 \times z(t)$;
2. the number screened was zero, or not recorded, or less than 20 ($z(t) \leq 20$).

Imputation of the number of negative tests takes place within the MCMC fitting procedure.

We use a Geometric prior for the number of negative screening tests in year $t$, $A_D^-(t) \sim \mathrm{Geom}(\lambda_t)$, where $\lambda_t = \frac{1}{1+\bar{N}_t}$ and $\bar{N}_t = \frac{\sum_{j=2000, j\neq t}^{2020} N_j e^{-|t-j|}}{\sum_{j=2000, j\neq t}^{2020} e^{-|t-j|}}$, a weighted mean of the number of people screened in years other than $t$. The proposal distribution for $A_D^-(t)$ was a negative binomial distribution:

$$A_D^-(t)|A_D(t), p(t) \sim \mathrm{NB}(A_D(t)+1, 1-(1-p(t))(1-\lambda_t))$$

where the probability of active case detection, $p(t)$, was sampled from the following Beta distribution:

$$p(t)|\theta \sim \text{Beta}\left(\hat{p}(t)\left(\frac{1}{\text{disp}_{\text{act}}(t)} - 1\right), (1 - \hat{p}(t))\left(\frac{1}{\text{disp}_{\text{act}}(t)} - 1\right)\right)$$

and $\hat{p}(t)$ is the probability of active case detection in year $t$ calculated from the ODE outputs.

#### A.6 Projections

↩ Return to the Supplement Table of Contents

Samples and future projections were run using a stochastic model analogous to the deterministic model used during the MCMC fitting. This approach allowed us to avoid the computational expense of fitting a stochastic model (e.g. via particle filter MCMC or approximate Bayesian computation), but still gain the advantages of stochastic model outputs, in particular being able to directly assess the model when elimination of transmission or elimination of infection has occurred. This is not possible in the deterministic framework without using a proxy threshold [5, 31]. Recent work by Davis et al. has demonstrated the very good alignment between posteriors from deterministic MCMC and stochastic pMCMC fitting of this gHAT model [32].

Model projections were carried out by taking 2,000 random samples from the joint posterior distribution of the model parameters, using these to simulate the stochastic model 10 times for each sample from the joint posterior distribution of the model parameters with observational samples for case reporting using the beta-binomial distributions described earlier. The stochastic model covered the period for which data were fitted (2000 to 2020) as well as projecting into the future under various intervention strategies (see section A.7).

##### Ensemble model

The results from the stochastic projections for the two model variants were combined into an ensemble model, with the proportion taken from each model based on the relative model evidence. The model evidence, or marginal likelihood, for each model, was estimated using the importance sampled estimator [33].

The following description of the important sampling method to estimate the model evidence has been adapted slightly from that in the Supplementary Information for Crump et al. [2].

The joint distribution of $(\boldsymbol{\theta}_m, \mathbf{x})$, for parameters $\boldsymbol{\theta}_m = (\theta_1, \theta_2, \ldots, \theta_{d_m})$ of model $m$ and data $\mathbf{x} = (x_1, x_2, \ldots, x_n)$ satisfies

$$\pi(\boldsymbol{\theta}_m|\mathbf{x})\,\pi(\mathbf{x}|\mathbf{m}) = \pi(\mathbf{x}|\boldsymbol{\theta}_m)\,\pi(\boldsymbol{\theta}_m)\,, \tag{7}$$

where $\pi(\boldsymbol{\theta}_m|\mathbf{x})$ is the joint posterior distribution of parameters $1 \ldots d$, $\pi(\mathbf{x}|\mathbf{m})$ is the marginal likelihood or *evidence*; $\pi(\mathbf{x}|\boldsymbol{\theta}_m)$ is the likelihood, and $\pi(\boldsymbol{\theta}_m)$ is the prior distribution.

By use of MCMC methods to investigate the posterior distribution of the parameters, calculation of $\pi(\mathbf{x}|\mathbf{m})$ is avoided. Calculation of the evidence for use in model comparison requires computing the integral:

$$\pi(\mathbf{x}|\mathbf{m}) = \int \pi(\mathbf{x}|\boldsymbol{\theta}_m)\,\pi(\boldsymbol{\theta}_m)\;\mathrm{d}\boldsymbol{\theta}_m \tag{8}$$

$$= \int \pi(\mathbf{x}|\boldsymbol{\theta}_m)\,\frac{\pi(\boldsymbol{\theta}_m)}{q(\boldsymbol{\theta}_m)}q(\boldsymbol{\theta}_m)\;\mathrm{d}\boldsymbol{\theta}_m \tag{9}$$

Equation 8 cannot be calculated analytically except for some small set of tractable models. It can, however, be rewritten as equation 9, where $q(\boldsymbol{\theta}_m)$ is a $d_m$-dimensional probability density function. From this, an importance sampled estimator of $\pi(\mathbf{x}|\mathbf{m})$ is:

$$\hat{P}_q = \frac{1}{N}\sum_{i=1}^{N}\pi(\mathbf{x}|\boldsymbol{\theta}_{m,i})\,\frac{\pi(\boldsymbol{\theta}_{m,i})}{q(\boldsymbol{\theta}_{m,i})}, \tag{10}$$

where the $\boldsymbol{\theta}_{m,i}$ are $N$ samples drawn from $q$.

A defence mixture [34] was used for $q(\boldsymbol{\theta}_m)$:

$$q(\boldsymbol{\theta}_m) = p\phi(\boldsymbol{\theta}^*_m; n, \boldsymbol{\mu}_1 \ldots \boldsymbol{\mu}_n, \mathbf{C}_1 \ldots \mathbf{C}_n)\left|\frac{\boldsymbol{\theta}^*_m}{\boldsymbol{\theta}_m}\right| + (1-p)\,\pi(\boldsymbol{\theta}_m) \tag{11}$$

where $\phi(\cdot)$ is a mixture of $n$ multivariate Gaussian distributions with vectors of means $\boldsymbol{\mu}_j$ ($j = \{1 \ldots n\}$), and covariance matrices $\mathbf{C}_j$, $\left|\frac{\theta^*_m}{\theta_m}\right|$ is the Jacobian transformation relating probability on transformed and original scales, and $p$ is a mixing proportion ($p = 0.95$ was chosen for use, being a typical value [33]).

In each of our health-zone-level MCMC analyses of the models with and without animal transmission, $2\,000$ samples from the joint posterior distribution were generated and stored, and $\phi\big(\boldsymbol{\theta}_m; n, \boldsymbol{\mu}_1 \ldots \boldsymbol{\mu}_n, \mathbf{C}_1 \ldots \mathbf{C}_n\big)$ for each health zone and model was chosen using the Matlab `fitgmdist` function, selecting $n$ based on Akaike's Information Criterion (AIC). To account for the high correlations between some of our model parameters, regularisation was applied to ensure that the covariance matrices, $\mathbf{C}_k$, would be positive semi-definite. Before passing to `fitgmdist`, transformations were applied to the posterior samples to put them in the range $(-\infty, \infty)$ – appropriate for Gaussian distributions – followed by scaling and centring to keep the regularisation consistent across analyses, at least at the simple, single overall covariance matrix level.

Having defined $\phi(\cdot)$ for a given analysis (health zone, model combination), $\hat{P}_q$ was calculated (equation 10) using $N = 10\,000$ samples drawn from $q(\boldsymbol{\theta}_m)$. Note that this is an increase from the $2\,000$ samples used previously [2], and is expected to reduce the possible influence of sampling on the model evidence results.

#### A.7 Strategies and interventions

↩ Return to the Supplement Table of Contents

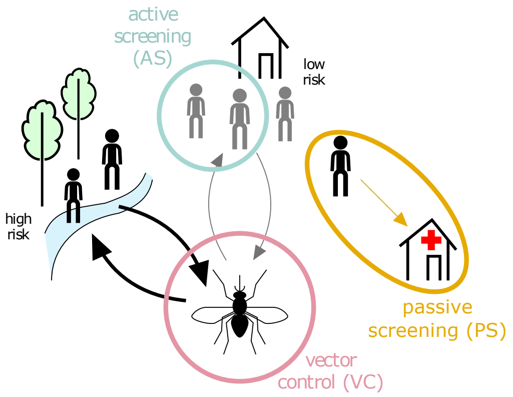

**Supplementary Figure 3**: Core gHAT intervention toolbox. Abbreviations: AS: active screening, PS: passive screening, RS: reactive screening, VC: vector control.

| Intervention | Description |
|---|---|
| **Active screening (AS)** | Mobile teams travelling to at-risk villages to test any person willing to participate |
| **High risk** | Individuals with greatest risk of gHAT infection |
| **Intensified (Int.) active screening** | Screening coverage (% people) at either the historic maximum or at 30% if the historic maximum is lower than this value |
| **Intervention** | Tools, treatments or approaches used to prevent or treat the infection |
| **Low risk** | Individuals with lowest risk of gHAT infection |
| **Mean active screening** | Screening coverage (% people) at the mean of the last five years for a region |
| **Passive screening (PS)** | Testing self-presenting individuals for gHAT at fixed health facilities |
| **Reactive screening (RS)** | Testing in specific locations in response to cases detected through passive screening |
| **Treatment** | Treatment of confirmed cases with either fexinidazole (oral drug course) if eligible, or pentamidine or NECT. Acoziborole (oral single-dose cure) may be used in the future if approved but is not considered in this analysis. |
| **Vector control (VC)** | Methods used to reduce the abundance of the vector, i.e. tsetse, that transmit infection. |
| **Targeted vector control (VC)** | An adapted method based on that previously used by LSTM to identify areas with high case density at which to focus Tiny Target deployment efforts along large rivers. |
| **Full Vector control (VC)** | Considers the deployment of Tiny Targets throughout all large rivers in a health zone. |

**Supplementary Table 8**: Intervention components which make up key strategies

##### A.7.1 Interventions

The definition of the interventions is shown in Supplementary Table 8 and depicted in 3. A summary of the levels of each of the interventions are in Supplementary Table 9.

##### Active Screening

Historically, AS has been the flagship activity of gHAT control, and therefore we simulate AS at two coverage levels: *Mean AS* which is the average coverage of the most recent five years in the data (i.e. 2016–2020), and *Intensified AS* which is the higher of the historical maximum (in 2000–2020) or 30% of the health zone. This *Int. AS* option was selected because the historical maximum is quite low in many health zones (less than 5% of people screened in 45 analysed health zones).

Two levels of active screening coverage are considered in our strategy sets. They are *Mean AS* and *Int. AS* in the general set, and *No AS* (sometimes skipped in the strategy names) and *Int. AS* for the special set for the Bas Uélé region. *Mean AS* represents maintaining AS at a recent level, we used the average numbers of people screened between 2016–2020. *Int. AS* refers to the realistic maximum level of AS that can be achieved in each health zone. For health zones outside the Bas Uélé region, it was assumed to be the higher of the historical maximum (in 2000–2020) or 30% of the health zone population in 2018. For health zones inside the Bas Uélé region, the *Int. AS* is assumed to be the average number of people screened between 2000–2014 because of the absence of recent HAT control activities. The values of *Mean AS* and *Int. AS* are calculated individually in each health zone. Finally, *No AS* means no activity at all in the future.

Although AS is organised at the coordination level, these units are mobile, require limited start-up costs to expand, can be scaled back, and often serve more than one health zone, so considering the resource use by the health zone will accurately reflect the collective costs at the coordination level.

**Cessation of active screening.** The planning of AS in the DRC is done by the end of the year and follows the WHO guidelines. AS should continue at the village level until 3 consecutive years of no case detection, and then pause for 1 year before a final screening in the fifth year. In other words, this is a two-step cessation algorithm for AS. The first step happens after no case reporting for 3 years, and the second step happens after no case reporting for 5 years. In our simulations, we assumed both steps occur in the future. To ensure *Mean AS* is a distinct strategy from *Int. AS*, we further assumed the earliest possible year for the first step cessation is 2024 and 2025 for strategies with mean and intensified coverage respectively.

**Reactive screening to case detections after cessation.** Reactive screening (RS) is the screening activities that react to any passive cases reporting after AS ceases. Its cessation also follows the same WHO guidelines explained above. Therefore, RS has a minimum length of 5 years (with 4 screening activities), but the actual duration depends on case finding.

##### Passive Screening

PS is simulated to continue at estimated 2020 levels into the future; this level reflects some improvements over the period of 2000-2020, estimated by model fitting. We do not include additional future improvements to the PS system. We used the 2019 WHO survey of clinics to calculate the cost of PS with two exceptions: for coordinations Bandundu Nord and Bandundu Sud. In those coordinations, in the provinces of Mai-Ndombe, Kwilu, and Kwango, there were ongoing trials at the time to expand the network of health posts that would have RDTs available for serological diagnosis of HAT. Patients would then be referred to the health zone's health centre for confirmation, staging (when necessary) and treatment [28]. However, that programme has since ceased, and most of those clinics are no longer getting support. Therefore, we have used the number of PS clinics that were listed for Bandundu Nord and Sud in the survey that was published for the year 2012 [35].

**Passive screening, including post-elimination surveillance.** Although we did not consider cessation of PS in the transmission model, we assumed that PS scales back to one health centre per health zone in the cost model after 5 years of no case reporting and this assumed post-elimination surveillance is equally effective as current PS (ignoring the possible impact of losing skilled clinicians and population awareness as the disease becomes rare).

##### Future VC

VC activities have been shown to decrease both tsetse populations and case reports in other locations [26, 36, 37], and therefore we simulate the addition of VC activities at two levels. In this analysis, any strategies including *Targeted VC* use an adapted algorithm based on that previously used by the Liverpool School of Tropical Medicine (LSTM) to identify areas with high case density at which to focus Tiny Target deployment efforts along large rivers. Strategies including *Full VC*, by contrast, involve a substantial expansion of VC interventions, considering the deployment of Tiny Targets throughout all large rivers in a health zone.

For our future strategies, we consider three possible levels of VC: *No VC*, *Targeted VC* and *Full VC*. The *Full VC* strategy consists of treating the banks of all candidate rivers in a health zone with tiny targets. Candidate rivers are either “large rivers", which we defined as those with an average long-term discharge estimate for river reach of $> 20m^3/s$, or any other rivers where VC has historically been deployed.

To select rivers for the *Targeted VC* strategy, we use a similar method to our coverage calculations, buffering each river segment by 5 km in every direction and taking the intersection of this buffer and the health zone in question. In all areas other than Bas Uélé, we then take all geolocated cases from 2016–2020 and intersect them with these buffered river segment areas. The number of cases falling in each buffered area is then counted; any cases that fall into multiple health buffers are split fractionally and equally between them so that they are counted as one case in total. We count "river length to be treated" as any river segment with a case density of at least 1 case per 10 km of treated riverbank. In the Bas Uélé region, we instead use cases from 2010 to 2014 since there was no capacity for detecting cases in this region between 2016–2020.

If our *Targeted VC* and *Full VC* scenarios have the same length of riverbank and case coverage, and VC has historically been deployed in the area, then instead of using the above calculations for *Targeted VC*, we use the maximum historical deployment length and case coverage of VC.

To compute the case coverage for the *Targeted VC* and *Full VC* strategies, we select the relevant rivers as above, then split them at any junctions, and prune any dead-end segments of less than 10 km. The rivers are then recombined and split again at health zone boundaries or any junctions between remaining river segments.

To calculate the length of the riverbank treated for each river segment, we apply the following steps. If a segment falls entirely along or within 5km of a health zone boundary (inside or outside), we treat it as if one bank (the interior bank) is being treated. If a segment falls entirely on the interior of the health zone (more than 5km from the boundary), we count it as having both banks treated. If it lies both near a boundary and also on the interior of the health zone, we count both banks on the portion in the interior, and one bank in the portion near the boundary. We then sum the length of the banks as above to estimate the total length of the riverbank where control is deployed under the strategy.

**Cessation of vector control.** Cessation of vertical interventions (which include vector control) was modelled as stopping based on consecutive years of zero detected cases by any screening modality. For VC, we used three years of no case detections (without the requirement for the fifth year of activity like with AS). Reactive VC is not considered in our simulations.

In addition to the criterion based on case reporting, we also considered a minimum of 3 years of VC to reflect the operational constraints such as training new people and sensitization to reach its maximum impact. In other words, we assume VC cessation can occur when there are no reported cases for three consecutive years. As the earliest possible year to scale back VC varies depending on the history of VC in individual health zones, our simulations do not allow cessation to happen before the analytical present and earlier than 2025 in order to ensure strategies with VC in the name have at least 1 year of VC. In our simulations, we checked simulated case numbers by the end of each year to decide whether the cessation criterion has been met from 2025 (for health zones with VC already, listed in Supplementary Table 7) or 2028 (for rest health zones). We assumed VC will stop from the following year if the cessation criterion is met and no reactive VC will take place to react to any simulated case reporting after scaling back.

##### Treatment

Historically, gHAT treatment could take place only in these referral hospitals and in many cases, the only health facilities where one would self-report for diagnosis with serological and parasitological tests and staging of the disease. In 2020, with the availability of the simpler oral treatment of fexinidazole treatment could be administered within the health areas or villages where the individual cases live for those individuals that do not need hospitalization according to recommendations [38].

In this analysis, Fexinidazole is simulated as being available immediately, but acoziborole – which is not yet approved for use outside clinical trials – is not included in this analysis. Further details on the treatment classifications and costs are described in section A.10.4.

| Coordination | PS clinics | Sum PS clinics | Sum PS clinics, in analysis | AS mean | AS historical maximum | AS total (thou-sands) | VC bank targeted (km) | VC bank full (km) |
|---|---|---|---|---|---|---|---|---|
| Bandundu Nord | 8 [2-21] | 169 | 54 | 0.18 [0-0.45] | 0.31 [0.02-0.64] | 621 | 83 [0-409] | 279 [0-916] |
| Bandundu Sud | 5 [0-25] | 111 | 39 | 0.09 [0-0.61] | 0.17 [0-0.74] | 699 | 11 [0-417] | 272 [26-483] |
| Equateur Nord | 1 [0-4] | 28 | 34 | 0.02 [0-0.12] | 0.22 [0.05-0.41] | 186 | 0 [0-0] | 161 [0-394] |
| Equateur Sud | 1 [0-4] | 12 | 13 | 0.01 [0-0.06] | 0.1 [0-0.2] | 22 | 0 [0-15] | 181 [25-892] |
| Isangi - Bas-Uélé | 0 [0-0] | 0 | 2 | 0 [0-0] | 0.21 [0.21-0.22] | 0 | 377 [0-753] | 890 [320-1460] |
| Isangi - Tschopo | 2 [0-11] | 16 | 17 | 0.09 [0.01-0.11] | 0.17 [0.04-0.21] | 61 | 108 [0-142] | 237 [197-300] |
| Kasai Occidental | 1 [0-3] | 17 | 23 | 0.02 [0-0.17] | 0.06 [0-0.23] | 143 | 0 [0-113] | 185 [0-639] |
| Kasai Oriental | 2 [0-8] | 54 | 57 | 0.02 [0-0.13] | 0.05 [0.02-0.23] | 182 | 0 [0-89] | 94 [0-329] |
| Kinshasa | 2 [0-6] | 24 | 28 | 0 [0-0.07] | 0.01 [0-0.1] | 37 | 11 [0-69] | 21 [11-542] |
| Kongo Central | 4 [0-11] | 81 | 82 | 0.01 [0-0.12] | 0.05 [0.01-0.27] | 53 | 0 [0-25] | 72 [0-379] |
| Maniema Katanga | 1 [0-2] | 9 | 14 | 0.02 [0-0.08] | 0.06 [0-0.12] | 94 | 0 [0-144] | 235 [0-628] |
| Sankuru | 1 [0-3] | 10 | 12 | 0.03 [0-0.23] | 0.08 [0-0.27] | 64 | 0 [0-190] | 258 [94-564] |
| **Total** | **2 [0-25]** | **531** | **375** | **0.02 [0-0.61]** | **0.09 [0-0.74]** | **2162** | **0 [0-753]** | **175 [0-1460]** |

**Supplementary Table 9**: Summary of the screening and vector control activities in health zones in the analysis, stratified by the coordinations delineated the programme national de lutte contre la Trypanosomiase humaine africaine (PNLTHA-RDC). Distributions are the median followed by the minimum and maximum values for health zones. The number of PS clinics in the analysis and the number in the 2019 WHO survey differ for two reasons. First, in Bandundu Nord and Bandundu Sud, TrypElim had expanded the network of clinics with capacity for serological confirmation followed by a referral to the main hospital, but the referral system did not work as most patients were lost-to-follow-up. For more information, see Snijders et. al [28]. Therefore, the clinics we assume can still diagnose and treat patients were equivalent to the clinics reported before the TrypElim project in Bandundu: 54 clinics in Bandundu Nord and 40 clinics in Bandundu Sud [35]. Second, we have assumed all health zones have at least one clinic that can screen for HAT with an RDT or a CATT test; even if the clinic is not present in that health zone there is a clinic in a nearby health zone committing resources to screen patients from the health zone in question. Abbreviations: HZ: health zone, PS: passive screening, AS: active screening, VC: vector control.

##### A.7.2 Strategies

There are two sets of strategies considered from the analytical present (i.e. from 2026 onward) for the special set for health zones in the Bas Uélé region and the general set for the rest of the analysed health zones (see Figure 1(b) and 1(c) in the main text). The Bas Uélé region had a very distinct strategy history (i.e. no HAT control activities such as active and PS since MSF left in 2015) due to operational feasibility, and therefore the general strategies are not suitable nor feasible for health zones in the Bas Uélé region.

The strategies are listed in Figure 1B for 163 health zones, and in Figure 1C for the two health zones of Bas Uélé region that required different strategies. The health zones are characterized by six typologies, which determine the strategies simulated and the comparator:

| Category | Strategies | Comparator | No. HZ |
|---|---|---|---|
| 1 | All six | *Mean AS* | 25 |
| 2 | All six | *Mean AS + Targeted VC* | 12 |
| 3 | No Targeted VC | *Mean AS* | 116 |
| 4 | No VC | *Mean AS* | 10 |
| 5 | Alternative eight | *No AS, PS, nor VC* | 1 |
| 6 | Alternative six | *No AS, PS, nor VC* | 1 |

Six typologies of health zones, depending on the strategies simulated and the comparator. Abbreviations: AS: active screening, PS: passive screening, VC: vector control.

1. **All six strategies, *Mean AS* is the comparator.** Health zones where there is no current VC but where there is potential for targeted VC based on recent case clustering near large rivers have a status quo strategy of *Mean AS* which includes PS in fixed health facilities and mobile screening activities that screen a number of the population equal to the mean number screened in AS in 2016–2020. One of these health zones, Bolobo, had VC in the recent past but has no ongoing VC. The five additional strategies are made up of different combinations of AS and VC – *Int. AS*, *Mean AS + Targeted VC*, *Int. AS + Targeted VC*, *Mean AS + Full VC*, and *Intensified AS + Full VC* – are simulated to compare the health benefits and the costs against this status quo strategy. The number of health zones in this category is 25.
2. **All six strategies, *Mean AS + Targeted VC* is the comparator.** Health zones where there is ongoing VC, will have a status quo strategy of *Mean AS + Targeted VC* but we still run the six strategies. The *Mean AS* and *Intensified AS* strategies would imply cessation of the current VC activities. Health zones with historical and ongoing deployments are Bagata, Bandundu, Bokoro, Bolobo, Kwamouth and Yumbi in Bandundu Nord and Bulungu, Kimputu Masi Manimba, Mokala in Bandundu Sud. Yasa Bonga in Bandundu Sud has had historical VC but the extent has approximated Full VC, so the comparator is *Mean AS + Full VC*. Kikongo started as VC in mid-2019 and switched to 'community' deployment in 2022, but we ran it as if nothing had changed. The total number of health zones in this category is 12.
3. **Four strategies; no Targeted VC strategies.** In health zones where there is no clear way to perform targeted VC, as the recent cases are not sufficiently geographically clustered– but where there are large rivers that have a status quo strategy equivalent to *Mean AS*. We still simulate VC along all large rivers and so the three additional strategies are: *Intensified AS*, *Mean AS + Full VC*, and *Intensified AS + Full VC*. The number of health zones in this category is 116.
4. **Two strategies; no VC strategies.** For health zones where no major rivers were detected during the analysis, we only simulated the two strategies that exclude VC. *Mean AS* is the comparator. These health zones were: Ntand Embelo in Bandundu Nord; Gemena and Tandala in Équateur Nord; Katende in Kasaï Occidental; Kabinda, Mpokolo, and Nzaba in Kasaï Orientale; Muanda and Seke Banza Kongo Central; and Mbulula in Maniema-Katanga for a total of 10 health zones.
5. **Bas Uélé region; eight and six alternative strategies.** Health zones in the Bas Uélé region including Ango and Doruma have an alternative set of strategies as shown in Figure 1C. These health zones have an alternative set of strategies because activities there were formerly run by an international organization, Médecins sans frontières (MSF), rather than under the purview of the national programme. MSF ran screening with alternative diagnostic algorithms [3] and therefore the data are interpreted separately. Since the departure of MSF, due to challenges in access, the national programme has not had the opportunity to run activities

there, and therefore, there is less data and more uncertainty about transmission in this region. As a result, we model activities that would keep the status quo (*No AS, PS, nor VC*) as the comparator, putting into place activities similar to the rest of the country, and conducting additional VC:

(a) in Ango: *Int. AS* + *no PS*, *Restart PS*, *Int. AS* + *restart PS*, *Targeted VC* + *restart PS*, *Int. AS* + *Targeted VC* + *restart PS*, *Full VC* + *restart PS*, *Int. AS* + *Full VC* + *restart PS*.

(b) Doruma has cases that are so diffuse throughout the health zone that the algorithm to determine VC extents was not able to find any hotspots. Therefore, no Targeted VC strategies were simulated.

**Note on the interpolation for the period of 2021-2023.** For those years of 2021-2023, for which decision analysis does not make sense but for which we did not have data at the time of analysis, we do not assume alternative strategies. In other words, we assumed that PS and VC carried on as they were, and actively screened covered populations at the average level from 2016–2020 during the period between the end of the data period and the analytical present (i.e. in 2021–2023).

#### A.8 Cost-effectiveness analysis

↩ Return to the Supplement Table of Contents

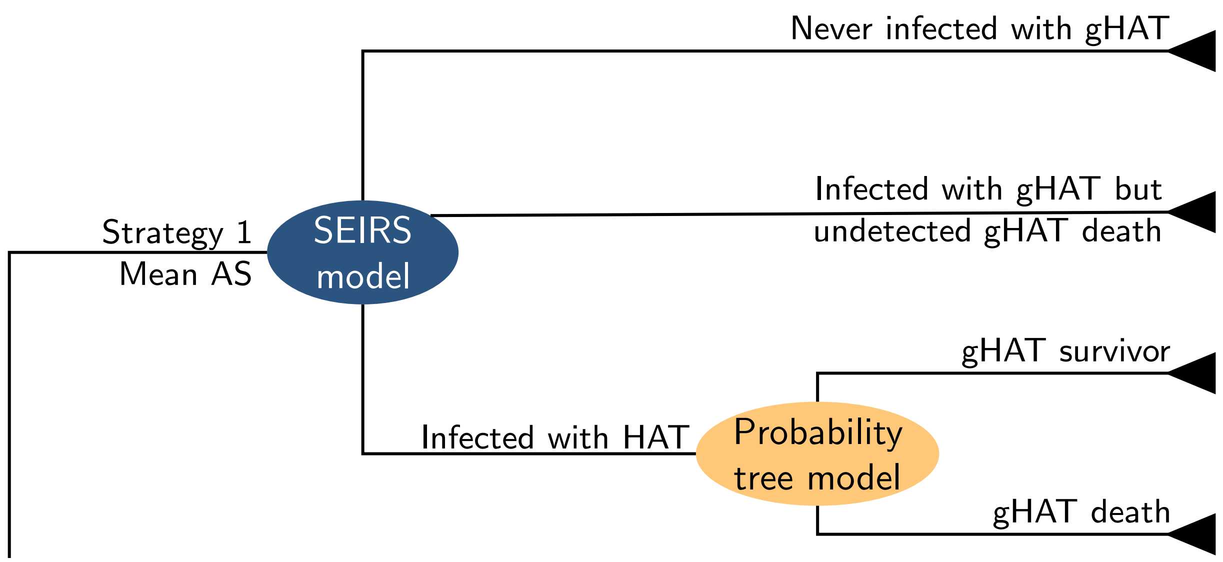

**Supplementary Figure 4**: Decision tree, close up of Supplementary Figure 5. Decision nodes are square, and probabilistic nodes are ellipses. The transmission model is depicted in Supplementary Figure 2 and the probability tree model is depicted in Supplementary Figure 6. Abbreviations: SEIRS: susceptible-exposed-infected-recovered-susceptible dynamic model of transmission, AS: active screening, PS: passive screening, RS: reactive screening, VC: vector control.

Lastly, we calculate costs, treatment outcomes and disease burden to estimate the cost-effectiveness of each of the strategies according to a decision tree shown in Figure 4 and shown in more full in Figure 5. The costs here are denominated in 2024 USD, and health burden and benefits are denominated in disability-adjusted life-years, although we also present cases and deaths as intermediate outcomes. We have selected to show costs, benefits, and cost-effectiveness taking into account a 2040 horizon to calculate the costs of activities until 2030, the goal date for elimination of transmission, and to be able to take into account any savings from scale-back of activities that may stretch into the decade following 2030.

(a) A) General set of strategies.

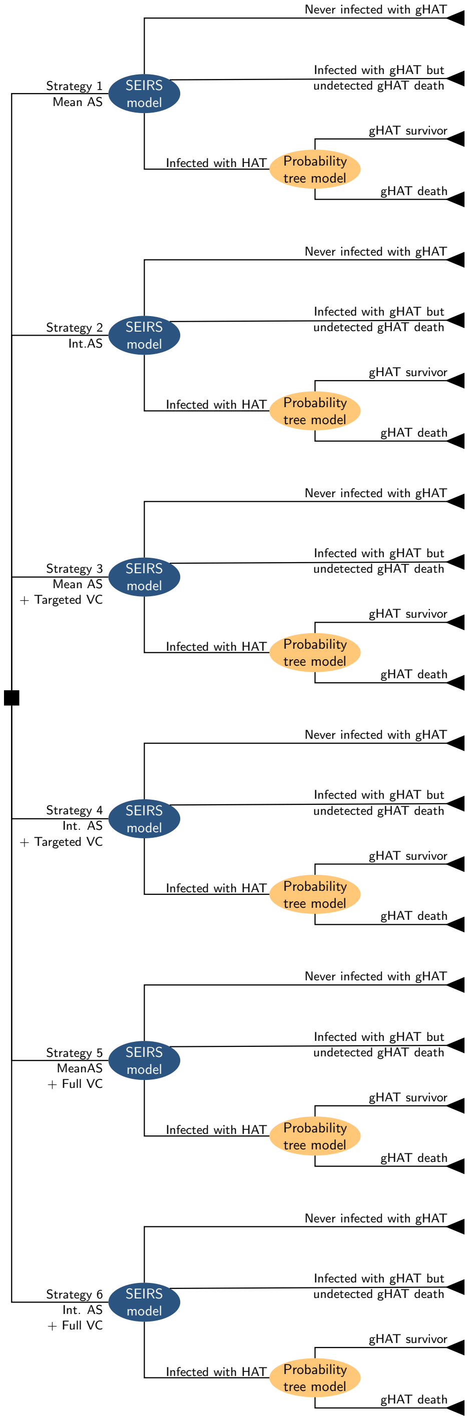

(b) B) Special set of strategies (for Bas Uélé region).

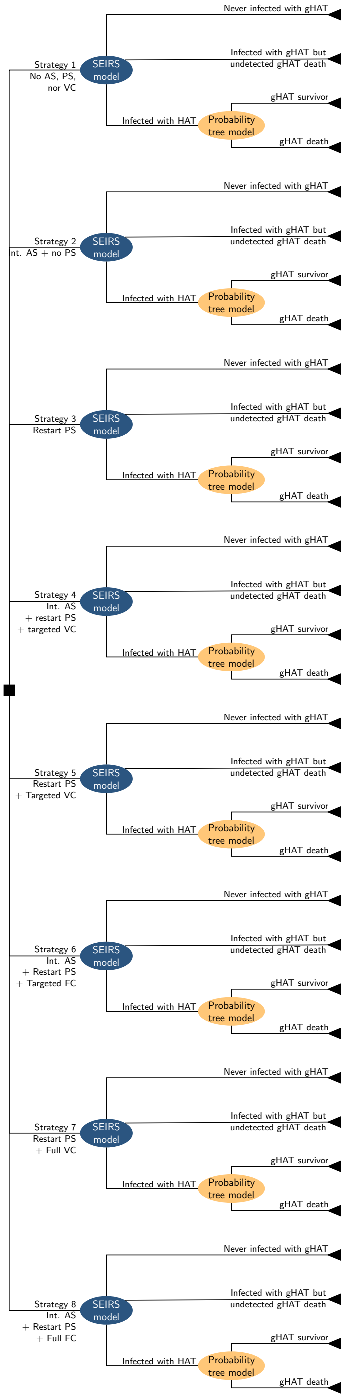

**Supplementary Figure 5**: Decision tree of disease suppression and prevention strategies. Decision nodes are square, and probabilistic nodes are ellipses. One branch (one strategy) is depicted in detail in Supplementary Figure 4. The transmission model is depicted in Supplementary Figure 2 and the probability tree model is depicted in Supplementary Figure 6. A) Strategies against gHAT, including active screening (AS) by mobile teams, passive screening (PS) in fixed health facilities, and vector control (VC). In three strategies (*Mean AS*, *Mean AS* + *Targeted VC*, and *Mean AS* + *Full VC*) the proportion screened equalled the mean number screened during 2016–2020. In the three other strategies (*Int. AS*, *Int. AS* + *Targeted VC*, *Int. AS* + *Full VC*), the coverage is the maximum number screened during 2000–2020. In strategies 3–6, vector control (VC) is simulated assuming a reduction as described in the Supplementary Methods, Section A.7.1. PS is in place under all strategies. B) Strategies against gHAT in Bas Uélé region. Abbreviations: SEIRS: susceptible-exposed-infected-recovered-susceptible dynamic model of transmission, AS: active screening, PS: passive screening, VC: vector control.

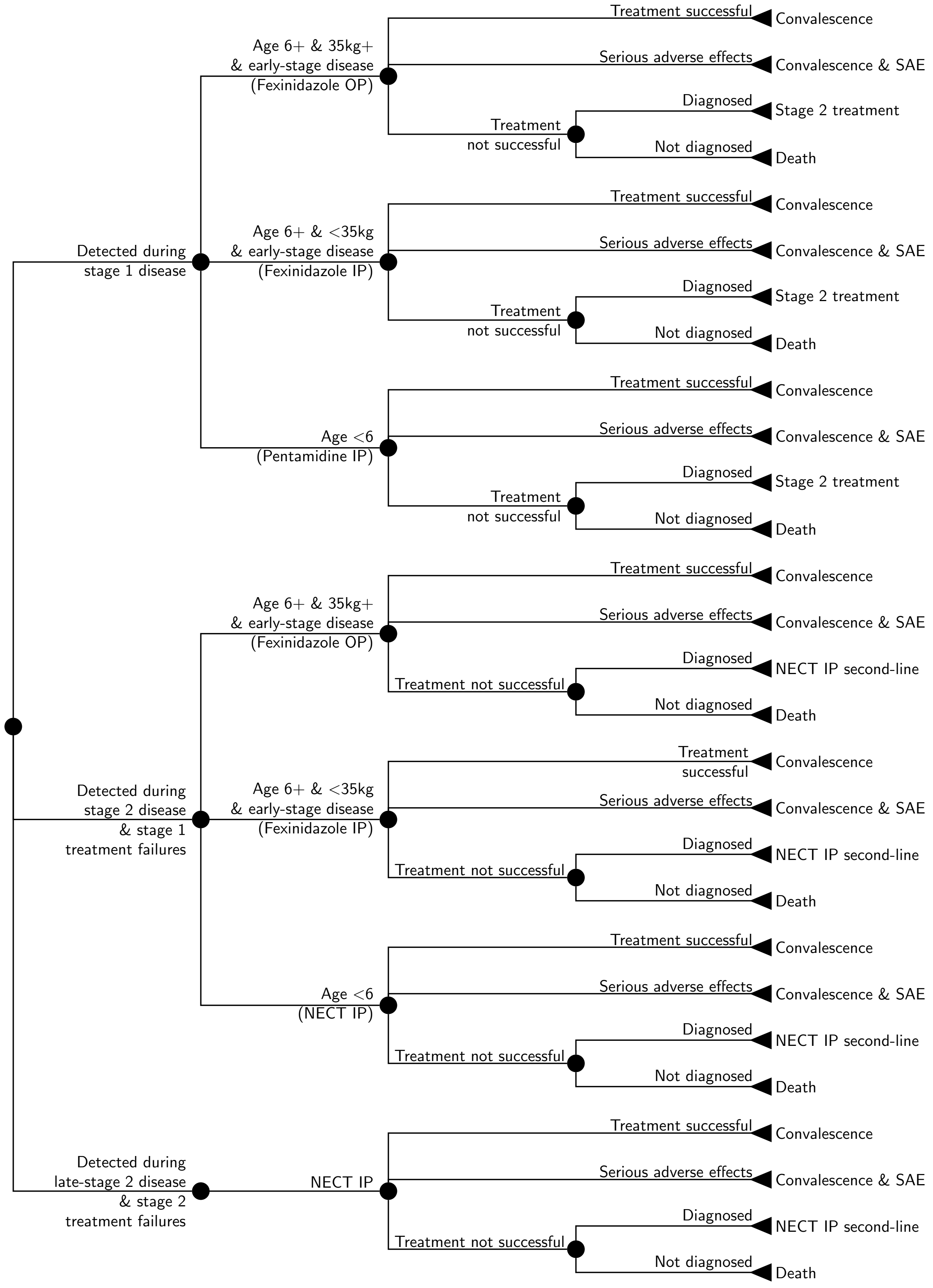

**Supplementary Figure 6**: Treatment model. Treatment for diagnosed gHAT patients is modelled as a branching tree process of possible health outcomes, including eligibility for novel Fexinidazole. Abbreviations: SAE: Serious adverse events, IP: inpatient care, OP: outpatient care, NECT: nifurtimox-eflornithine combination therapy.

#### A.9 Health outcomes denominated as disability-adjusted life-years

↩ Return to the Supplement Table of Contents

(Please note that sections A.9-A.10.4 contain some amount of recycled text from the authors' previous publications [6, 27].)

For incremental cost-effectiveness calculation, we define the health effects of gHAT interventions in terms of disability-adjusted life-years (DALYs) [39, 40]. Using DALYs is in line with the recommendations of the Bill and Melinda Gates Foundation's reference case and WHO's guidelines for the conduct of cost-effectiveness analyses [39–41]. DALYs allow policy-makers to compare interventions across different disease programs with one common metric.

DALYs were discounted at a rate of 3% per year [40, 41]. We follow established conventions to calculate DALYs and evaluate the estimates in present-day terms (after applying discounting) [40–42].

A more general discussion of this is found in [6] (Supplementary Methods, pages 10-11), but we provide a brief description here for convenience.

$$\text{Disability-adjusted life-years (DALY)} = \text{Years of life lost to disability (YLD)} + \text{Years of life lost due to death (YLL)}$$

$$\text{YLD} = (\text{YLDs before detection} + \text{YLDs during treatment} + \text{YLDs due to side effects}) \times \text{DALY weight}$$

where the DALY weight is a metric to measure the relative severity of gHAT compared to living with other diseases.

$$\text{YLL} = \text{life-expectancy at age of death} - \text{age of death from gHAT}$$

One important difference between the method described in [6] and the current study is that here we use the life-table method of accounting to calculate the years of life that a person with a fatal case would have lived absent gHAT. The previous study used the life-of-years lived at birth to less the age at death. In 2019 (the most recent year with data from the WHO), the life expectancy in DRC was 62.4 and the age of death from gHAT is 26.62 (See Section E.7.1 of this supplement). The life expectancy at age 25-29 is about 70 in DRC (usually, life expectancy is higher in older ages after surviving the risky first few years of life). Therefore, the current method estimates a slightly higher number of YLLs, and by consequence, DALYs, than the previous method.

#### A.10 Treatment outcomes and cost functions

↩ Return to the Supplement Table of Contents

The cost functions are described in this section separately for active screening, passive screening, vector control, and treatment. The health zone of Kikongo is used as an example to show intermediate calculations and final costs.

The total costs can be characterised by the expression

$$\text{Total costs} = \sum_{i \in \text{all sub-categories}} \left(U_i \times C_i\right)$$

$i$ is the cost sub-category. Where $U$ is the units of a resources(i.e. people screened, teams in operation, testing centres, etc) and $C$ is the cost per resource unit. All costs were denominated in 2024 US$. The process to update costs from the literature is detailed in Supplementary Note 3, Section E.1.

##### A.10.1 Active screening

Yearly costs of AS were calculated as a function of two groups of expenses: capital and management costs, as well as the costs of screening the population and confirming suspected cases. Because gHAT program activities are not combined with activities of any other disease programs, we employed a full costing, rather than an incremental costing, method. A more thorough explanation of active screening campaigns and their costs is available from Snijders and colleagues [43].

Notably, we do not include the costs of lumbar punctures in this portion of the analysis; rather, we include it in the costs of treatment. Because many patients are eligible for fexinidazole treatment, which does not require lumbar punctures, we include lumbar puncture costs in the treatment portion of the analysis for those patients who are not eligible for fexinidazole. See Supplementary Methods, section A.10.4.

- Overhead costs: overhead costs are split between capital costs and recurrent (management) costs to run an active screening team. AS teams serve a “coordination”– a subnational designation of the PNLTHA program that manages a set of health zones. Therefore, a health zone where fewer than 60,000 people are targeted for screening does not necessarily incur larger overhead costs for active screening than a health zone where the coverage is closer to a multiple of the yearly capacity of a team.
  - Capital costs consist of vehicles, medical equipment, energy (solar panels) and training (which occurs once every few years).
  - Recurrent costs consist of management and consumables that are spent on the team: fuel, staff time, etc.
- Costs related to the population screened:
  - Card Agglutination Trypanosomiasis Tests (CATT) are scaled up according to the number of people that are screened per year, and a factor of wastage of CATT tests is included.
  - Confirmation tests are counted for all of those who are positive according to the CATT test, including false positives (which are estimated as 1-specificity of the test) and true positives, calculated by the dynamic model.
- For all costs, the national PNLTHA is assumed to consume resources, and there is a mark-up to account for the central management at the national program headquarters. We have made the same assumption as Snijders and colleagues [43] who estimate that PNLTHA management equals approximately 15% of costs.

The parameters for AS are described in Supplementary Note 3: Parameter Glossary, section E.5 and the cost parameters are described in Supplementary Note 3: Parameter Glossary, section E.9.

The cost for Kikongo health zone per year, given the number of people screened in that health zone under Mean and Intensified AS, is therefore:

| Variable Name | Parameterization | Summary |
|---|---|---|
| AS coverage per year | Fixed | See Table 9 |
| Wastage factor for CATT administration in AS context | Beta(8, 92) | 0.08 (0.04, 0.14) |
| CATT specificity | Beta(4523, 22) | 1.00 (0.99, 1.00) |
| AS capital costs (annualized) | Gamma(820, 20.1) | 16,525 (15,405, 17,693) |
| AS recurrent costs (annualized) | Gamma(4153.88, 17.21) | 71,486 (69,309, 73,689) |
| Cost of CATT test | Gamma(1140, 0.0006897) | 0.79 (0.74, 0.83) |
| Cost confirmation (microscopy) | Gamma(8.47, 1.71) | 14.59 (6.49, 26.08) |
| Cost of delivery for CATT (markup) | Fixed, 0.10 | 0.10 |

**Supplementary Table 10**: Components of active screening costs

| Item | Units ($U$) | Cost ($C$) |
|---|---|---|
| Capital (annualized) | AS coverage per year ÷ patients screened by a team | AS capital × (1+PNLTHA markup) |
| Management/recurrent expenses | AS coverage per year ÷ patients screened by a team | AS recurrent × (1+PNLTHA markup) |
| CATT testing (See Note 1) | AS coverage per year (traditional) × (1+wastage factor for CATT in AS context) | CATT × (1+delivery mark-up) × (1+PNLTHA markup) |
| Microscopy/confirmation | (1-CATT specificity) × (AS coverage per year × Population) | Microscopy × (1+PNLTHA markup) |

[1] Ideally, CATT tests would be used for active screening and RDT tests would be used for passive screening because of the high wastage of CATT tests in the context of passive screening settings. We simulate screening costs as if screening in the context of AS takes place with CATT only.

**Supplementary Table 11**: Active screening: cost function. Abbreviations: AS: active screening, CATT: card agglutination trypanosomiasis test, PNLTHA: Programme National de Lutte contre la Trypanosomiase Humaine, the national control program housed at the Ministry of Health.

| Item | Units ($U$) | Cost per unit ($C$) | Cost per category |
|---|---|---|---|
| **Mean AS** | | | |
| Capital (annualized) | 0.74 (0.54, 1.07) | 16,525 (15,405, 17,693) | 12,245 (8,881, 17,789) |
| Recurrent expenses | 0.74 (0.54, 1.07) | 71,486 (69,309, 73,689) | 52,973 (38,583, 76,864) |
| Microscopy | 208 (131, 305) | 14.59 (6.49, 26.08) | 3,042 (1,173, 6,048) |
| CATT testing | 46,479 (44,558, 49,057) | 1.14 (1.04, 1.24) | 46,478 (44,557, 49,056) |
| **Total** | | | **114,737 (96,617, 144,383)** |
| **Int. AS** | | | |
| Capital (annualized) | 1.23 (0.90, 1.78) | 16,525 (15,405, 17,693) | 20,341 (14,753, 29,551) |
| Recurrent expenses | 1.23 (0.90, 1.78) | 71,486 (69,309, 73,689) | 88,001 (64,096, 127,690) |
| Microscopy | 346 (217, 506) | 14.59 (6.49, 26.08) | 5,053 (1,949, 10,048) |
| CATT testing | 77,213 (74,021, 81,496) | 1.14 (1.04, 1.24) | 77,212 (74,021, 81,495) |
| **Total** | | | **190,607 (160,504, 239,856)** |

**Supplementary Table 12**: Cost breakdown for active screening activities for 1 year in Kikongo health zone.

##### A.10.2 Passive screening

The costs of passive screening each year were calculated as a function of two groups of expenses: 1) overhead costs, and 2) the number of consultations, screening and confirmation tests that are performed in any clinic in the health zone capable of performing a serological or confirmatory test for a person that comes in with suspected gHAT according to the 2019 WHO survey of clinics in DRC [44]. The process of passive screening and its costs is described in more detail by Snijders and colleagues [28].

- Overhead costs: overhead costs include capital costs and recurrent (management) costs to equip a health centre to perform serological screening for HAT and microscopic confirmation, as well as to keep personnel in the clinics trained. These costs are scaled by the number of health clinics in the health zone capable of doing serological testing for gHAT [44].
  - Capital costs consist of medical equipment, energy (solar panels) and training (which occurs periodically every few years). These costs are scaled by the number of facilities that can perform serological confirmation.
  - Recurrent/management costs consist of health zone and provincial management and supervision.
- Costs that scale by population served:
  - Rapid diagnostic tests (RDT) are scaled up according to the number of people that are screened per year, including a markup to account for the wastage of tests.
  - Confirmation tests are accounted for all of those who are positive according to the RDT test: both false positives (which are modelled as a factor equal to 1-specificity of the test) and the true positives outputted by the dynamic model.
  - Lumbar punctures are not depicted as part of the passive surveillance diagnosis costs but are included as part of the treatment costs. Because many patients are eligible for Fexinidazole treatment, which does not require lumbar punctures, we include lumbar puncture costs in the treatment portion of the analysis for those patients who are not eligible for Fexinidazole. See Supplemental Methods, section A.10.4.
- For all costs, there is a PNLTHA mark-up added for the central management at the national program headquarters. Snijders *et al* [43] estimate that PNLTHA management equals approximately 15% of costs, and we have made the same assumption.

The parameters for PS are described in Supplementary Note 3: Parameter Glossary, section E.5 and the cost parameters are described in Supplementary Note 3: Parameter Glossary, section E.9.

| Variable Name | Parameterization | Summary |
|---|---|---|
| PS coverage per year per population per clinic | Fixed, by coordination, see E.5.2. Gamma(55.53, 1.55) for Bandundu Sud. | 85.96 (64.80, 110.05) |
| Wastage factor for RDT | Beta(1, 99) | 0.01 (<0.01, 0.04) |
| RDT specificity | Beta(1134, 11) | 0.99 (0.98, 1.00) |
| PS capital costs (annualized, per facility) | Gamma(8.475, 283.46) | 2,400 (1,069, 4,251) |
| PS management costs (annualized, per health zone) | Gamma(8.475, 145) | 1,228 (548, 2,169) |
| Cost of RDT test | Gamma(1140, 0.002205) | 2.51 (2.37, 2.66) |
| Cost confirmation (microscopy) | Gamma(8.475, 1.71) | 14.59 (6.49, 26.08) |
| Cost of delivery (markup) | 1.10 | 1.10 |

**Supplementary Table 13**: Components of passive screening costs

The cost for each health zone per year, given the number of people screened per health zone and the number of health centres available for PS, is therefore:

| Item | Units ($U$) | Cost ($C$) |
|---|---|---|
| Capital - clinics | Number of facilities capable of screening and confirmation within the focus | Capital costs (clinic) × (1+PNLTHA markup) |
| District management | Per district | District management costs × (1+PNLTHA markup) |
| OP visit | PS coverage per year per clinic × Clinics in the focus | OP visit cost × (1+PNLTHA markup) |
| RDT testing | PS coverage per year per clinic × Clinics in the focus × (1+wastage for RDT) | RDT × (1+delivery mark-up) × (1+PNLTHA markup) |
| Microscopy/confirmation (suspects first identified in screening-only sites) | (1-RDT specificity) × (PS coverage per year per clinic × Clinics in the focus) | Microscopy × (1+PNLTHA markup) |

[1] We assume that all testing in passive screening is done with RDT tests. This is not always true, but the costs will be approximately similar if the testing is done with CATT.

**Supplementary Table 14**: Passive screening: cost function. Abbreviations: PS: passive screening, RDT: rapid diagnostic test, OP: outpatient, PNLTHA: Programme National de Lutte contre la Trypanosomiase Humaine, the national control program housed at the Ministry of Health.

| Item | Units ($U$) | Cost per unit ($C$) | Cost per category |
|---|---|---|---|
| Capital - clinics | 3 | 2,400 (1,069, 4,251) | 7,199 (3,207, 12,754) |
| Management | 1 | 1,228 (548, 2,169) | 1,228 (548, 2,169) |
| OP visit | 258 (194, 330) | 3 (2, 4) | 811 (475, 1,243) |
| RDT | 261 (196, 333) | 2.64 (2.49, 2.79) | 687 (514, 887) |
| Microscopy for false positives | 2.48 (1.16, 4.40) | 14.59 (6.49, 26.08) | 36.10 (11.83, 79.59) |
| **Total** | | | **9,961 (5,842, 15,572)** |

**Supplementary Table 15**: Cost breakdown for passive screening activities for 1 year in Kikongo health zone. The same calculation were made and shown in Antillon et. al [6] and [27], but these calculations feature specific parameters for Kikongo (and every other health zone in this analysis) as well as costs updated to 2024 USD values. Abbreviations: RDT: rapid diagnostic test, OP: outpatient.

##### A.10.3 Vector control

The costs of vector control (VC) were calculated as a function of two features: 1) the extent of the rivers where VC is deployed, and 2) the number of targets per kilometre of river where the targets were deployed. The costs of entomological surveys (tsetse monitoring), sensitisation of the population (information campaigns), and district management were assumed to scale with the extent of the health zone where VC would be deployed. The materials and labour time related to target deployment were scaled according to the number of targets deployed. A full costing method was used. A more detailed account of vector control in DRC is given in publications by Tirados and colleagues [30] and the costs are detailed in Snijders and colleagues' publication [45].

The extent of the riverbank that is necessary to cover for adequate coverage is determined by the case reports of the previous five years. The method is further explained in Supplementary Methods, Section A.7.1. Thirty targets per kilometre are assumed to be used, per current practice in DRC.

The parameters for VC are described in Supplementary Note 3: Parameter Glossary, section E.8 and the VC cost parameters are described in Supplementary Note 3: Parameter Glossary, section E.11.

| Variable Name | Parameterization | Summary |
|---|---|---|
| Linear km of river bank | Fixed | Varies by health zone. See section E.8.1. For Kikongo, it is 79 km for targeted VC and 601 km for full VC. |
| Units per km of river bank | Fixed | 15 |
| Deployments per year | Fixed | 2 |
| Cost for entomological surveys, sensitisation and district management per kilometer | Gamma(8.475, 57.7) | 487 (216, 868) |
| Cost per target deployment per target | Gamma(8.475, 0.50) | 4.23 (1.88, 7.49) |
| PNLTHA markup | Uniform(0.1, 0.2) | 0.15 (0.10, 0.20) |

**Supplementary Table 16**: Components of vector control costs

| Item | Units ($U$) | Cost ($C$) |
|---|---|---|
| Entomological surveys, sensitization and management | Kilometers of river covered | Cost for entomological surveys, sensitisation and district management per kilometer × (1+PNLTHA markup) |
| Target deployment | Kilometers of river covered × Targets per kilometer × Number of deployments per year | Cost for target deployment per target × (1+PNLTHA markup) |

**Supplementary Table 17**: Vector control: cost function. Abbreviation: PNLTHA: Programme National de Lutte contre la Trypanosomiase Humaine, the national control program housed at the Ministry of Health.

Per year, the simulated costs according to the above formulation and parameters result in the following estimates:

| Item | Units ($U$) | Cost ($C$) |
|---|---|---|
| **Targeted VC** | | |
| Entomological surveys, sensitization and management | 79 km | 44,222 (19,548, 78,966) |
| Target deployment | 4740 targets | 23,046 (10,188, 40,814) |
| **Total** | | **61,502 (35,541, 95,120)** |
| **Full VC** | | |
| Entomological surveys, sensitization and management | 601 km | 336,422 (148,713, 600,741) |
| Target deployment | 36060 targets | 175,327 (77,508, 310,497) |
| **Total** | | **467,883 (270,382, 723,635)** |

**Supplementary Table 18**: Cost breakdown for vector control activities

##### A.10.4 Treatment

Traditionally, the stage of disease determined by microscopic examination of the cerebro-spinal fluid, which is extracted via lumbar puncture for cases that have been confirmed by visualisation of the trypnosome (e.g. cases in which trypanosomes are present in the blood). If trypanosomes are present in the cerebro-spinal fluid, the patient is considered to have stage 2 disease; if not, he or she is considered to have stage 1 disease. According to the stage of disease, patients are referred to the appropriate health centre or health district hospital for treatment.

In the context of fexinidazole treatment, which has been present in DRC since 2020, lumbar punctures will not be performed by the active screening team, but once patients are referred to a health centre, the health centre will determine eligibility for fexinidazole treatment.

We assumed the treatment algorithm based on the WHO interim recommendations of 2019 [38].

- **Step 1, Group A. Patients without clinical symptoms of severe gHAT.** These patients would be eligible for fexinidazole treatment if their presentation fulfils the following criteria:
  - **Patient age** $<$ **6 years or weight** $<$ **20 kg.** These patients would be ineligible for Fexinidazole treatment. For simplicity, we assumed that all patients over 6 years old were also over 20 kg due to scant data on patient characteristics. See Step 2, Group A.
  - **Patient age** $>$ **6 years old and weight** $>$ **20 kg.** These patients would be eligible for Fexinidazole treatment. See Step 2, Group B. WHO recommendations stipulated that a doctor ought to be certain of the adherence on the part of the patient in order to prescribe fexinidazole on an outpatient basis. For simplicity, we have assumed that this is not an issue because it doesn't make a substantial difference in the total costs or effects of this particular analysis.
- **Step 1, Group B. Patients with clinical symptoms of severe HAT.** Patients whose clinical assessment would be consistent with severe gHAT (see Annex 1 of the WHO Interim guidelines [38]) would undergo a lumbar puncture to determine the concentration of white blood cells (WBCs) in the cerebrospinal fluid. For a concentration $<$ 100 WBC per microlitre ($\mu$L), the patient would be considered eligible for Fexinidazole treatment, depending on age and weight, as detailed in Step 1, Group A. In our model, we assumed that no stage 1 patient would show more than 100 WBC/$\mu$L of CSF as no trypanosomes should be present in the CSF. Moreover, we assumed that some proportion of stage 2 patients will be in late-stage disease (see Supplementary Tables 19-20).
- **Step 2, Group A. Patients ineligible for Fexinidazole treatment.** Patients age $<$ 6 years old or weight $<$ 20 kg are assumed to submit to a lumbar puncture to determine disease stage with 100% adherence. In our treatment model, we consider the cost of a lumbar puncture in these patients (see Supplementary Table-23) but we take the outcome of the lumbar puncture (stage 1 or stage 2) from the transmission model, the stage of disease is a critical output of the transmission model.
  - WHO recommendations stipulate the following criteria: if there is no trypanosoma in the CSF, then the patient undergoes Pentamidine treatment on an inpatient basis. If there are more than 5 leukocytes (or WBC)/$\mu$L then the patient undergoes NECT treatment on an inpatient basis.
  - Pentamidine treatment would consist of intra-muscular infections for 7 days.
  - NECT (Nifurtimox-eflornithine combination therapy): Nifurtimox is administered orally for 10 days, while Eflornithine is administered intravenously for 7 days [38].
- **Step 2, Group B. Patients eligible for Fexinidazole treatment.** We assumed that patients would be treated on an inpatient basis if age $>$ 6 years old and 20 kg $<$ weight $<$ 35 kg, otherwise, they would be treated on an outpatient basis as directly observed therapy.

**Uncertainty** Uncertainty in the treatment and cost parameters is parameterized according to the standard errors of estimates in the literature (see Supplementary Note 3: Parameter Glossary, sections E.6 and E.10).

We show here the components of the costs per case treated, depending on the stage and the treatment. The parameters for the above table are available in Supplementary Note 3: Parameter Glossary, section E.10 and eligibility distributions are described in Supplementary Table 20.

| Patient Characteristic | Parameterization | Summary |
|---|---|---|
| Under 6 years old | Beta(152.5, 2427.9) | 0.06 (0.05, 0.07) |
| Under 35 kg of weight | Beta(8.3, 359.6) | 0.02 (<0.01, 0.04) |
| Late stage-2 disease | Beta(76.9, 44.9) | 0.63 (0.54, 0.72) |

**Supplementary Table 19**: Parameters for treatment eligibility. Reproduced with permission from [6] under a CC-BY license.

| Eligibility | Rationale | Summary |
|---|---|---|
| **Stage 1** | | |
| Pentamidine | Under 6 years old (1) | 0.06 (0.05, 0.07) |
| Fexinidazole-inpatient | Over 6 years old but under 35 kg of weight | 0.02 (<0.01, 0.04) |
| Fexinidazole-outpatient | Over 6 years old and over 35 kg of weight | 0.92 (0.90, 0.93) |
| **Stage 2** | | |
| NECT | Under 6 years old or late-stage disease | 0.65 (0.57, 0.73) |
| Fexinidazole-inpatient | Over 6 years old but under 35 kg of weight and early stage-2 disease | <0.01 (<0.01, 0.01) |
| Fexinidazole-outpatient | Over 6 years old, over 35 kg of weight, and early stage-2 disease | 0.34 (0.26, 0.42) |

[1] For simplicity, all patients over 6 years old were assumed to be over 20 kg in weight.

**Supplementary Table 20**: Eligibility for treatment

| Treatment | Outcomes | Estimate |
|---|---|---|
| **Stage 1** | | |
| Pentamidine | Cured | 0.05 (0.05, 0.06) |
| | Cured with SAEs | <0.01 (<0.01, <0.01) |
| | Rescue treatment | <0.01 (<0.01, <0.01) |
| | Death | <0.01 (<0.01, <0.01) |
| Fexinidazole - inpatient | Cured | 0.02 (<0.01, 0.04) |
| | Cured with SAEs | <0.01 (<0.01, <0.01) |
| | Rescue treatment | <0.01 (<0.01, <0.01) |
| | Death | <0.01 (<0.01, <0.01) |
| Fexinidazole - outpatient | Cured | 0.89 (0.87, 0.91) |
| | Cured with SAEs | 0.01 (<0.01, 0.02) |
| | Rescue treatment | 0.02 (<0.01, 0.03) |
| | Death | <0.01 (<0.01, <0.01) |
| **All treatments** | **Cured** | **0.97 (0.95, 0.98)** |
| | **Cured with SAEs** | **0.01 (<0.01, 0.02)** |
| | **Rescue treatment** | **0.02 (0.01, 0.03)** |
| | **Death** | **<0.01 (<0.01, <0.01)** |
| **Stage 2** | | |
| NECT | Cured | 0.56 (0.49, 0.64) |
| | Cured with SAEs | 0.06 (0.04, 0.08) |
| | Rescue treatment | 0.03 (0.01, 0.04) |
| | Death | <0.01 (<0.01, <0.01) |
| Fexinidazole - inpatient | Cured | <0.01 (<0.01, 0.01) |
| | Cured with SAEs | <0.01 (<0.01, <0.01) |
| | Rescue treatment | <0.01 (<0.01, <0.01) |
| | Death | <0.01 (<0.01, <0.01) |
| Fexinidazole - outpatient | Cured | 0.33 (0.25, 0.41) |
| | Cured with SAEs | <0.01 (<0.01, <0.01) |
| | Rescue treatment | <0.01 (<0.01, 0.01) |
| | Death | <0.01 (<0.01, <0.01) |
| **All treatments** | **Cured** | **0.90 (0.88, 0.92)** |
| | **Cured with SAEs** | **0.07 (0.05, 0.09)** |
| | **Rescue treatment** | **0.03 (0.02, 0.05)** |
| | **Death** | **<0.01 (<0.01, <0.01)** |

**Supplementary Table 21**: Treatments and outcomes distributions for stage 1 and 2 patients, calculated according to the probability tree in 6. SAE: severe adverse events.

| Variable Name | Parameterization | Summary |
|---|---|---|
| Lumbar puncture and laboratory exam - cost | Gamma(2.42, 3.66) | 13.72 (3.32, 31.10) |
| Duration of hospital stay for NECT treatment in days | Fixed | 10 |
| Duration of hospital stay fexinidazole for stage 1 or 2 disease in days | Fixed | 10 |
| Duration of severe adverse events in days | Gamma (1.219, 2.377) | 2.91 (0.13, 9.84) |
| Probability of serious adverse events - pentamidine | Beta(1,499) | 0.0026 (0.0002, 0.0083) |
| Probability of serious adverse events - NECT | Beta(11.6,226.4) | 0.10 (0.07, 0.13) |
| Probability of serious adverse events - fexinidazole | Beta(3,261) | 0.01 (<0.01, 0.03) |
| Outpatient consultation - cost | Gamma(24.2, 0.13) | 3.14 (2.02, 4.48) |
| Hospital day - cost | Gamma(5.81, 0.59) | 3.42 (1.24, 6.74) |
| Course of pentamidine - cost | Fixed | 54 |
| Course of NECT - cost | Fixed | 360 |
| Course of fexinidazole - cost | Fixed | 50 |
| Delivery mark-up | Beta(45,55) | 0.45 (0.36, 0.55) |
| PNLTHA markup | Uniform(0.1, 0.2) | NA (NA, NA) |

**Supplementary Table 22**: Parameters for treatment costs

| | Pentamidine | NECT | Fexinidazole - inpatient | Fexinidazole - outpatient |
|---|---|---|---|---|
| Staging | 15.09 (3.65, 34.21) | 15.09 (3.65, 34.21) | 0 | 0 |
| Doctor's consult | 34.57 (22.23, 49.24) | 3.46 (2.22, 4.92) | 3.46 (2.22, 4.92) | 34.57 (22.23, 49.24) |
| Inpatient care | 0 | 37.64 (13.63, 74.18) | 37.64 (13.63, 74.18) | 0 |
| Medicine | 86.16 (80.51, 91.96) | 574.38 (536.75, 613.05) | 79.78 (74.55, 85.15) | 79.78 (74.55, 85.15) |
| Treatment for SAE | <0.01 (<0.01, 0.03) | 1 (0, 5) | 0 (0, 1) | 0 (0, 1) |
| **Total** | **136 (117, 160)** | **632 (583, 688)** | **110 (87, 144)** | **114 (101, 130)** |

**Supplementary Table 23**: Cost per person for different gHAT treatments. Because these are costs averaged over all patients and SAEs are rare, the average cost per patient for SAE is low.

##### A.10.5 Treatment and deaths in the pre-2026 period

To calculate the DALYs and deaths from 2000-25, which are featured in GUI as well as in one selection of our supplemental results, we made certain assumptions about the ubiquity of certain treatments from during that period. The following is a description of the data and assumptions we took.

###### Untreated

- Before 2013, we have no records of how many people were untreated, so we assume 1% of people were untreated. Starting in 2013, we know from the fourth stakeholders' meeting report that 91%, 97%, 98%, 100%, 99% of all cases in the DRC were treated between 2013-17 [46], then from the program, we know that 100%, 92%, 96%, 85%, 94% from 2018-2022. We will assume that 2023-5 had the same proportion untreated as 2022. We suspect that there was not 100% treatment adherence in any year, so for those years that indicate 100% treatment, we assume that 1% of cases remain untreated.

###### Stage 1

- From 2000-2012, there was no development for stage 1 drugs, so we put in that 99% of all cases were treated by Pentamidine, the standard of care since the 1940's [21].
- In 2013, the Fexinidazole trials started, so some proportion of S1 cases were then treated with Fexinidazole through trials [47]. Therefore, we assume 2% treatment with Fexinidazole in those trials in the period of 2013-16. From Nov 2016-Aug 2019, 7 clinics ran a prospective cohort with acoziborole treatment which was published along with data from 1 clinic in Guinea. We do not know how many of the 174 patients in that study were from DRC vs Guinea, but we know that fewer than 174 of the 4141 cases detected in DRC in those years cumulatively according to the HAT Atlas, which would equate to at most 4% of cases. If we only count the 1/6 of the cases in 2016, since the trial only ran for two months that year, and 8/12 cases in 2019, since the trial only ran for 8 months that year, we would consider a cumulative 2463 cases, so 174 cases would have been at most 7% of cases, which will be the proportion we will assume. Then we assume Fexinidazole again starting in 2020 when it was adopted as standard treatment. We will discuss below the process to parse the Fexinidazole treatments (which are unclassified between stage 1 and stage 2).
- See below for counts of S1 treated by Acoziborole during the trials.
- See below for post-2020 estimates.

###### Stage 2

- From 1948 until 2003, only Melarsoprol was available for the treatment of stage 2. In the 1990's a short course was developed, so for 2000-02, 99% of cases were treated this way. Rescue treatment consisted of a long course of Melarsoprol [21].
- In 2003-2006, Eflornithine monotherapy was approved, but it was rarely used due to needs in training and complicated protocols. Therefore, by 2006, only 20% of cases (anywhere) were treated by Eflornithine monotherapy, so I will assume that the prevalence was 5%, 10%, 15%, and 20% in 2003-06. The rest would be treated with Melarsoprol short-course protocol [21].
- In 2006, the WHO prepared a kit to administer Eflornithine monotherapy more easily, so its use increased to 64% by 2009 [21]. The rest would be assumed to be treated by Melarsoprol. Therefore, from 2007-2009, we assumed that the percent of cases treated by Eflornithine monotherapy would be 33%, 49%, 64% (essentially a linear interpolation from 20-64%). The rest would be treated by a short course of Melarsoprol.
- From 2003-2006 there were about 240 patients enrolled in NECT trials in the DRC [48, 49]. We are going to ignore those proportions because they constitute less than half of a percent of all cases in that period.
- In 2009, Nifurtimox-Eflornithine Combination Therapy (NECT) was endorsed for use by the WHO, and they developed a kit for delivery. NECT then became the dominant treatment for stage 2 starting in 2010, such that 88% of cases in that year were treated by NECT and the remaining 12% by Melarsoprol [21]. We will assume 88% of cases treated with NECT, 11% with Melarsoprol, and 1% remained untreated.

- Fexinidazole trials did not begin until 2013, so from 2010-2012 we assumed the use of NECT increased even further, to 94% and then 99% in 2011 and 2012. The rest would be treated with Melarsoprol.
- In 2013, Fexinidazole trials began, so we apportion 2% of cases to be treated with that drug for 2013-16, and 7% for 2016-2019 (see stage 1 for reasoning) until Fexinidazole is rolled out in 2020 [47, 50]. In the meantime, S2 cases are assumed to be treated with NECT.
- We will discuss below the process to parse the Fexinidazole treatments (which are unclassified between stage 1 and stage 2).
- See below for counts of S1 treated by Acoziborole during the trials.
- See below for post-2020 estimates.

**Acoziborole trial in 2016-2019**

- Between October 2016 and March 2019, there were 174 patients enrolled in DRC in the Acoziborole trial [51]. That would be 10 trimesters. Therefore, we assumed that 1/10 were enrolled in 2016, 1/10 in 2019, and 4/10 in 2017 and 2018. So 17 cases were treated in 2016 with Acoziborole, 70 cases in 2018, 70 cases in 2018, and 17 cases in 2019. Most of these were assumed to be in S2 of the disease, as only 20% of the cases enrolled were in "early or intermediate-stage of disease", which was defined as an absence of trypanosome and white blood cells in the CSF; in other words, the definition of stage 1.
  - Therefore, we assumed that in 2016-2019, 3, 12, 12, and 3 S1 cases were treated with Acoziborole, and 14, 58, 58, and 14 S2 cases were treated with Acoziborole.
  - Therefore, we assume about 2% of all S2 cases in 2016 and 2019 and 8% of all S2 cases in 2017 and 2018 were treated by Acoziborole, but we ignore the S1 cases treated by Acoziborole because these are too few.

**Calculating treatments as a proportion of S1 and S2 cases after 2020**

- In 2020, Fexinidazole was rolled out as a standard treatment. Because Fexinidazole is prescribed for both stages, much of the data we now have about cases is not stratified by disease stage.
- We know there were 660, 613, 395, 424, and 516 cases in 2020-2022, and when one breaks those cases down by stages, we get:

| Year | Cases, Stage UK | Cases, S1 | Cases, S2 | Total Cases |
|---|---|---|---|---|
| 2018 | 21 | 305 | 334 | 660 |
| 2019 | 24 | 289 | 296 | 609 |
| 2020 | 65 | 127 | 203 | 395 |
| 2021 | 151 | 81 | 190 | 424 |
| 2022 | 287 | 75 | 154 | 516 |

Cases in DRC during the period of 2018-2022. Abbreviations: UK: unknown stage, S1: stage 1 disease, S2: stage 2 disease.

- For 2018 and 2019, the proportion of cases that are not staged is marginal, and about 46-47% of cases are in S1.
- We have data that 123, 84, and 34 cases from 2020-2022 were treated with Pentamidine (PNLTHA program information). If we consider that 46% of all cases were in S1, then these Pentamidine treatment account for 68%, 43% and 14% of all S1 cases in 2020-2022.
- We have data that 161, 142, and 132 cases from 2020-2022 were treated with NECT (PNLTHA program information). If we consider that 54% of all cases were in S2, then these NECT treatments account for 75%, 62% and 47% of all S2 cases in 2020-2022.

- The remaining cases were treated with Fexinidazole, after accounting for the untreated. In S1, that would be 28%, 32%, and 80% for 2020-2022. In S2, that would be 21%, 23%, and 47% for 2020-2022. We assume a larger proportion of S1 cases are treated with Fexinidazole than S2 cases because Fexinidazole is contraindicated for late/severe disease.
- We assume that 2023-25 proportions are equal to 2022 proportions because we do not yet have any data on treatments in those years.

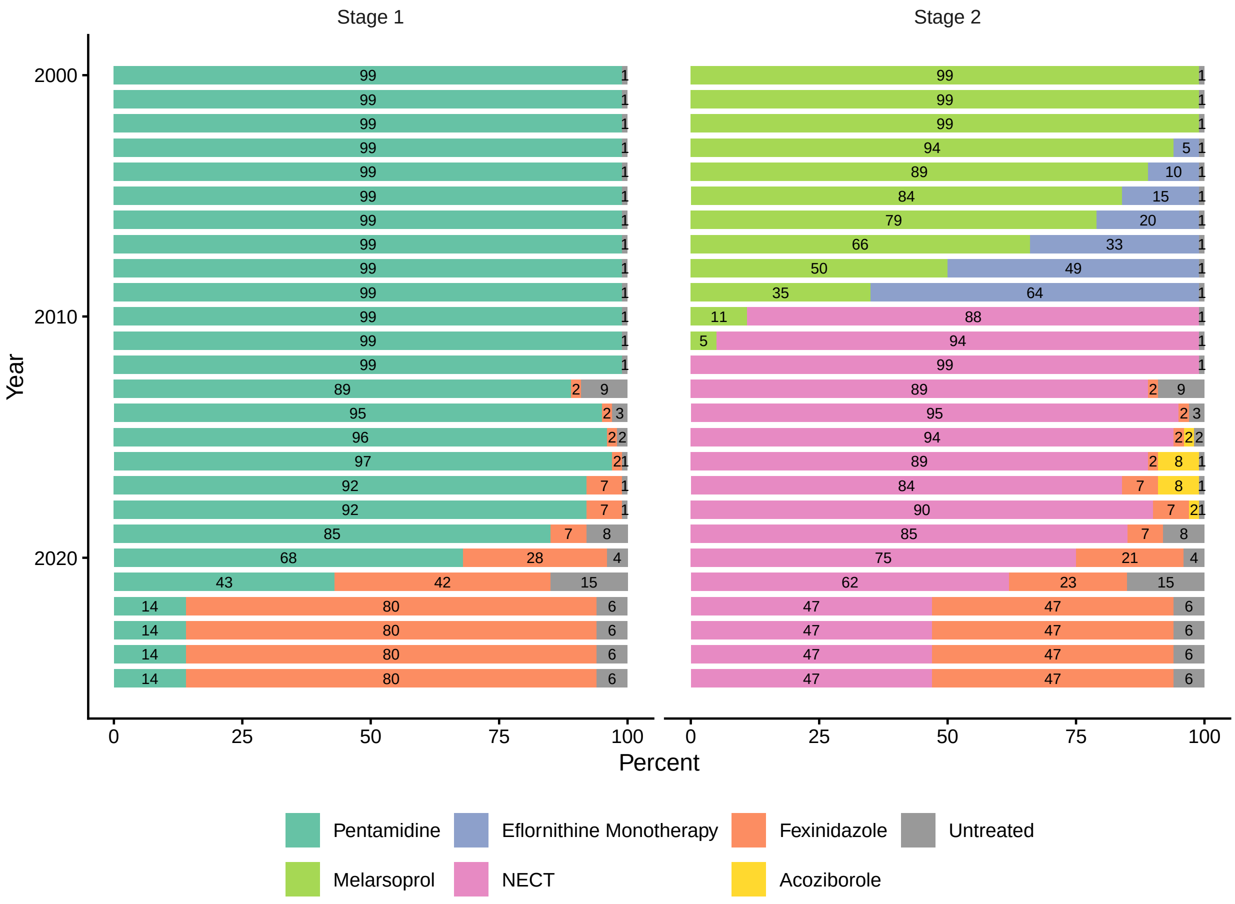

Treatments 2000-25. Since we have no data yet for what happened in 2023-25, we have assumed the same values as 2022. Abbreviations: NECT: nifurtimox-eflornithine combination therapy.

| Parameter | Distribution | Summary | Sources | Supplementary Note 3 Section |
|---|---|---|---|---|
| Treatment failure for Pentamidine, S1 | Beta(50.3, 665.48) | 0.07 (0.05-0.09) | [52–57] | E.6.7 |
| SAEs for Pentamidine, S1 | Beta(1.43, 551.42) | 0.0026 (0.0002-0.0081) | [53, 56, 57] | E.6.10 |
| Treatment failure for Melarsoprol, S2 | Beta(3, 247) | 0.01 (0.003-0.03) | [58] | |
| SAEs for Melarsoprol, S2 | Beta(47, 203) | 0.19 (0.14-0.24) | [58] | |
| CFR for Melarsoprol | Beta(12, 408) | 0.03 (0.02-0.05) | [58, 59] | |
| Treatment failure for Eflornithine, S2 | Beta(14, 149) | 0.09 (0.05-0.13) | [59] | |
| SAEs for Eflornithine, S2 | Beta(41, 102) | 0.29 (0.22-0.36) | [49] | |
| CFR for Eflornithine | Beta(9, 351) | 0.02 (0.01-0.04) | [59] | |
| Treatment failure for NECT, S2 | Beta(15.87, 378.55) | 0.04 (0.02-0.06) | [47–49, 60–62] | E.6.8 |
| SAEs for NECT, S2 | Beta(40.88, 367.8) | 0.10 (0.07-0.13) | [47–49, 60–62] | E.6.11 |
| CFR for NECT | 0 | 0 | [47–49, 60–62] | |
| Treatment failure for Fexinidazole, S1 & S2-early | Beta(9.49, 497) | 0.02 (<0.01–0.03) | [38, 47] | E.6.9 |
| SAEs for Fexinidazole, S1 & S2-early | Beta(3.00, 261) | 0.01 (<0.01–0.03]) | [38, 47] | E.6.12 |
| CFR for Fexinidazole | 0 | 0 | [38, 47] | |
| Treatment failure for Acoziborole, S1 & S2 | Beta(8, 167) | 0.05 (0.02-0.08) | [51] | |
| SAEs for Acoziborole, S1 & S2 | Beta(1, 208) | 0.005 (0.0001-0.02) | [51] | |
| CFR for Acoziborole, S1 & S2 | 0 | 0 | [51] | |
| Sensitivity of RDT | Beta(230, 1) | 1.00 (0.98-1.00) | [63] | E.5.7 |
| Age of death from infection (for years of life lost per fatal case) | Gamma(148, 0.18) | 26.65 (22.56, 31.07) | [47–49, 52–57, 60–62, 64–67] | E.7.1 |
| Average years lost at age of death | Interpolation of life expectancy at ages 20–35 | 45.4 [41.4–49.9] | [68] | E.7.2 |
| DALY weight, S1 | Beta(22.96, 147.21) | 0.14 (0.09, 0.19) | [69] | E.7.3 |
| DALY weight, S2 | Beta(18.37, 15.63) | 0.54 (0.37, 0.70) | [69] | E.7.4 |
| DALY weight, SAE | Uniform(0.04, 0.11) | 0.08 (0.04, 0.11) | [69] | E.7.5 |
| Duration S1 treatment | Fixed | 7 | [38] | E.6.4 |
| Duration S2 treatment | Fixed | 10 | [38] | E.6.5 & E.6.5 |
| Duration SAE | Gamma(1.22, 2.38) | 2.88 (0.14, 9.65) | [67] | E.6.13 |

Parameters for calculation of treatments, DALYs, and deaths 2000-23. Parameters are discussed in more detail in Supplemental Note 3. Parameters that are not features in Supplemental Note 3 are not part of the main analysis (e.g. drugs that are no longer used in 2024 such as melarsoprol and eflornithine, or acoziborole which has only been used as part of a trial). Abbreviations: S1: stage 1 disease, S2: stage 2 disease, SAE: severe adverse events, CFR: case fatality rate, NECT: nifurtimox-eflornithine combination therapy, DALY: disability-adjusted life-year.

Summary of treatment outcomes for historical treatments. FP = treatments and outcomes given to false positives.

| Drug | Outcome | Summary |
|---|---:|---:|
| Pentamidine | Convalescence & SAE | 0.002 (<0.001, 0.008) |
| | Convalescence & no SAE | 0.927 (0.907, 0.945) |
| | Detected relapse | 0.070 (0.052, 0.090) |
| | Undetected relapse | <0.001 (<0.001, 0.001) |
| | Treatment-related death | 0 |
| Pentamidine, FP | Convalescence & SAE | 0.003 (<0.001, 0.008) |
| | Convalescence & no SAE | 0.997 (0.992, 1.000) |
| | Detected relapse | 0 |
| | Undetected relapse | 0 |
| | Treatment-related death | 0 |
| Melarsoprol | Convalescence & SAE | 0.180 (0.136, 0.229) |
| | Convalescence & no SAE | 0.779 (0.727, 0.826) |
| | Detected relapse | 0.012 (0.002, 0.028) |
| | Undetected relapse | <0.001 (<0.001, <0.001) |
| | Treatment-related death | 0.029 (0.015, 0.047) |
| Melarsoprol, FP | Convalescence & SAE | 0.182 (0.138, 0.232) |
| | Convalescence & no SAE | 0.789 (0.738, 0.836) |
| | Detected relapse | 0 |
| | Undetected relapse | 0 |
| | Treatment-related death | 0.029 (0.015, 0.047) |
| Eflornithine | Convalescence & SAE | 0.255 (0.191, 0.325) |
| | Convalescence & no SAE | 0.634 (0.560, 0.706) |
| | Detected relapse | 0.086 (0.047, 0.133) |
| | Undetected relapse | <0.001 (<0.001, 0.001) |
| | Treatment-related death | 0.025 (0.011, 0.043) |
| Eflornithine, FP | Convalescence & SAE | 0.280 (0.211, 0.354) |
| | Convalescence & no SAE | 0.695 (0.620, 0.765) |
| | Detected relapse | 0 |
| | Undetected relapse | 0 |
| | Treatment-related death | 0.025 (0.011, 0.043) |
| NECT | Convalescence & SAE | 0.096 (0.070, 0.127) |
| | Convalescence & no SAE | 0.864 (0.828, 0.895) |
| | Detected relapse | 0.040 (0.023, 0.062) |
| | Undetected relapse | <0.001 (<0.001, <0.001) |
| | Treatment-related death | 0 |
| NECT, FP | Convalescence & SAE | 0.100 (0.073, 0.132) |
| | Convalescence & no SAE | 0.900 (0.868, 0.927) |
| | Detected relapse | 0 |
| | Undetected relapse | 0 |
| | Treatment-related death | 0 |
| Fexinidazole | Convalescence & SAE | 0.011 (0.002, 0.027) |
| | Convalescence & no SAE | 0.970 (0.950, 0.984) |
| | Detected relapse | 0.019 (0.009, 0.032) |
| | Undetected relapse | <0.001 (<0.001, <0.001) |
| | Treatment-related death | 0 |
| Fexinidazole, FP | Convalescence & SAE | 0.011 (0.002, 0.027) |
| | Convalescence & no SAE | 0.989 (0.973, 0.998) |
| | Detected relapse | 0 |
| | Undetected relapse | 0 |
| | Treatment-related death | 0 |
| | Convalescence & SAE | 0.005 (<0.001, 0.017) |

Summary of treatment outcomes for historical treatments. FP = treatments and outcomes given to false positives. *(continued)*

| Drug | Outcome | Summary |
|---|---:|---:|
| Acoziborole | Convalescence & no SAE | 0.947 (0.913, 0.973) |
| | Detected relapse | 0.048 (0.023, 0.081) |
| | Undetected relapse | <0.001 (<0.001, <0.001) |
| | Treatment-related death | 0 |
| Acoziborole, FP | Convalescence & SAE | 0.005 (<0.001, 0.018) |
| | Convalescence & no SAE | 0.995 (0.982, 1.000) |
| | Detected relapse | 0 |
| | Undetected relapse | 0 |
| | Treatment-related death | 0 |
| Untreated | Convalescence & SAE | 0 |
| | Convalescence & no SAE | 0 |
| | Detected relapse | 0 |
| | Undetected relapse | 0 |
| | Treatment-related death | 1.000 (1.000, 1.000) |
| Melarsoprol, second-line | Convalescence & SAE | 0.180 (0.136, 0.229) |
| | Convalescence & no SAE | 0.779 (0.727, 0.826) |
| | Detected relapse | 0 |
| | Undetected relapse | 0 |
| | Treatment-related death | 0.041 (0.022, 0.064) |
| NECT, second-line | Convalescence & SAE | 0.096 (0.070, 0.127) |
| | Convalescence & no SAE | 0.864 (0.828, 0.895) |
| | Detected relapse | 0 |
| | Undetected relapse | 0 |
| | Treatment-related death | 0.040 (0.023, 0.062) |

#### A.11 Sensitivity analysis

↩ Return to the Supplement Table of Contents

Our GUI (https://hatmepp.warwick.ac.uk/DRCCEA/v8/) provides health impact, cost calculations, and cost-effectiveness results for alternative horizons of 2026–2030 and 2026–2050 with and without 3% discounting so the impact of these choices can be seen. By exploring the "Optimal Strategy" map it is noted that changing the time horizon or discounting only results in minor changes to a few health zones if the objective is elimination of transmission by 2030. Longer time horizons and lack of discounting do change recommended strategies to more intensive interventions in more health zones (such as adding vector control or increasing screening coverage) when we consider the minimum cost objective or objectives based on different WTP thresholds.

### B Supplementary Results

#### B.1 Modeling results for sample health zone: Kikongo

↩ Return to the Supplement Table of Contents

Here we briefly show the results for one health zone to give a sense of the outputs for the fitting step for each health zone. All outcomes, AS and PS cases as well as new infections decrease over time thanks for screening. Active and Passive detections are usually a subset of 'new infections' from previous years. There is no 'validation' of New Infections against the data because new infections are an unobserved process. The new infections that are never detected are assumed to result in death, which is further discussed in Section B.3. Similar results are available for all health zones in the graphical user interphase https://hatmepp.warwick.ac.uk/DRCCEA/v6/.

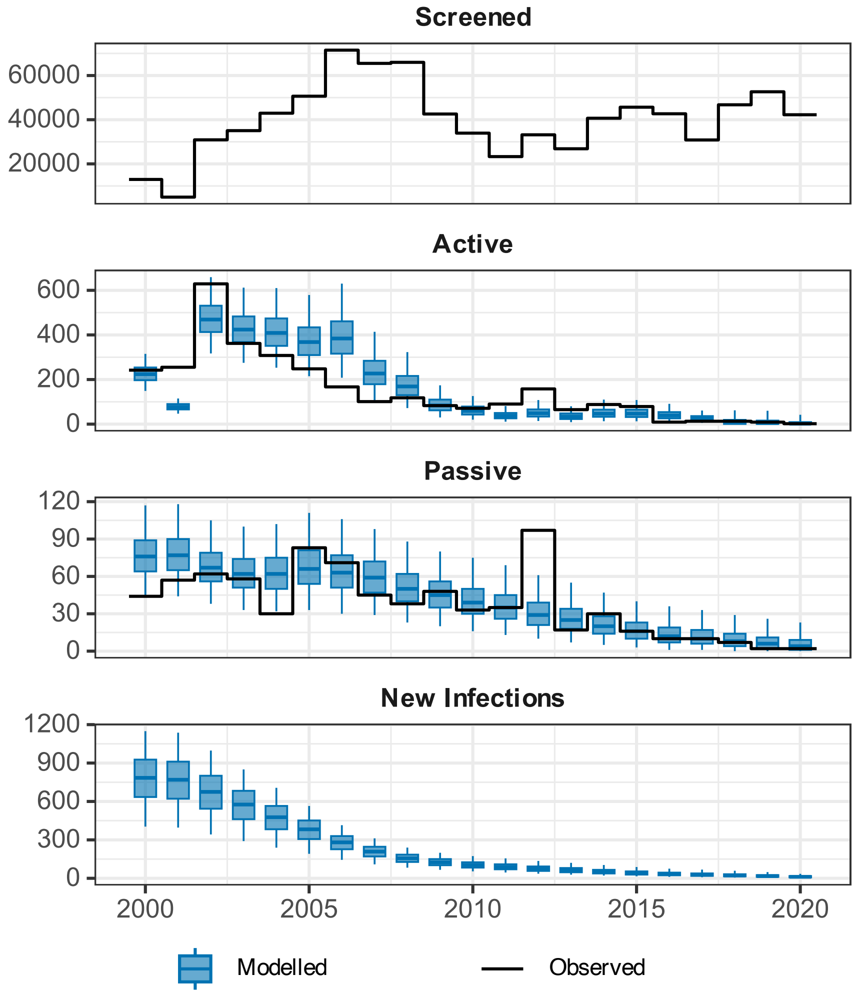

**Supplementary Figure 7**: **Fit to historical case data (2000–2020) from active and passive screening in the Kikongo health zone of the DRC.** Outputs include estimating unobservable new infections per year (bottom row). Blue box-and-whiskers present within-year summaries of model fits (median for centre line; and 50% and 95% credible intervals for the box and whiskers, respectively).

#### B.2 Model selection

↩ Return to the Supplement Table of Contents

We take this opportunity to update some of the results presented in our previous paper on the fitting of the model including animal transmission [2]. We do this in light of the availability of a considerable amount of new data (2000–2020 rather than the previous 2000–2016) and modifications made to the modelling process.

To obtain the statistical support for the two model variants in each health zone following fitting we categorised the Bayes factors (BF), where BF may be used to describe the statistical support for either model relative to the other, in a widely accepted way [70]:

| Support for Model | Bayes Factor |
|---:|---|
| Weak ('Barely worth mentioning') | $10^0 < \mathrm{BF} < 10^{\frac{1}{2}}$ |
| Substantial | $10^{\frac{1}{2}} < \mathrm{BF} < 10^1$ |
| Strong | $10^1 < \mathrm{BF} < 10^{\frac{3}{2}}$ |
| Very Strong | $10^{\frac{3}{2}} < \mathrm{BF} < 10^2$ |
| Decisive | $\mathrm{BF} > 10^2$ |

If BF for the model with animal transmission was greater than 1, the categorisation indicating statistical support for the model with animal transmission was used, otherwise, the categorisation used was for statistical support for the model without animal transmission [2]. Figure 8 shows the results from the current analyses and can be compared to Figure 3 of the previous article [2]. There was less support for the model with animal transmission than there was in our previous analyses [2]. In the previous article, 24 health zones were found to have 'Substantial' to 'Decisive' support for the model with animal transmission, whereas as a result of additional data and modifications to the modelling, there is now only a single health zone in this category.

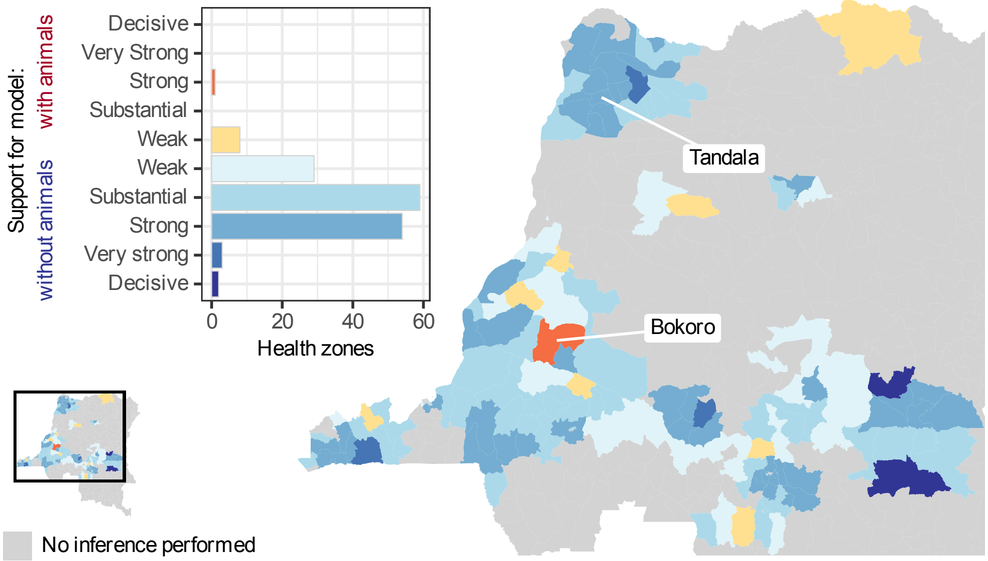

**Supplementary Figure 8**: Support for the model either with or without animals contributing to the transmission of gHAT. Levels of support taken from whichever of the two Bayes Factors (BF, with either evidence for the model with or without animal transmission as the denominator) exceeded 1. Weak, $10^0 < \mathrm{BF} < 10^{\frac{1}{2}}$; Substantial, $10^{\frac{1}{2}} < \mathrm{BF} < 10^1$; Strong, $10^1 < \mathrm{BF} < 10^{\frac{3}{2}}$; Very Strong, $10^{\frac{3}{2}} < \mathrm{BF} < 10^2$; and Decisive, $\mathrm{BF} > 10^2$. Health zones used as examples in Crump et al (2022), Bokoro and Tandala, are indicated. Shapefiles used to produce this map were provided by Nicole Hoff and Cyrus Sinai under a CC-BY licence (current versions can be found at https://data.humdata.org/dataset/drc-health-data).

Figure 9 is a bivariate choropleth map which combines support for models with and without animal transmission with differences in the probability of achieving the end of transmission by 2030 under the strategy where active screening in the health zone continues at the mean level observed in the period 2012–2016 ($P_d = \mathbb{P}(\text{EOT by 2030}|\text{Model without animal transmission}) - \mathbb{P}(\text{EOT by 2030}|\text{Model with animal transmission})$. This is an update of Figure B in the Supplementary Information of the previous animal transmission modelling paper [2]. For presentation in Figure 9 both variables were put into three categories:

| Choice of Model | Bayes Factor Values |
|---|---|
| without animals | $\mathrm{BF_{wo}} > 10^{\frac{1}{2}}$ |
| | or $\mathrm{BF_{wo}} < 10^{\frac{1}{2}} \wedge \mathrm{BF_{w}} < 10^{\frac{1}{2}}$ |
| with animals | $\mathrm{BF_{w}} > 10^{\frac{1}{2}}$ |

for model support, where $\mathrm{BF_{wo}}$ is the Bayes Factor value for support for the model without animals, and $\mathrm{BF_{w}}$ is the Bayes Factor value for support for the model with animals. For the difference in probability of achieving EoT by 2030 ($P_d$):

| Category | $\Delta$ Pr. EoT by 2030 |
|---|---|
| low | $P_d \leq 0.05$ |
| medium | $0.05 < P_d \leq 0.1$ |
| high | $P_d > 0.1$ |

In Figure 9 there are 31 health zones (dark purple) with (i) more than 10% reduction in the probability of meeting the EoT goal under the model with animal transmission compared to the model without animal transmission and (ii) with weak support for either model variant. In the previous article, this category contained 18 health zones. This discrepancy likely reflects both the additional data included and a change to the priors associated with the animal transmission model to remove the nesting of the models. In these 31 health zones, there is considerable uncertainty in whether animals contribute to transmission and this could alter policy recommendations for future strategy based on model predictions. The single health zone with strong support for the model with animal transmission also has a $> 10\%$ reduction in the probability of reaching the 2030 EoT goal.

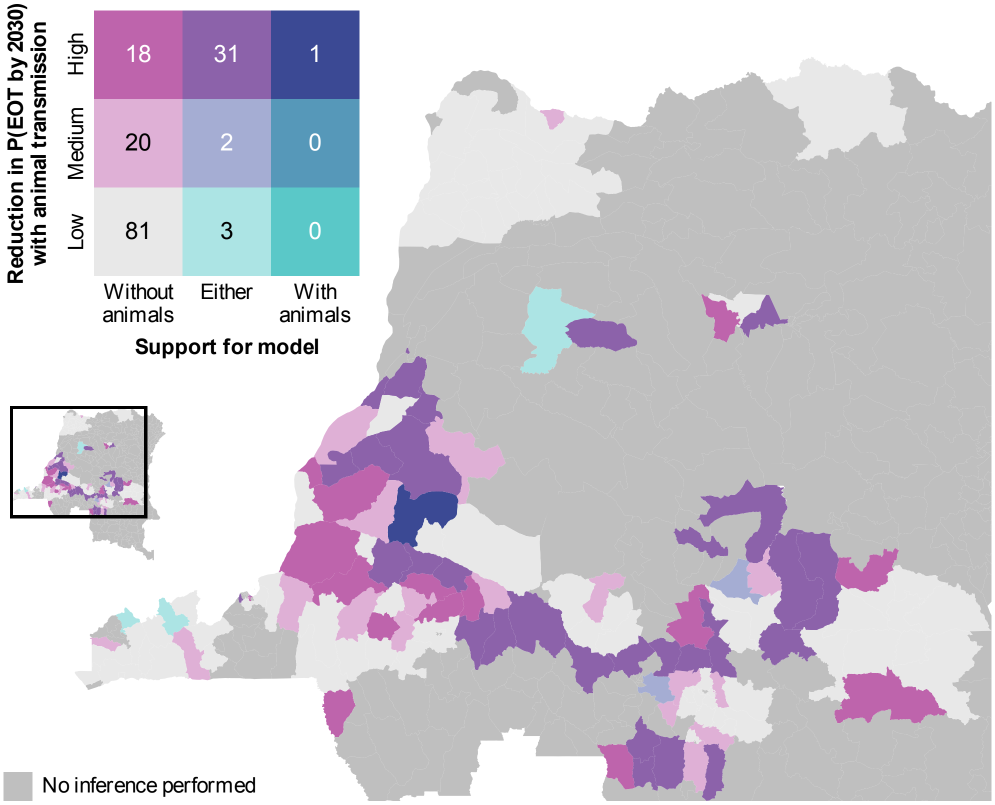

**Supplementary Figure 9**: Bivariate choropleth showing support for the models with or without animals contributing to transmission and the difference in the probability $(P_d)$ of achieving EoT to humans by 2030 from these two models ("High" is more than 10% difference, "Medium" is 5-10% difference, and "Low" is less than 5% difference). Shapefiles used to produce this map were provided by Nicole Hoff and Cyrus Sinai under a CC-BY licence (current versions can be found at https://data.humdata.org/dataset/drc-health-data).

#### B.3 Detected cases and undetected deaths

↩ Return to the Supplement Table of Contents

As there is no data available to inform this projection in the DRC, we have to infer the unreported deaths. In the model, the number of unreported cases (assumed to all result in deaths) are triangulated by the number of active cases screened, the presence or absence of VC, the cases detected and the proportion of cases detected that are S1 vs S2 cases. For a more extended discussion of the subject of detected and undetected cases in the model, see the S1 Text for Antillon et. al (2023) [27], pages 11-14. An illustration of the cases (reported, unreported, and false-positives) as well as the proportion of cases reported is illustrated in Supplementary Figures 10.

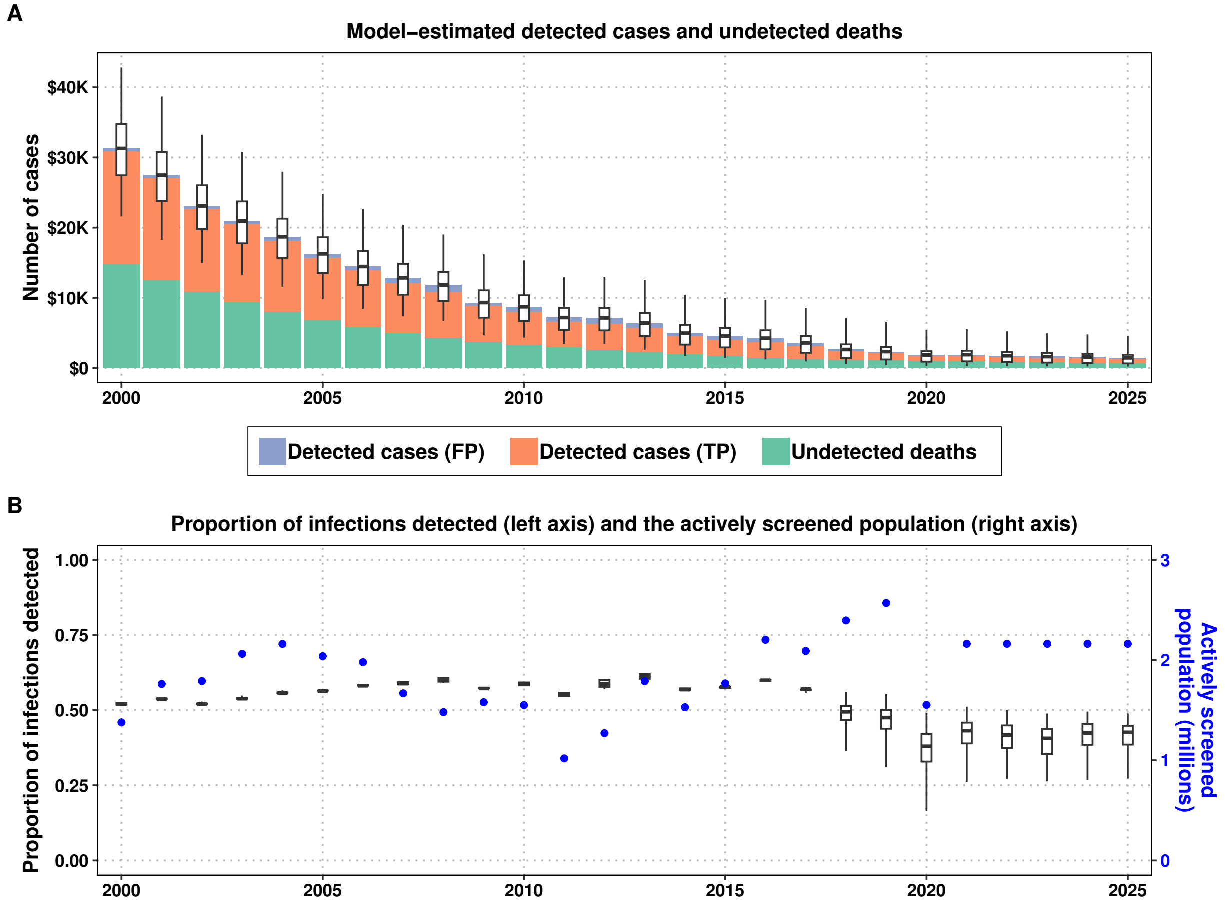

**Supplementary Figure 10**: Comparison of detected and undetected infections generated by the model and their relationship to active screening intensity. A) detected cases, undetected cases – which result in deaths – and false positive cases. Box-and-whisker plots show the mean estimate, interquartile range, and 95% prediction intervals of the total number of cases. B) the proportion of cases detected, and the population tested by mobile screening teams. Abbreviations: TP: true positives, FP: false positives.

#### B.4 Cost-effectiveness for example health zones: Kikongo and Kwamouth

↩ Return to the Supplement Table of Contents

| | Mean AS | Int. AS | Mean AS + Targeted VC | Int. AS + Targeted VC | Mean AS + Full VC | Int. AS + Full VC |
|---|---|---|---|---|---|---|
| **Health effects** | | | | | | |
| Reported cases | 23 (0, 103) | 22 (0, 109) | 14 (0, 61) | 15 (0, 76) | 12 (0, 53) | 12 (0, 67) |
| Deaths undetected | 25 (0, 111) | 18 (0, 76) | 15 (0, 59) | 12 (0, 48) | 12 (0, 47) | 10 (0, 38) |
| Cases total | 47 (0, 197) | 40 (0, 168) | 29 (0, 106) | 27 (0, 107) | 24 (0, 86) | 22 (0, 91) |
| Deaths detected[a] | 0 (0, 0) | 0 (0, 0) | 0 (0, 0) | 0 (0, 0) | 0 (0, 0) | 0 (0, 0) |
| YLD | 29 (0, 113) | 21 (0, 84) | 18 (0, 69) | 15 (0, 56) | 15 (0, 58) | 12 (0, 48) |
| YLL | 1,101 (0, 4,988) | 792 (0, 3,424) | 677 (0, 2,616) | 534 (0, 2,112) | 550 (0, 2,114) | 448 (0, 1,698) |
| DALYs | 1,129 (0, 5,086) | 813 (0, 3,487) | 696 (0, 2,669) | 548 (0, 2,154) | 565 (0, 2,155) | 460 (0, 1,730) |
| ΔDALYs | 0 (0, 0) | 317 (-1,475, 3,113) | 434 (-1,039, 3,443) | 581 (-711, 3,754) | 565 (-708, 3,752) | 669 (-495, 3,987) |
| **Costs, in thousands US$ (not discounted)** | | | | | | |
| AS costs | 990 (204, 1926) | 2436 (1801, 3332) | 816 (203, 1629) | 2343 (1795, 3147) | 738 (201, 1479) | 2303 (1785, 3058) |
| PS costs | 681 (451, 958) | 668 (444, 945) | 658 (443, 934) | 646 (438, 917) | 643 (439, 908) | 633 (433, 897) |
| VC costs | 0 (0, 0) | 0 (0, 0) | 169 (26, 428) | 158 (26, 389) | 1182 (198, 2837) | 1123 (199, 2659) |
| Treatment | 12 (0, 55) | 12 (0, 57) | 8 (0, 33) | 8 (0, 40) | 6 (0, 29) | 7 (0, 36) |
| Costs total | 1683 (728, 2758) | 3116 (2336, 4130) | 1651 (756, 2735) | 3155 (2386, 4160) | 2569 (964, 4757) | 4066 (2634, 6037) |
| ΔCosts | 0 (0, 0) | 1,433 (449, 2,476) | -33 (-1,111, 1,049) | 1,471 (497, 2,486) | 886 (-599, 2,815) | 2,383 (1,032, 4,135) |
| **EoT** | | | | | | |
| Year of EoT | 2035 (2023, 2057) | 2033 (2023, 2054) | 2032 (2023, 2053) | 2031 (2023, 2050) | 2030 (2023, 2050) | 2030 (2023, 2047) |
| Prob EoT 2030 | 0.37 | 0.44 | 0.53 | 0.61 | 0.67 | 0.72 |
| Prob EoT 2040 | 0.80 | 0.87 | 0.91 | 0.93 | 0.94 | 0.95 |
| **Cost-effectiveness without uncertainty (discounted)[b]** | | | | | | |
| ΔDALYs | 0 | 148 | 199 | 273 | 262 | 315 |
| ΔCosts | 0 | 1,129,905 | 8,734 | 1,189,285 | 883,821 | 2,054,185 |
| ICER | Min Cost | Dominated | 44 | Weakly Dominated | 13,839 | 22,072 |
| **Cost-effectiveness with uncertainty (discounted), conditional on WTP[c].** | | | | | | |
| WTP: $0 | 0.55(p) | 0 | 0.41 | 0 | 0.03 | 0 |
| WTP: $250 | 0.51 | 0 | 0.45(p) | 0 | 0.04 | 0 |
| WTP: $500 | 0.48 | 0 | 0.47(p) | 0 | 0.04 | 0 |
| WTP: $750 | 0.46 | 0 | 0.49(p) | 0 | 0.05 | 0 |
| WTP: $1000 | 0.45 | 0.01 | 0.49(p) | 0.01 | 0.05 | 0 |
| WTP: $1500 | 0.42 | 0.01 | 0.49(p) | 0.02 | 0.06 | 0 |

[a] Detected deaths are those that occur due to treatment failure of loss-to-follow-up.

[b] Cost-effectiveness results are given for discounted DALYs and costs as per convention

[c] (p) is the preferred strategy; the strategy with the highest mean net monetary benefits

**Supplementary Table 25**: Summary of effects, costs, elimination of transmission (EoT) 2030, and cost-effectiveness with and without uncertainty in Kikongo health zone. Means are given along with 95% prediction intervals (PIs). YLL: years of life lost (to fatal disease), YLD: years of life lost to disability, DALYs: disability-adjusted life-years, PS: passive screening, AS: active screening, VC: vector control, ICER: incremental cost-effectiveness ratio, WTP: willingness to pay (USD per DALY averted), EoT: elimination of transmission.

| | Mean AS | Int. AS | Mean AS + Targeted VC | Int. AS + Targeted VC | Mean AS + Full VC | Int. AS + Full VC |
|---|---|---|---|---|---|---|
| **Health effects** | | | | | | |
| Reported cases | 10 (0, 76) | 10 (0, 78) | 4 (0, 27) | 4 (0, 29) | 4 (0, 27) | 4 (0, 28) |
| Deaths undetected | 3 (0, 24) | 3 (0, 21) | 1 (0, 9) | 1 (0, 8) | 1 (0, 9) | 1 (0, 8) |
| Cases total | 14 (0, 97) | 13 (0, 95) | 5 (0, 32) | 5 (0, 34) | 5 (0, 32) | 5 (0, 33) |
| Deaths detected[a] | 0 (0, 0) | 0 (0, 0) | 0 (0, 0) | 0 (0, 0) | 0 (0, 0) | 0 (0, 0) |
| YLD | 5 (0, 34) | 4 (0, 29) | 2 (0, 9) | 2 (0, 9) | 2 (0, 9) | 2 (0, 8) |
| YLL | 146 (0, 1,086) | 125 (0, 930) | 59 (0, 387) | 53 (0, 358) | 58 (0, 382) | 53 (0, 365) |
| DALYs | 151 (0, 1,111) | 129 (0, 960) | 61 (0, 394) | 55 (0, 364) | 60 (0, 388) | 55 (0, 372) |
| ΔDALYs | 0 (0, 0) | 21 (-627, 780) | 90 (-187, 928) | 96 (-177, 942) | 91 (-185, 941) | 96 (-176, 938) |
| **Costs, in thousands US$ (not discounted)** | | | | | | |
| AS costs | 862 (273, 2438) | 3030 (2410, 4038) | 678 (273, 1496) | 2959 (2405, 3822) | 676 (273, 1468) | 2958 (2405, 3818) |
| PS costs | 700 (468, 1059) | 694 (468, 1049) | 664 (463, 952) | 661 (464, 941) | 663 (464, 947) | 660 (463, 939) |
| VC costs | 0 (0, 0) | 0 (0, 0) | 522 (117, 1410) | 509 (116, 1357) | 1037 (231, 2769) | 1010 (232, 2681) |
| Treatment | 5 (0, 40) | 5 (0, 41) | 2 (0, 15) | 2 (0, 15) | 2 (0, 14) | 2 (0, 15) |
| Costs total | 1567 (799, 3415) | 3729 (3001, 4930) | 1866 (960, 3549) | 4131 (3189, 5593) | 2378 (1101, 4808) | 4630 (3346, 6743) |
| ΔCosts | 0 (0, 0) | 2,162 (599, 3,181) | 299 (-1,146, 1,621) | 2,564 (1,039, 3,723) | 811 (-687, 2,802) | 3,063 (1,536, 4,771) |
| **EoT** | | | | | | |
| Year of EoT | 2027 (2021, 2040) | 2027 (2021, 2039) | 2026 (2021, 2032) | 2026 (2021, 2031) | 2026 (2021, 2031) | 2026 (2021, 2031) |
| Prob EoT 2030 | 0.81 | 0.83 | 0.96 | 0.97 | 0.97 | 0.97 |
| Prob EoT 2040 | 0.98 | 0.98 | >0.99 | >0.99 | >0.99 | >0.99 |
| **Cost-effectiveness without uncertainty (discounted)[b]** | | | | | | |
| ΔDALYs | 0 | 11 | 44 | 47 | 44 | 47 |
| ΔCosts | 0 | 1,677,738 | 328,875 | 2,083,904 | 818,036 | 2,561,643 |
| ICER | Min Cost | Dominated | 7,504 | 514,693 | Weakly Dominated | Dominated |
| **Cost-effectiveness with uncertainty (discounted), conditional on WTP[c].** | | | | | | |
| WTP: $0 | 0.83(p) | 0 | 0.14 | 0 | 0.03 | 0 |
| WTP: $250 | 0.82(p) | 0 | 0.14 | 0 | 0.03 | 0 |
| WTP: $500 | 0.82(p) | 0 | 0.15 | 0 | 0.03 | 0 |
| WTP: $750 | 0.81(p) | 0 | 0.15 | 0 | 0.03 | 0 |
| WTP: $1000 | 0.81(p) | 0 | 0.16 | 0 | 0.03 | 0 |
| WTP: $1500 | 0.8(p) | 0 | 0.16 | 0 | 0.03 | 0 |

[a] Detected deaths are those that occur due to treatment failure of loss-to-follow-up.
[b] Cost-effectiveness results are given for discounted DALYs and costs as per convention
[c] (p) is the preferred strategy; the strategy with the highest mean net monetary benefits

**Supplementary Table 26**: Summary of effects, costs, elimination of transmission (EoT) 2030, and cost-effectiveness with and without uncertainty in Kwamouth health zone. Means are given along with 95% prediction intervals (PIs). YLL: years of life lost (to fatal disease), YLD: years of life lost to disability, DALYs: disability-adjusted life-years, PS: passive screening, AS: active screening, VC: vector control, ICER: incremental cost-effectiveness ratio, WTP: willingness to pay (USD per DALY averted), EoT: elimination of transmission.

#### B.5 Cost-effectiveness in Bas Uélé region: Ango and Doruma health zones

↩ Return to the Supplement Table of Contents

We also show detailed results for the two special health zones, Ango (Supplementary Table 27) and Doruma (Supplementary Table 28).

Because there are no activities simulated in these health zones, the status quo would detect zero cases and incur no costs. In Ango and Doruma, we estimate 939 [95% PI: 0–3098] and 980 [95% PI: 2–3249] deaths, respectively, leading to 42,516 [95% PI: 0–140,203] and 44,368 [95% PI: 89–148,286] DALYs, respectively. The lack of activities not only yields a high expected number of undetected cases, which are almost all resulting in deaths, but also a large degree of uncertainty.

If only one intervention were to be instituted, in terms of detecting cases *Restart PS* would be cost-effective for Ango and Doruma, in both health zones dominating the re-start of AS, therefore allowing more cases to be reported under PS at a lower cost. However, combinations of interventions are best, and the addition of *Int. AS* to any combination would substantially lower the number of total cases. Unfortunately, the probability of EoT by 2030 for both health zones is exceedingly low: 5%–13% in Ango and 5%–16% in Doruma, depending on the strategy. By 2040, there is an 8% and 10% chance of reaching EoT if nothing is done, but there is up to a 50% and 79% change of reaching EoT with the most ambitious strategy (*Int. AS* + *Full VC* + *Restart PS*).

In terms of cost-effectiveness in Ango and Doruma, no strategy would minimize costs, since nothing is assumed to be done and therefore nothing is being spent. However, the model indicates that *Restart PS* has a low ICER of $20/DALY averted in Ango and $7/DALY averted in Doruma. In Ango, *Int. AS* + *Restart PS*, would have an ICER of 356 and *Targeted VC* + *Restart PS* would have an ICER of $457/DALY, indicating cost-effectiveness at a moderately low WTP. In Ango, the only other efficient strategy would constitute the addition of AS (*Int. AS* + *Targeted VC* + *Restart PS*) has an ICER of $1,082/DALY–still under the threshold equivalent to 3 GDPs per capita. In Doruma, the only other sensible strategy after *Restart PS* is *Int. AS* + *Restart PS*, which has an ICER of %809/DALY averted.

The strategies that would maximize EoT in Ango are dominated because Full VC seems to spend more money without an advantage in terms of DALYs averted. In Doruma, such strategies are either dominated because they lack Int. AS that treats current cases, or are decidedly not cost-effective with an ICER of $24,713/DALY averted (*Int. AS* + *Full VC* + *Restart PS*).

Accounting for uncertainty in Ango, *Restart PS* would be the optimal strategy at a WTP <$250, and *Targeted VC* + *Restart PS* would be optimal at higher threholds. In Doruma, *Restart PS* would be the optimal strategy at $250<WTP<$1000; and above that, *Int. AS* + *Restart PS* would be optimal.

| | No AS, PS, nor VC | Int. AS + no PS | Restart PS | Int. AS + Restart PS | Targeted VC + Restart PS | Int. AS + Targeted VC + Restart PS | Full VC + Restart PS | Int. AS + Full VC + Restart PS |
|---|---|---|---|---|---|---|---|---|
| **Health effects** | | | | | | | | |
| Reported cases | 0 (0, 0) | 175 (0, 563) | 214 (0, 594) | 270 (0, 754) | 57 (0, 152) | 84 (0, 241) | 57 (0, 154) | 84 (0, 240) |
| Deaths undetected | 939 (0, 3098) | 555 (0, 1679) | 484 (0, 1525) | 307 (0, 946) | 129 (0, 374) | 89 (0, 259) | 129 (0, 371) | 89 (0, 256) |
| Cases total | 939 (0, 3098) | 730 (0, 2178) | 698 (0, 2059) | 576 (0, 1658) | 186 (0, 503) | 173 (0, 473) | 186 (0, 505) | 173 (0, 473) |
| Deaths detected[a] | 0 (0, 0) | 0 (0, 0) | 0 (0, 1) | 0 (0, 1) | 0 (0, 0) | 0 (0, 0) | 0 (0, 0) | 0 (0, 0) |
| YLD | 556 (0, 1943) | 333 (0, 1081) | 320 (0, 1035) | 207 (0, 656) | 83 (0, 254) | 58 (0, 174) | 83 (0, 253) | 58 (0, 172) |
| YLL | 41,960 (0, 138,144) | 24,808 (0, 75,258) | 21,638 (0, 68,248) | 13,719 (0, 42,248) | 5,775 (0, 16,750) | 3,987 (0, 11,553) | 5,778 (0, 16,655) | 3,991 (0, 11,461) |
| DALYs | 42,516 (0, 140,203) | 25,141 (0, 76,399) | 21,958 (0, 69,203) | 13,926 (0, 42,746) | 5,858 (0, 17,001) | 4,046 (0, 11,715) | 5,861 (0, 16,903) | 4,049 (0, 11,619) |
| ΔDALYs | 0 (0, 0) | 17,375 (-9,983, 80,409) | 20,558 (-3,410, 86,446) | 28,590 (0, 106,753) | 36,658 (0, 124,071) | 38,470 (0, 128,964) | 36,655 (0, 124,191) | 38,467 (0, 128,948) |
| **Costs, in thousands US$ (not discounted)** | | | | | | | | |
| AS costs | 0 (0, 0) | 1437 (197, 2101) | 0 (0, 0) | 1537 (207, 2119) | 0 (0, 0) | 1230 (206, 1887) | 0 (0, 0) | 1231 (205, 1897) |
| PS costs | 0 (0, 0) | 0 (0, 0) | 89 (60, 125) | 89 (60, 125) | 89 (60, 125) | 89 (60, 125) | 89 (60, 125) | 89 (60, 125) |
| VC costs | 0 (0, 0) | 0 (0, 0) | 0 (0, 0) | 0 (0, 0) | 3331 (850, 6645) | 3259 (877, 6584) | 6454 (1655, 12946) | 6315 (1679, 12805) |
| Treatment | 0 (0, 0) | 86 (0, 280) | 125 (0, 352) | 145 (0, 403) | 33 (0, 90) | 45 (0, 127) | 33 (0, 91) | 45 (0, 127) |
| Costs total | 0 (0, 0) | 1523 (197, 2251) | 214 (80, 443) | 1771 (299, 2444) | 3454 (943, 6792) | 4624 (1278, 8369) | 6576 (1752, 13087) | 7681 (2110, 14593) |
| ΔCosts | 0 (0, 0) | 1,523 (197, 2,251) | 214 (80, 443) | 1,771 (299, 2,444) | 3,454 (943, 6,792) | 4,624 (1,278, 8,369) | 6,576 (1,752, 13,087) | 7,681 (2,110, 14,593) |
| **EoT** | | | | | | | | |
| Year of EoT | 2054 (2022, 2057) | 2053 (2022, 2057) | 2053 (2022, 2057) | 2051 (2022, 2057) | 2044 (2022, 2057) | 2042 (2022, 2057) | 2044 (2022, 2057) | 2042 (2022, 2057) |
| Prob EoT 2030 | 0.05 | 0.06 | 0.06 | 0.08 | 0.10 | 0.13 | 0.10 | 0.13 |
| Prob EoT 2040 | 0.08 | 0.12 | 0.13 | 0.19 | 0.44 | 0.50 | 0.44 | 0.50 |
| **Cost-effectiveness without uncertainty (discounted)[b]** | | | | | | | | |
| ΔDALYs | 0 | 7,385 | 8,667 | 12,190 | 15,553 | 16,462 | 15,551 | 16,461 |
| ΔCosts | 0 | 1,231,854 | 170,039 | 1,425,562 | 2,961,219 | 3,945,076 | 5,641,839 | 6,578,235 |
| ICER | Min Cost | Dominated | 20 | 356 | 457 | 1,082 | Dominated | Dominated |
| **Cost-effectiveness with uncertainty (discounted), conditional on WTP[c].** | | | | | | | | |
| WTP: $0 | 1(p) | 0 | 0 | 0 | 0 | 0 | 0 | [illegible] |
| WTP: $250 | 0.1 | 0.02 | 0.63(p) | 0.16 | 0.06 | 0.01 | 0 | [illegible] |
| WTP: $500 | 0.08 | 0.03 | 0.35 | 0.24(p) | 0.21 | 0.09 | 0 | [illegible] |
| WTP: $750 | 0.07 | 0.03 | 0.21 | 0.21 | 0.28(p) | 0.19 | 0 | [illegible] |
| WTP: $1000 | 0.06 | 0.03 | 0.15 | 0.18 | 0.29(p) | 0.28 | 0 | [illegible] |
| WTP: $1500 | 0.05 | 0.02 | 0.1 | 0.13 | 0.27 | 0.41(p) | 0.01 | 0.01 |

[a] Detected deaths are those that occur due to treatment failure of loss-to-follow-up.
[b] Cost-effectiveness results are given for discounted DALYs and costs as per convention
[c] (p) is the preferred strategy; the strategy with the highest mean net monetary benefits

**Supplementary Table 27**: Summary of effects, costs, elimination of transmission (EoT) 2030, and cost-effectiveness with and without uncertainty in Ango health zone. Means are given along with 95% prediction intervals (PIs). YLL: years of life lost (to fatal disease), YLD: years of life lost to disability, DALYs: disability-adjusted life-years, PS: passive screening, AS: active screening, VC: vector control, ICER: incremental cost-effectiveness ratio, WTP: willingness to pay (USD per DALY averted), EoT: elimination of transmission.

| | No AS, PS, nor VC | Int. AS + no PS | Restart PS | Int. AS + Restart PS | Full VC + Restart PS | Int. AS + Full VC + Restart PS |
|---|---|---|---|---|---|---|
| **Health effects** | | | | | | |
| Reported cases | 0 (0, 0) | 103 (0, 411) | 117 (0, 397) | 114 (0, 369) | 113 (0, 374) | 111 (0, 360) |
| Deaths undetected | 980 (2, 3249) | 630 (1, 2233) | 128 (0, 461) | 87 (0, 278) | 123 (0, 439) | 84 (0, 265) |
| Cases total | 980 (2, 3249) | 733 (1, 2539) | 245 (1, 832) | 201 (1, 634) | 236 (1, 802) | 195 (1, 610) |
| Deaths detected[a] | 0 (0, 0) | 0 (0, 0) | 0 (0, 0) | 0 (0, 0) | 0 (0, 0) | 0 (0, 0) |
| YLD | 572 (1, 2085) | 371 (0, 1403) | 83 (0, 309) | 57 (0, 194) | 80 (0, 298) | 55 (0, 187) |
| YLL | 43,796 (86, 146,530) | 28,156 (44, 99,475) | 5,703 (0, 20,565) | 3,895 (0, 12,423) | 5,491 (0, 19,694) | 3,777 (0, 11,933) |
| DALYs | 44,368 (89, 148,286) | 28,527 (44, 100,864) | 5,786 (1, 20,846) | 3,953 (0, 12,621) | 5,571 (1, 20,015) | 3,832 (0, 12,080) |
| ΔDALYs | 0 (0, 0) | 15,842 (-29,798, 94,177) | 38,582 (0, 131,662) | 40,416 (0, 138,235) | 38,797 (0, 132,572) | 40,536 (0, 138,415) |
| **Costs, in thousands US$ (not discounted)** | | | | | | |
| AS costs | 0 (0, 0) | 847 (179, 1353) | 0 (0, 0) | 850 (238, 1301) | 0 (0, 0) | 840 (226, 1297) |
| PS costs | 0 (0, 0) | 0 (0, 0) | 80 (52, 115) | 80 (52, 115) | 80 (52, 115) | 80 (52, 115) |
| VC costs | 0 (0, 0) | 0 (0, 0) | 0 (0, 0) | 0 (0, 0) | 1699 (409, 3262) | 1579 (415, 3107) |
| Treatment | 0 (0, 0) | 51 (0, 205) | 62 (0, 213) | 59 (0, 190) | 60 (0, 202) | 57 (0, 186) |
| Costs total | 0 (0, 0) | 898 (179, 1464) | 142 (68, 295) | 989 (319, 1511) | 1839 (489, 3442) | 2556 (729, 4414) |
| ΔCosts | 0 (0, 0) | 898 (179, 1,464) | 142 (68, 295) | 989 (319, 1,511) | 1,839 (489, 3,442) | 2,556 (729, 4,414) |
| **EoT** | | | | | | |
| Year of EoT | 2054 (2026, 2057) | 2052 (2026, 2057) | 2038 (2026, 2057) | 2036 (2025, 2051) | 2038 (2026, 2057) | 2036 (2025, 2051) |
| Prob EoT 2030 | 0.05 | 0.06 | 0.12 | 0.16 | 0.13 | 0.16 |
| Prob EoT 2040 | 0.10 | 0.15 | 0.66 | 0.77 | 0.67 | 0.79 |
| **Cost-effectiveness without uncertainty (discounted)[b]** | | | | | | |
| ΔDALYs | 0 | 6,619 | 16,140 | 17,010 | 16,236 | 17,064 |
| ΔCosts | 0 | 734,178 | 118,386 | 822,180 | 1,531,753 | 2,145,123 |
| ICER | Min Cost | Dominated | 7 | 809 | Dominated | 24,713 |
| **Cost-effectiveness with uncertainty (discounted), conditional on WTP[c].** | | | | | | |
| WTP: $0 | 1(p) | 0 | 0 | 0 | 0 | 0 |
| WTP: $250 | 0.06 | 0 | 0.83(p) | 0.1 | 0 | 0 |
| WTP: $500 | 0.05 | 0.01 | 0.69(p) | 0.24 | 0.01 | 0 |
| WTP: $750 | 0.04 | 0.01 | 0.58(p) | 0.32 | 0.02 | 0.02 |
| WTP: $1000 | 0.04 | 0.01 | 0.51 | 0.37(p) | 0.04 | 0.04 |
| WTP: $1500 | 0.04 | 0.01 | 0.42 | 0.4(p) | 0.06 | 0.08 |

[a] Detected deaths are those that occur due to treatment failure of loss-to-follow-up.
[b] Cost-effectiveness results are given for discounted DALYs and costs as per convention
[c] (p) is the preferred strategy; the strategy with the highest mean net monetary benefits

**Supplementary Table 28**: Summary of effects, costs, elimination of transmission (EoT) 2030, and cost-effectiveness with and without uncertainty in Doruma health zone. Means are given along with 95% prediction intervals (PIs). YLL: years of life lost (to fatal disease), YLD: years of life lost to disability, DALYs: disability-adjusted life-years, PS: passive screening, AS: active screening, VC: vector control, ICER: incremental cost-effectiveness ratio, WTP: willingness to pay (USD per DALY averted), EoT: elimination of transmission.

#### B.6 Results by coordination

↩ Return to the Supplement Table of Contents

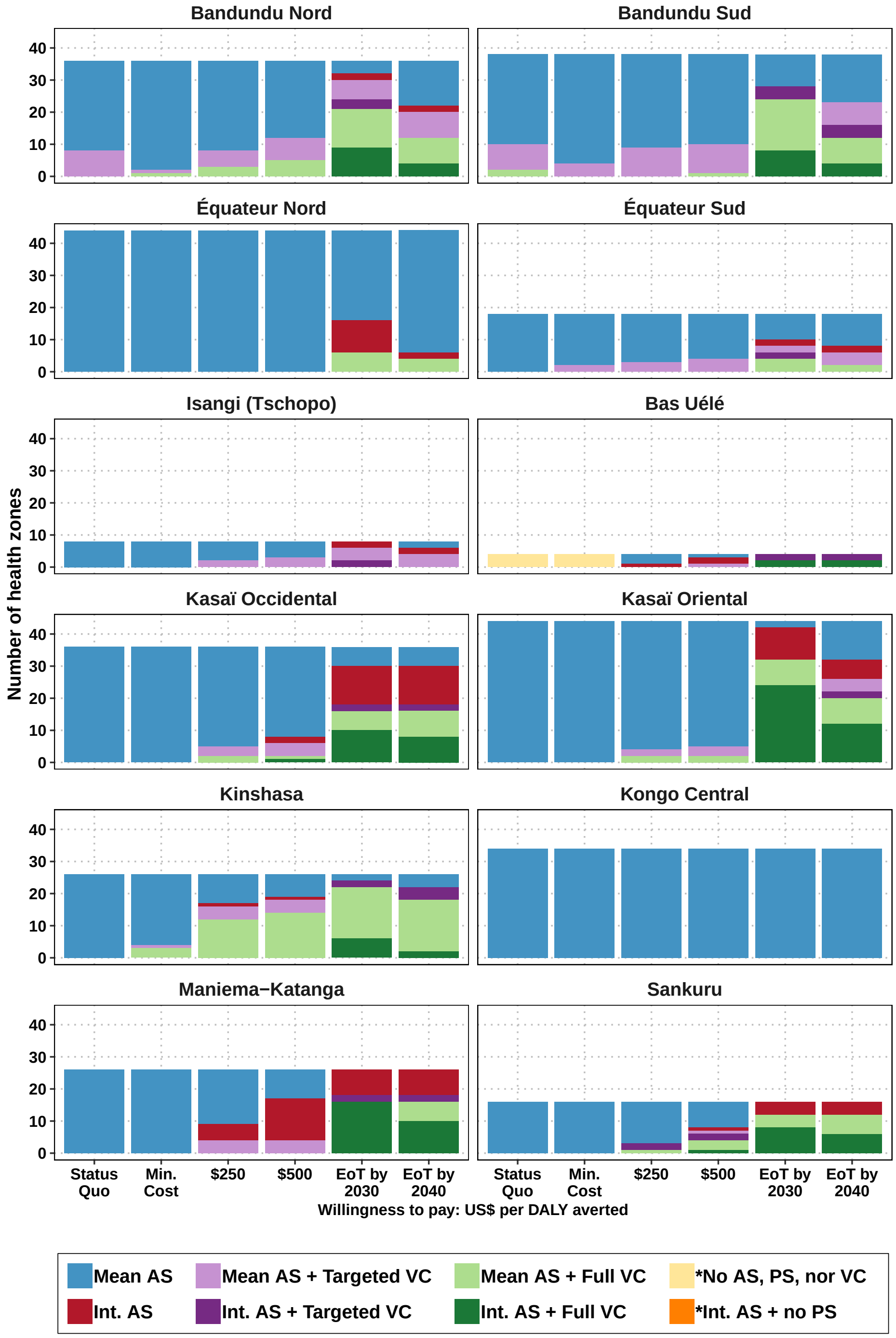

**Supplementary Figure 11**: Histrogram of optimal strategies by coordination. Abbreviations: AS: active screening, PS: passive screening, VC: vector control, DALY: disability-adjusted life-year, EoT: elimination of transmission.

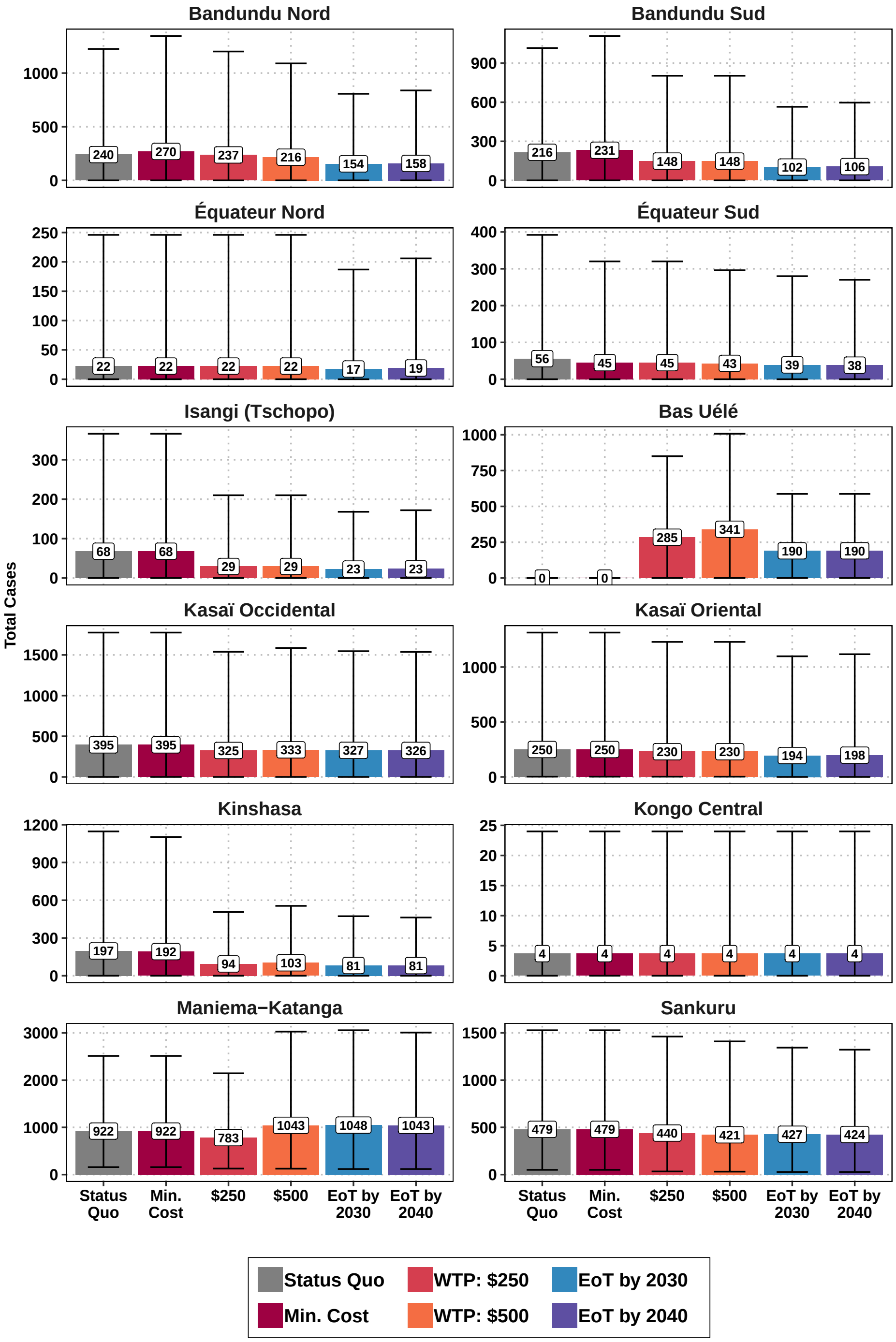

**Supplementary Figure 12**: Total cases reported 2026-2040 at different levels of investment. Abbreviations: DALY: disability-adjusted life-year, WTP: willingness-to-pay (denominated in USD per DALY averted), EoT: elimination of transmission.

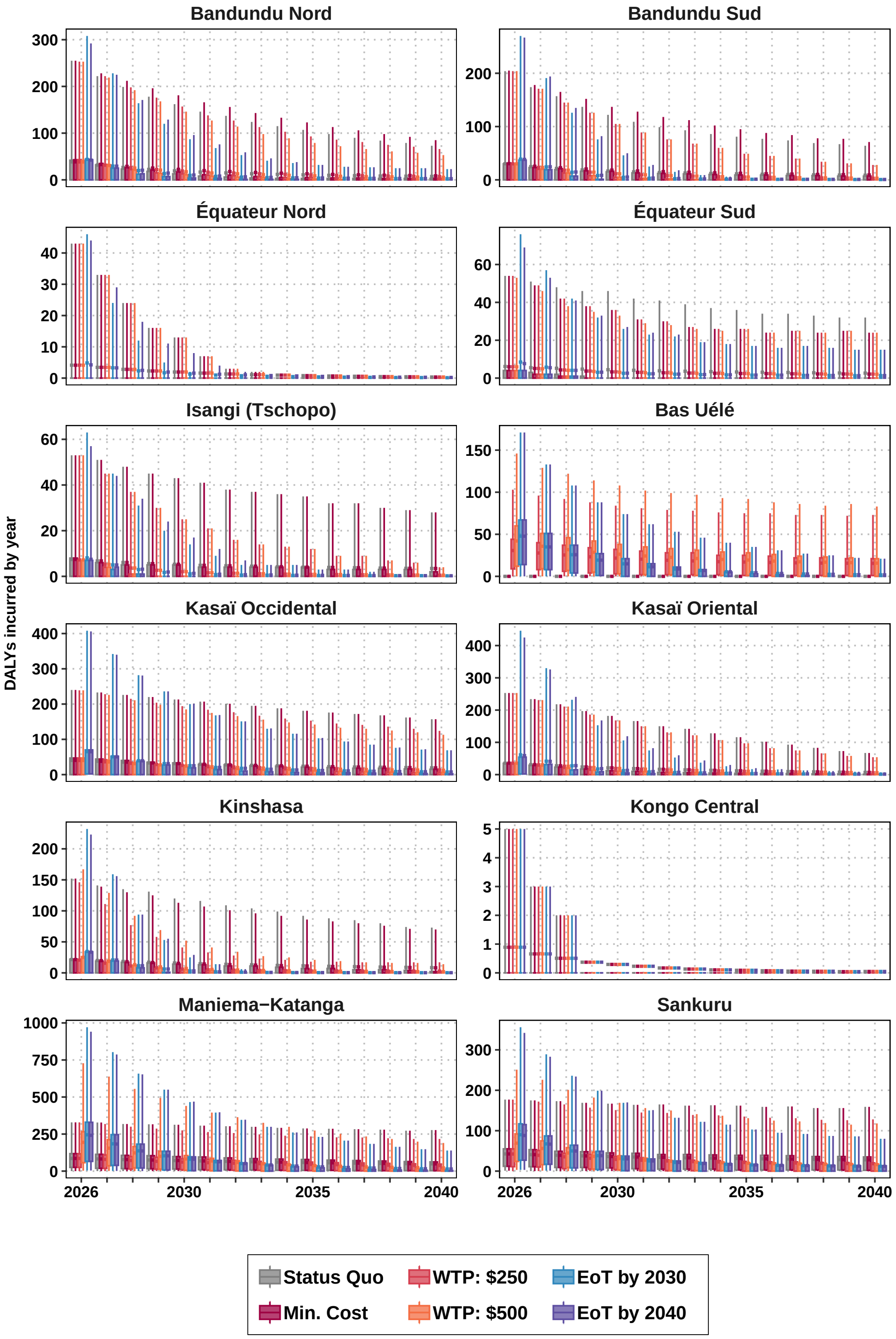

**Supplementary Figure 13**: Cases by the year 2026-2040 at different levels of investment.Abbreviations: DALY: disability-adjusted life-year, WTP: willingness-to-pay (denominated in USD per DALY averted), EoT: elimination of transmission.

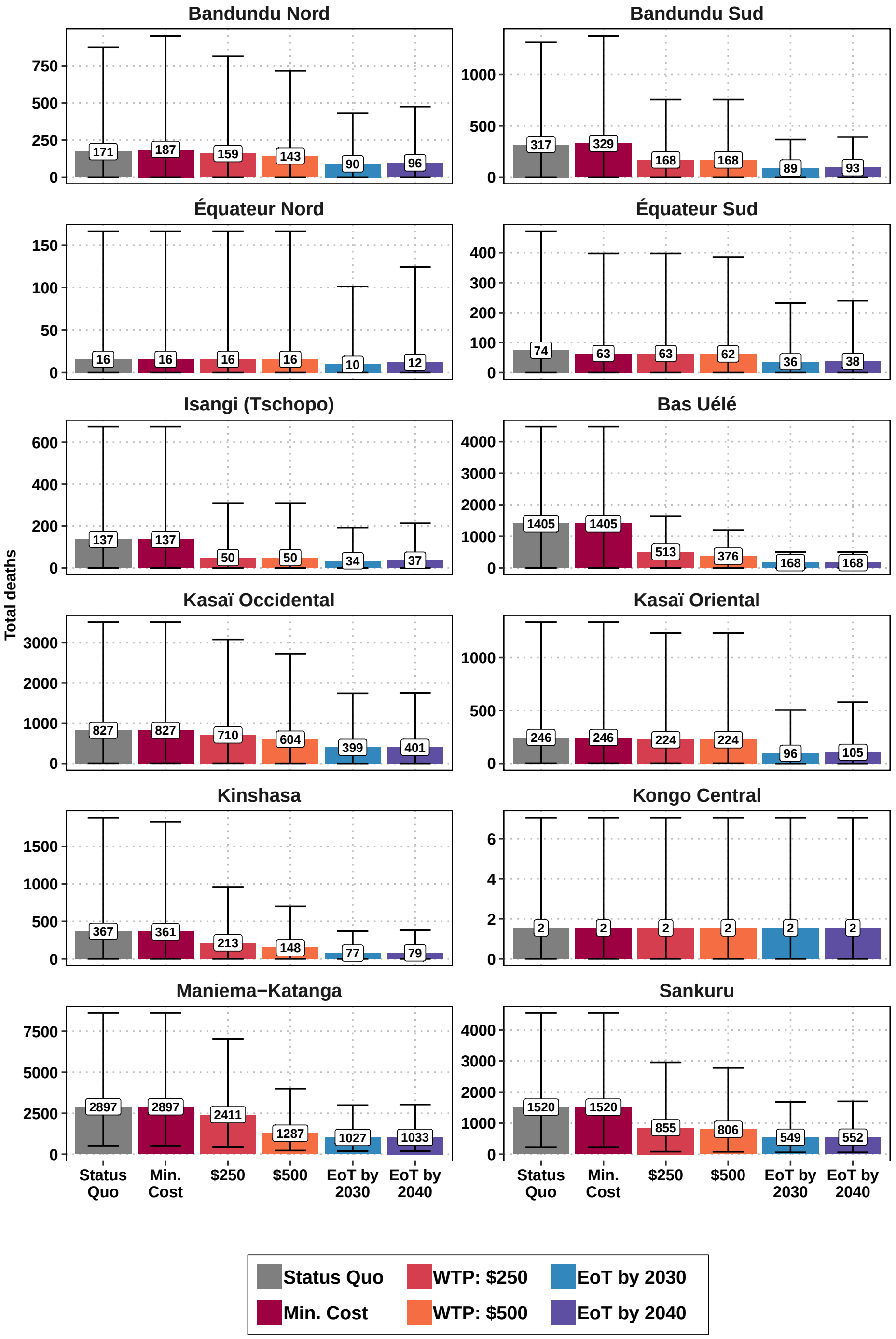

**Supplementary Figure 14**: Total deaths 2026-2040 at different levels of investment. Abbreviations: DALY: disability-adjusted life-year, WTP: willingness-to-pay (denominated in USD per DALY averted), EoT: elimination of transmission.

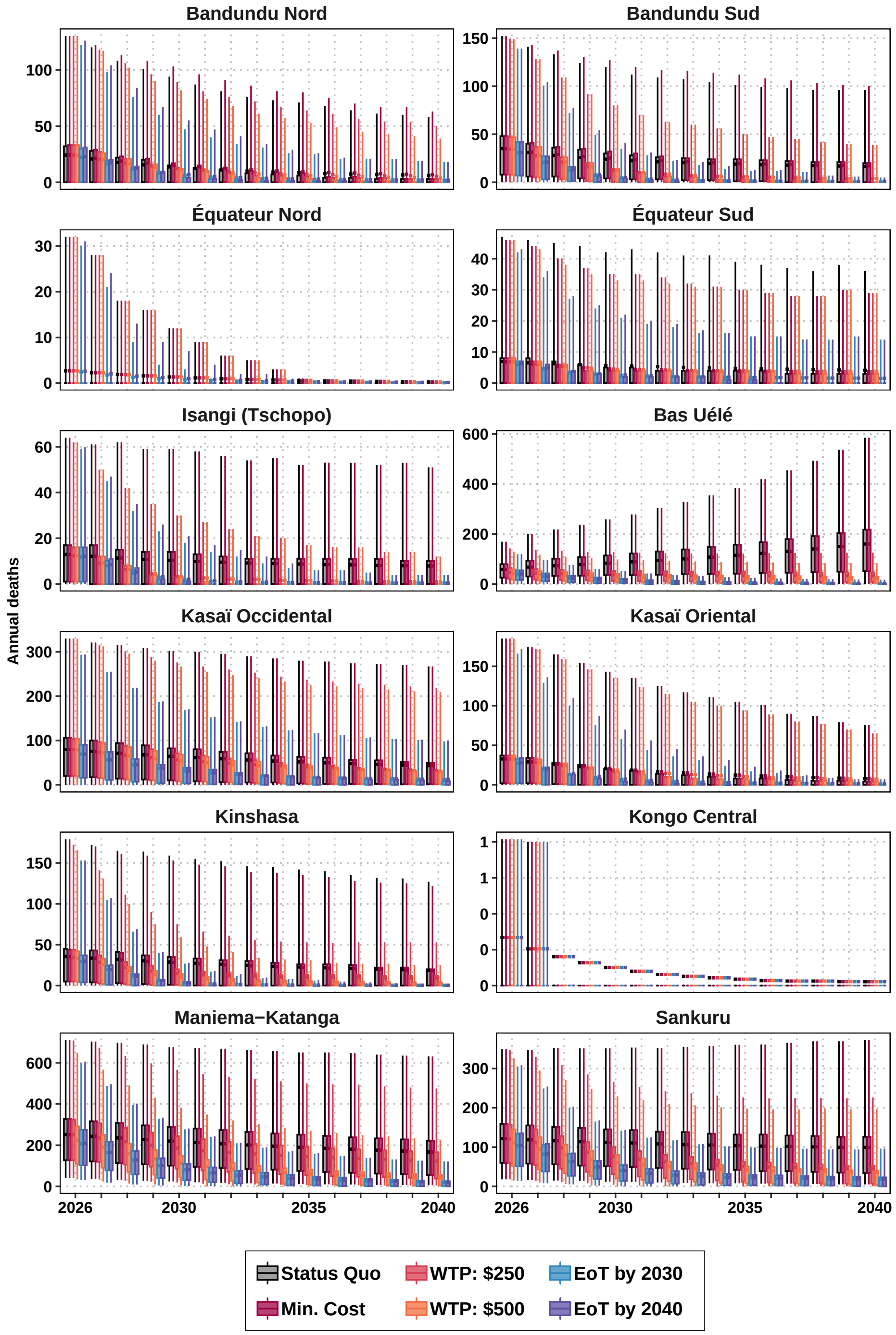

**Supplementary Figure 15**: Deaths by the year 2026-2040 at different levels of investment. Abbreviations: DALY: disability-adjusted life-year, WTP: willingness-to-pay (denominated in USD per DALY averted), EoT: elimination of transmission.

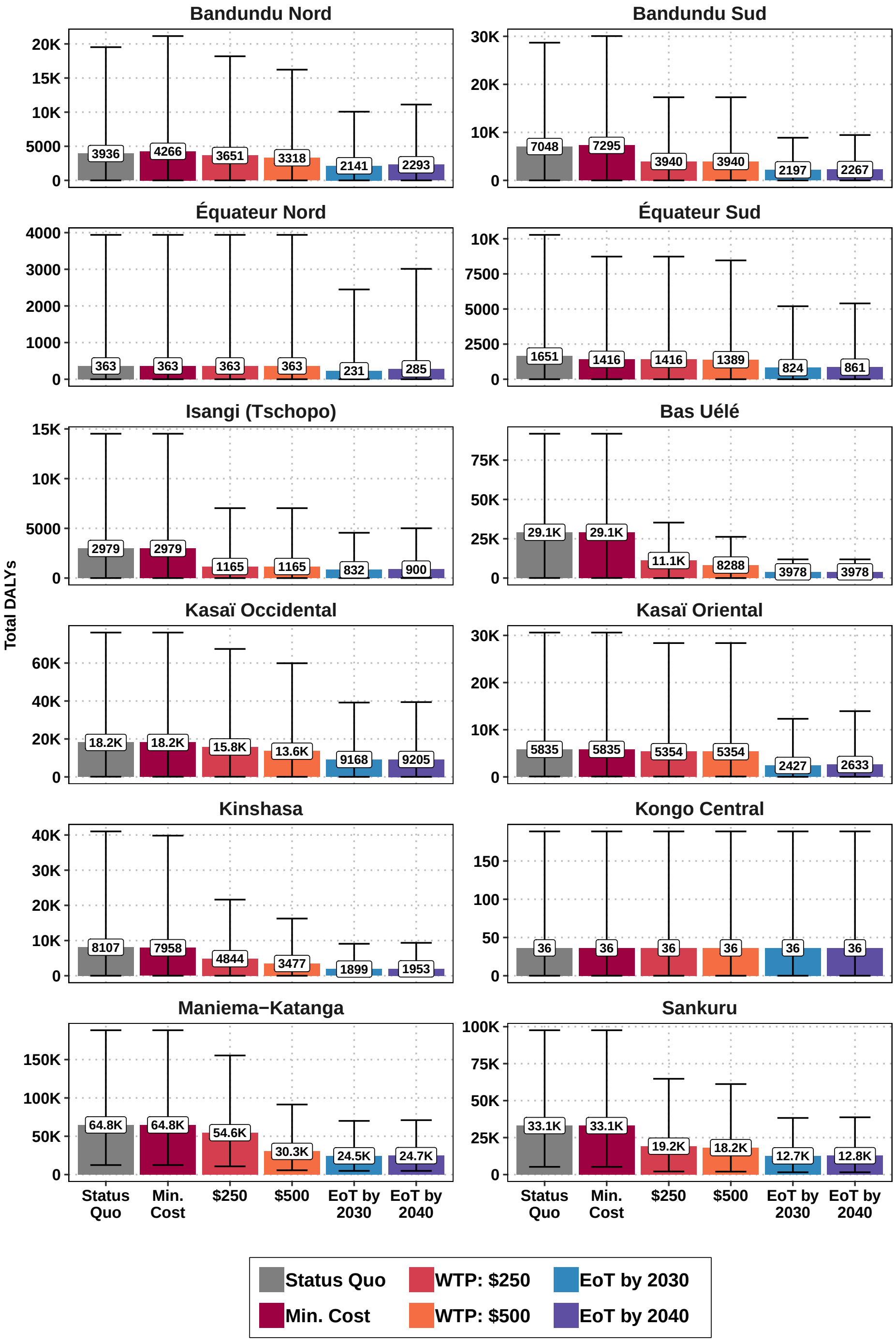

**Supplementary Figure 16**: Total DALYs 2026-2040 at different levels of investment. Abbreviations: DALY: disability-adjusted life-year, WTP: willingness-to-pay (denominated in USD per DALY averted), EoT: elimination of transmission.

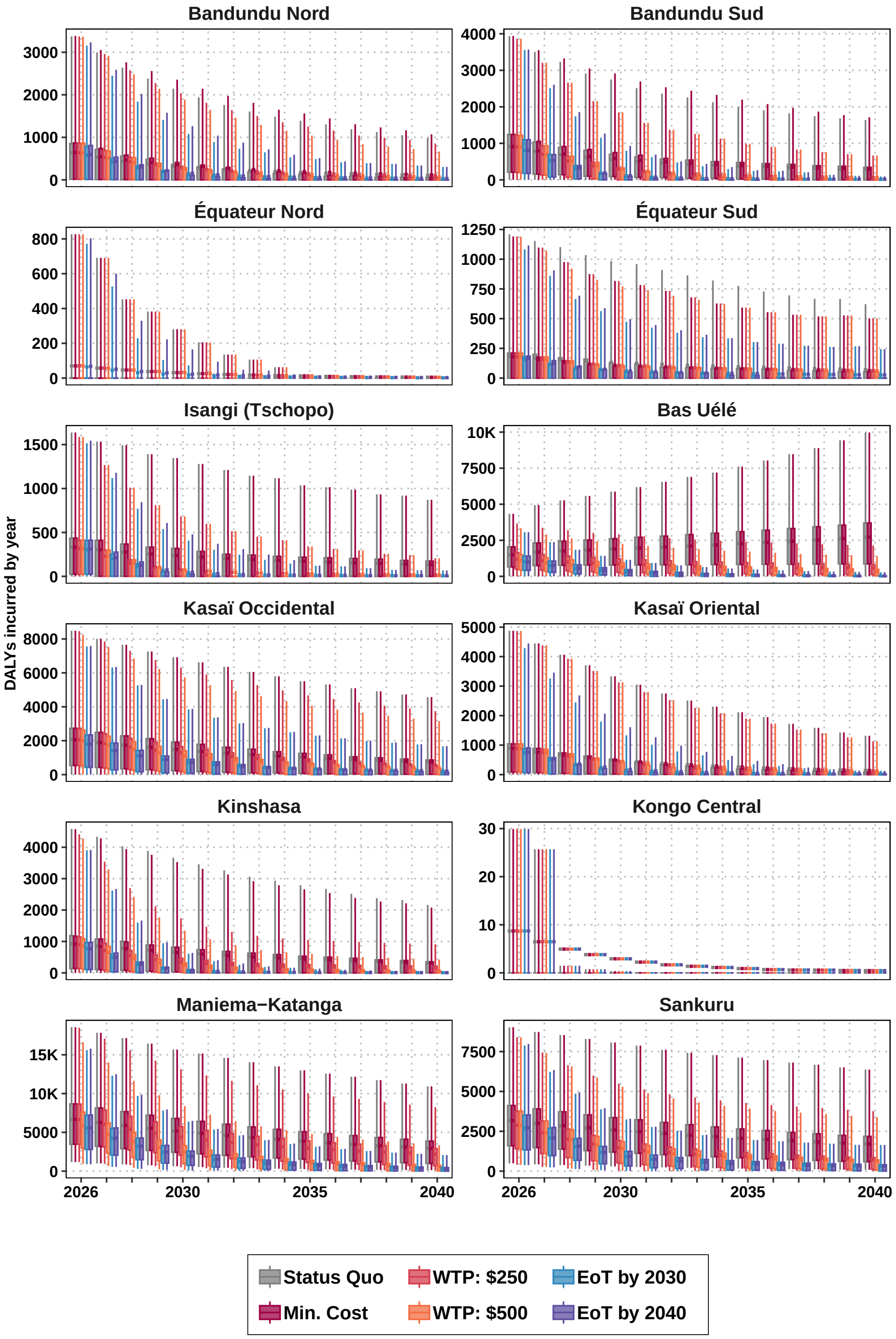

**Supplementary Figure 17**: DALYs by the year 2026-2040 at different levels of investment. Abbreviations: DALY: disability-adjusted life-year, WTP: willingness-to-pay (denominated in USD per DALY averted), EoT: elimination of transmission.

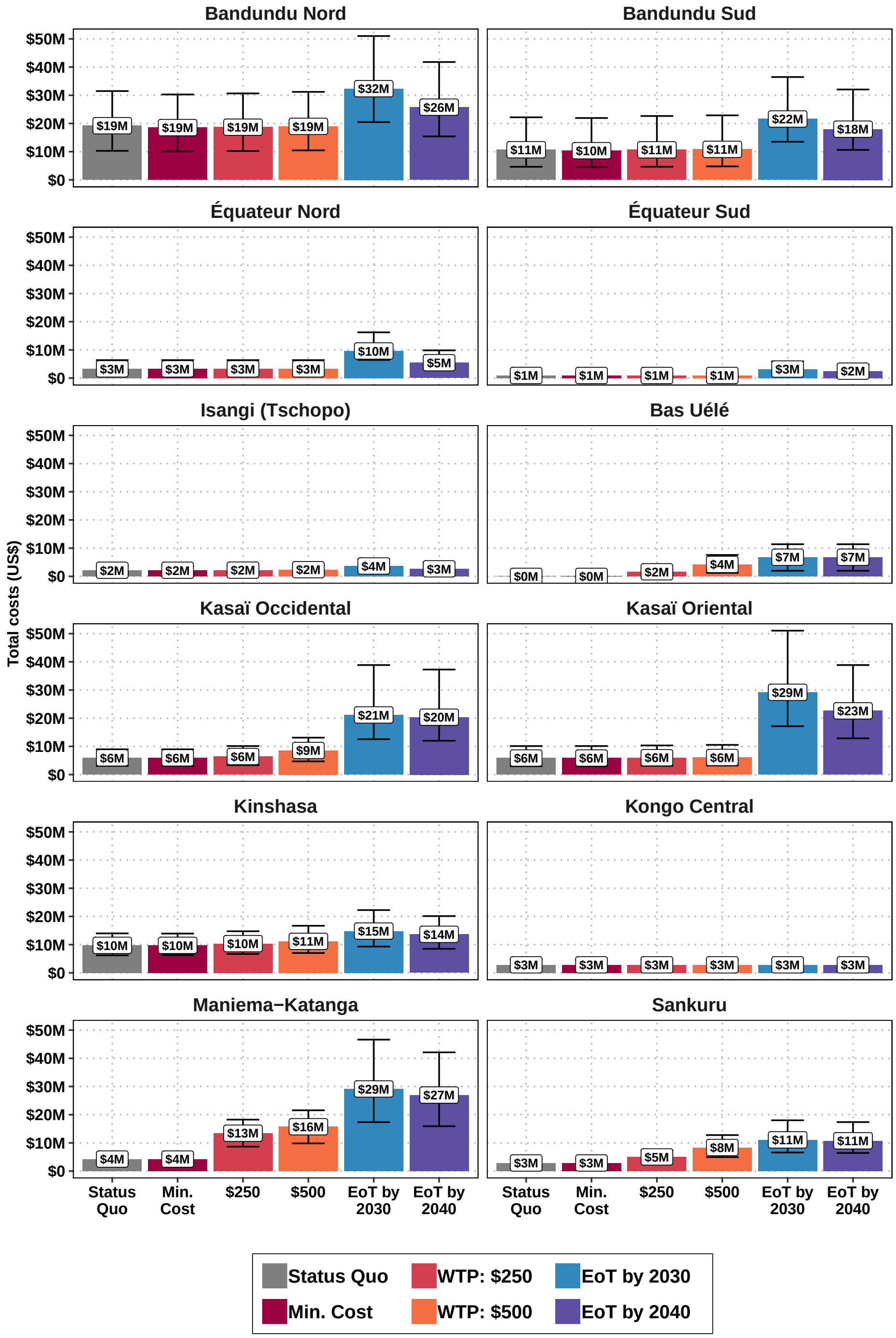

**Supplementary Figure 18**: Total costs 2026-2040 at different levels of investment. Abbreviations: DALY: disability-adjusted life-year, WTP: willingness-to-pay (denominated in USD per DALY averted), EoT: elimination of transmission.

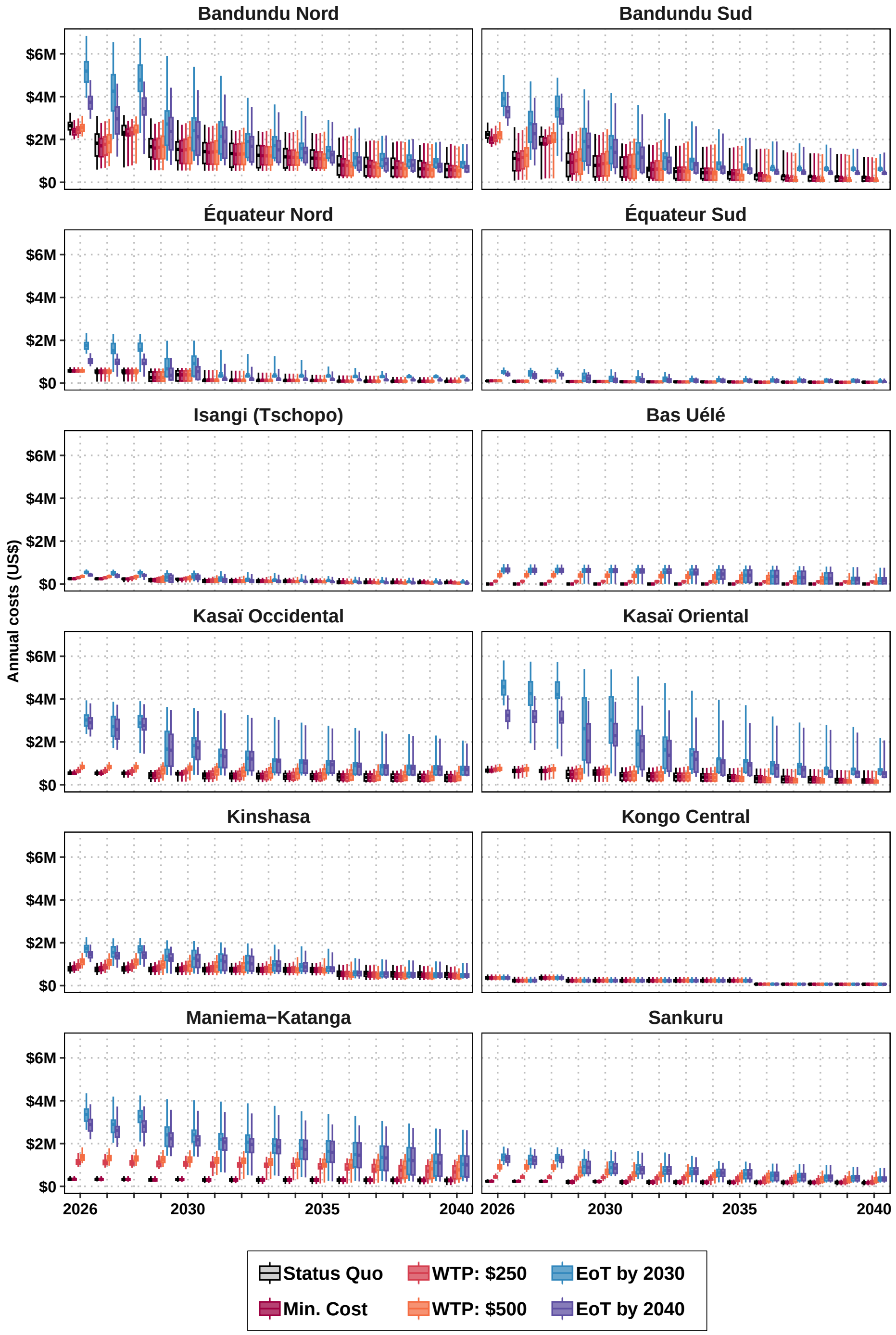

**Supplementary Figure 19**: Total costs 2026-2040 at different levels of investment. Abbreviations: DALY: disability-adjusted life-year, WTP: willingness-to-pay (denominated in USD per DALY averted), EoT: elimination of transmission.

#### B.7 Resource forecasts

↩ Return to the Supplement Table of Contents

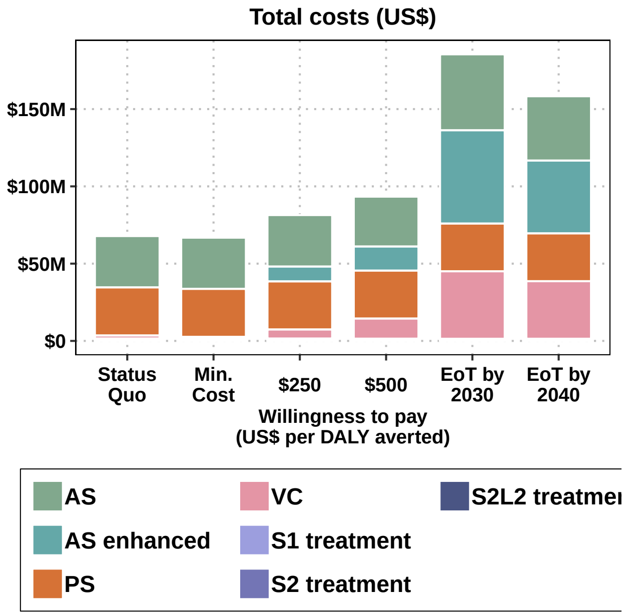

**Supplementary Figure 20**: Costs allocated to different activities, at different levels of investment, for the period 2026-2040. Abbreviations: AS: active screening, PS: passive screening, VC: vector control, S1: Stage 1 disease, S2: Stage 2 disease, S2L2: Stage 2 rescue medicine, DALY: disability-adjusted life-year, EoT: elimination of transmission.

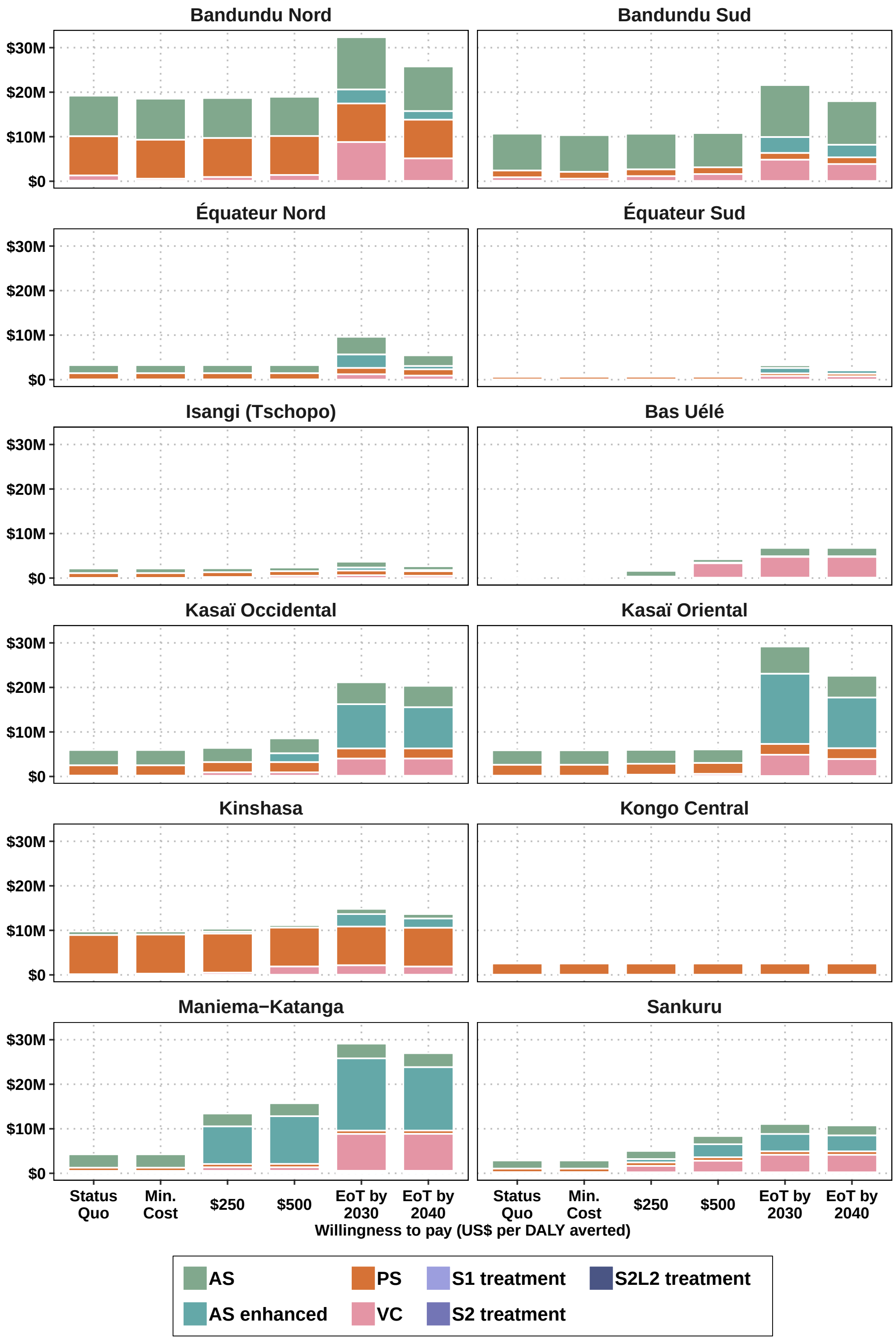

**Supplementary Figure 21**: Costs allocated to different activities, at different levels of investment by coordination, for the period 2026-2040. Abbreviations: AS: active screening, PS: passive screening, VC: vector control, S1: Stage 1 disease, S2: Stage 2 disease, S2L2: Stage 2 rescue medicine, DALY: disability-adjusted life-year, EoT: elimination of transmission.

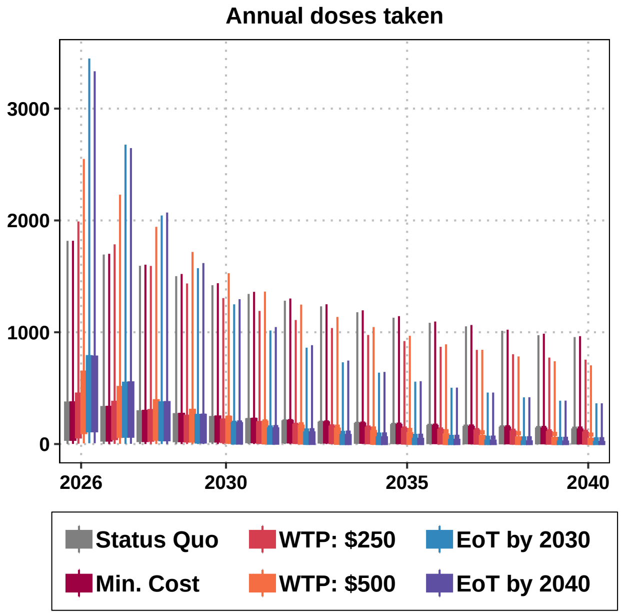

**Supplementary Figure 22**: Drugs used by the year 2026-2040 at different levels of investment. Abbreviations: DALY: disability-adjusted life-year, WTP: willingness-to-pay (denominated in USD per DALY averted), EoT: elimination of transmission.

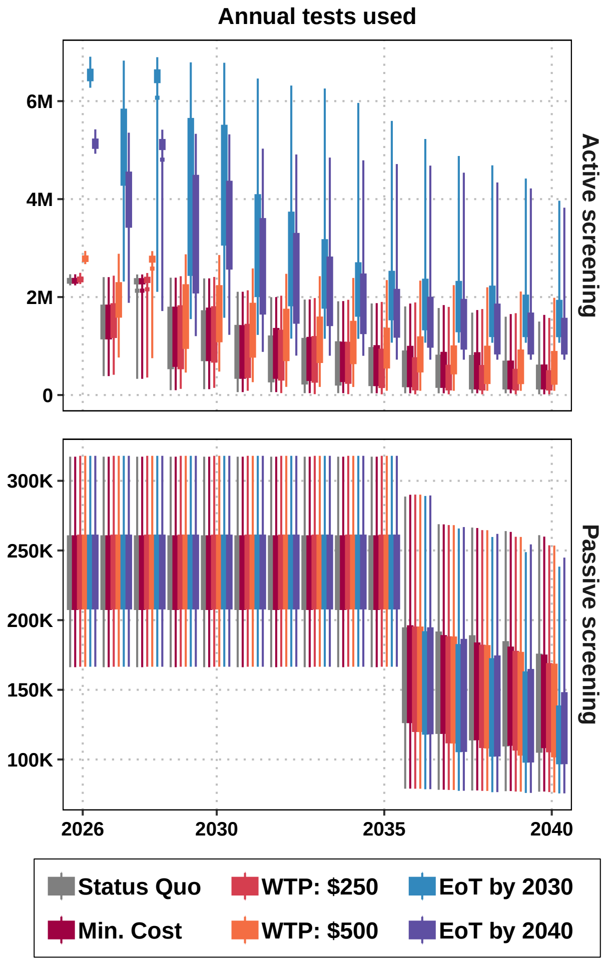

**Supplementary Figure 23**: Tests used by the year 2026-2040 at different levels of investment. Abbreviations: DALY: disability-adjusted life-year, WTP: willingness-to-pay (denominated in USD per DALY averted), EoT: elimination of transmission.

### C Supplementary Note 1: Glossary of Technical Terms

Box 1: Glossary (adapted from Antillon et. al 2022 [6] under a CC-BY 4.0 licence.)

##### EPIDEMIOLOGY TERMS

**Intervention** Interventions are separate activities to address a health need (e.g. active screening (AS) or vector control (VC)).

**Strategy** A strategy combines interventions with specific coverage and in parallel. In this paper, we simulate strategies with and without an improvement in AS and with and without VC (e.g. Strategy 1 is passive screening (PS) and mean AS, and Strategy 6 is PS, intensified AS and full VC).

**Elimination of transmission (EoT)** Globally this is the 2030 goal for gHAT; here we also consider local EOT for health zones. The feasibility of EoT is expressed as a probability equal to the proportion of our simulations in which new infections are zero before a given year (usually 2030).

**Objective** The objective is the overarching goal of the decision maker; this could be EoT by 2030, minimising costs, or delivering a cost-effective programme compared to other diseases. We assume the objective is country-wide and different regions may need different strategies to meet the objective.

**Disability-adjusted life-year (DALY)** In order to present the burden of disease as one common metric across diseases, DALYs are calculated in cost-effectiveness analyses. This is the sum of the years lived with disability due to the disease and the years of life lost by fatal cases.

##### HEALTH ECONOMICS TERMS

**Parameter uncertainty** Uncertainty in the level of transmission or the costs of interventions due to unknown underlying parameters (see Supplementary Note 3 for an explanation of our parameterisation of the health outcomes and cost model).

**Willingness-to-pay (WTP) or cost-effectiveness threshold** The amount of money that payers would pay to avert one DALY arising from the disease in the analysis (gHAT). No specific threshold is recommended, but a recent analysis shows that the WTP in DRC is between 5–230 USD per DALY averted [71–73].

**Incremental cost-effectiveness ratio** A ratio of marginal cost for a marginal benefit, calculated as follows:

$$\text{ICER} = \frac{\Delta\text{Costs}}{\Delta\text{DALYs}} = \frac{\text{Costs}_{\text{strategy}} - \text{Costs}_{\text{next best}}}{\text{Effects}_{\text{strategy}} - \text{Effects}_{\text{next best}}}$$

**Cost-effective strategy** The strategy where the ICER is less than the WTP (or cost-effectiveness threshold). We say that the cost-effective strategy is “conditional" on the WTP.

**Dominated strategy** A strategy that costs more than the minimum cost intervention while reducing the burden by a smaller degree. This strategy ought not to be implemented.

**Weakly dominated strategy (or strategies under extended dominance)** A strategy in which the ICER is higher than the next more expensive strategy. This strategy is less efficient than the next more expensive one and should not be implemented. For further illustration of weak dominance, see Supplementary Section, page 46 of Antillon et. al 2022 [6].

**Net monetary benefit** The net benefits (NMB) framework is derived from ICERs but also takes uncertainty into account.

$$\text{NMB}|\text{WTP} = \text{WTP} \times \Delta\text{DALYs} - \Delta\text{Costs}$$

The optimal strategy at a given WTP is the strategy with the highest mean NMB at that value of WTP.

**Optimal strategy** Analogous to the cost-effective strategy when no uncertainty is assumed, this is the strategy that is recommended by the NMB framework.

### D Supplementary Note 2: Health Zone-Specific Parameters

Health zone-specific parameters, grouped by coordination. For a map of the coordinations, see Supplementary Figure 1.

| Health Zone | Province | Population (2023) | AS mean | AS int. | PS clinics | VC targeted (km) | VC full (km) |
|---|---|---|---|---|---|---|---|
| **Bandundu Nord** | | | | | | | |
| Bagata | Kwilu | 210,271 | 51,314 | 73,911 | 3 | 395.18 | 458.60 |
| Bandjau | Mai-Ndombe | 147,021 | 28,749 | 54,485 | 1 | 0.00 | 49.14 |
| Bandundu | Kwilu | 242,325 | 51,895 | 103,021 | 8 | 133.47 | 178.93 |
| Bokoro | Mai-Ndombe | 276,470 | 69,388 | 97,879 | 3 | 224.85 | 444.97 |
| Bolobo | Mai-Ndombe | 173,678 | 77,928 | 111,073 | 6 | 33.33 | 211.22 |
| Bosobe | Mai-Ndombe | 160,728 | 27,352 | 52,073 | 1 | 0.00 | 178.06 |
| Djuma | Kwilu | 269,689 | 35,783 | 69,791 | 6 | 88.09 | 105.62 |
| Inongo | Mai-Ndombe | 225,939 | 1,352 | 58,469 | 1 | 0.00 | 742.18 |
| Kikongo | Kwilu | 239,132 | 43,035 | 71,489 | 3 | 79.38 | 600.64 |
| Kiri | Mai-Ndombe | 140,587 | 0 | 36,381 | 1 | 0.00 | 633.15 |
| Kwamouth | Mai-Ndombe | 161,140 | 62,336 | 89,363 | 5 | 409.29 | 813.11 |
| Mushie | Mai-Ndombe | 159,222 | 29,415 | 43,774 | 5 | 86.61 | 345.87 |
| Nioki | Mai-Ndombe | 202,109 | 20,837 | 60,441 | 3 | 355.97 | 392.12 |
| Ntand Embelo | Mai-Ndombe | 109,868 | 28,561 | 58,098 | 2 | 0.00 | 0.00 |
| Oshwe | Mai-Ndombe | 185,952 | 4,216 | 48,121 | 1 | 0.00 | 916.39 |
| Sia | Kwilu | 140,256 | 21,403 | 36,296 | 1 | 139.69 | 139.69 |
| Vanga | Kwilu | 366,524 | 38,636 | 94,850 | 2 | 93.42 | 93.42 |
| Yumbi | Mai-Ndombe | 149,793 | 29,297 | 53,380 | 2 | 44.51 | 76.02 |
| **Bandundu Sud** | | | | | | | |
| Boko | Kwango | 264,635 | 7,727 | 68,484 | 2 | 10.95 | 482.91 |
| Bulungu | Kwilu | 347,987 | 60,251 | 90,053 | 3 | 56.09 | 137.35 |
| Idiofa | Kwilu | 358,183 | 29,260 | 92,692 | 1 | 107.04 | 152.77 |
| Ipamu | Kwilu | 250,413 | 35,472 | 64,803 | 4 | 232.63 | 400.63 |
| Kasongo Lunda | Kwango | 214,302 | 17,931 | 55,458 | 1 | 0.00 | 197.65 |
| Kenge | Kwango | 358,461 | 48,759 | 92,764 | 2 | 130.44 | 400.96 |
| Kikwit Nord | Kwilu | 273,345 | 51 | 70,737 | 1 | 25.83 | 25.83 |
| Kimbau | Kwango | 227,664 | 1,511 | 58,914 | 2 | 0.00 | 429.13 |
| Kimputu | Kwilu | 248,679 | 45,957 | 73,093 | 3 | 102.59 | 174.77 |
| Koshibanda | Kwilu | 233,393 | 18,874 | 60,398 | 1 | 0.00 | 384.72 |
| Lusanga | Kwilu | 298,567 | 26,179 | 77,263 | 1 | 0.00 | 248.57 |
| Masi Manimba | Kwilu | 245,840 | 91,636 | 107,678 | 3 | 161.82 | 293.46 |
| Moanza | Kwilu | 212,771 | 9,863 | 55,061 | 2 | 0.00 | 271.92 |
| Mokala | Kwilu | 273,648 | 53,173 | 70,815 | 5 | 54.93 | 253.37 |
| Mosango | Kwilu | 153,827 | 56,588 | 76,654 | 1 | 5.50 | 54.89 |
| Mungindu | Kwilu | 155,591 | 1,624 | 40,264 | 1 | 0.00 | 313.07 |
| Pay Kongila | Kwilu | 196,891 | 1,182 | 50,951 | 1 | 0.00 | 204.74 |
| Popokabaka | Kwango | 229,297 | 25,945 | 72,688 | 1 | 0.00 | 354.76 |
| Yasa Bonga | Kwilu | 272,929 | 166,959 | 201,251 | 4 | 417.00 | 417.00 |
| **Équateur Nord** | | | | | | | |
| Bangabola | Sud-Ubangi | 173,405 | 282 | 47,084 | 1 | 0.00 | 106.05 |
| Bogose Nubea | Sud-Ubangi | 222,573 | 23,957 | 57,598 | 1 | 0.00 | 356.39 |
| Bokonzi | Sud-Ubangi | 170,691 | 300 | 44,172 | 2 | 0.00 | 266.14 |
| Bominenge | Sud-Ubangi | 198,664 | 10,807 | 51,411 | 2 | 0.00 | 25.49 |
| Boso Manzi | Mongala | 159,168 | 788 | 41,190 | 1 | 0.00 | 293.21 |
| Bosobolo | Nord-Ubangi | 204,421 | 1,570 | 52,901 | 1 | 0.00 | 220.61 |
| Boto | Sud-Ubangi | 266,045 | 14,840 | 68,848 | 1 | 0.00 | 119.72 |
| Budjala | Sud-Ubangi | 164,096 | 367 | 44,632 | 2 | 0.00 | 142.19 |
| Bulu | Sud-Ubangi | 170,517 | 814 | 44,127 | 1 | 0.00 | 87.68 |

| | | | | | | | |
|---|---|---|---|---|---|---|---|
| Businga | Nord-Ubangi | 158,634 | 3,685 | 41,052 | 2 | 0.00 | 393.59 |
| Bwamanda | Sud-Ubangi | 289,733 | 16,481 | 79,273 | 3 | 0.00 | 232.05 |
| Gbadolite | Nord-Ubangi | 179,656 | 0 | 46,492 | 1 | 0.00 | 68.26 |
| Gemena | Sud-Ubangi | 426,803 | 23,391 | 110,449 | 1 | 0.00 | 0.00 |
| Karawa | Nord-Ubangi | 310,957 | 33,788 | 110,145 | 4 | 0.00 | 78.58 |
| Kungu | Sud-Ubangi | 274,325 | 9,724 | 83,355 | 3 | 0.00 | 87.13 |
| Libenge | Sud-Ubangi | 285,692 | 0 | 73,932 | 1 | 0.00 | 273.28 |
| Loko | Nord-Ubangi | 156,329 | 18,065 | 63,411 | 1 | 0.00 | 168.60 |
| Mawuya | Sud-Ubangi | 196,193 | 0 | 50,771 | 1 | 0.00 | 301.66 |
| Mbaya | Sud-Ubangi | 84,186 | 0 | 21,786 | 1 | 0.00 | 155.51 |
| Mobayi | Nord-Ubangi | 139,236 | 0 | 36,033 | 1 | 0.00 | 256.20 |
| Ndage | Sud-Ubangi | 160,806 | 3,644 | 41,614 | 1 | 0.00 | 167.03 |
| Tandala | Sud-Ubangi | 348,292 | 23,421 | 93,818 | 2 | 0.00 | 0.00 |
| **Équateur Sud** | | | | | | | |
| Befale | Tshuapa | 212,378 | 1,384 | 54,960 | 1 | 0.00 | 891.60 |
| Bikoro | Équateur | 192,101 | 4,212 | 49,712 | 1 | 0.00 | 180.53 |
| Bomongo | Équateur | 137,605 | 485 | 35,610 | 1 | 0.00 | 547.31 |
| Iboko | Équateur | 126,985 | 673 | 32,862 | 1 | 0.00 | 61.42 |
| Irebu | Équateur | 44,625 | 1,320 | 11,549 | 1 | 0.00 | 157.94 |
| Lukolela | Équateur | 178,878 | 10,574 | 46,291 | 4 | 15.19 | 214.43 |
| Mompono | Tshuapa | 170,697 | 1,606 | 44,173 | 1 | 0.00 | 430.76 |
| Ntondo | Équateur | 82,085 | 2,213 | 21,242 | 2 | 0.00 | 61.75 |
| Wangata | Équateur | 186,862 | 0 | 48,358 | 1 | 6.16 | 24.78 |
| **Isangi-Tschopo** | | | | | | | |
| Isangi | Tshopo | 198,596 | 22,633 | 51,392 | 4 | 107.63 | 268.80 |
| Yabaondo | Tshopo | 208,445 | 21,831 | 53,942 | 11 | 141.60 | 204.47 |
| Yahisuli | Tshopo | 106,345 | 1,190 | 27,520 | 1 | 0.00 | 300.01 |
| Yakusu | Tshopo | 198,205 | 15,378 | 51,292 | 1 | 108.32 | 196.93 |
| **Kasaï Occidental** | | | | | | | |
| Bena Leka | Kasaï-Central | 357,282 | 834 | 92,458 | 1 | 0.00 | 259.52 |
| Bulape | Kasaï | 217,982 | 6,172 | 56,411 | 2 | 0.00 | 182.50 |
| Demba | Kasaï-Central | 420,288 | 157 | 108,763 | 1 | 0.00 | 187.33 |
| Dibaya | Kasaï-Central | 273,154 | 0 | 70,687 | 1 | 0.00 | 22.81 |
| Kakenge | Kasaï | 208,410 | 18,675 | 53,933 | 2 | 0.00 | 166.24 |
| Katende | Kasaï-Central | 117,038 | 14,606 | 30,286 | 1 | 0.00 | 0.00 |
| Luambo | Kasaï-Central | 349,796 | 3,422 | 90,521 | 1 | 0.00 | 286.83 |
| Lubunga 2 | Kasaï-Central | 128,006 | 21,383 | 33,126 | 1 | 112.54 | 133.41 |
| Luebo | Kasaï | 311,576 | 0 | 80,630 | 1 | 0.00 | 321.49 |
| Luiza | Kasaï-Central | 220,576 | 5,604 | 57,081 | 1 | 0.00 | 257.85 |
| Masuika | Kasaï-Central | 267,402 | 30,125 | 69,199 | 1 | 109.88 | 234.58 |
| Mikope | Kasaï | 239,645 | 114 | 62,016 | 1 | 0.00 | 475.07 |
| Mushenge | Kasaï | 209,231 | 13,078 | 54,145 | 2 | 0.00 | 27.56 |
| Mweka | Kasaï | 259,573 | 10,770 | 67,174 | 1 | 0.00 | 124.38 |
| Tshibala | Kasaï-Central | 312,867 | 2,332 | 80,965 | 1 | 0.00 | 88.52 |
| Tshikula | Kasaï-Central | 171,435 | 12,555 | 44,364 | 3 | 0.00 | 32.23 |
| Yangala | Kasaï-Central | 196,524 | 3,073 | 50,857 | 1 | 0.00 | 221.68 |
| Kapanga | Lualaba | 188,362 | 382 | 48,745 | 1 | 0.00 | 639.29 |
| **Kasaï Oriental** | | | | | | | |
| Bibanga | Kasaï-Oriental | 169,065 | 22,423 | 43,750 | 8 | 74.10 | 152.90 |
| Bonzola | Kasaï-Oriental | 259,302 | 3 | 67,103 | 2 | 20.03 | 20.03 |
| Cilindu | Kasaï-Oriental | 234,503 | 7,303 | 60,685 | 1 | 49.70 | 98.67 |
| Kabeya Kamuanga | Kasaï-Oriental | 210,427 | 9,046 | 54,455 | 2 | 0.00 | 132.99 |
| Kabinda | Lomami | 353,298 | 3,763 | 91,427 | 1 | 0.00 | 0.00 |
| Kalambayi Kabanga | Lomami | 201,056 | 19,411 | 52,030 | 2 | 70.92 | 280.32 |
| Kalenda | Lomami | 255,034 | 3,165 | 65,998 | 4 | 0.00 | 241.28 |
| Kanda Kanda | Lomami | 299,395 | 3,544 | 77,477 | 2 | 0.00 | 172.26 |
| Kasansa | Kasaï-Oriental | 265,689 | 20,311 | 68,756 | 3 | 0.00 | 138.25 |
| Lubao | Lomami | 265,120 | 1,437 | 68,608 | 1 | 0.00 | 258.07 |

| | | | | | | | |
|---|---|---|---|---|---|---|---|
| Luputa | Lomami | 372,027 | 2,389 | 96,274 | 1 | 0.00 | 251.76 |
| Miabi | Kasaï-Oriental | 188,071 | 11,256 | 48,669 | 2 | 0.00 | 40.48 |
| Mpokolo | Kasaï-Oriental | 393,264 | 3,612 | 101,770 | 3 | 0.00 | 0.00 |
| Mukumbi | Kasaï-Oriental | 152,578 | 12,057 | 39,484 | 3 | 0.00 | 84.37 |
| Mulumba | Lomami | 368,899 | 10,006 | 95,465 | 4 | 0.00 | 118.39 |
| Mwene Ditu | Lomami | 542,962 | 7,746 | 140,508 | 1 | 0.00 | 46.05 |
| Ngandajika | Lomami | 399,796 | 7,133 | 103,459 | 7 | 34.33 | 78.68 |
| Nzaba | Kasaï-Oriental | 377,559 | 2,033 | 97,707 | 1 | 0.00 | 0.00 |
| Tshilenge | Kasaï-Oriental | 382,087 | 21,963 | 98,878 | 4 | 83.99 | 83.99 |
| Tshitenge | Kasaï-Oriental | 297,337 | 7,089 | 76,946 | 3 | 88.72 | 88.72 |
| Tshitshimbi | Kasaï-Oriental | 246,246 | 6,119 | 63,724 | 1 | 0.00 | 46.32 |
| Tshofa | Lomami | 172,721 | 372 | 44,698 | 1 | 0.00 | 329.18 |
| **Kinshasa** | | | | | | | |
| Kimbanseke | Kinshasa | 385,527 | 144 | 99,768 | 3 | 19.16 | 19.16 |
| Kingabwa | Kinshasa | 287,364 | 0 | 74,366 | 1 | 0.00 | 22.75 |
| Kisenso | Kinshasa | 593,525 | 0 | 153,594 | 2 | 0.00 | 15.67 |
| Maluku 1 | Kinshasa | 246,514 | 9,808 | 63,794 | 2 | 69.42 | 345.91 |
| Maluku 2 | Kinshasa | 90,243 | 5,871 | 23,353 | 6 | 0.00 | 542.18 |
| Masa | Kongo-Central | 129,020 | 693 | 33,388 | 1 | 0.00 | 111.81 |
| Masina 1 | Kinshasa | 457,864 | 0 | 118,486 | 1 | 13.80 | 13.80 |
| Masina 2 | Kinshasa | 378,110 | 0 | 97,848 | 1 | 10.97 | 10.97 |
| Matete | Kinshasa | 395,222 | 0 | 102,277 | 1 | 0.00 | 12.02 |
| Mont Ngafula 1 | Kinshasa | 338,221 | 6,791 | 87,525 | 3 | 21.15 | 21.15 |
| Mont Ngafula 2 | Kinshasa | 198,140 | 3,479 | 51,275 | 3 | 23.15 | 23.15 |
| Ndjili | Kinshasa | 441,354 | 0 | 114,215 | 1 | 0.00 | 14.54 |
| Nsele | Kinshasa | 241,160 | 9,807 | 62,409 | 3 | 50.40 | 116.90 |
| **Kongo Central** | | | | | | | |
| Boma | Kongo-Central | 251,100 | 0 | 64,980 | 3 | 24.71 | 24.71 |
| Boma Bungu | Kongo-Central | 105,720 | 5,158 | 27,358 | 4 | 0.00 | 91.14 |
| Gombe Matadi | Kongo-Central | 132,897 | 735 | 34,391 | 4 | 0.00 | 185.77 |
| Inga | Kongo-Central | 105,758 | 12,190 | 28,177 | 9 | 0.00 | 95.61 |
| Kangu | Kongo-Central | 131,965 | 1,200 | 34,150 | 2 | 0.00 | 72.35 |
| Kibunzi | Kongo-Central | 83,098 | 3,369 | 21,504 | 5 | 0.00 | 119.40 |
| Kimpese | Kongo-Central | 234,061 | 10,811 | 60,571 | 11 | 0.00 | 240.08 |
| Kinkonzi | Kongo-Central | 91,565 | 0 | 23,695 | 1 | 0.00 | 18.55 |
| Kwilu Ngongo | Kongo-Central | 189,452 | 4,507 | 49,027 | 6 | 0.00 | 78.91 |
| Lukula | Kongo-Central | 240,077 | 7,480 | 62,128 | 9 | 0.00 | 56.53 |
| Luozi | Kongo-Central | 117,418 | 831 | 30,386 | 6 | 0.00 | 93.29 |
| Mangembo | Kongo-Central | 93,713 | 397 | 24,251 | 2 | 0.00 | 30.45 |
| Matadi | Kongo-Central | 242,113 | 146 | 62,655 | 1 | 0.00 | 43.33 |
| Muanda | Kongo-Central | 176,470 | 625 | 45,667 | 8 | 0.00 | 0.00 |
| Nsona Pangu | Kongo-Central | 142,257 | 4,672 | 36,814 | 6 | 0.00 | 378.61 |

| | | | | | | | |
|---|---|---|---|---|---|---|---|
| Seke Banza | Kongo-Central | 186,948 | 675 | 48,378 | 4 | 0.00 | 0.00 |
| Tshela | Kongo-Central | 118,731 | 0 | 30,725 | 1 | 0.00 | 51.71 |
| **Maniema Katanga** | | | | | | | |
| Kabalo | Tanganika | 313,960 | 0 | 81,247 | 1 | 0.00 | 337.91 |
| Kabambare | Maniema | 147,506 | 0 | 38,172 | 1 | 0.00 | 407.46 |
| Kalemie | Tanganika | 416,591 | 0 | 107,807 | 1 | 0.00 | 141.02 |
| Kasongo | Maniema | 282,945 | 20,606 | 73,221 | 1 | 0.00 | 181.92 |
| Kibombo | Maniema | 132,833 | 7,493 | 34,376 | 1 | 0.00 | 628.31 |
| Kongolo | Tanganika | 416,590 | 20,434 | 107,806 | 1 | 143.89 | 184.61 |
| Kunda | Maniema | 337,462 | 8,144 | 87,328 | 1 | 88.78 | 223.89 |
| Lusangi | Maniema | 215,408 | 1,797 | 55,744 | 1 | 0.00 | 234.60 |
| Mbulula | Tanganika | 236,974 | 19,509 | 61,326 | 1 | 0.00 | 0.00 |
| Nyunzu | Tanganika | 322,497 | 7,633 | 83,457 | 1 | 0.00 | 318.34 |
| Salamabila | Maniema | 152,657 | 5,218 | 39,505 | 1 | 0.00 | 127.77 |
| Samba | Maniema | 183,967 | 2,760 | 47,609 | 2 | 0.00 | 342.70 |
| Tunda | Maniema | 107,209 | 197 | 27,743 | 1 | 0.00 | 359.77 |
| **Sankuru** | | | | | | | |
| Dikungu | Sankuru | 183,360 | 19,792 | 47,449 | 3 | 0.00 | 94.21 |
| Katako Kombe | Sankuru | 174,648 | 0 | 45,196 | 1 | 0.00 | 564.42 |
| Lodja | Sankuru | 239,464 | 0 | 61,969 | 1 | 0.00 | 236.39 |
| Lusambo | Sankuru | 117,517 | 1 | 30,410 | 1 | 0.00 | 380.86 |
| Minga | Sankuru | 207,498 | 11,473 | 53,697 | 3 | 0.00 | 113.85 |
| Pania Mutombo | Sankuru | 95,225 | 611 | 24,642 | 1 | 0.00 | 233.41 |
| Tshumbe | Sankuru | 126,130 | 6,473 | 32,640 | 1 | 189.89 | 312.89 |
| Wembo Nyama | Sankuru | 109,592 | 25,408 | 29,871 | 1 | 72.90 | 279.63 |
| **Bas Uélé** | | | | | | | |
| Ango | Bas Uélé | 132,690 | 0 | 10,286 | 1 | 753.41 | 1,460.35 |
| Doruma | Haut Uélé | 87,509 | 0 | 6,273 | 1 | 0.00 | 319.71 |

### E Supplementary Note 3: Parameter Glossary

#### E.1 Principles for parameterization

(Please note that this Supplemental Note 3 contains recycled text from the authors' previous publications [6, 27]. Parameter values and citations have been updated where appropriate.)

Below are the guidelines our team follows to parameterize the parameters in the treatment and intervention model. The rationale behind the choices in the tables that describe parameters is easier with these guidelines in mind.

- **Transferability of costs across time** Costs from the literature are updated to 2024 USD values by converting to local currency units in the year of the study in the literature, inflated to 2024 values using the consumer price index (CPI) of the country, and then converted to USD using the exchange rate in 2024. It should be noted that the 2003 WHO Guide to Cost-effectiveness recommends that the GDP inflator be used (see 3.2.6 Transferability of costs across time, page 43) but we found that the data on this measure (from the World Bank) were sometimes sparse so we relied on the consumer price index instead (`'NY.GDP.DEFL.KD.ZG'` in the World Bank Development Indicator Database) [40].

- **Transferability of costs across settings** To 'borrow' data from other countries, we follow the 2003 WHO Guide to Cost-effectiveness recommendations in section 3.2.7 Transferability of costs across settings) [40]. For non-traded items (i.e. nurse and doctor time) we convert USD or LCU prices into PPP (international dollars) values in the year of the cost study and then turn the value in international dollars to local currency (still in the year of the study) of the country where a cost estimate is needed. Then, we use the CPI to inflate costs to 2024 levels and then use the exchange rate with USD to get 2024 USD values.

- **Combining multiple sources of information** Values from different publications are combined using meta-analytic methods.

- **Choice of probability distributions** Costs and ratios were modelled via gamma distributions and proportions or probability were modelled with beta distributions. These distributions were parameterized using the method of moments (see Briggs 2006 [74], Chapter 4).

- **Missing information on uncertainty: Gamma distributions.**

1. Option A: Whenever uncertainty was missing for a cost or a ratio in the literature, we assigned a gamma distribution for the parameter that would yield credible intervals between half and double the estimate.
2. Option B: If at least 2 studies listed a cost, then we take the range of the costs to parameterize a gamma distribution in which the range matched the 95 percent confidence interval (e.g. the 2.5th and 97.5th percentile). In these cases, we use a method to parameterize gamma distributions using quantiles rather than using the mean and standard error of a sample (e.g. "method of moments") [75].

- **Missing information on uncertainty: Beta distributions.**

1. Option A: Usually modeled assuming that 100 trials were observed with the **proportion_estimate x 100** as the alpha parameter and **(1-proportion_estimate) x 100** as a beta parameter.
2. Option B: If at least 2 studies listed a probability or a proportion, then we take the range of the costs to parameterize a beta distribution by assuming the range matches the 95 percent confidence interval (the 2.5th and 97.5th percentile). We use a method to parameterize Beta distributions using quantiles rather than the mean and standard error of a sample (method of moments) [76].

#### E.2 Organization of parameters

In the code repository, found in https://osf.io/ezjxb/, the parameters are in an sql database: `parameters.sqlite3`.

Additionally, a list named `epi_output` (read in from Matlab output) holds the output from the dynamic model: stage 1 and stage 2 cases detected by passive and active screening, as well as person-time spent in stage 1 and 2 before detection.

#### E.3 Summary of health outcome parameters

Below are all the parameters that model health outcomes, as well as a summary of their characteristics. An extended discussion of our choices is featured in the sections annotated in the table.

| Variable description | Variable name | Statistical Distribution | Descriptive Summary | Notes |
|---|---|---|---|---|
| **Screening** | | | | |
| Population | pop | Fixed value | Varies by health zone | See section E.5.1 |
| PS: coverage of the population per facility | ps_coverage | Beta(14, 2094) | 0.007 (0.004, 0.010) | See section E.5.2 |
| PS: number of facilities | ps_facilities | Fixed value | Varies by health zone | See section E.5.3 |
| AS: coverage | as_traditional & as_traditional_int | Fixed value | Varies by health zone | See section E.5.4 |
| AS: capacity per team per year | as_capacity_traditional | Normal(60000, 10000) | 60,055 (40,448, 79,471) | See section E.5.5 |
| CATT algorithm: diagnostic specificity | dx-spec-catt-1-in-8 | Beta(4523, 22) | 0.995 (0.993, 0.997) | See section E.5.6 |
| RDT algorithm: diagnostic sensitivity | dx-sens-rdt | Beta(230, 1) | 1.00 (0.98, 1.00) | See section E.5.7 |
| RDT algorithm: diagnostic specificity | dx_spec_rdt | Beta(226, 31) | 0.88 (0.84, 0.92) | See section E.5.8 |
| CATT algorithm: wastage during AS | dx_wastage_catt_as | Beta(8, 92) | 0.08 (0.03, 0.14) | See section E.5.9 |
| RDT algorithm: wastage during PS | dx_wastage_rdt_ps | Beta(1, 99) | 0.01 (<0.01, 0.04) | See section E.5.10 |
| **Treatment** | | | | |
| Proportion of cases age<6 | treat_prob_under6yo | Beta(152.53, 2427.9) | 0.06 (0.05, 0.07) | See section E.6.1 |
| Proportion of cases weight<35 kg among age>6 | treat_prob_under35kg | Beta(8.3, 359.6) | 0.02 (<0.01, 0.04) | See section E.6.2 |
| Proportion of S2 cases that are severe | prob_late_stage2 | Beta(76.93, 44.87) | 0.63 (0.54, 0.72) | See section E.6.3 |
| Age of death from infection | age_of_death | Gamma(148, 0.18) | 26.63 (22.41, 31.08) | See section E.7.1 |
| Length, treatment: pentamidine (days) | treat-duration-penta | Fixed value | 7 | See section E.6.4 |
| Length of hospital stay: NECT treatment | treat-duration-nect | Fixed value | 10 | See section E.6.5 |
| Length of hospital stay: fexinidazole treatment | treat-duration-fexi | Fixed value | 10 | See section E.6.6 |
| Pr. of relapse (treatment failure): pentamidine | treat_prob_failure_pent_s1 | Beta(50.3, 665.48) | 0.07 (0.05, 0.09) | See section E.6.7 |
| Pr. of relapse (treatment failure): NECT | treat_prob_failure_nect_s2 | Beta(15.87, 378.55) | 0.05 (0.02, 0.08) | See section E.6.8 |
| Pr. of relapse: fexinidazole | treat_prob_failure_fexi | Beta(9.49, 496.54) | 0.02 (<0.01, 0.03) | See section E.6.9 |
| SAE: pentamidine treatment | treat-prob-sae-pent-s1 | Beta(1.43, 551.42) | 0.002 (<0.001, 0.008) | See section E.6.10 |
| SAE: NECT treatment | treat-prob-sae_nect_s2 | Beta(40.88, 367.8) | 0.05 (0.03, 0.08) | See section E.6.11 |
| SAE: fexinidazole treatment | treat_prob_sae_fexi | Beta(3, 261) | 0.01 (<0.01, 0.03) | See section E.6.12 |
| Days lost to disability: due to SAE | treat_duration_sae | Gamma(1.22, 2.38) | 2.96 (0.14, 9.99) | See section E.6.13 |
| **Life-years lost (DALY)** | | | | |
| Life expectancy | life_expectancy | Fixed values, interpolated | Varies | See section E.7.2 |
| Disability weights: S1 disease | disability_weighting_s1 | Beta(22.96, 147.21) | 0.14 (0.09, 0.19) | See section E.7.3 |
| Disability weights: S2 disease | disability_weighting_s2 | Beta(18.37, 15.63) | 0.54 (0.37, 0.70) | See section E.7.4 |
| Disability weights: SAE | disability_weighting_sae | Uniform(0.04, 0.11) | 0.08 (0.04, 0.11) | See section E.7.5 |
| **Vector control** | | | | |
| Linear km of targets | vc_length_default & vc_length_enhanced | Fixed value | Varies by health zone | See section E.8.1 |
| Targets per km | vc_density_linear | Fixed value | Varies by health zone | See section E.8.2 |
| Replacement rate of targets per year | vc_deployments_yr | Fixed value | 2 | See section E.8.3 |

**Supplementary Table 30**: Health outcome parameters

#### E.4 Summary of cost parameters

Below are all the cost parameters and a summary of their characteristics in three tables for screening, treatment, and vector control costs. An extended discussion of our choices is featured in the sections annotated in the tables.

| Variable description | Variable name | Statistical Distribution | Descriptive Summary | Notes |
|---|---|---|---|---|
| **Screening** | | | | |
| AS: capital costs of a team | `as_cost_team_capital` | Gamma(820, 20.1) | 16,514 (15,410, 17,671) | See section E.9.1 |
| AS: fixed management costs of a team | `as_cost_team_management` | Gamma(4154, 17.21) | 71,472 (69,311, 73,658) | See section E.9.2 |
| CATT algorithm: cost per test used | `dx_cost_catt` | Gamma(1140, 0.0006897) | 0.79 (0.74, 0.83) | See section E.9.3 |
| Staging: lumbar puncture & lab exam | `dx_cost_lumbar_exam` | Gamma(3.73, 2.96) | 13.76 (3.37, 31.08) | See section E.9.4 |
| Confirmation: microscopy | `dx_cost_microscopy` | Gamma(8.47, 1.27) | 10.68 (4.70, 18.84) | See section E.9.5 |
| RDT algorithm: costs per test used | `dx_cost_rdt` | Gamma(1140, 0.002205) | $2.51 (2.37, 2.66) | See section E.9.6 |
| Indirect management costs (PNLTHA mark-up) | `program-markup` | Uniform(0.1, 0.2) | 0.15 (0.10, 0.20) | See section E.9.7 |
| PS: capital costs of a facility | `ps_cost_facility_capital` | Gamma(8.47, 226) | 2393 (1067, 4269) | See section E.9.8 |
| PS: management costs | `ps_cost_management` | Gamma(8.47, 145) | 1223 (560, 2157) | See section E.9.9 |
| **Treatment** | | | | |
| Hospital stay: cost per day | `treat_cost_ip_day` | Gamma(5.81, 0.59) | 1.39 (0.50, 2.71) | See section E.10.1 |
| Outpatient consultation: cost | `treat_cost_op_visit` | Gamma(24.2, 0.13) | 3.13 (2.02, 4.47) | See section E.10.2 |
| Course of pentamidine: cost | `rx_cost_pentamidine` | Fixed value | 54 | See section E.10.3 |
| Course of NECT: cost | `rx_cost_nect` | Fixed value | 360 | See section E.10.4 |
| Course of fexinidazole: cost | `rx_cost_fexinidazole` | Fixed value | 50 | See section E.10.5 |
| Drug delivery mark-up | `rx_delivery_markup` | Beta(45, 55) | 0.45 (0.35, 0.55) | See section E.10.6 |
| **Vector control** | | | | |
| Operational cost per kilometer of riverbank covered | `vc_cost_management` | Gamma(8.47, 57.7) | 491 (217, 869) | See section E.11.1 |
| Deployment cost per target | `vc_cost_deployment` | Gamma(8.47, 0.50) | 4.21 (1.88, 7.50) | See section E.11.2 |

**Supplementary Table 31**: Cost parameters

#### E.5 Screening parameters

##### E.5.1 Population

↩ Return to the Summary of Health Outcome Parameters.

- Name in the code: `pop`
- Source: [9]
- Country of estimate: DRC
- Statistical distribution and parameters: Fixed value
- Summary statistics (mean and 95% CI or fixed value): Varies by health zone

**Notes**

Our population data comes from the United Nations Office for the Coordination of Humanitarian Affairs censuses for national vaccine days [9]. Determined by taking the population from [9] and assuming a 3% population growth. See population summaries for the health zones included and excluded in Supplementary Table 2. In Supplementary Note 2, we provide the population for each health zone.

##### E.5.2 PS: coverage of the population per facility

↩ Return to the Summary of Health Outcome Parameters.

- Name in the code: `ps_coverage`
- Source: PNLTHA administrative data.
- Country of estimate: DRC
- Statistical distribution and parameters: Specific per coordination, see table below.
- Summary statistics (mean and 95% CI or fixed value): Varies by coordination, see table below.

**Notes**

For a summary of the clinics per coordination and nationwide, see Supplementary Table 9 in Supplementary Section A.3. Data from 2019-2021 comes from the national program, and for 2022 from the annual report.

| | Kongo Cen-tral | Bandunc Nord | Bandunc Sud | Equateu Nord | Equateu Sud | Kasai Orien-tal | Kasai Occi-dental | Maniema - Katanga | Kinshasa | Isangi | Sankuru | Whole |
|---|---|---|---|---|---|---|---|---|---|---|---|---|
| **Clinics in 2019 WHO Survey** | | | | | | | | | | | | |
| Clinics | 81 | 169 | 111 | 28 | 12 | 54 | 17 | 9 | 24 | 16 | 7 | 528 |
| **Screened** | | | | | | | | | | | | |
| 2019 | 7,187 | 98,589 | 23,167 | 179,984 | 14,282 | 35,371 | 26,623 | 18,043 | 5,355 | 9,473 | 8,006 | 426,080 |
| 2020 | 27,079 | 151,914 | 23,915 | 141,817 | 5,831 | 5,985 | 24,290 | 13,932 | 3,028 | 6,391 | 11,739 | 415,912 |
| 2021 | 4 | 104,309 | 22,342 | 244,177 | 3,743 | 13,836 | 13,418 | 9,589 | 3,162 | 12,560 | 8,086 | 435,226 |
| 2022 | 10,313 | 88,789 | 276,553 | 762 | 5,461 | 12,077 | 24,951 | 4,921 | 3,000 | 6,819 | 5,889 | 459,535 |
| **Population in health zones with clinics (millions)** | | | | | | | | | | | | |
| 2019 | 2.27 | 3.16 | 4.14 | 3.23 | 1.14 | 4.74 | 2.55 | 1.49 | 2.45 | 0.54 | 0.75 | 26.47 |
| 2020 | 2.34 | 3.26 | 4.27 | 3.32 | 1.18 | 4.88 | 2.63 | 1.53 | 2.52 | 0.55 | 0.78 | 27.26 |
| 2021 | 2.41 | 3.36 | 4.39 | 3.42 | 1.21 | 5.03 | 2.71 | 1.58 | 2.60 | 0.57 | 0.80 | 28.08 |
| 2022 | 2.48 | 3.46 | 4.53 | 3.53 | 1.25 | 5.18 | 2.79 | 1.63 | 2.68 | 0.59 | 0.82 | 28.92 |
| **Screened per site** | | | | | | | | | | | | |
| 2019 | 89 | 583 | 209 | 6,428 | 1,190 | 655 | 1,566 | 2,005 | 223 | 592 | 1,144 | 807 |
| 2020 | 334 | 899 | 215 | 5,065 | 486 | 111 | 1,429 | 1,548 | 126 | 399 | 1,677 | 788 |
| 2021 | 0 | 617 | 201 | 8,721 | 312 | 256 | 789 | 1,065 | 132 | 785 | 1,155 | 824 |
| 2022 | 127 | 525 | 2,491 | 27 | 455 | 224 | 1,468 | 547 | 125 | 426 | 841 | 870 |
| **Screened per site per 10K pop** | | | | | | | | | | | | |
| 2019 | 7 | 33 | 10 | 438 | 94 | 30 | 110 | 175 | 12 | 77 | 121 | 51 |
| 2020 | 24 | 50 | 10 | 335 | 37 | 5 | 98 | 131 | 6 | 50 | 173 | 48 |
| 2021 | 0 | 33 | 9 | 561 | 23 | 11 | 52 | 88 | 7 | 96 | 116 | 49 |
| 2022 | 9 | 27 | 105 | 2 | 33 | 9 | 95 | 44 | 6 | 51 | 82 | 50 |
| **Mean and standard error in 2019-2022** | | | | | | | | | | | | |
| Mean | 10 | 36 | 33 | 334 | 47 | 14 | 89 | 109 | 8 | 69 | 123 | 49 |
| St. Err. | 5 | 5 | 24 | 120 | 16 | 6 | 13 | 28 | 1 | 11 | 19 | 1 |
| **Distribution simulated** | | | | | | | | | | | | |
| Shape | 3.72 | 55.53 | 1.93 | 7.75 | 8.58 | 6.24 | 49.74 | 15.00 | 32.09 | 38.06 | 42.70 | 6,860.98 |
| Rate | 0.37 | 1.55 | 0.06 | 0.02 | 0.18 | 0.45 | 0.56 | 0.14 | 4.14 | 0.55 | 0.35 | 139.12 |
| Summary | 9 (3, 22) | 36 (27, 46) | 28 (4, 93) | 320 (142, 607) | 45 (21, 83) | 13 (5, 27) | 88 (66, 115) | 107 (61, 171) | 8 (5, 11) | 68 (49, 92) | 122 (89, 162) | 49 (48, 50) |

**Supplementary Table 32**: People screened in passive screening, 2019-22, and the parameters for simulations for the future. In the analysis, both Isangi - Tschopo and Isangi - Bas-Uelé will have the same parameters. The summary shows the median and 95% confidence intervals of the distribution used to simulate the coverage in the future.

##### E.5.3 PS: number of facilities

↩ Return to the Summary of Health Outcome Parameters.

- Name in the code: `ps_facilities`
- Source: [7, 35, 44]; see Supplementary Table 9 for summary by coordination.
- Country of estimate: DRC
- Statistical distribution and parameters: Fixed value
- Summary statistics (mean and 95% CI or fixed value): Varies by health zone

**Notes**

These values were retrieved from Simarro et. al. (see the supplement) and WHO surveys [35, 44]. These are facilities that can perform serological tests (CATT or RDT), microbiological confirmation, and/or treatment. In Supplementary Note 2, we provide the number of facilities for each health zone.

##### E.5.4 AS: coverage

↩ Return to the Summary of Health Outcome Parameters.

- Name in the code: `as_traditional` & `as_traditional_int`
- Source: HAT Atlas data
- Country of estimate: DRC
- Statistical distribution and parameters: Fixed value
- Summary statistics (mean and 95% CI or fixed value): Varies by health zone

**Notes**

The percent of the population that is screened by mobile teams in their villages each year. These were determined by the average percent of the population in each health zone that was screened over the years 2016-2020 (for *Mean AS*) and the maximum that was screening over the years in 2000-2020 (for *Int. AS*). See Supplementary Note 2 for the number of AS people screened per health zone.

##### E.5.5 AS: capacity per team per year

↩ Return to the Summary of Health Outcome Parameters.

- Name in the code: `as_traditional_capacity`
- Source: [43, 77, 78]
- Country of estimate: DRC
- Statistical distribution and parameters: Normal(60,000, 10,000)
- Summary statistics (mean and 95% CI or fixed value): 60,055 (40,448, 79,471)

**Notes**

The capacity of an active screening team in DRC has a mean of 60,000, a lower bound of 40,000 [77] and an upper bound of 80,000 with a work year of 220 days [43]. Teams are managed by the coordination and serve a span of multiple health zones and dozens of villages.

To parameterize the model, we chose a normal distribution with upper and lower bounds of 40-80 thousand people, therefore the parameters are Normal(60,000, 10,000).

##### E.5.6 CATT 1:8 algorithm: diagnostic specificity

↩ Return to the Summary of Health Outcome Parameters.

- Name in the code: `dx_spec_catt_1_in_8`
- Source: [79]
- Country of estimate: Various
- Statistical distribution and parameters: Beta(4523, 22)
- Summary statistics (mean and 95% CI or fixed value): 0.995 (0.993, 0.997)

**Notes**

Lumbala *et al*'s 2017 publication, which reported a CATT 1:8 specificity of 99.5% [99.3%, 99.7%] for active screening in the DRC. This was in line with their 2018 publication which reported a specificity of 99.5% [99.5%; 99.6%] in active screening and 97.6% [97.3%; 97.9%] for passive screening in the DRC. The beta distribution that corresponds with that (99.3, 99.7) as the 95% confidence interval is Beta(4523, 22).

As a form of validation, in a screening of 1.4M people (the total in 2022), that would equal 7,000 [4,200–9,800] false positives. In 2022, there were 7195 seropositives in AS, mostly done with CATT testing (Annual Report).

##### E.5.7 RDT algorithm: diagnostic sensitivity

↩ Return to the Summary of Health Outcome Parameters.

- Name in the code: `dx_sens_rdt`
- Source: [63]
- Country of estimate: Guinea and Côte d'Ivoire
- Statistical distribution and parameters: Beta(230, 1)
- Summary statistics (mean and 95% CI or fixed value): 1.00 (0.98, 1.00)

**Notes**

Based on a study in Guinea and Côte d'Ivoire [63], there was 1 sample from a gHAT patient that tested negative out of 231. Therefore, the parameter distribution for specificity is Beta(230, 1).

##### E.5.8 RDT algorithm: diagnostic specificity

↩ Return to the Summary of Health Outcome Parameters.

- Name in the code: `dx_spec_rdt`
- Source:
- Country of estimate: Guinea and Cote d'Ivoire
- Statistical distribution and parameters: Beta(1134, 11)
- Summary statistics (mean and 95% CI or fixed value): 0.990 (0.984, 0.995)

**Notes**

Over the last 5 years, the positivity rate for RDTs in PS is between 98.40 (in 2018, Fourth Stakeholders' meeting report) to 99.52 in 2022 (Annual Report). We have assigned that range as the 95% confidence interval of a beta distribution describing the specificity of the RDTs. When we derived a Beta distributions with that 95% confidence interval, we arrived at parameters of (1134, 11), or a distribution of 0.990 (0.984, 0.995).

Previous studies assigned a lower specificity, but the positivity rate would indicate specificity is now better. Based on a study in Guinea and Côte d'Ivoire [63], there were 31 samples from non-HAT patients that tested positive out of 257. Therefore, the parameter distribution for specificity is Beta(226, 31), or a specificity of 88% (95% CI: 84-92%). However, if we consider that about 400,000 people are tested in PS and a false positivity rate of 12% (the complement of the specificity), then that means that we have to confirm 48,000 patients. In 2018, 531,863 people were tested in DRC and only 8,485 RDT-positives were identified (for microscopic confirmation), and in 2022 459,535 people were tested and only 2181 people came out positive.

##### E.5.9 CATT algorithm: wastage during AS

↩ Return to the Summary of Health Outcome Parameters.

- Name in the code: `dx_wastage_catt_as`
- Source: [43]
- Country of estimate: DRC
- Statistical distribution and parameters: Beta(8, 92)
- Summary statistics (mean and 95% CI or fixed value): 0.08 (0.03, 0.14)

**Notes**

CATT tests come in packs of 50, and the list cost is assumed to consider that a pack is used on 50 patients. Once a pack is opened, one test is used as a positive control and one test is used as a negative control, so wastage is at least 4 percent. The shelf life of the test is one week in refrigeration and wastage in active screening activities is relatively low; generally, wastage of CATT tests in the context of active screening occurs at the end of the day when there are tests remaining in an open pack. To be conservative, we doubled the 4-percent lower bound for wastage and assigned the parameter a distribution of Beta(8, 92).

##### E.5.10 RDT algorithm: wastage during PS

↩ Return to the Summary of Health Outcome Parameters.

- Name in the code: `dx_wastage_rdt_ps`
- Source: [28]
- Country of estimate: DRC
- Statistical distribution and parameters: Beta(1, 99)
- Summary statistics (mean and 95% CI or fixed value): 0.01 (<0.01, 0.04)

**Notes**

We followed the same assumption as Snijders and colleagues that less than 1 percent of RDT tests would not be used [28]. Because there was no sense of uncertainty in this parameter, we assumed a Beta (1,99) distribution.

#### E.6 Treatment parameters

##### E.6.1 Proportion of cases age<6

↩ Return to the Summary of Health Outcome Parameters.

- Name in the code: `treat_prob_under6yo`
- Source: [53, 65]
- Country of estimate: South Sudan
- Statistical distribution and parameters: Beta(152.53, 2427.9)
- Summary statistics (mean and 95% CI or fixed value): 0.06 (0.05, 0.07)

**Notes**

There were only two studies where the number of children under 5 or 6 years of age was stated explicitly [53, 65].

| Study | Groups | Under 6 y.o. | Total | Est. | 95% CI |
|---|---|---|---|---|---|
| *Schmid 2012* | Single arm | 35 | 629 | 0.06 | (0.04−0.08) |
| *Eperon 2006* | Pentamidine | 56 | 850 | 0.07 | (0.05−0.08) |
| *Eperon 2006* | Melarsoprol | 62 | 1108 | 0.06 | (0.04−0.07) |
| **Common effect model** | | | **2587** | **0.06** | **(0.05−0.07)** |
| **Random effects model** | | | | **0.06** | **(0.05−0.07)** |

Heterogeneity: $I^2 = 0\%$, $\tau^2 = 0$, $p = 0.60$

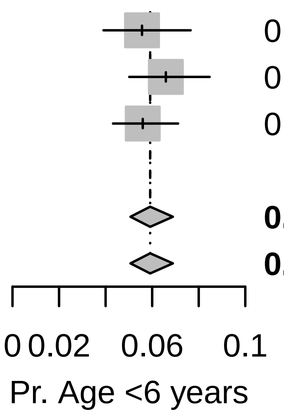

Because the data showed non-significant heterogeneity according to the tau-squared test for heterogeneity, we have chosen to use the fixed-effects combined estimate: 0.059 (0.051, 0.069). The beta parameters of the random-effects estimate Beta(153.53, 2427.90) for the probability of that a patient is under 6 years old.

##### E.6.2 Proportion of cases weight<35 kg among age>6

↩ Return to the Summary of Health Outcome Parameters.

- Name in the code: `treat_prob_under35kg`
- Source: [47–49, 54, 56, 57, 60–62, 65]
- Country of estimate: Various
- Statistical distribution and parameters: Beta(8.3, 359.6)
- Summary statistics (mean and 95% CI or fixed value): 0.02 (<0.01, 0.04)

**Notes**

To determine whether a patient is eligible for fexinidazole treatment, we could not find any studies that would tell us the number of HAT patients who weighed less than 35 kg, but we have estimated the number of people who might weigh less than 35 kg by examining the distribution of weight among patients in the trials in the literature. Furthermore, we have examined how this variable is related to potential selection by age of the study population. We are interested in the proportion of older children and adults that might weigh less than 35 kg, as age under 6 is a contraindication for fexinidazole.

We fit a gamma distribution by the method of moments to the reported mean and standard deviations of each of the studies. For Priotto 2012, no SD was reported, but an interquartile range was reported, so we fit a gamma distribution by the method of Cook [75].

We then took the expected number of people under 35 kg, and then performed a single-proportion meta-analysis with the expected number of people in each study under and over the 35 kg threshold.

| Citation | Group | Age Group | Mean weight | Measure of spread | No. of obser-vations | Gamma distr. alpha par. | Gamma distr. beta par. | Prop. <35kg | Simulated No. <35kg |
|---|---|---|---|---|---|---|---|---|---|
| Priotto 2006 | Melarsoprol and Nifurtimox | All ages | 49.20 | SD = 14.4 | 18 | 11.67 | 4.21 | 0.16 | 3 |
| Priotto 2006 | Melarsoprol and Eflornithine | All ages | 50.00 | SD = 10.3 | 19 | 23.56 | 2.12 | 0.06 | 1 |
| Priotto 2006 | NECT | All ages | 51.40 | SD = 8.4 | 17 | 37.44 | 1.37 | 0.02 | 0 |
| Priotto 2007 | NECT | Over 15 years old | 51.70 | SD = 7.4 | 52 | 48.81 | 1.06 | 0.01 | 0 |
| Priotto 2007 | Eflornithine | Over 15 years old | 53.10 | SD = 7.2 | 51 | 54.39 | 0.98 | 0.00 | 0 |
| Checchi 2007 | NECT | All ages | 44.80 | SD = 15.1 | 31 | 8.80 | 5.09 | 0.28 | 9 |
| Priotto 2009 | NECT | Over 15 years old | 53.00 | SD = 8.7 | 143 | 37.11 | 1.43 | 0.01 | 2 |
| Priotto 2009 | Eflornithine | Over 15 years old | 53.90 | SD = 8.3 | 143 | 42.17 | 1.28 | 0.01 | 1 |
| Ngoyi 2010 | Pentamidine and Melarsoprol | Over 12 years old | 56.00 | SD = 10.0 | 360 | 31.36 | 1.79 | 0.01 | 3 |
| Priotto 2012 | Single arm | All ages | 49.00 | IQR: 40-56 | 2190 | 16.37 | 2.96 | 0.12 | 265 |
| Schmid 2012 | Single arm | All ages | 45.00 | SD = 16.0 | 629 | 7.91 | 5.69 | 0.29 | 182 |
| Burri 2016 | Pentamidine | Over 15 years old and >35 kg | 48.50 | SD = 7.6 | 40 | 40.83 | 1.19 | 0.03 | 1 |
| Pohlig 2016 | Pentamidine | Over 12 years old and >30 kg | 45.70 | SD = 7.8 | 137 | 34.15 | 1.34 | 0.08 | 10 |
| Pohlig 2016 | Pafuramidine | Over 12 years old and >30 kg | 44.70 | SD = 7.9 | 136 | 32.02 | 1.40 | 0.10 | 14 |
| Kansiime 2018 | All | Over 15 years old | 51.69 | SD = 9.7 | 109 | 28.22 | 1.83 | 0.03 | 3 |
| Mesu 2018 | NECT | Over 15 years old | 50.70 | SD = 9.6 | 130 | 27.89 | 1.82 | 0.04 | 5 |
| Mesu 2018 | Fexinidazole | Over 15 years old | 50.50 | SD = 8.2 | 264 | 37.93 | 1.33 | 0.02 | 5 |

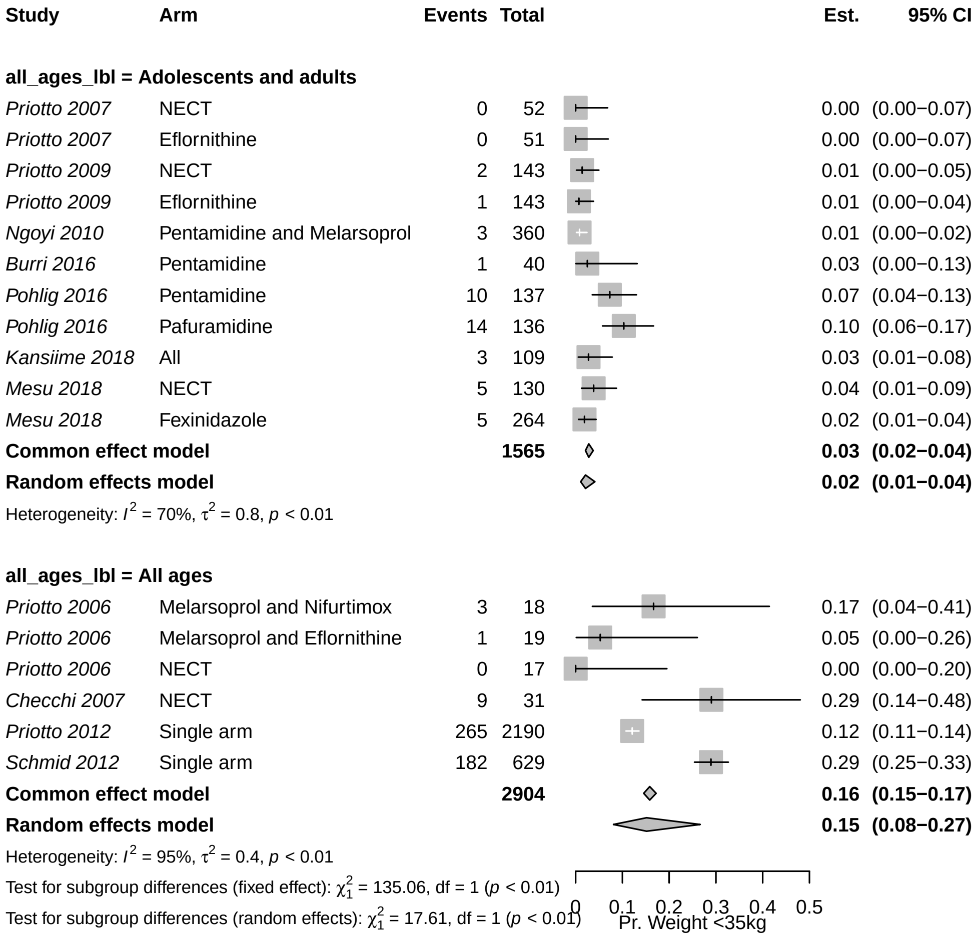

Because the data showed significant heterogeneity according to the tau-squared test for heterogeneity, we have chosen to use the random-effects estimate: 0.02 (0.01-0.04), represented by probability distribution: Beta(8.30, 359.61).

##### E.6.3 Proportion of S2 cases that are severe

↩ Return to the Summary of Health Outcome Parameters.

- Name in the code: `prob_late_stage2`
- Source: [38, 48, 49, 53, 54, 60, 61, 65]
- Country of estimate: Various
- Statistical distribution and parameters: Beta(76.93, 44.87)
- Summary statistics (mean and 95% CI or fixed value): 0.63 (0.54, 0.72)

**Notes**

The definition of severe stage 2 gHAT disease by the WHO is when there are more than 100 white blood cells (WBC, leukocytes) per micro-litre in the cerebrospinal fluid. We have searched the clinical trials for the proportion of stage 2 patients that have high concentrations of leukocytes upon admission to treatment.

| Study | Arm | Events | Total | Est. | 95% CI |
|---|---|---|---|---|---|
| *Priotto 2006* | Melarsoprol and Nifurtimox | 9 | 18 | 0.50 | (0.26−0.74) |
| *Priotto 2006* | Melarsoprol and Eflornithine | 12 | 19 | 0.63 | (0.38−0.84) |
| *Priotto 2006* | NECT | 6 | 17 | 0.35 | (0.14−0.62) |
| *Eperon 2006* | Melarsoprol | 407 | 1108 | 0.37 | (0.34−0.40) |
| *Priotto 2007* | NECT | 31 | 52 | 0.60 | (0.45−0.73) |
| *Priotto 2007* | Eflornithine | 39 | 51 | 0.76 | (0.63−0.87) |
| *Checchi 2007* | NECT | 18 | 31 | 0.58 | (0.39−0.75) |
| *Priotto 2009* | NECT | 107 | 143 | 0.75 | (0.67−0.82) |
| *Priotto 2009* | Eflornithine | 115 | 143 | 0.80 | (0.73−0.87) |
| *Ngoyi 2010* | Pentamidine and Melarsoprol | 209 | 272 | 0.77 | (0.71−0.82) |
| *Priotto 2012* | Single arm | 1373 | 2190 | 0.63 | (0.61−0.65) |
| *Schmid 2012* | Single arm | 397 | 629 | 0.63 | (0.59−0.67) |
| **Common effect model** | | | **4673** | **0.58** | **(0.57−0.60)** |
| **Random effects model** | | | | **0.63** | **(0.55−0.71)** |

Heterogeneity: $I^2 = 96\%$, $\tau^2 = 0.3$, $p < 0.01$

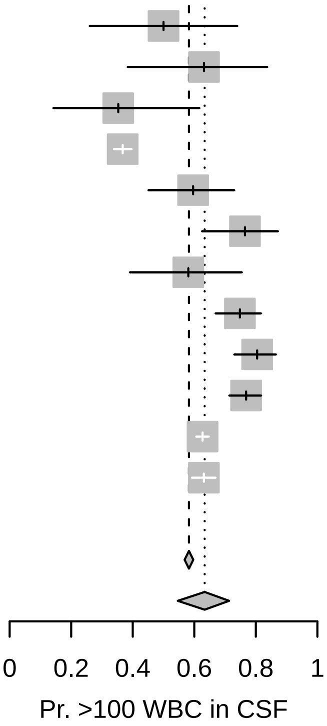

Because the data showed significant heterogeneity according to the tau-squared test for heterogeneity, we have chosen to use the random-effects combined estimate: 0.634 (0.546, 0.713), represented by the probability distribution Beta(76.93, 44.87).

##### E.6.4 Length of hospital stay: Pentamidine treatment

↩ Return to the Summary of Health Outcome Parameters.

- Name in the code: `treat_duration_penta`
- Source: [38]
- Country of estimate: Global recommendations
- Statistical distribution and parameters: Fixed value
- Summary statistics (mean and 95% CI or fixed value): 7

**Notes**

For NECT treatment, patients must stay in inpatient care for a minimum of 7 days for the eflornithine infusions, and whether they stay for a total of 10 days for nifurtimox administration is unclear. For the most recent clinical trial [47], NECT patients were released on days 13-18 after admission, but we have assumed that for the most recent trial, the average patient can be released from care after 10 days in the hospital.

##### E.6.5 Length of hospital stay: NECT treatment

↩ Return to the Summary of Health Outcome Parameters.

- Name in the code: `treat_duration_nect`
- Source: [38, 47]
- Country of estimate: Global recommendations
- Statistical distribution and parameters: Fixed value
- Summary statistics (mean and 95% CI or fixed value): 10

**Notes**

For NECT treatment, patients must stay in inpatient care for a minimum of 7 days for the eflornithine infusions, and whether they stay for a total of 10 days for nifurtimox administration is unclear. For the most recent clinical trial [47], NECT patients were released on days 13-18 after admission, but we have assumed that for the most recent trial, the average patient can be released from care after 10 days in the hospital.

##### E.6.6 Length of hospital stay: fexinidazole treatment

↩ Return to the Summary of Health Outcome Parameters.

- Name in the code: `treat_duration_fexi`
- Source: [38, 47]
- Country of estimate: Global recommendations
- Statistical distribution and parameters: Fixed value
- Summary statistics (mean and 95% CI or fixed value): 10

**Notes**

For the only trial that is published [47], patients were released on days 13-18 after the initiation of treatment, although the treatment only took 10 days, so we have assumed that in routine care the average patient will be in inpatient care for 10 days.

##### E.6.7 Pr. of relapse: pentamidine

↩ Return to the Summary of Health Outcome Parameters.

- Name in the code: `treat_prob_failure_pent_s1`
- Source: [52–57]
- Country of estimate: Various
- Statistical distribution and parameters: Beta(50.3, 665.48)
- Summary statistics (mean and 95% CI or fixed value): 0.07 (0.05, 0.09)

**Notes**

The WHO guidelines for the treatment of HAT in 2019 [38] presented existing data on treatment failure of pentamidine treatment. To produce one comprehensive estimate of treatment failure, we performed a meta-analysis on proportions within a single group.

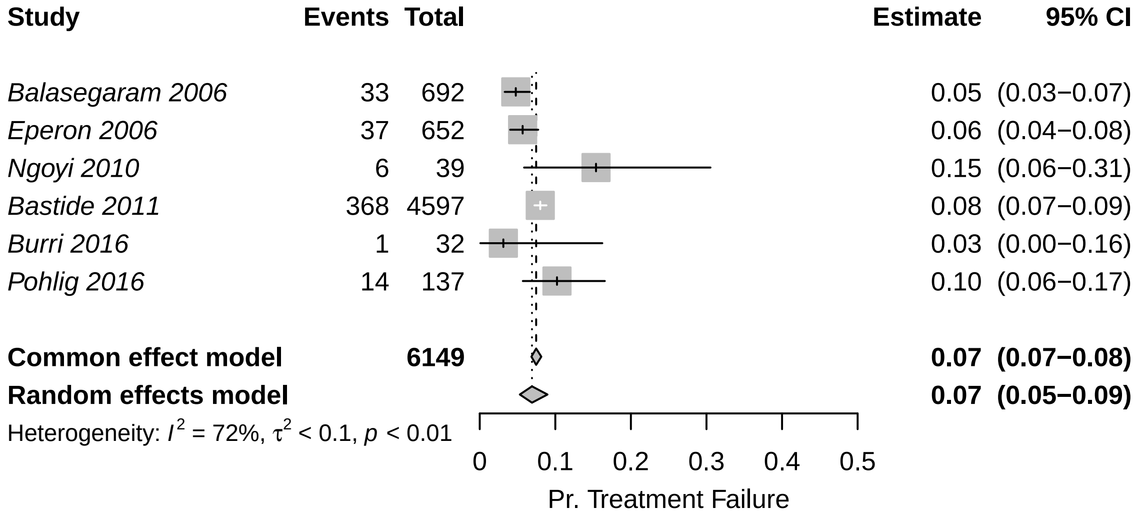

Because the data showed significant heterogeneity according to the tau-squared test for heterogeneity, we have chosen to use the random-effects estimate, to which we assigned a distribution of Beta(50.30, 665.47).

##### E.6.8 Pr. of relapse (treatment failure): NECT

↩ Return to the Summary of Health Outcome Parameters.

- Name in the code: `treat_prob_failure_nect_s2`
- Source: [47–49, 60–62]
- Country of estimate: Various
- Statistical distribution and parameters: Beta(15.87, 378.55)
- Summary statistics (mean and 95% CI or fixed value): 0.05 (0.02, 0.08)

**Notes**

The WHO guidelines for the treatment of HAT in 2019 [38] presented existing data on treatment failure of NECT. Kansiime and colleagues [62] also performed a systematic review of studies estimating the outcomes of NECT treatment. To produce one comprehensive estimate of treatment failure, we performed a meta-analysis on proportions within single groups.

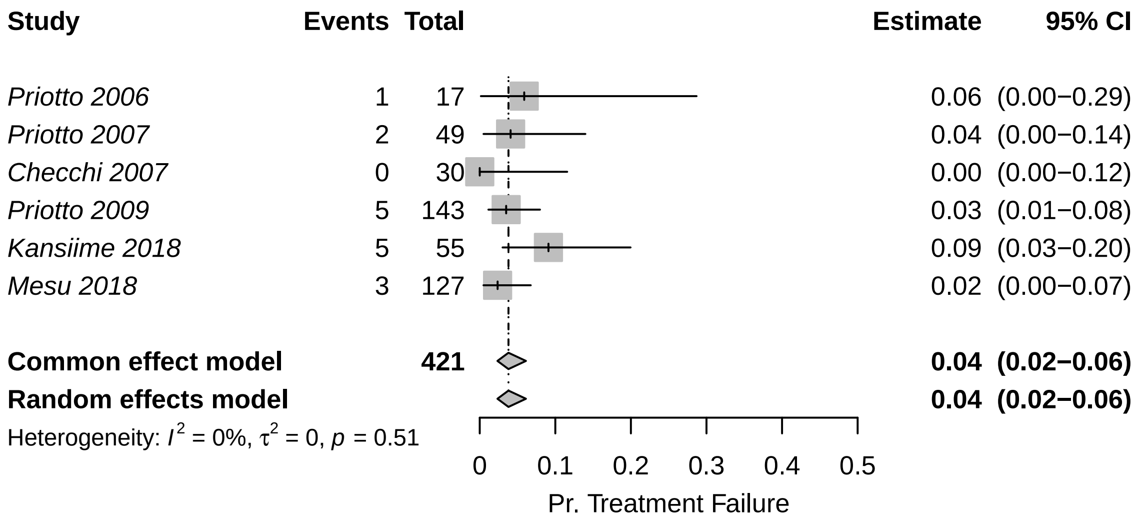

Because the data showed non-significant heterogeneity according to the tau-squared test for heterogeneity, we have chosen to use the fixed-effects estimate, to which we assigned a distribution of Beta(15.87, 378.55)

##### E.6.9 Pr. of relapse: fexinidazole

↩ Return to the Summary of Health Outcome Parameters.

- Name in the code: `treat_prob_failure_fexi`
- Source: [38]
- Country of estimate: DRC
- Statistical distribution and parameters: Beta(9.49, 496.54)
- Summary statistics (mean and 95% CI or fixed value): 0.02 (<0.01, 0.03)

**Notes**

Mesu and colleagues [47] have published the only study on fexinidazole treatment effectiveness in late-stage 2 cases. Moreover, the accompanying meta-analysis for the WHO treatment guidelines released in 2019 shows the outcomes of an additional extension study on stage 1, both early and late-stage 2 disease as well for the data from Mesu et al stratified by the concentration of WBC in the CSF [38].

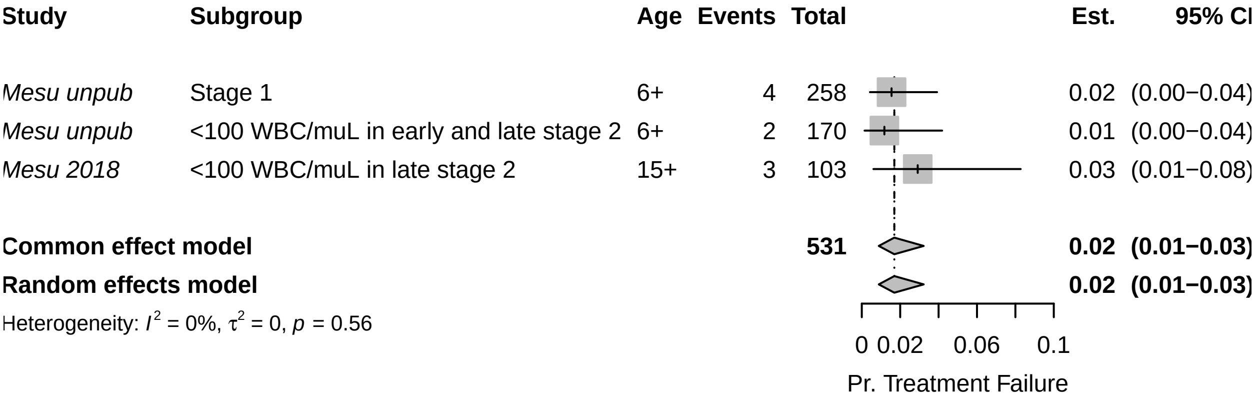

Because the data showed non-significant heterogeneity according to the tau-squared test for heterogeneity, we have chosen to use the fixed-effects combined estimate: 0.017 (0.009, 0.032), for which we assigned a distribution of Beta(9.49, 496.54).

##### E.6.10 SAE: pentamidine treatment

↩ Return to the Summary of Health Outcome Parameters.

- Name in the code: `treat_prob_sae_pent_s1`
- Source: [53, 56, 57]
- Country of estimate: DRC and South Sudan
- Statistical distribution and parameters: Beta(1.43, 551.42)
- Summary statistics (mean and 95% CI or fixed value): 0.002 (<0.001, 0.008)

**Notes**

As part of the WHO guidelines for the treatment of HAT in 2019 ([38]), Cochrane performed a systematic review of studies that evaluated the efficacy of NECT compared to fexinidazole studies and presented the probability of serious adverse events. Severe or serious adverse events in studies for S1 treatment were defined as "significant hazard, contra-indication, side effect, or precaution" [53, 56, 57].

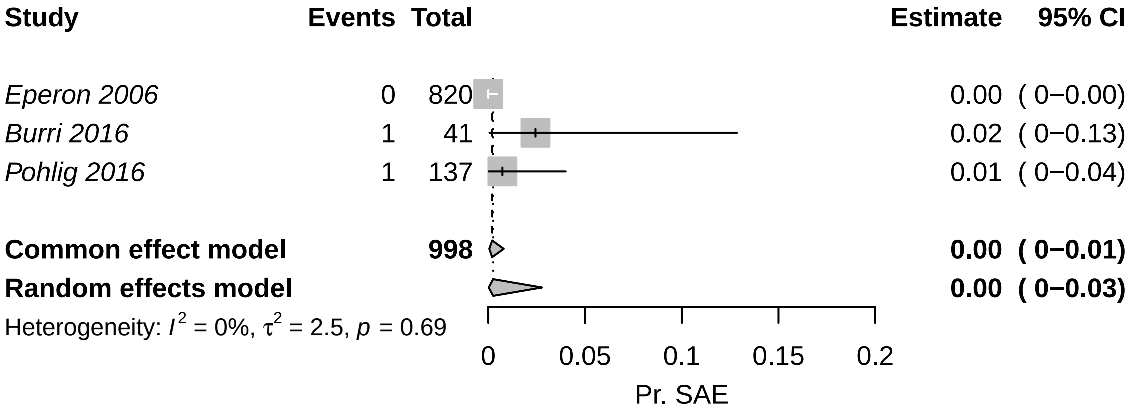

Because the results do not contain evidence of significant heterogeneity, we have chosen to use the fixed (pooled) estimate of 0 (0-0.01), which would result from a beta distribution of Beta(1.43, 551.42).

##### E.6.11 SAE: NECT treatment

↩ Return to the Summary of Health Outcome Parameters.

- Name in the code: `treat_prob_sae_nect_s2`
- Source: [47–49, 60–62]
- Country of estimate: DRC
- Statistical distribution and parameters: Beta(40.88, 367.8)
- Summary statistics (mean and 95% CI or fixed value): 0.05 (0.03, 0.08)

**Notes**

The WHO guidelines for treatment of gHAT in 2019 [38], presented all NECT studies to date, as did Kansiime and colleagues [62]. We searched through these studies for evidence of the probability of severe adverse vents (SAEs), described as events of Grade 3 or higher according to the National Cancer Institute Common Toxicity Criteria for Adverse Events (CTCAE).

To produce one comprehensive estimate of the probability of SAE, we performed a meta-analysis on proportions within single groups.

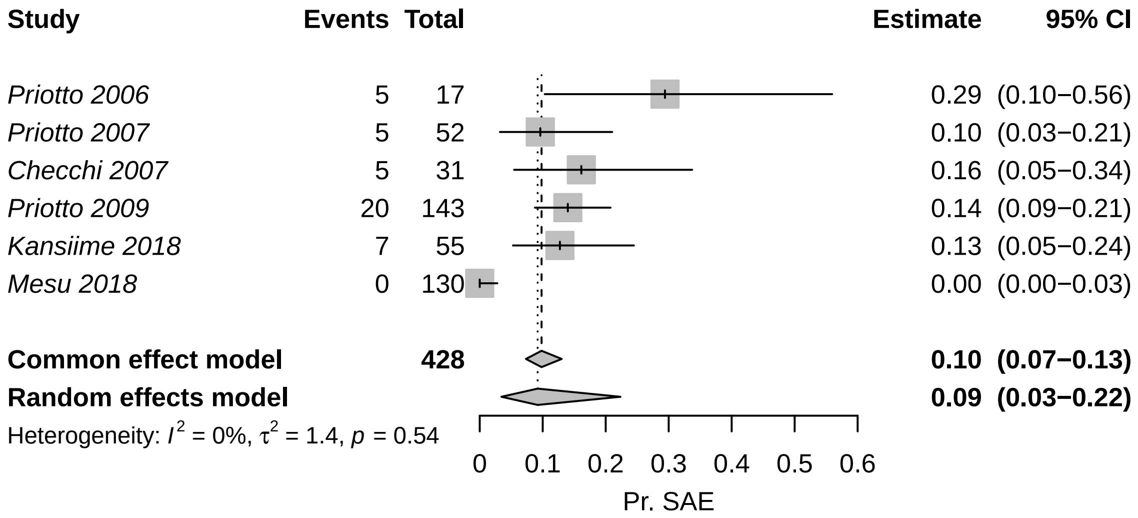

Reproduced from Antillon *et al* [6] with permission under a CC-BY license.

Because the data showed non-significant heterogeneity according to the tau-squared test for heterogeneity, we have chosen to use the fixed-effects estimate: 0.098 (0.073, 0.130), which would result from a beta distribution of Beta(40.88, 367.80).

##### E.6.12 SAE: fexinidazole treatment

↩ Return to the Summary of Health Outcome Parameters.

- Name in the code: `treat_prob_sae_fexi`
- Source: [47]
- Country of estimate: DRC
- Statistical distribution and parameters: Beta(3, 261)
- Summary statistics (mean and 95% CI or fixed value): 0.01 (<0.01, 0.03)

**Notes**

There is only one published study on fexinidazole, so the probability of serious adverse events will be parameterized with the observations from that study: 4 adverse events attributable to fexinidazole in 3 people among 264 people, so we have assigned a distribution of Beta(3, 261).

There were additional data reported in the appendix to the WHO's interim guidelines ([38]) related to studies that are ongoing. However, since we do not know details about whether those SAEs were attributable to treatment, we have chosen to omit those data.

##### E.6.13 Days lost to disability: due to SAE

↩ Return to the Summary of Health Outcome Parameters.

- Name in the code: `treat_duration_sae`
- Source: [67]
- Country of estimate: DRC
- Statistical distribution and parameters: Gamma(1.22, 2.38)
- Summary statistics (mean and 95% CI or fixed value): 2.96 (0.14, 9.99)

**Notes**

Our only source of information for the duration of severe adverse events (SAEs) is Alirol 2013 [67], which lists the most common adverse events and the median duration of these events. Most events last a median of 1-2 days (with interquartile ranges reaching up to 4 days).

For simplicity, we have fit a gamma distribution with interquartile range of 1-4 days. Our distribution is therefore Gamma(1.22, 2.38) with a mean and 95% confidence interval of 2.92 (0.12, 9.95), which provides a sufficiently large range of values in light of the scarce information we have.

#### E.7 Life-years lost (DALY) parameters

↩ Return to the Summary of Health Outcome Parameters.

##### E.7.1 Age of death from infection

↩ Return to the Summary of Health Outcome Parameters.

- Name in the code: `age_of_death`
- Source: [47–49, 52–54, 56, 57, 60–62, 65–67]
- Country of estimate: DRC and South Sudan
- Statistical distribution and parameters: Gamma(148, 0.18)
- Summary statistics (mean and 95% CI or fixed value): 26.63 (22.41, 31.08)

**Notes**

No good registry of the age of infection exists, so we have searched through the literature that we have used to parameterize the model for the average age of HAT patients. Among the studies that we have used to inform other parameters, eight studies reported age information in a sample of patients of all ages, and nine studies reported the age information in a sample of older children or adults (12-15 years and older).

However, the data exists in a state that is difficult to synthesize:

Therefore, we have fit a gamma distribution to the means and medians of the studies that included patients of all ages. We have omitted the median from Alirol et al. 2013 [67] as this median seems unusually high – even higher than the mean age of patients in studies where only adults (over the age of 15) were recruited. Our distribution is therefore Gamma(147.93, 0.18) with a mean and 95% confidence interval of 27 (23, 31), which provides a sufficiently large bound of uncertainty in lieu of the information we have.

##### E.7.2 Life expectancy

- Name in the code: `life_expectancy`
- Source: [80]
- Country of estimate: DRC
- Statistical distribution and parameters: Interpolation
- Summary statistics (mean and 95% CI or fixed value): See below.

**Notes**

We took age-specific life expectancy at around the time when people die of HAT in DRC for 2019, the last year for which there are estimates.

We see here that the expected years of life left at each of the ages is:

Using these data, we made an interpolating function in R (function: `approxfun`) that would calculate the life years left for each age of death (see previous parameter, `age_of_death`).

##### E.7.3 Disability weights: S1 disease

- Name in the code: `disability_weighting_s1`
- Source: [69]
- Country of estimate: GBD
- Statistical distribution and parameters: Beta(22.96, 147.21)
- Summary statistics (mean and 95% CI or fixed value): 0.14 (0.09, 0.19)

**Notes**

The Global Burden of Disease listed the impact of sleeping sickness as equivalent to the health state labelled “Motor plus cognitive impairments, severe” and their estimate for a disability weight is 0.542 (0.374-0.702), using the 2013 weight values. No distinction was made between stage 1 and 2 of the disease.

| Citation | Group | Age Group | Summary |
|---|---|---|---|
| Priotto 2006 | Melarsoprol and Nifurtimox | All ages | Mean: 29.1 range: 5-56 |
| Priotto 2006 | Melarsoprol and Eflornithine | All ages | Mean: 28.1 range: 11-61 |
| Priotto 2006 | NECT | All ages | Mean: 29.1 range: 9-62 |
| Balasegaram 2006 | Pentamidine | All ages | 148 under 15 and 504 over 15 |
| Eperon 2006 | Pentamidine | All ages | 56 patients 0-5, 226 patients 6-15, 568 patients 15+ |
| Eperon 2006 | Melarsoprol | All ages | 63 patients 0-5, 249 patients 6-15, 796 patients 15+ |
| Priotto 2007 | NECT | Over 15 years old | Mean: 33.1 range: 15-69 |
| Priotto 2007 | Eflornithine | Over 15 years old | Mean: 36.1 range: 15-70 |
| Checchi 2007 | NECT | All ages | Mean: 23.9 range: 4-45 |
| Priotto 2009 | NECT | Over 15 years old | Mean: 32.8 SD: 12.5 |
| Priotto 2009 | Eflornithine | Over 15 years old | Mean: 34.6 SD: 13.5 |
| Ngoyi 2010 | Pentamidine | Over 12 years old | Mean: 35 SD: 13 |
| Ngoyi 2010 | Pentamidine and Melarsoprol | Over 12 years old | Mean: 34 SD: 12 |
| Priotto 2012 | Single arm | All ages | Median: 24 IQR: 15-35 |
| Schmid 2012 | Single arm | All ages | 35 patients 0-4 yo, 65 patients 5-11 yo, and 529 patients 12 yo or more. |
| Hasker 2012 | All | All ages | Median: 27 IQR: 16-40 |
| Alirol 2013 | Single arm | All ages | Median: 36 IQR: 20-50 |
| Burri 2016 | Pentamidine | Over 15 years old and >35 kg | Median: 31 range: 15-50 |
| Pohlig 2016 | Pentamidine | Over 12 years old and >30 kg | Median: 31 range: 13-75 |
| Pohlig 2016 | Pafuramidine | Over 12 years old and >30 kg | Median: 30 range: 12-64 |
| Kansiime 2018 | NECT | Over 15 years old | Mean: 27.23 SD: 12.07 |
| Kansiime 2018 | Eflornithine | Over 15 years old | Mean: 27.33 SD: 8.59 |
| Mesu 2018 | NECT | Over 15 years old | Mean: 35.2 SD: 13.2 |
| Mesu 2018 | Fexinidazole | Over 15 years old | Mean: 34.5 SD: 12.6 |

| Age | Years left |
|---|---|
| 15-19 | 52.4 |
| 20-24 | 48.1 |
| 25-29 | 44.0 |
| 30-34 | 39.9 |
| 35-39 | 35.7 |
| 40-44 | 31.6 |
| 45-49 | 27.6 |
| 50-54 | 23.6 |

Life-years left for each age group. Source: https://www.who.int/data/gho/data/indicators/indicator-details/GHO/gho-ghe-life-tables-by-country. Variable: *expectation of life at age x*.

While this seems appropriate for the second stage of sleeping sickness, for stage 1 disability we chose to use the disability weights for equivalent to “infectious disease, acute episode, severe”, which is described as “has a high fever and pain, and feels very weak, which causes great difficulty with daily activities” and has a much lower disability weight equivalent to 0.133 (0.088-0.190). The distribution for this parameter is therefore Beta(22.96, 147.21).

It should be noted that other cost-effectiveness analyses have used different values for disability weights [78, 81, 82]. These values arise from the 1994 Global Burden of Disease Study but we prefer to consider updated values. Since most of the disability is due to deaths rather than illness during life, we do not believe that this difference is cause for concern.

##### E.7.4 Disability weights: S2 disease

- Name in the code: `disability_weighting_s2`
- Source: [69]
- Country of estimate: GBD
- Statistical distribution and parameters: Beta(18.37, 15.63)
- Summary statistics (mean and 95% CI or fixed value): 0.54 (0.37, 0.70)

**Notes**

The Global Burden of Disease listed the impact of sleeping sickness as equivalent to the health state labeled “Motor plus cognitive impairments, severe” and their estimate for a disability weight is 0.542 (0.374-0.702), using the 2013 weight values. No distinction was made between stage 1 and 2 of the disease. The distribution for the parameter is Beta(18.37, 15.63).

It should be noted that other cost-effectiveness analyses have used different values for disability weights [78, 81, 82]. These values arise from the 1994 Global Burden of Disease Study but we prefer to consider updated values. Since most of the disability is due to deaths rather than illness during life, we do not believe that this difference is cause for concern.

##### E.7.5 Disability weights: SAE

↩ Return to the Summary of Health Outcome Parameters.

- Name in the code: `disability_weighting_sae`
- Source: [69]
- Country of estimate: GBD
- Statistical distribution and parameters: Uniform(0.04, 0.11)
- Summary statistics (mean and 95% CI or fixed value): 0.08 (0.04, 0.11)

**Notes**

As far as we are aware, no one has considered the disability due to severe adverse events attributable to gHAT treatment, but the most common adverse events are gastrointestinal problems and headaches.

We consulted the Global Burden of Disease for disability weights. The health state labeled “symptomatic tension-type headache” was described as “moderate headache that also affects the neck, which causes difficulty in daily activities” and was estimated to have a disability weight equal to 0.037 (0.022–0.057). The health state labeled “moderate symptomatic gastritis and duodenitis without anaemia” was described as “abdominopelvic problem, moderate has pain in the belly and feels nauseous; the person has difficulties with daily activities” and was estimated to have a disability weight equal to 0.114 (0.078–0.159).

Our distribution is therefore Uniform 0.037-0.114. Since most of the disability is due to death rather than illness during life, we do not believe that the uncertainty in this parameter is cause for concern for the purpose of the conclusions of this analysis.

#### E.8 Vector control parameters

↩ Return to the Summary of Health Outcome Parameters.

##### E.8.1 Linear km of targets

- Name in the code: `vc_length_default` and `vc_length_enhanced`
- Source: [30]
- Country of estimate: DRC
- Statistical distribution and parameters: Fixed value
- Summary statistics (mean and 95% CI or fixed value): Varies by health zone

**Notes**

The parameter for the extent of the riverbank covered by vector targets is a fixed value. The values for each health zone are detailed in the Supplementary Methods, section A.7.1, and see Supplementary Note 2 for the length of rivers covered by VC per health zone.

##### E.8.2 Target per km

↩ Return to the Summary of Health Outcome Parameters.

- Name in the code: `vc_density_linear`
- Source: [30]
- Country of estimate: DRC
- Statistical distribution and parameters: Fixed value, varies by health zone
- Summary statistics (mean and 95% CI or fixed value): Varies by health zone

**Notes**

In this analysis, we are using bank length, because, in some health zones, we do not cover with targets on both sides. In DRC, on average, while the goal is to cover rivers with 40 targets per kilometre, placing them 25m apart on alternating sides, or 50m apart on each side, records have shown that approximately 30 targets are used per kilometre. We parameterize the targets per kilometre as half that since we are considering the bank length, rather than the river length.

We used a fixed value for the parameter for the extent of the riverbank covered by vector targets.

##### E.8.3 Replacement rate of targets per year

↩ Return to the Summary of Health Outcome Parameters.

- Name in the code: `vc_deployments_yr`
- Source: [30, 83]
- Country of estimate: DRC
- Statistical distribution and parameters: Fixed value
- Summary statistics (mean and 95% CI or fixed value): 2

**Notes**

We have set this parameter as a fixed number, as this is the number of times that one must replace a set of targets in order to provide continuous protection throughout the year [30, 83].

#### E.9 Screening cost parameters

##### E.9.1 AS: capital costs of a team

↩ Return to the Summary of Cost Parameters.

- Name in the code: `as_cost_team_capital`
- Source: [43, 77, 78]
- Country of estimate: DRC
- Distribution and parameters: Gamma(820, 20.1)
- Summary statistics (mean and 95% CI or fixed value): 16,514 (15,410, 17,671)

**Notes**

Capital costs are denominated in 2024 US dollars.

To our knowledge, no active surveillance costs have been estimated via a detailed costing study [84]. We looked at the three studies that calculated costs using an ingredients approach, but decided on using the estimates from the more recent studies only [43, 77, 78].

Lutumba and colleagues [77] calculated that the total cost of screening 40,000 patients in 2003 was 46,734.29 Euros (1 Euro = 0.86 USD in 2003) (see table 3 of [77]). Of that value, 21 percent of the costs were capital costs, or a cost of 32,913 in 2024 USD. The publication did not indicate whether that value was annualized or not.

According to Bessel and colleagues [78] the cost for a mobile team that screens 250 patients per day for 220 days a year has capital investments (annualized for five years) of $12,000 for a team that administers CATT and $12,781 for a team that administers RDT (in 2013 USD) (see [78] Table S1). Adjusting for inflation, that would equal 17,671 in 2024 USD.

According to Snijders et al [43], the capital cost of an active surveillance team was $11,406 in 2018 for a team that administers CATT. Adjusting for inflation, that would equal 15,410 in 2024 USD.

Although no publication gave us a sense of the uncertainty in capital costs, we had three studies, but we felt that the Lutumba publication was too old, the costs were too high, and the question of annualization could influence the comparability of that cost. Nevertheless, we assumed that the range was equivalent to the 95% confidence interval. Therefore, our probability distribution is Gamma(820, 20.1), which yields a distribution with mean and confidence intervals of 16,514 (15,410, 17,671).

##### E.9.2 AS: recurrent & management costs of a team

↩ Return to the Summary of Cost Parameters.

- Name in the code: `as_cost_team_management`
- Source: [43, 77, 78]
- Country of estimate: DRC
- Distribution and parameters: Gamma(4153.88, 17.21)
- Summary statistics (mean and 95% CI or fixed value): 71,472 (69,311, 73,658)

**Notes**

Recurrent management costs are denominated in 2022 US dollars.

To our knowledge, no active surveillance costs have been estimated via a detailed costing study [84]. We looked at the three studies that calculated costs using an ingredients approach, but decided on using the estimates from the more recent studies only [43, 77, 78].

Lutumba and colleagues [77] calculated that the total cost of screening 40,000 patients in 2003 was 46,734.29 Euros (1 Euro = 0.86 USD in 2003, for an equivalent of $40,191) (see table 3 of [77]). Of that value, 79 percent of the costs were recurrent fixed costs ($31,751), or a cost of 123,817 in 2024 USD after adjusting for inflation and changes in the exchange rate.

According to Bessel and colleagues [78] the cost for a mobile team that screens 250 patients per day for 220 days a year has annual recurrent costs of $30,307 and daily recurrent costs of $97 in 2013 values (see [78], Table S1). Summing those costs ($51,647) and adjusting for inflation and changes in the exchange rate, would equal 73,658 in 2024 USD. Although Bessel and colleagues considered the cost of RDT use in AS, they assumed that the teams that would be using RDTs were traditional mobile units (with sport-utility vehicles) rather than the mini-mobile units with motorcycles that constitute the majority of RDT use in AS nowadays.

According to Snijders and colleagues [28], the management and recurrent costs of an active surveillance team were $42,408 in 2018, and $7,961 for the management costs from the provincial and central level PNLTHA (including training and supervision) for a total of $51031. Adjusting for inflation and changes in the exchange rate, would equal 69,311 in 2024 USD. It should be noted that these were the costs for a traditional team using sport utility vehicles and CATT tests, rather than a 'mini-team' using motorcycles and RDTs.

Although no publication gave us a sense of the uncertainty in capital costs, we had three studies, but we felt that the Lutumba publication was too old. If we assumed that the range of the other two observations was equal to the 95% confidence interval (69,311, 73,658), our probability distribution is Gamma(4153.88, 17.21), which yields a distribution with mean and confidence intervals of 71,472 (69,311, 73,658).

##### E.9.3 CATT algorithm: cost per test used

↩ Return to the Summary of Cost Parameters.

- Name in the code: `dx_cost_catt`
- Source: [85]
- Country of estimate: international market (Belgium)
- Distribution and parameters: Gamma(1140, 0.0006897)
- Summary statistics (mean and 95% CI or fixed value): $0.79 (0.74, 0.83)

**Notes**

The CATT test is sold in the international market by Institute of Tropical Medicine in Antwerp.

A kit of reagents is 280.18€ for 500 tests and accessories are 35.82€ for 250 tests, which yields a total of 0.70€ per person [85]. The capital necessary to carry out the test (the rotator field kit) is considered under the parameter for capital costs.

The price of the CATT varies due to uncertainty in the exchange with the USD. Judging by variation in the five years previous to and including 2024 (0.85-0.95), the range of the cost of the test can be between US$0.74-0.83, which is the 95% confidence interval of a distribution given by Gamma(1140, 0.0006897). The final distribution is given by $0.79 (0.74, 0.83) in USD.

Although delivery costs (described in E.10.6) are considered to be about 45% of the cost, after conversations with ITM colleagues, we decided to assume that the delivery cost for this is equal to a 10% markup per year. The markup is higher than the RDT tests because the CATT test shipping may need a cold-chain, whereas the RDT shipping would not (CATT reagents must be stored in temperatures of 2-8C; see E.9.6). The 10% estimate was also the assumption taken in a previous costing paper by Snijders and colleagues [43].

##### E.9.4 Staging: lumbar puncture & lab exam

↩ Return to the Summary of Cost Parameters.

- Name in the code: `dx_cost_lumbar_exam`
- Source: [43, 86]
- Country of estimate: DRC and Chad
- Distribution and parameters: Gamma(3.73, 2.96)
- Summary statistics (mean and 95% CI or fixed value): 13.76 (3.37, 31.08)

**Notes**

To our knowledge, costs for lumbar puncture tests were listed in detail only by Bessel and colleagues [78] and will be featured as part of an upcoming publication by Snijders and colleagues [43]. The cost listed by Bessel et al was 2.38 in terms of 2013 USD, equivalent to 3.39 in 2024 USD. Snijders and colleagues report a cost of 23.02 in 2018 USD, or 31.10 in 2024 USD. Irurzun-Lopez [86] have reported a similar value, so we will assume a value equal to that of Snijders and colleagues.

Because we had more than two studies to inform this parameter, we chose a gamma distribution where the 2.5th and the 97.5th percentiles would match the minimum and maximum values in the literature, so our parameter distribution is Gamma(3.59, 3.82), which yields a distribution with a mean and confidence interval of 13.76 (3.37, 31.08).

##### E.9.5 Confirmation: microscopy

↩ Return to the Summary of Cost Parameters.

- Name in the code: `dx_cost_microscopy`
- Source: [28]
- Country of estimate: DRC
- Distribution and parameters: Gamma(8.47, 1.71)
- Summary statistics (mean and 95% CI or fixed value): 13.92 (6.44, 25.75)

**Notes**

The per-patient price to confirm a patient with a full microscopy procedure is reported in an upcoming publication by Snijders et al [28]. They report that a microscopy procedure consisting of mAECT and LNA (lymph node aspiration) costs 9.53 in 2018 USD, or 12.88 in 2024 USD. Because we had no report of the standard error around that estimate, we assigned a gamma distribution that had a confidence interval that spanned half the estimate and twice the estimate, yielding a distribution of Gamma(8.47, 1.71), and a mean and confidence interval of 13.92 (6.44, 25.75).

##### E.9.6 RDT: costs per test used

↩ Return to the Summary of Cost Parameters.

- Name in the code: `dx_cost_rdt`
- Source: Co-authors from ITM
- Country of estimate: international market (Belgium)
- Distribution and parameters: Gamma(1140, 0.002205)
- Summary statistics (mean and 95% CI or fixed value): $2.51 (2.37, 2.66)

**Notes**

If one takes the Snijders et al estimate from the micro-costing analysis the cost is between 0.85 and 1.97 in 2018 USD, from Abbott and Coris, respectively. The Abbott RDT is no longer available, and therefore, one can only purchage tests from Coris. Those costs would be equivalent to 1.50€. However, as of 2025, Coris Sero-K-Set RDTs 2.25€ per test.

The price of the RDT varies due to uncertainty in the exchange with the USD. Judging by variation in the five years previous to and including 2024 (0.85-0.95), the range of the cost of the test can be between US$1.904-2.128, which is the 95% confidence interval of a distribution given by Gamma(1139.58093, 0.002205). The final distribution is given by $2.51 (2.37, 2.66).

Although delivery costs (described in E.10.6) are considered to be about 45% of the cost, after conversations with ITM colleagues, we decided to assume that the delivery cost for this is equal to a 5% markup per year. The markup is lower than the CATT tests because the CATT test shipping may need a cold-chain, whereas the RDT shipping would not (CATT reagents must be stored in temperatures of 2-8C; see E.9.3).

##### E.9.7 Indirect management costs (PNLTHA mark-up)

↩ Return to the Summary of Cost Parameters.

- Name in the code: `program_markup_vc`
- Source: [28]
- Country of estimate: DRC
- Distribution and parameters: Uniform(0.1, 0.2)
- Summary statistics (mean and 95% CI or fixed value): 0.15 (0.10, 0.20)

**Notes**

Snijder's and colleagues [28] have assumed that there is a component of management at the national programme (PNLTHA) level that is approximately 15% of the expenses at the local and coordination level (both fixed costs and variable/consumable costs for both active screening and passive screening in fixed health posts). However, the mark-up was not applied to the consult in the fixed health post, as these were consultations paid for by patients for symptoms in general, but testing and confirmation for HAT specifically is administered and paid for by PNLTHA, so we have included a mark-up for these items.

In this analysis, this was only included in this paper for VC costs and treatment, as the overhead for AS and PS activities was included in the general management of those activities.

##### E.9.8 PS: capital costs of a facility

↩ Return to the Summary of Cost Parameters.

- Name in the code: `ps_cost_facility_capital`
- Source: [28]
- Country of estimate: DRC
- Distribution and parameters: Gamma(8.475, 283.46)
- Summary statistics (mean and 95% CI or fixed value): 2393 (1067, 4269)

**Notes**

Capital costs are denominated in 2024 USD and apply to each health centre or hospital that is capable of HAT diagnosis.

We took into account the results of a micro-costing study in Yasa Bonga and Mosango [28], two health zones of Kwilu Province (Bandundu Sud coordination). The study reported a cost of 1580 USD in 2018 values, for an equivalent of 2135 in 2024 USD.

To parameterize the model, we assign a distribution with 95% confidence intervals equal to half and double the costs. The distribution is Gamma(8.47, 283.46), which yields a distribution with mean and confidence intervals of 2393 (1067, 4269).

##### E.9.9 PS: management costs

↩ Return to the Summary of Cost Parameters.

- Name in the code: `ps_cost_management`
- Source: [28]
- Country of estimate: DRC
- Distribution and parameters: Gamma(8.475, 145)
- Summary statistics (mean and 95% CI or fixed value): 1223 (560, 2157)

**Notes**

In 2025, the program costs for coordination management (which is the only management for PS) was $650,000 for the whole country for 179 health zones for support of both AS and PS activities. The micro-costing study in Yasa Bonga and Mosango [28] noted that about 30% of coordination effort is spent on support of fixed health facilities.

We do not adjust the cost between years because this has stayed roughly stable over the years, and the funding is all external (denominated in Euros or US Dollars, not in Congolese Francs).

$650K divided by 179 and taking only 30% of that would equal to $1090. Because we do not have a sense of how much this may change over the next few years, we still know there is some uncertainty in this estimate, so we apply a Gamma distribution where the 95% CI equals half and double our estimate: Gamma(8.475, 145) with mean estimate and PI of 1223 (560, 2157).

#### E.10 Treatment cost parameters

##### E.10.1 Hospital stay: cost per day

↩ Return to the Summary of Cost Parameters.

- Name in the code: `treat_cost_ip_day`
- Source: [40, 87, 88]
- Country of estimate: DRC
- Distribution and parameters: Gamma(5.81, 0.59)
- Summary statistics (mean and 95% CI or fixed value): 3.42 (1.22, 6.76)

**Notes**

We got the estimates of inpatient treatment costs from the 2010 WHO CHOICE cost estimates (recently updated by [87, 88]). In 2010, a consult at a primary hospital in DRC would be 2.41 (0.90, 5.73) I$. In 2010, a consult at a secondary hospital in DRC would be 2.59 (0.98, 5.81) I$, and a consult at a tertiary hospital in DRC would be 3.25 (1.32, 7.20) I$.

The equivalent estimates in 2010 USD are 1.40 (0.52, 3.33) per day at a primary hospital, 1.51 (0.57, 3.38) per day at a secondary hospital, and 1.89 (0.77, 4.19) per day at a tertiary hospital. After converting to local currency, applying the inflation index, and converting to 2024 USD, the estimates are 2.25 (0.84, 5.36) per day at a primary hospital, 2.42 (0.92, 5.43) per day at a secondary hospital, and 3.04 (1.23, 6.73) per day at a tertiary hospital.

At the moment, we do not know how many of each kind of hospital the population of HAT patients attend, nor do we understand how costs at district hospitals, referral hospitals, etc resemble those of the two kinds of hospitals under analysis by the WHO CHOICE program. Therefore, we take the estimate with a higher mean (hospital day in a tertiary hospital) in an effort not to under-state the costs of treatment and interventions.

To parameterize the model, we assign a gamma distribution with 95% confidence intervals equal to those reported by WHO CHOICE [40]. The distribution is Gamma(5.81, 0.59), which yields a distribution with median and confidence intervals of 3.42 (1.22, 6.76). Although this yields a higher mean, the uncertainty is adequately characterized.

##### E.10.2 Outpatient consultation: cost

↩ Return to the Summary of Cost Parameters.

- Name in the code: `treat_cost_op_visit`
- Source: [28, 89]
- Country of estimate: DRC
- Distribution and parameters: Gamma(24.2, 0.13)
- Summary statistics (mean and 95% CI or fixed value): 3.13 (2.02, 4.47)

**Notes**

We got the estimates of outpatient consultation costs from two sources:

1) Laokri and colleagues [89] presented an estimate with mean $2.33 and standard deviation 0.27 in 2013 values, or 2.01 (0.23) in 2024 USD values.
2) Snijders and colleagues [28] reported that the cost of a consultation is $3.33 in 2018 USD values, or $4.50 in 2024 values.

We will use the two mean estimates as the range of a gamma distribution, so we get Gamma(24.2, 0.13) and a sample with mean and 95% CI of 3.13 (2.02, 4.47)

##### E.10.3 Course of pentamidine: cost

↩ Return to the Summary of Cost Parameters.

- Name in the code: `rx_cost_pentamidine`
- Source: [84]
- Country of estimate: WHO
- Distribution and parameters: Fixed value
- Summary statistics (mean and 95% CI or fixed value): 54

**Notes**

The cost of pentamidine, for stage 1 disease. Because it is available on the international market, where is it sold in USD, and not subject to the inflationary pressures of any particular country, we have not inflated the cost or converted them to any other currency.

In the future pentamidine treatment it may be replaced with fexinidozole treatment, which would circumvent the need for a lumbar puncture.

##### E.10.4 Course of NECT: cost

↩ Return to the Summary of Cost Parameters.

- Name in the code: `rx_cost_nect`
- Source: [90]
- Country of estimate: WHO
- Distribution and parameters: Fixed value
- Summary statistics (mean and 95% CI or fixed value): 360

**Notes**

This represents the cost of NECT to the capital for stage 2 disease. Simarro and colleagues listed a cost of 1440 USD for the treatment of four patients.

Because it is available on the international market, where is it sold in USD, and not subject to the inflationary pressures of any particular country, we have not inflated the cost or converted them to any other currency.

In the future, it may be replaced with fexinidozole and this would be the drug for treatment failures or very severe patients.

##### E.10.5 Course of fexinidazole: cost

↩ Return to the Summary of Cost Parameters.

- Name in the code: `rx_cost_fexinidazole`
- Source: [82]
- Country of estimate: WHO
- Distribution and parameters: Fixed value
- Summary statistics (mean and 95% CI or fixed value): 50

**Notes**

The cost of fexanidozole, for stage 1 and 2 disease. In the near future this will be the drug of choice for first-line treatment for both stages of disease. It may require hospitalization, but eventually, it should be taken on an outpatient basis.

##### E.10.6 Drug delivery mark-up

↩ Return to the Summary of Cost Parameters.

- Name in the code: `rx_delivery_markup`
- Source: [87, 88]
- Country of estimate: DRC
- Distribution and parameters: Beta(45, 55)
- Summary statistics (mean and 95% CI or fixed value): 0.45 (0.35, 0.55)

**Notes**

Because we do not know the delivery price of drugs for each country, we have applied the standard value for the mark up of traded goods recommended by the WHO CHOICE programme for AFRO E: https://www.who.int/teams/health-systems-governance-and-financing/economic-analysis/costing-and-technical-efficiency/quantities-and-unit-pri

#### E.11 Vector control cost parameters

##### E.11.1 Operational cost per kilometer of riverbank covered

↩ Return to the Summary of Cost Parameters.

- Name in the code: `vc_cost_management`
- Source: [45]
- Country of estimate: DRC
- Distribution and parameters: Gamma(8.47, 57.7)
- Summary statistics (mean and 95% CI or fixed value): 491 (217, 869).

**Notes**

Vector control operational costs are denominated in 2024 US dollars on a per-kilometer basis.

To our knowledge, only one vector control micro-costing study has been performed in DRC by Snijders and colleagues [45]. In that study, centred in Yasa Bonga, Kwilu Province, targets were laid out across 210 km of river length (or 420km of river bank).

The operational costs came to 61,796 USD, or 294.27 USD (in 2016 values) per km. The equivalent cost is 435 in 2024 USD per kilometer.

As there was no sense of the uncertainty in VC operational costs, we assigned a distribution with confidence intervals equal to half and double the costs. The distribution is Gamma(8.47, 57.7), which yields a distribution with mean and confidence intervals of 491 (217, 869).

##### E.11.2 Deployment cost per target

↩ Return to the Summary of Cost Parameters.

- Name in the code: `vc_cost_deployment`
- Source: [45]
- Country of estimate: DRC
- Distribution and parameters: Gamma(8.47, 0.50)
- Summary statistics (mean and 95% CI or fixed value): 4.21 (1.88, 7.50)

**Notes**

Target deployment costs are denominated in 2024 US dollars on a per-target basis.

To our knowledge, only one vector control micro-costing study has been performed in DRC by Snijders and colleagues [45]. In that study, centred in Yasa Bonga, Kwilu Province, 22,622 targets were laid out across 210 km of river length (or 420 of river bank). The target deployment activities cost 57,571 USD, or per target 2.54 USD (in 2016 values). The equivalent cost is 3.75 in 2024 USD per target.

Uncertainty: as there was no sense of the uncertainty in target deployment costs, we assigned a distribution with confidence intervals equal to half and double the costs. The distribution is Gamma(8.47, 0.50), which yields a distribution with mean and confidence intervals of 4.21 (1.88, 7.50).

### F Supplementary Note 4: NTD PRIME Criteria

| **Principle and what has been done to satisfy the principle?** | **Where in the manuscript is this described?** |
|---|---|
| **1. Stakeholder engagement**<br>Strategy components were determined along with the country director of PNLTHA, Erick Miaka (co-author). Implementation of simulations of AS and PS costs was aided by Rian Snijders, who has helped run field operations in former Bandundu. Implementation of costs of VC was aided by Andrew Hope, Iñaki Tirados, Sophie Dunkley, and Rian Snijders and collaborators at the Liverpool School of Tropical Medicine, who have run field operations in DRC since 2015. | Authorship list and acknowledgements |
| **2. Complete model documentation**<br>Full model (including the fitting code) and documentation are available through OpenScienceFramework (OSF). The epidemiological model is fully described in the fitting study of Crump et al. [1] and cost model in the Supplementary Methods. | Description in Supplementary Methods, Sections A.1, through A.9 access the code via DRC_wholecountryCEA[1]. |
| **3. Complete description of data used**<br>Information about the data used for fitting is described in Crump et al. [1]. The data used for clinical outcomes and costs were estimates from the literature. No data from intervention operations was used. Assumptions and estimates were parameterised according to conventions in the economic evaluation literature [91]. | For assumptions around intervention, treatment effects and costs, see Supplemental Methods, Sections A.7, A.9, and A.10. |
| **4. Communicating uncertainty**<br>*Structural uncertainty*:<br>Uncertainty arising from the choice of vector control operation inputs, discounting, and time horizon are shown by re-running the entire analysis with alternative assumptions. | Main text results and discussion. Results with alternative time horizons and discounting are available in our GUI[2]. |
| *Parameter uncertainty*:<br>The epidemiological parameters are the posterior distributions of a model fitted to time-series data, and full details are available in another publication [1]. For the parameters to model health outcomes and costs, assumptions and estimates were parameterized according to conventions in the economic evaluation literature [91], taking care to sample from large distributions for aspects for which we knew very little.<br>*Prediction uncertainty*:<br>Observational uncertainty in epidemiological model predictions. Figures 4 present both means and 95% prediction intervals. The GUI[2] includes box and whisker plots to show uncertainty in cases, deaths and DALYs. We include the probability of meeting EOT by 2030 as well as the expected year of EOT. For cost-effectiveness results, we present the optimal decisions (the probability of each strategy being cost-effective) at different willingness-to-pay thresholds rather than solely providing ICERs using the net benefits framework (see Supplementary Tables 25, 26, 27, 28). | Epidemiological parameters are available on OSF DRC_wholecountryCEA [1]. Health outcome and cost-effectiveness parameters: see Supplementary Methods, Sections A.9 and A.10, and Supplementary Note 4.<br>The GUI[4]. |
| **5. Testable model outcomes**<br>Epidemiological model outputs are routinely reported metrics of the disease course: detected active and passive case detections. Therefore, these predictions can be compared to future data as long as the data is put into context alongside measures of active screening coverage and the number of fixed health posts equipped for passive screening. Some components of cost predictions can be validated against expenditures, but it must be noted that these are economic costs, and so resource use for which there is no explicit invoice is taken into account as well. | Epidemiological projections for the period 2026-2040 are shown in Figure 4 for the whole country, and by coordination (aggregated and by year in Supplementary Figures 12-19.<br>Epidemiological outcomes for each year until 2050 and by coordination and health zone can be viewed in the GUI[2].<br>All the ingredients to the economic costs were shown in detail in the supplement section A.10. |

[1] DRC_wholecountryCEA with full address: https://osf.io/ezjxb/.
[2] GUI with full address: https://hatmepp.warwick.ac.uk/DRCCEA/v8/

**Supplementary Table 33**: PRIME-NTD criteria fulfilment. We summarise how the NTD Modelling Consortium's "5 key principles of good modelling practice" have been met in the present study.

### G Supplementary Note 5: CHEERS Checklist

| Section/item | Item No | Recommendation | Reported on page no, line no |
|---|---|---|---|
| **Title** | | | |
| Title | 1 | Identify the study as an economic evaluation or use more specific terms such as "cost-effectiveness analysis", and describe the interventions compared. | Cover page |
| **Abstract** | | | |
| Abstract | 2 | Provide a structured summary of objectives, perspective, setting, methods (including study design and inputs), results (including base case and uncertainty analyses), and conclusions. | Abstract |
| **Introduction** | | | |
| Background and objectives | 3 | Provide an explicit statement of the broader context for the study. Present the study question and its relevance for health policy or practice decisions. | Introduction section, in particular the second-to-last and the last paragraph. |
| **Methods** | | | |
| Health economic analysis plan | 4 | Indicate whether a health economic analysis plan was developed and where available. | No previous protocol was published. |
| Study population | 5 | Describe characteristics of the study population (such as age range, demographics, socioeconomic, or clinical characteristics). | The population is described in the Supplementary Methods, Section A.3 and Supplementary Tables 2, and 3. |
| Settings and location | 6 | Provide relevant contextual information that may influence findings. | The location is described in the Supplementary Methods, Section A.3 and Supplementary Tables 2, and 3. |
| Comparators | 7 | Describe the interventions or strategies being compared and state why they were chosen. | Figure 1; second paragraph of the Results section; third section of the methods; and more detail in the Supplementary Methods, Section A.7, Supplementary Table 8, Supplementary Figure 3, and Supplementary Table 9. |
| Perspective | 8 | State the perspective(s) adopted by the study and the rationale. | Fourth subsection of the methods, "Cost-effectiveness analysis", in a subsection called "Costs". |
| Time horizon | 9 | State the time horizon over and why the horizon is appropriate. | Fourth subsection of the methods, "Cost-effectiveness analysis", in a subsection called "Economic evaluation and investment horizon". |
| Discount rate | 10 | Report the discount rate(s) and reason chosen. | Fourth subsection of the methods, "Cost-effectiveness analysis", in a subsection called "Economic evaluation and investment horizon". 3% is the recommended rate by WHO-CHOICE and the Gates Reference Case. |

*(continued)*

| Section/item | Item No | Recommendation | Reported on page no, line no |
|---|---|---|---|
| Selection of outcomes | 11 | Describe what outcomes were used as the measure(s) of benefit(s) and harm(s). | Fourth subsection of the methods, “Cost-effectiveness analysis”, in a subsection called “Health outcomes”. Moreover, Supplementary Methods, Sections A.9 and A.10.4 contain detailed explanations of how DALYs are calculated and how the natural history of HAT was considered. |
| Measurement of outcomes | 12 | Describe how outcomes used to capture benefit(s) and harm(s) were measured. | Outcomes were not measured, but were simulated. See the fourth subsection of the methods, "Outcome metrics". Effectiveness of AS, PS and VC strategies: model-based, treatment was sourced from the literature. Described in detail in Supplementary Methods, Section A.10.4, Supplementary Tables 20-21, and Supplementary Note 4, Section E.6. |
| Measurement and valuation of resources and costs | 12 | If applicable, describe the population and methods used to elicit preferences for outcomes. | The construction of programme costs are detailed in Supplementary Methods, Section A.10. As can be seen, in a publication of this scope, detailing the cost inputs in the main body would be unfeasible. However, the resulting expected costs are in Figure 4 of the main body of the paper, as well as in 20 broken down by activity, and in 21 broken down by coordination. In the `GUI`, one may find the costs broken down by activity by health zone, coordination, and the country for under a variety of sensitivity analyses. |
| Currency, price date, and conversion | 15 | Report the dates of the estimated resource quantities and unit costs, plus the currency and year of conversion. | Fourth subsection of the methods, “Cost-effectiveness analysis”, in a subsection called “Costs”. Our general approach for this is in the section "Principles for parameterization" in Supplementary Note 4, Section E.1, followed by the specific choices for each parameter. |

*(continued)*

| Section/item | Item No | Recommendation | Reported on page no, line no |
|---|---|---|---|
| Rationale and description of model | 16 | If modelling is used, describe in detail and why used. Report if the model is publicly available and where it can be accessed. | The decision analytic model is illustrated in detail in Supplementary Figure 4 and in full in Supplementary Figure 5, but the components models feeding into the decision analytic model are described as follows: the dynamic transmission (SEIRS) model is described briefly in the third methods section and Supplementary Methods, Section A.1 and pictured in Supplementary Figure 2, and the treatment model is described briefly in Supplementary Methods, Section A.9 and shown in Supplementary Figure 6. |
| Analytics & assumptions | 17 | Describe any methods for analysing or statistically transforming data, any extrapolation methods, and approaches for validating any model used. | The transmission and treatment models are discussed in the Supplementary Methods, Sections A.1-A.6. The treatment model is described in detail in Sections A.9. Assumptions are described in detail in Supplementary Methods, Section A.10 and the parameter glossary in Supplementary Note 4. |
| Characterising heterogeneity | 18 | Describe any methods used for estimating how the results of the study vary for subgroups. | Heterogeneity across health zones and across coordinations was characterised by aggregation and disaggregation, showing how different geographic portions of the country may need different resources and investments to reach disease control and elimination goals. |
| Characterising distributional effects | 19 | Describe how impacts are distributed across different individuals or adjustments made to reflect priority populations. | Because there were no subgroups, there were no distributional effects. The differential impacts of treatment on poorer or less poor individuals were beyond the scope of this paper. |
| Characterising uncertainty | 20 | Describe methods to characterise any sources of uncertainty in the analysis. | The epidemiological parameters are the posterior distributions of a model fitted to time-series data, and full details are available in the Supplementary Methods, SectionsA.5-A.6. For the parameters to model health outcomes and costs, assumptions and estimates were parameterized according to conventions in the economic evaluation literature [91], taking care to sample from large distributions for aspects for which we knew very little. Epidemiological parameters are available on OSF `https://osf.io/ezjxb/`. Health outcome and cost-effectiveness parameters: see Table 1 and Supplementary Note 4. |

*(continued)*

| Section/item | Item No | Recommendation | Reported on page no, line no |
|---|---|---|---|
| Approach to engagement with patients and others affected by the study | 21 | Describe any approaches to engage patients or service recipients, the general public, communities, or stakeholders (such as clinicians or payers) in the others affected by the study. | Strategy components were determined along with the country director of PNLTHA, Dr. Erick Miaka and other PNLTHA members, Chancy Shampa, and Junior Lebuki (co-authors). Implementation of simulations of AS and PS costs was aided by information from Rian Snijders and Paul Verlé (co-authors), who has helped with operations in Bandundu Coordination and collaborators at the Liverpool School of Tropical Medicine, Andrew Hope, Iñaki Tirados, and Sophie Dunkley (co-authors) who have run vector control field operations in DRC since 2015. |
| **Results** | | | |
| Study parameters | 22 | Report the values, ranges, references, and, if used, probability distributions for all parameters. Report reasons or sources for distributions used to represent uncertainty where appropriate. Providing a table to show the input values is strongly recommended. | Table 1 and described in more detail in Supplementary Note 4. |
| Incremental costs and outcomes | 23 | For each intervention, report mean values for the main categories of estimated costs and outcomes of interest, as well as mean differences between the comparator groups. If applicable, report incremental cost-effectiveness ratios. | For four sample health zones, the intermediate outcomes are in Supplementary Tables 25-28; for the whole country, the outcomes can be found in 4; for each coordination, the results are in 12-19. For the whole country, the cost-effectiveness results are in Figures 2 and 3 and the same results can be found per coordination in Supplementary Figure 11. |
| Effect of uncertainty | 24 | Model-based economic evaluation: Describe the effects on the results of uncertainty for all input parameters, and uncertainty related to the structure of the model and assumptions. | For four sample health zones, the effect of uncertainty on the cost-effectiveness are shown in Supplementary Tables 25-28. The interpretation is in the Supplementary Methods, Section B.4. For all other health zones, the results of uncertainty on the CEA is shown in the GUI: `GUI`. The effect of uncertainty on the Elimination of Transmission goal is shown in Figure 3. The interpretation of the uncertainty is in the Results section. |
| Effect of engagement with patients and others affected by the study | 25 | Report on any difference patient/service recipient, the general public, community, or stakeholder involvement made to the approach or findings of the study | Our engagement with the stakeholders (co-authors) was iterative throughout the process |
| **Discussion** | | | |

*(continued)*

| Section/item | Item No | Recommendation | Reported on page no, line no |
|---|---|---|---|
| Study findings, limitations, generalisability, and current knowledge | 26 | Summarise key study findings and describe how they support the conclusions reached. Discuss limitations and the generalisability of the findings and how the findings fit with current knowledge. | Discussion. |
| **Other relevant information** | | | |
| Source of funding | 27 | Describe how the study was funded and the role of the funder in the identification, design, conduct, and reporting of the analysis. Describe other non-monetary sources of support. | Funding statement. |
| Conflicts of interest | 28 | Describe any potential for conflict of interest of study contributors in accordance with journal policy. In the absence of a journal policy, we recommend authors comply with the International Committee of Medical Journal Editors recommendations. | Conflict of interest statement. |

51. Betu Kumeso VK, Kalonji WM, Rembry S, Valverde Mordt O, Ngolo Tete D, Prêtre A, Delhomme S, Ilunga Wa Kyhi M, Camara M, Catusse J, Schneitter S, Nusbaumer M, Mwamba Miaka E, Mahenzi Mbembo H, Makaya Mayawula J, Layba Camara M, Akwaso Massa F, Kaninda Badibabi L, Kasongo Bonama A, Kavunga Lukula P, Mutanda Kalonji S, Mariero Philemon P, Mokilifi Nganyonyi R, Embana Mankiara H, Asuka Akongo Nguba A, Kobo Muanza V, Mulenge Nasandhel E, Fifi Nzeza Bambuwu A, Scherrer B, Strub-Wourgaft N, and Tarral A. Efficacy and safety of acoziborole in patients with human African trypanosomiasis caused by Trypanosoma brucei gambiense: a multicentre, open-label, single-arm, phase 2/3 trial. The Lancet Infectious Diseases 2023 Apr; 23:463–70. DOI: 10.1016/S1473-3099(22)00660-0

74. Briggs A, Claxton K, and Sculpher M. Decision Modelling for Health Economic Evaluation. First. Oxford, UK: Oxford University Press, 2006

83. Liverpool School of Tropical Medicine. Use of Tiny Targets to control tsetse flies in Gambian HAT foci: standard operating procedures. Tech. rep. October. 2016

84. Keating J, Yukich JO, Sutherland CS, Woods G, and Tediosi F. Human African trypanosomiasis prevention, treatment and control costs: A systematic review. Acta Tropica 2015 Oct; 150:4–13. DOI: 10.1016/j.actatropica.2015.06.003. Available from: http://dx.doi.org/10.1016/j.actatropica.2015.06.003%20https://linkinghub.elsevier.com/retrieve/pii/S0001706X15300267

85. CATT/T.b.g. REAGENTS (KIT R500 tests) - ITM Webshop for in vitro diagnostics. Available from: https://diagnostics.itg.be/product/catt-t-b-g-reagents-kit-r500-tests/
